# The information in diagnostic tests

**DOI:** 10.64898/2026.09.13.26362573

**Authors:** Shuhan He, Brian W. Locke, Joshua W. Joseph, David M. Liebovitz, Cory Rohlfsen, Allison Goff, Pedram Safari, Christopher Kabrhel, Amal Mohamed, Ali S. Raja, Joshua N. Goldstein

## Abstract

**Background:** Diagnostic accuracy measures describe test performance, but expected learning also depends on the probability of disease before testing. We explain diagnostic information as expected uncertainty reduction and construct an empirical reference for its interpretation.

**Methods:** We reanalysed structured study-level diagnostic accuracy data from Cochrane reviews, public repositories and PubMed Central review tables. A common continuity-corrected bivariate random-effects model pooled sensitivity and specificity for defined diagnostic profiles. We calculated mutual information in bits and as the percentage of starting uncertainty resolved at stated disease probabilities. Profiles contributed equally to the reference distribution. Sensitivity analyses examined alternative estimators, equal-review weighting and whole-review resampling.

**Results:** The reference included 273 pooled profiles from 210 reviews, comprising 4104 study-result appearances. Median uncertainty resolved was 23.3%, 29.2% and 31.4% at starting probabilities of 5%, 20% and 50%, respectively. At 20%, the median represented 0.211 of 0.722 bits, with a profile-bootstrap 95% confidence interval of 27.6% to 32.7% for the percentage resolved. Equal-review weighting gave 30.0%; whole-review resampling gave an interval of 27.5% to 33.5%. Two components of the head impulse, nystagmus and test of skew examination with nearly equal Youden indices resolved 18.7% and 28.1% at a 5% starting probability. Carcinoembryonic-antigen thresholds illustrated probability-dependent information ordering; the Ottawa ankle rule distinguished expected learning from uncertainty change after one result.

**Conclusions:** Diagnostic information makes expected learning explicit alongside conventional accuracy measures. Reporting bits and percentage of starting uncertainty resolved, with the starting probability and clinical context, supports interpretation and comparison of diagnostic evidence.

## Introduction

Clinicians begin with a probability that a condition is present and revise it as evidence arrives. Sensitivity and specificity, combined with that starting probability, determine how much uncertainty a test is expected to resolve. Information theory quantifies learning from signals,[1,2] and diagnostic information applies that measure to the question of what to investigate next. Evidence-based medicine (EBM) does not routinely report this expected learning.

Mutual information has been used to evaluate single diagnostic tests since the 1970s; Benish developed it as an index of test performance and reviewed the field,[3,4] and Somoza and Mossman used it to compare and optimise tests.[5] Tsalatsanis and colleagues showed that it can be meta-analysed, with two worked examples, and proposed interpretive thresholds by analogy with treatment effect sizes.[6] Our own studies applied entropy removal to diagnostic tools and clinical features.[7,8] We extend that work with an empirical reference scale: 273 pooled estimates of sensitivity and specificity for defined tests and thresholds from 210 reviews. We calculate expected uncertainty reduction at different starting probabilities, compare it with conventional accuracy measures, and show how the expectation before testing differs from the evidence provided by one result.

### Summary points

- Diagnostic information makes expected learning explicit when comparing candidate investigations before their results are known, using sensitivity, specificity, and a stated starting probability.
- Diagnostic reviews can report the percentage of starting uncertainty a test is expected to resolve alongside sensitivity and specificity, with bits as the underlying absolute measure.
- Across 273 pooled profiles from 210 reviews, the median profile resolved 29.2% of starting uncertainty at a standardised 20% disease probability.
- Diagnostic information depends on the starting disease probability and the test’s performance in the relevant population and setting.

### What is diagnostic information?

Diagnostic uncertainty is greatest when disease and no disease are equally likely. One bit is the uncertainty in that balanced yes-or-no question. At a probability of 20%, the starting uncertainty is 0.722 bits; at 5%, it is 0.286 bits. A question that already has a very unlikely or very likely answer contains less uncertainty for a test to remove.

Let *D* denote disease status (1 for condition present, 0 for absent), and let *p* = *P* (*D* = 1) be the starting disease probability, where *P* means probability. The starting uncertainty *H*(*D*), called binary entropy, is

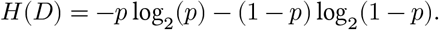

Here, 1 ™ *p* is the probability that the condition is absent, and log_2_ is the base-2 logarithm, giving entropy in bits. Let *T* denote the test result, with *t* = + for a positive result and *t* = ™ for a negative result. *P* (*T* = *t*) is the probability of that result, and *H*(*D* ∣ *T* = *t*) is the uncertainty remaining after it. Expected diagnostic information *I*(*D*; *T*) is the starting uncertainty minus the average remaining uncertainty:

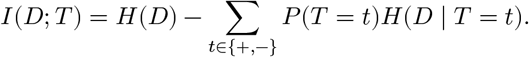

The sum adds the uncertainty after each possible result, weighted by that result’s probability; this mutual information is therefore a summary derived from sensitivity, specificity, and starting probability. Reporting it as the percentage of starting uncertainty removed, the uncertainty coefficient of statistics,[9] shows how much of the diagnostic question the test is expected to resolve; bits preserve the absolute unit used to calculate and compare that information.

Each pooled diagnostic estimate represents a defined test or positivity threshold for a stated target condition, population, setting, and reference standard, so its information value belongs to that diagnostic profile and starting probability; it is not a context-free property of a test name, and it concerns one binary target condition, not the whole differential diagnosis or all uncertainty in an encounter.

**Box 1. What can one more test tell us?**

- **Starting point:** In this hypothetical illustration, a 64-year-old man presents with chest pain. After the history, examination, and ECG, suppose his probability of myocardial infarction is 20% before troponin testing.
- **If the result is positive:** With stipulated sensitivity of 90% and specificity of 95%, the probability of myocardial infarction rises to about **82%**.
- **If the result is negative:** The probability of myocardial infarction falls to about **3%**.
- **Before the result is known:** Either branch can occur, so expected diagnostic information combines both possible results using their model-implied probabilities (22% positive and 78% negative).
- **Expected diagnostic information:** The test is expected to resolve **60.6% of the uncertainty present before testing** (0.437 of 0.722 bits). This is not a post-test probability; the observed result determines the patient’s post-test probability.

Supplementary figure S10 shows this weighted expectation on the entropy curve and repeats the hypothetical calculation at a 5% starting probability.

### Putting diagnostic information on an empirical scale

#### Evidence base

The protocol was registered with ConductScience (CSR-DI-2026-001; https://doi.org/10.55157/csr.di.2026.001).

We built the empirical reference scale from structured diagnostic accuracy data in the 2018 Cochrane corpus and recent Cochrane reviews, public repositories, and machine-readable PubMed Central review tables.[10,11] Each included study result had a study identifier, the four counts of its 2×2 table, and a link to its source; these counts give sensitivity and specificity, which we pooled across studies of the same test and definition and combined with a stated starting probability to calculate diagnostic information.

Each source-defined test, target condition, threshold, or analysis definition formed a candidate group. Groups with nonnegative 2×2 counts, at least five unique study identifiers, one result per identifier, and at least three studies with both disease states represented entered bivariate synthesis in their source-defined test direction; these rules produced 444 model-ready groups, and all 444 fits converged.

The primary-selection rules retained 273 pooled profiles from 210 reviews; the other 171 groups were alternative analyses retained in the audit record (supplement S1). Cochrane reviews could contribute multiple eligible profiles, whereas public-repository and PubMed Central routes retained at most one per review. The selected profiles contained 4104 study-result appearances and contributed equally to the reference distribution.

#### Estimating sensitivity and specificity for each test or threshold

We pooled each selected test or threshold independently, meta-analysing sensitivity and specificity jointly with a bivariate random-effects model fitted by restricted maximum likelihood (REML), which lets both measures vary between studies and estimates their between-study association.[12] The fitted mean logits, transformed back to the probability scale, gave the summary sensitivity and specificity used in the information calculations.

Because zero cells create undefined study-level logits, the prespecified primary analysis added 0.5 to every cell in every study before calculating logits and their sampling variances; sensitivity analyses applied the correction only to studies with a zero cell and refitted the previously specified regularised direct-binomial model. We report the 95% pooled-mean interval, which describes uncertainty in the estimated average sensitivity and specificity, and the predictive range, which adds estimated between-study heterogeneity; both are plug-in summaries conditional on the fitted heterogeneity parameters. Estimation details, numerical diagnostics, zero-cell correction analyses, recovery checks, and estimator sensitivity analyses are in the supplement.

#### Calculating and examining diagnostic information

For each pooled estimate, we calculated how much diagnostic uncertainty the test was expected to remove at starting disease probabilities of 5%, 20%, and 50%, reported primarily as the percentage of starting uncertainty removed with the corresponding bits where useful, and across starting probabilities from 1% to 50% in 1% increments; when two information profiles changed order, numerical root finding estimated the crossing. The three standard probabilities were standardised comparison conditions, not prevalence estimates or clinical action thresholds. We estimated 95% confidence intervals for the median percentages by bootstrap resampling of the 273 pooled estimates (2000 replicates, percentile intervals, fixed seed); these intervals describe sampling of the reference sample and do not propagate uncertainty within each pooled estimate.

We assessed the influence of reviews contributing multiple profiles by giving each review equal total weight and by resampling whole reviews with their profiles kept together (supplement S11).

We compared diagnostic information with Youden’s J using Spearman correlations at each starting probability, sensitivity-specificity maps with equal-Youden contours, and within-review profiles with nearly equal J. Mathematical examples examined equal J with different information, similar information with different post-negative probabilities, and ordering changes across starting probabilities (supplements S3 and S6-S8). These establish mathematical distinctions without estimating their clinical frequency.

We then used the 2.5 and 5 µg/L carcinoembryonic antigen (CEA) thresholds for detecting recurrent colorectal cancer as a clinical example of why starting probability matters,[13] calculating their information across the prespecified range and identifying where their ordering changed under the primary and alternative synthesis models; the crossings compare expected uncertainty reduction, whereas an action threshold would require result-specific post-test probabilities and the consequences of acting or not acting.

Mutual information describes expected uncertainty reduction before testing. For each result, we also calculated its probability, likelihood ratio, post-test probability, entropy reduction *H*(prior)™*H*(posterior), and information gain *D*_*KL*_(posterior∥prior). Entropy reduction can be negative when a result moves probability towards 50%; posterior-to-prior Kullback-Leibler (KL) gain is non-negative. Each quantity averages to mutual information when weighted by result probabilities. Supplement S9 gives the definitions and sign conventions.

OpenAI Codex assisted with code development, numerical checks, source retrieval and document preparation under author review; supplement S12 describes the tasks and verification.

### How much information do diagnostic tests provide?

At starting probabilities of 5%, 20%, and 50%, the median profiles resolved 23.3%, 29.2%, and 31.4% of starting uncertainty, respectively. At 20%, this was 0.211 of 0.722 bits. Table 1 gives the distributions and bootstrap intervals.

**Table 1.** Empirical reference scale for diagnostic uncertainty resolved.

footnote:* IQR is the 25th–75th percentile.
| Starting disease probability | Median uncertainty resolved, % (IQR) | 95% CI of the median, % | 10th–90th percentile range, % |
| --- | --- | --- | --- |
| 5% | 23.3% (17.3%–37.6%) | 22.0%–26.7% | 10.7%–50.4% |
| 20% | 29.2% (22.1%–45.6%) | 27.6%–32.7% | 14.0%–58.5% |
| 50% | 31.4% (23.3%–48.3%) | 29.5%–34.9% | 15.2%–58.3% |

Reviews contributing multiple profiles did not materially drive the reference median in these checks: equal-review weighting changed it from 29.2% to 30.0% at a 20% starting probability, while whole-review resampling gave a similar 95% interval (27.5-33.5%; supplementary table S1).

Figure 1 shows the reference distribution from 1% to 50% starting probability: the median and percentile bands summarise the same 273 pooled estimates, three reference medians are labelled directly, and four clinical examples orient familiar tests within it.

**Figure 1.**
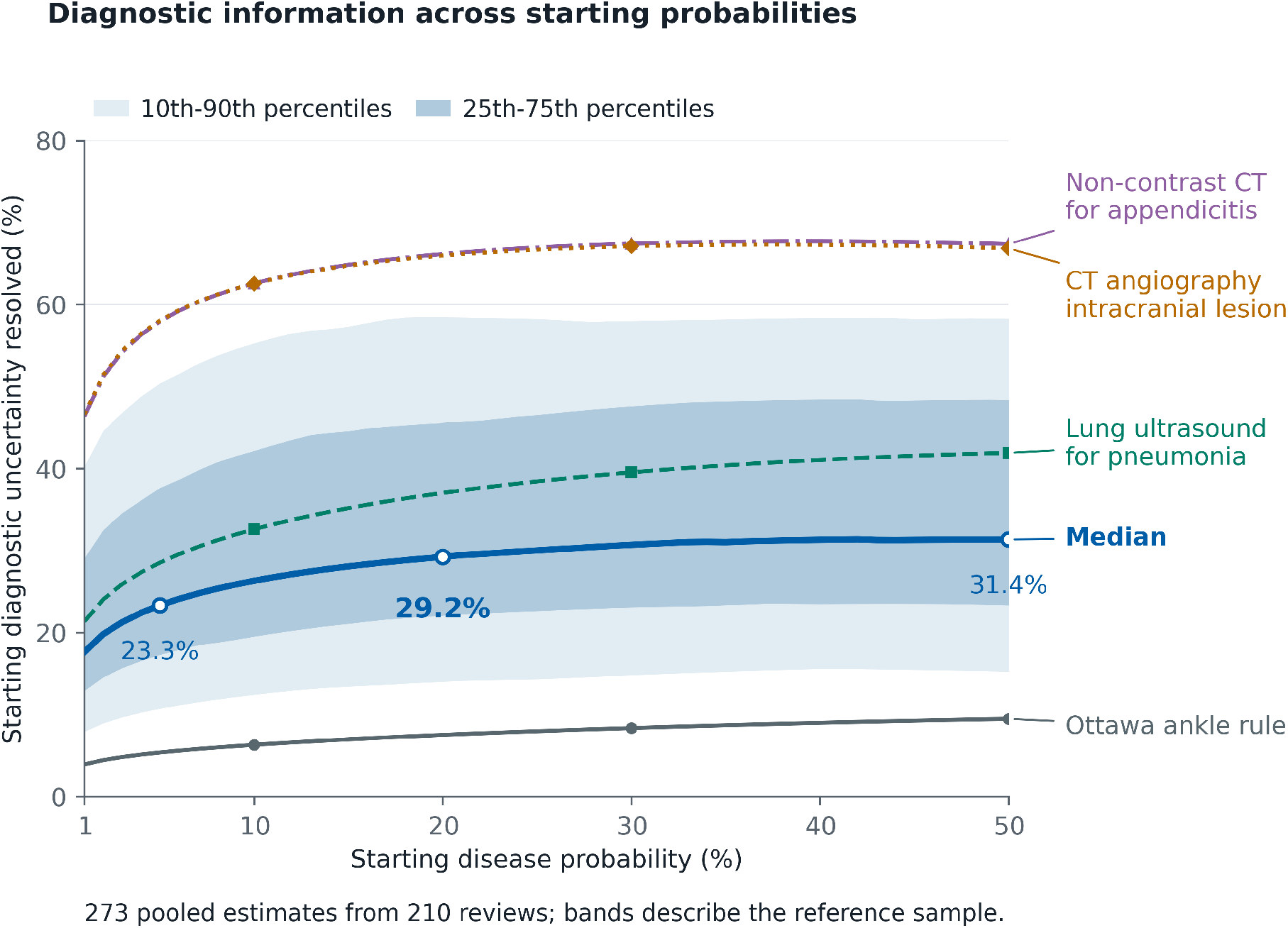
Empirical reference map of diagnostic information across starting probabilities. The median line and 25th–75th and 10th–90th percentile bands show the percentage of starting diagnostic uncertainty resolved by the same 273 pooled estimates from 210 reviews across starting disease probabilities from 1% to 50%. Median reference values at 5%, 20%, and 50% are labelled; the 29.2% median at 20% is emphasized. The bands describe the distribution of pooled estimates, not confidence intervals. The Ottawa ankle rule, lung ultrasound for pneumonia, non-contrast computed tomography for appendicitis, and computed tomography angiography for an intracranial lesion are clinical examples used to orient the scale. The examples do not rank clinical value or represent how often tests are used.

### Where information fits among diagnostic measures

Diagnostic information sits between test performance and decision value: it measures expected reduction in uncertainty before the result is known. Figure 2 maps all 273 pooled estimates over the sensitivity-specificity plane at starting probabilities of 5%, 20%, and 50%; equal-Youden contours stay fixed while the information surface changes with starting probability. Information and Youden’s J were strongly related because both derive from sensitivity and specificity: their Spearman correlations were 0.954, 0.975, and 0.992.

**Figure 2.**
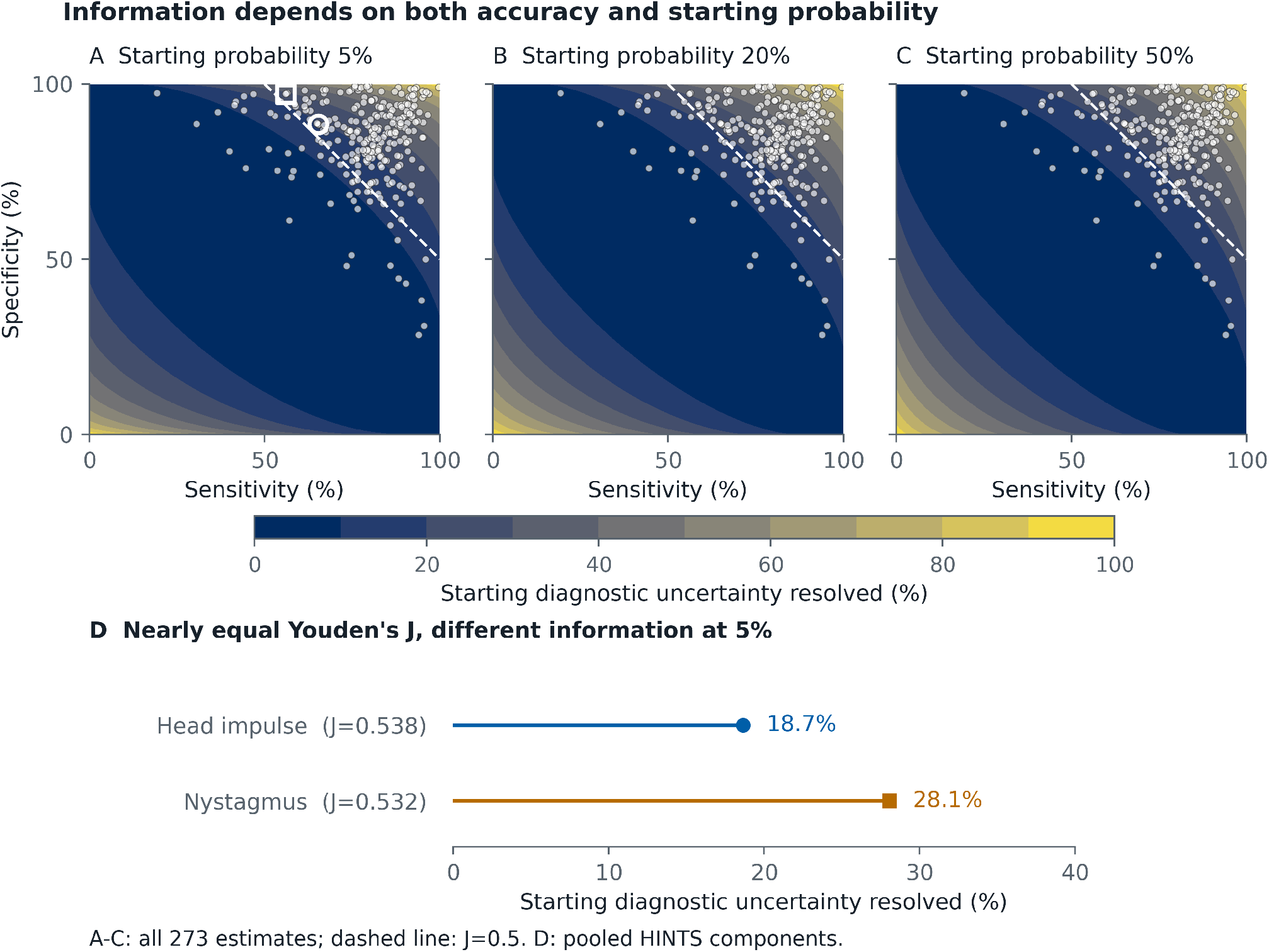
What diagnostic information adds beyond sensitivity, specificity, and Youden’s J. (A–C) Matched surfaces show the percentage of starting diagnostic uncertainty resolved at 5%, 20%, and 50%; all 273 pooled diagnostic estimates are overlaid. A dashed line in each map marks J=0.50. (D) A separate comparison shows the two highlighted HINTS component profiles at a 5% starting probability. Youden’s J is unchanged by starting probability, while the information surface changes. In pooled component profiles from review CD015089, the directly labelled head-impulse and nystagmus profiles had nearly equal J values (0.538 and 0.532) but resolved 18.7% and 28.1% of starting uncertainty at 5%. The comparison describes expected uncertainty reduction and does not establish clinical superiority or net benefit. HINTS denotes head impulse, nystagmus and test of skew.

The measures answer different questions. Youden’s J is sensitivity plus specificity minus one and contains no starting probability, whereas diagnostic information weights both result branches by how often each is expected. In pooled profiles from one review of central causes of acute vestibular syndrome, the head-impulse and nystagmus components had nearly identical Youden’s J values (0.538 and 0.532) but different sensitivity-specificity trade-offs,[14] and at a 5% starting probability they resolved 18.7% and 28.1% of starting uncertainty (figure 2). A near-tie in Youden’s J is therefore not a tie in expected diagnostic clarification, and the more informative profile is not automatically preferable: the consequences of false negatives and false positives still decide the strategy.

**Box 2. What each diagnostic measure tells us**.

| Measure | Question answered |
| --- | --- |
| Sensitivity and specificity | How often the test is positive among people with the target and negative among people without it |
| Youden's J or area under the receiver operating characteristic curve (AUROC) | How well the test or model separates the target states |
| Likelihood ratio and post-test probability | How one observed result changes the odds and probability for a patient |
| Diagnostic information | How much uncertainty about the target the evidence is expected to resolve at a stated starting probability |
| Calibration | Whether predicted probabilities agree with observed event frequencies |
| Brier score or log loss | How well probability forecasts predict observed outcomes under a specified proper scoring rule |
| Net benefit, utility, or expected value of information | Whether acting on the evidence improves outcomes, or how much a test is expected to improve a decision, once consequences and preferences are specified |

Value-of-information analysis asks how much a test is expected to improve a decision, given consequences and preferences.[15] Shannon entropy is the minimum expected logarithmic loss when forecasting the target probability; its expected reduction is mutual information. More generally, uncertainty can be expressed as the minimum expected loss over available actions, with a test valued by its expected reduction.[16] This connects learning to decision theory without supplying clinical losses: equal false-positive and false-negative costs do not recover Shannon entropy. A result can also guide the next investigation before it changes treatment; evaluating that sequence requires a model of subsequent tests and actions.

Additional mathematical comparisons and their proofs are reported in the supplement.

### Clinical case: recurrence probability changes how much information each CEA threshold provides

After treatment for colorectal cancer, carcinoembryonic antigen (CEA) is measured during surveillance for recurrence. Lowering the positive threshold to 2.5 µg/L labels more results positive, detecting more recurrences while creating more false alarms than the 5 µg/L threshold.[13] Their fitted Youden’s J values were 0.60 and 0.56; because J contains no starting probability, it ranks the lower threshold higher everywhere, whereas information can change the ordering because the expected frequencies of positive and negative results change with starting probability.

At a 5% starting probability, the 2.5 and 5 µg/L thresholds removed 19.1% and 19.5% of starting uncertainty; the greater specificity of the higher threshold preserved slightly more information when recurrence was uncommon. In the primary continuity-corrected bivariate REML analysis, their information curves crossed at 12.8%. At a 20% starting probability, the two thresholds removed 24.8% and 24.4%, so the greater sensitivity of the lower threshold resolved slightly more uncertainty (figure 3A).

**Figure 3.**
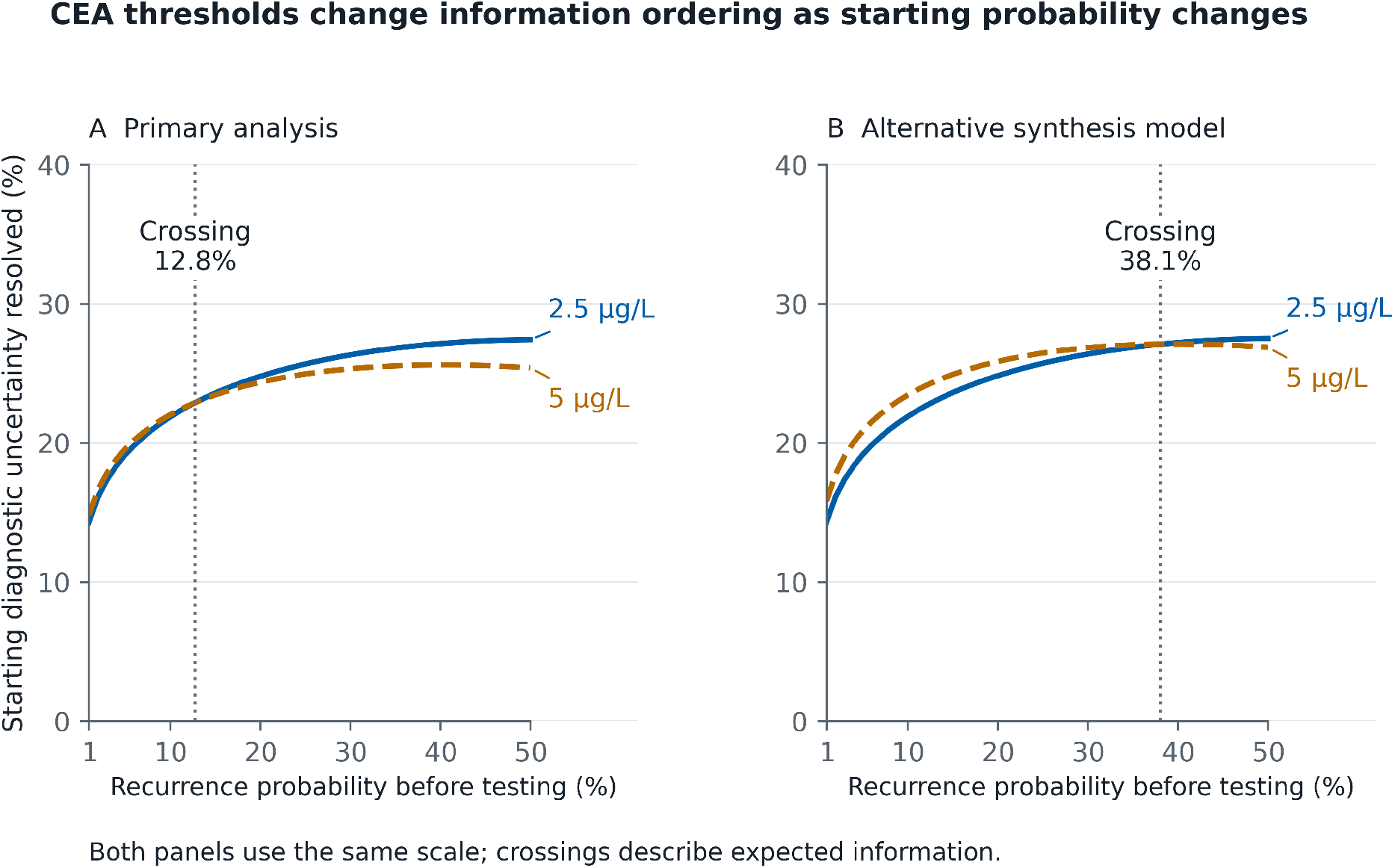
CEA information profiles for detecting recurrent colorectal cancer. (A) In the primary analysis, the 2.5 and 5 µg/L thresholds removed 19.1% and 19.5% of starting uncertainty at 5%, crossed at 12.8%, and removed 24.8% and 24.4% at 20%. (B) The alternative synthesis model crossed at 38.1%. The lower threshold favours sensitivity; the higher threshold favours specificity. The starting probability can therefore change which threshold is expected to resolve more uncertainty. Both panels use the same axes; the crossings do not imply a large information difference or clinical superiority. CEA denotes carcinoembryonic antigen.

In the regularised direct-binomial sensitivity analysis, the curves crossed at 38.1% (figure 3B): the ordering reversed under both models, while the exact crossing depended on the synthesis model. Information therefore identifies which threshold is expected to clarify the diagnosis more at the stated starting probability; the consequences of missed recurrence, false alarms, follow-up testing, and patient preferences determine which threshold should guide action.

### What an observed result can do

Mutual information is an average before the result is known; the result one patient receives may tell a different clinical story. For the Ottawa ankle rule under a standardised 5% starting probability, the pooled model implies a 70.4% probability of a positive result, which raises the probability of fracture only to 6.8%. This weak rule-in result moved the probability toward 50% and increased uncertainty by 0.071 bits. The model implies a 29.6% probability of a negative result, which lowers the probability to 0.8%, reducing uncertainty by 0.222 bits and supporting the rule’s intended rule-out role. Across both possible results, the rule is expected to remove 5.4% of starting uncertainty on average (0.015 bits; figure 4).

**Figure 4.**
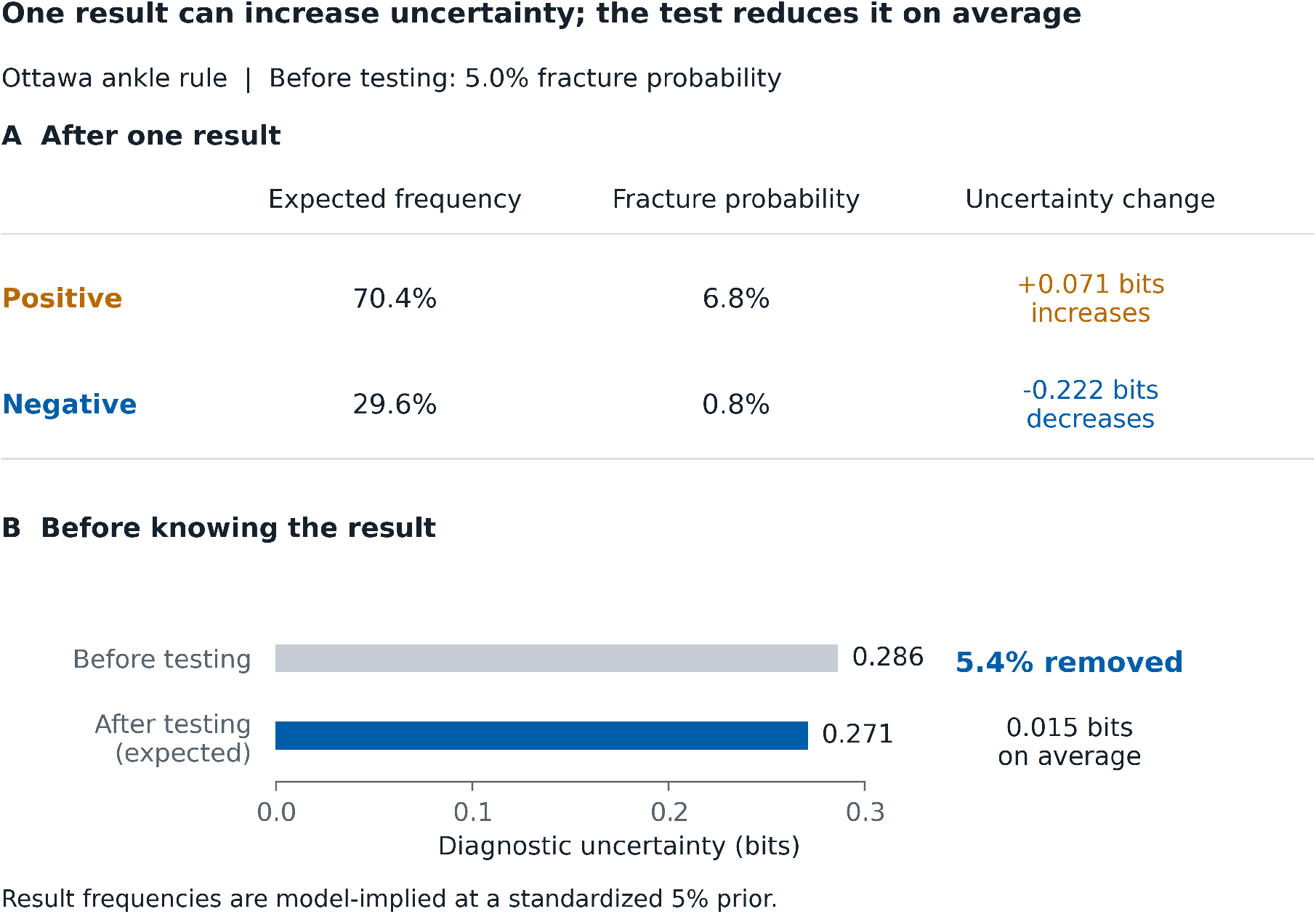
Individual outcomes and expected uncertainty for the Ottawa ankle rule at a 5% starting probability. (A) A positive result has probability 70.4%, raises fracture probability to 6.8%, and increases uncertainty by 0.071 bits. A negative result has probability 29.6%, lowers fracture probability to 0.8%, and reduces uncertainty by 0.222 bits. (B) Weighting the two conditional entropies by their result probabilities gives 0.271 bits of expected remaining uncertainty, compared with 0.286 bits before testing. The expected reduction is 0.015 bits, or 5.4% of starting uncertainty. An individual result can increase uncertainty while the test reduces it on average. Values use the primary pooled estimate from 16 studies; they describe a standardized 5% starting probability. Differences and percentages are calculated before rounding. Result frequencies are implied by the pooled sensitivity and specificity at this assumed prior, not observed frequencies in a 5% prevalence cohort. Uncertainty change means posterior entropy minus prior entropy; its weighted mean is minus mutual information. The negative branch supplies 80.1% of probability-weighted posterior-to-prior KL gain; this share does not partition entropy reduction. KL denotes Kullback-Leibler divergence.

Under this model, negative results are less frequent but supply 80.1% of expected information, measured as their share of probability-weighted posterior-to-prior KL gain; this is not their share of entropy reduction. The likelihood ratios are 1.38 for a positive result and 0.15 for a negative result.[17] Signed changes in log odds are reported separately in supplement S9. Choosing an action also requires consequences and patient preferences.[18,19]

## Discussion

### Principal findings

This study advances diagnostic information as an EBM measure through two linked contributions: interpretable reporting of expected learning and an empirical reference for its magnitude. Across 273 included pooled profiles from 210 reviews, the median profile resolved 29.2% of uncertainty at the standardised 20% starting probability (0.211 of 0.722 bits). This locates a profile within the collected evidence at a stated probability.

### What diagnostic information adds to EBM

Existing measures answer different questions (box 2): operating performance, discrimination, the evidence in an observed result, the quality of probability predictions, and the value of acting. Diagnostic information answers the missing prospective question: how much uncertainty about the target is the evidence expected to resolve before its value is known?

The HINTS comparison shows why nearly equal Youden’s J values can conceal different expected clarification at the stated starting probability. The CEA case gives the practical reason for stating that probability: the lower threshold removed more uncertainty at 20%, the higher threshold more at 5%.[13] The Ottawa ankle rule gives the patient-level distinction: mutual information describes what a test is expected to teach before testing, and result-specific measures describe what one patient learns after a result. Information measures learning; clinical value additionally depends on consequences, costs, available actions, and patient preferences.[18,19]

### Using diagnostic information

Expected information supplies a criterion for comparing candidate tests, thresholds, or model inputs when the objective is diagnostic learning. Compare alternatives for the same target and clinical context, using the evidence already available; a larger value means greater expected clarification under those assumptions. Test burdens and clinical consequences still inform the choice.

Diagnostic reviews should report the target condition, population and setting, result definition, sensitivity and specificity with their source, starting probability, expected information in bits, percentage of starting uncertainty resolved, and the uncertainty method. Report a profile over relevant starting probabilities; values at 5%, 20%, and 50% permit comparison with this collection. At 20%, 29.2% is its median, 45.6% its 75th percentile, and 58.5% its 90th percentile. These percentiles locate an estimate within the collected profiles. An interactive map of all 273 estimates and a calculator are at https://diagnosticuncertainty.org.

### From diagnostic tests to diagnostic models

Diagnostic information can describe conventional tests and model outputs in the same language.[20] The expectation extends to several result levels, including output bands and indeterminate results (supplement S9). To quantify what a new test adds after history or previous tests *X* = *x*, both *P* (*D* ∣ *X* = *x*) and *P* (*T* ∣ *D, X* = *x*) are needed.[2] An individual prior alone cannot account for dependence between tests: substituting marginal sensitivity and specificity also assumes their transport to that patient context. The atlas uses pooled binary profiles at standardised priors; the general measure requires neither binary results nor conditionally independent tests. Model-implied entropy describes target-state uncertainty, while calibration and proper scoring require comparison with observed outcomes.

### Strengths and limitations

Each defined test or threshold for a stated target, setting, population, and reference standard was a separate estimate; one prespecified model was applied throughout, fixed starting probabilities standardised comparisons, and equal weighting defined the empirical reference distribution. The primary 29.2% median at 20% changed to 30.6% with zero-cell-only correction and 34.8% with the regularised direct-binomial estimator. The reference describes the reproducibly accessible review sample; estimating how often tests, conditions, or patients occur in practice would require a separately designed sample. Incomplete clinical context and risk-of-bias reporting limit subgroup interpretation.

### What remains to be tested

Three questions need distinct evaluations. First, can prior probabilities and result distributions be estimated accurately and transported across populations, settings, and reference standards? Assess calibration and out-of-sample predictive performance, including dependence on previous evidence. Second, does reporting bits and percentage resolved improve readers’ understanding of expected versus observed information? Test comprehension against conventional summaries. Third, do strategies that use information to choose investigations improve diagnostic accuracy, test burden, time to diagnosis, or patient outcomes? Compare complete strategies with costs and consequences specified; the present atlas does not establish their clinical performance.

### Conclusion

Diagnostic information fills a missing place in EBM: a common measure of how much uncertainty evidence is expected to resolve at a stated starting probability. The empirical scale demonstrates its use across diagnostic tests and provides a foundation for extending the same language to diagnostic models and other medical evidence.

## Supporting information

Supplementary methods, tables, figures, source records, and sensitivity analyses

Executable analysis code, derived inputs, source-acquisition records, tests, and release metadata

## Data Availability

The Cochrane DTA Reference Dataset is available at https://zenodo.org/records/1303259 under CC BY-NC 4.0. Modern source packages remain available through their published Cochrane reviews subject to Cochrane’s terms; raw packages are not redistributed. The interactive map and calculator are at https://diagnosticuncertainty.org. The scrubbed analysis code, derived analysis inputs, source-acquisition records, tests, and release metadata are available at https://github.com/EMAI-Research/diagnostic-information.

https://zenodo.org/records/1303259

https://doi.org/10.1002/14651858.CD014911.pub2

https://doi.org/10.1002/14651858.CD015089.pub2

## Declarations

### Ethics approval

This study used published aggregate data and did not involve identifiable human participants. Institutional ethics approval was not required.

## Patient and public involvement

Patients and the public were not involved in the design, conduct, reporting, or dissemination planning of this secondary methodological analysis.

## Contributors and sources

SH is an emergency physician and health data researcher. The authors bring perspectives from emergency medicine (SH, JWJ, CK, AM, ASR, JNG), pulmonary and critical care (BWL), medicine and preventive medicine (DML), internal medicine (CR), and health data analytics (AG, PS). SH conceived the study, assembled the review data, specified and performed the analyses, interpreted the findings, created the figures, and wrote the article. BWL, JWJ, DML, CR, AG, PS, CK, AM, ASR and JNG contributed to interpretation and critical revision. Sources were diagnostic systematic reviews with accessible study-level 2×2 tables; common eligibility rules and acquisition records are described in the supplement. OpenAI Codex assisted with code, document preparation, numerical checks, source retrieval and language editing under author review. SH is the guarantor, had full access to all study data and statistical outputs, and controlled the decision to publish.

## Transparency statement

SH, the guarantor, affirms that this manuscript is an honest, accurate, and transparent account of the study reported; that no important aspects of the study have been omitted; and that any discrepancies from the planned analysis have been explained.

## Data and code availability

The Cochrane DTA Reference Dataset is available at https://zenodo.org/records/1303259 under CC BY-NC 4.0. Modern source packages remain available through their published Cochrane reviews subject to Cochrane’s terms; raw packages are not redistributed. The interactive map and calculator are at https://diagnosticuncertainty.org. The analysis-code supplement includes the executable methods, derived analysis inputs and source-acquisition records.

