## Supplementary methods, tables, figures, source records, and sensitivity analyses for "The information in diagnostic tests"

This appendix documents the fixed evidence base, source validation, estimator and numerical checks, complete sensitivity analyses, exploratory within-review contrasts, expanded CEA and mathematical checks, and result-branch distributions. All main claims use the primary continuity-corrected bivariate restricted maximum likelihood (REML) model unless explicitly labelled otherwise.

### Contents

S1 Evidence sources and selection; S2 Source fidelity and author audit; S3 Information distributions; S4 Model and numerical checks; S5 Estimator sensitivity; S6 Within-review contrasts; S7 Near ties and study overlap; S8 CEA and mathematical comparisons; S9 Observed-result information; S10 Hypothetical entropy illustration; S11 Review influence; S12 Reproducibility and protocol history.

### S1. Evidence sources and selection

The protocol was registered with ConductScience (CSR-DI-2026-001; https://doi.org/10.55157/csr.di.2026.001). This section explains how accessible structured evidence yielded the 273-profile reference sample.

| Source | Documented activity | Date |
| --- | --- | --- |
| Cochrane review datasets | Fixed 2018 corpus and two held packages | 2026-08-16 |
| OSF public deposits | Recorded deposit queries | 2026-08-17 |
| Zenodo public deposits | Recorded deposit queries | 2026-08-17 |
| Figshare public deposits | Recorded deposit queries | 2026-08-17 |
| Dryad public deposits | Recorded deposit queries | 2026-08-17 |
| SRDR+ public deposits | Recorded source inventory | 2026-08-17 |
| PubMed Central open-access review tables | Fixed query, 2020-2026 publications | 2026-08-17 |

The analysis-code supplement contains the complete recorded queries, acquisition limits and review inventories. Dates describe documented acquisition or audit activity; the first download date of already-held Cochrane packages was not recorded.

We assembled structured diagnostic accuracy evidence through three complementary routes: fixed Cochrane diagnostic accuracy sources, public review deposits, and machine-readable review tables identified through a fixed PubMed Central query. Dryad and SRDR+ were also audited and contributed no primary pooled estimates. Route-specific discovery counts describe different units and are therefore shown by source without an overall sum.

| Source | Acquisition role | Route-specific records or projects screened | Reviews represented after parsing |
| --- | --- | --- | --- |
| 2018 Cochrane corpus | Reference dataset | 63 | 63 |
| Recent Cochrane reviews | Format validation | 2 | 2 |
| OSF | Review data search | 1,152 | 10 |
| Zenodo | Review data search | 799 | 5 |
| Figshare | Review data search | 262 | 2 |
| Dryad | Review data search | 11 | 0 |
| SRDR+ | Review data search | 172 | 0 |
| PubMed Central | Search of structured review tables | 1,518 | 300 |

Each review record could contain one or more source-defined test, target, threshold, rule, or analysis definitions. Each definition linked to study results with an explicit identifier and non-negative integer counts for true positive, false positive, false negative, and true negative results. These four counts yield the study sensitivity and specificity needed for bivariate pooling and subsequent information calculation.

Of 1,830 assessable candidate groups, 444 met the requirements for modelling. A group required at least five unique study identifiers, one row per identifier, at least three studies with both target-positive and target-negative denominators, non-negative 2×2 counts, aggregate sensitivity plus specificity of at least one in the source-defined orientation, and removal of exact duplicate study-result rows. The structural exclusions were 1,219 groups with fewer than five unique studies, 155 with repeated identifiers, 11 below chance in the source-defined orientation, and one with fewer than three studies containing both disease states. All 444 model fits converged.

Rules for selecting the primary profiles were developed during successive source checks and fixed before the final reference analysis. The 2018 Cochrane corpus contributed 83 of 225 model-ready groups linked to review-author-reported pooled findings. The recent Cochrane reviews contributed 9 of 13 analyses labelled as main. OSF, Zenodo, and PubMed Central retained at most one main table per review record using fixed file-name, table-name, row-count, and identifier priorities. Figshare produced no model-ready group. The 171 remaining model-ready alternatives are documented in the selection record: 142 secondary analyses from the 2018 Cochrane corpus, four modern Cochrane subgroup analyses, and 25 public-source alternatives.

| Source | Reviews represented after parsing | Model-ready groups | Primary pooled estimates | Contributing reviews | Final study-result appearances |
| --- | --- | --- | --- | --- | --- |
| 2018 Cochrane corpus | 63 | 225 | 83 | 27 | 1,250 |
| Recent Cochrane reviews | 2 | 13 | 9 | 2 | 201 |
| OSF | 10 | 13 | 8 | 8 | 155 |
| PubMed Central | 300 | 182 | 169 | 169 | 2,432 |
| Zenodo | 5 | 11 | 4 | 4 | 66 |
| Figshare | 2 | 0 | 0 | 0 | 0 |
| Total | 382 | 444 | 273 | 210 | 4,104 |

The 273 selected groups contained 4,127 normalised study-result appearances. One recorded Zenodo workbook rule retained the 22 results from its declared `Master_Extraction` sheet and excluded 23 matching-group results from another sheet. The final analysis therefore contains 4,104 primary-analysis study-result appearances. A study identifier can appear in more than one pooled diagnostic estimate. Pooling occurred separately for each defined test or threshold, and the empirical reference distribution gave each pooled estimate equal weight.

### How the reference sample was selected

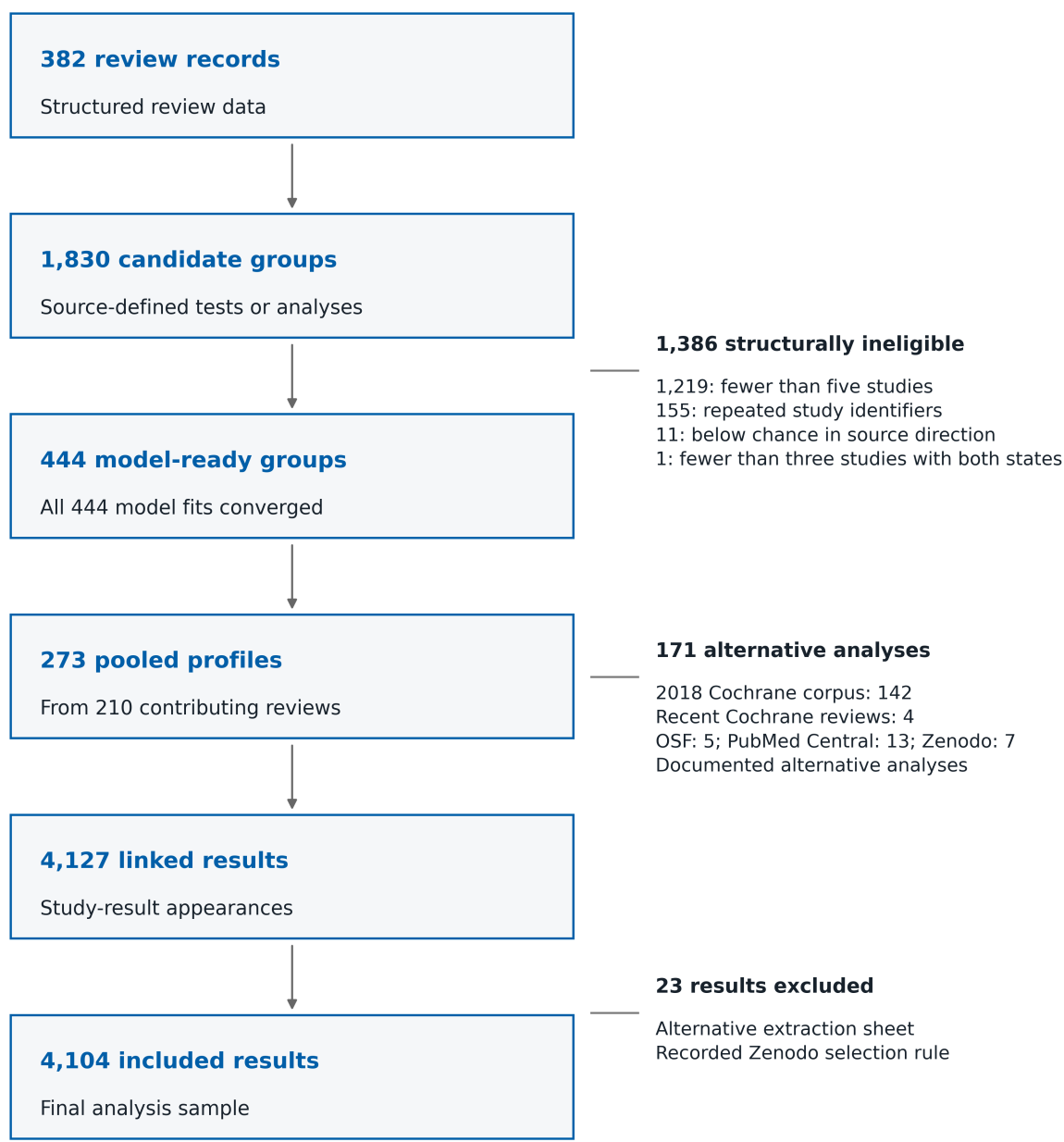

**Figure S1:** Construction of the empirical reference sample. The parsed corpus represented 382 review records and 1,830 assessable source-defined analysis groups. Common structural rules identified 444 model-ready groups, and all models converged. Route-specific primary designation retained 273 pooled diagnostic estimates from 210 reviews. These groups joined to 4,127 normalised study-result appearances; the rule selecting one extraction sheet excluded 23 results, leaving 4,104 primary-analysis study-result appearances. Counts beside each transition give the exact structural and source-route accounting. The diagram describes selection within the accessible source collection, not a comprehensive search of diagnostic medicine.

### S2. Source fidelity, audit denominators, and route stability

We checked whether the reconstructed pooled estimates agreed with source summaries and whether source routes gave similar reference values. The source-summary comparison included 92 pooled diagnostic estimates. Median absolute source-versus-primary differences were 0.021745 for sensitivity and 0.016653 for specificity; maxima were 0.398427 and 0.090881. The recorded author audit found 0/1640 transcription errors and 0/50 acceptance errors. These are fidelity and author-audit results, not independent adjudication or a truth standard.

Source-defined orientation was audited, and every primary row retains its source identity. Figure S2 shows the route-specific medians and interquartile ranges at the 20% prior; these compare accessible source collections, not interchangeable clinical populations.

#### Source agreement and route-specific reference values

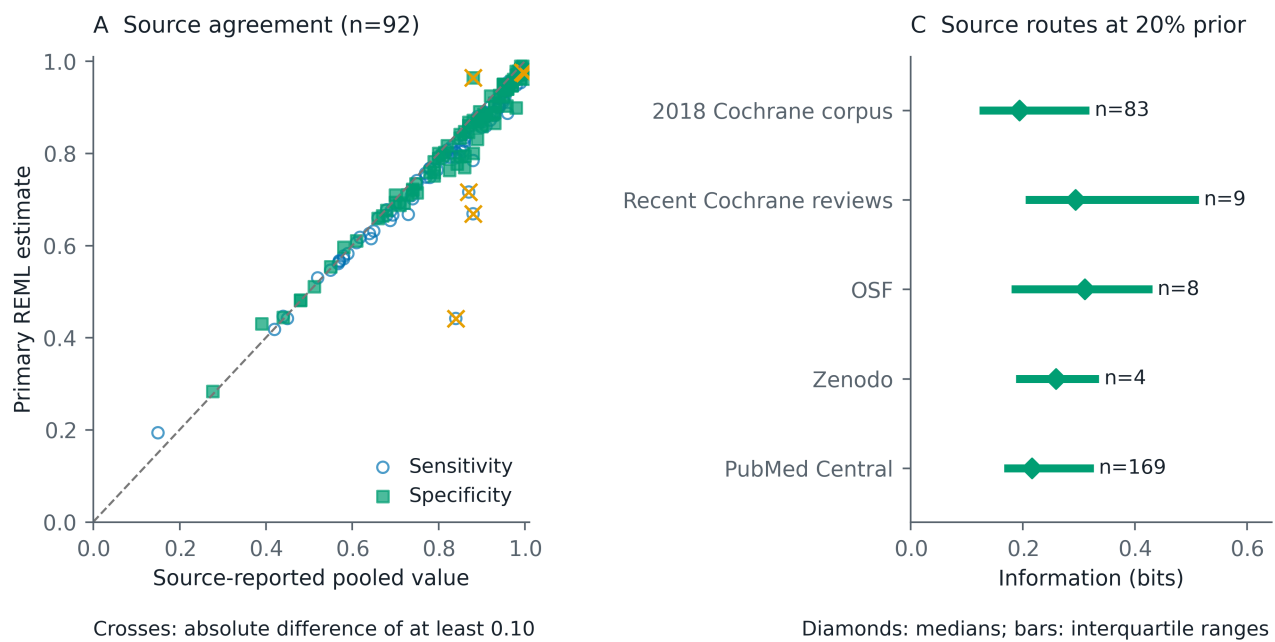

**Figure S2:** Source fidelity, author-audit denominators, and fixed-sample route variation. (A) Source-reported versus primary-analysis pooled sensitivity and specificity for n=92 source-defined test groups; median absolute differences were 0.0217 and 0.0167, maxima were 0.3984 and 0.0909, and 3 points differed by at least 0.10. For CD007394 a 6-versus-7 row mismatch may contribute; the other two cannot be attributed because threshold and pooling metadata are incomplete. This is a fidelity check, not a truth standard. (B) The recorded author audit found 0 transcription errors/1640 cells and 0 acceptance errors/50 decisions; it is not independent adjudication. (C) Route-specific descriptive distributions at 20%, with pooled-estimate n for each of 5 routes. Route summaries describe this sample only, not reproducibility or representativeness.

#### S3. Full information distributions and concordance

To show the spread behind the reference medians, we examined the complete distributions and their relation to Youden's J. The three standard anchors contain the same 273 pooled diagnostic estimates. At 5%, 20%, and 50%, median expected information was 0.067, 0.211, and 0.314 bits. Spearman correlations with Youden's J were 0.954, 0.975, and 0.992. These are fixed-sample descriptive distributions.

### Expected-information distributions and concordance with Youden's J

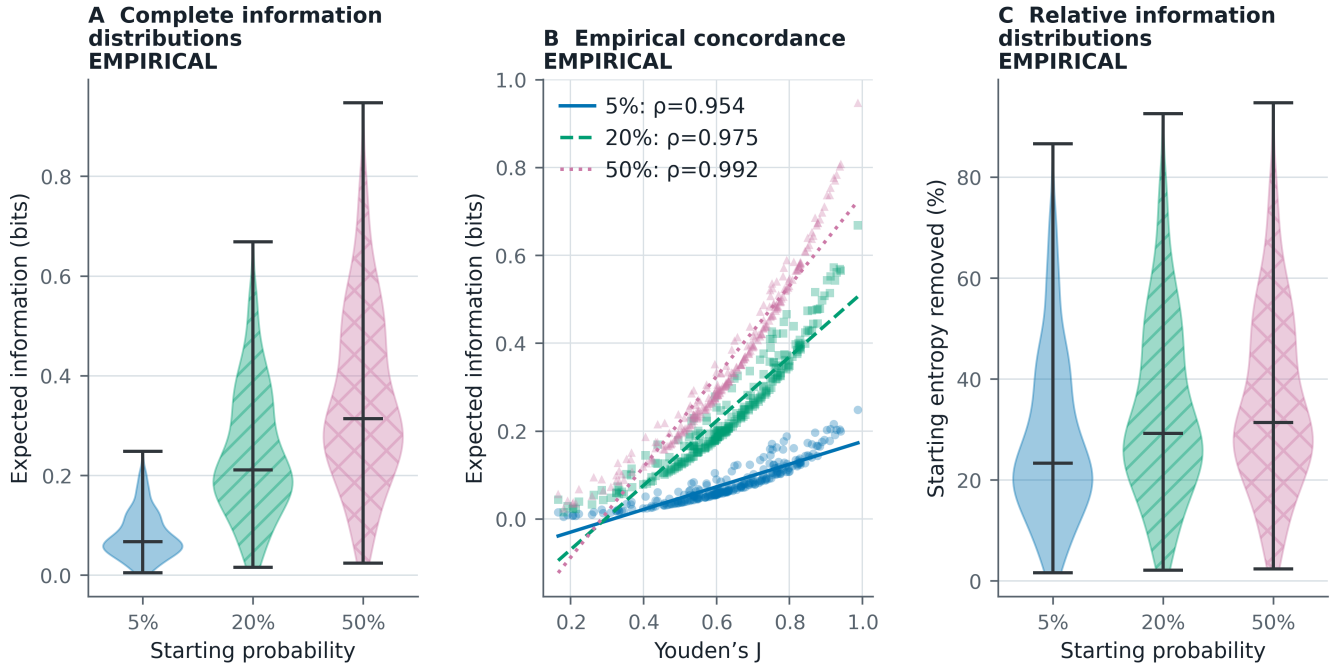

Each anchor contains all 273 primary operating points. Concordance does not imply mathematical equivalence.

**Figure S3:** Fixed-sample expected-information distributions and empirical concordance with Youden's J across 5%, 20%, and 50% starting probabilities. Each anchor contains $n=273$ source-defined meta-analytic test groups. Median expected information was 0.067, 0.211, 0.314 bits, and Spearman $\rho$ was 0.954, 0.975, 0.992. Correlations are descriptive and sample-composition dependent; shared sensitivity and specificity inputs do not guarantee them.

#### S4. Primary model, uncertainty, recovery, and diagnostics

We examined numerical convergence and limited recovery under the common pooling model. The primary continuity-corrected bivariate REML model added 0.5 to all four cells before calculating logits and sampling variances. Deterministic multistart optimisation selected the lowest valid recomputed objective. Pooled-mean intervals used the estimated mean covariance. Predictive ranges added fitted between-study covariance. Both are plug-in summaries conditional on fitted heterogeneity; heterogeneity-parameter uncertainty was not propagated.

The limited recovery exercise used one generated dataset in each of 24 scenarios. Pooled-mean intervals included the generating sensitivity and specificity in 87.5% and 87.5% of scenarios. This is a limited numerical check and cannot estimate repeated-sampling coverage. Fit diagnostics flagged 69 correlations with absolute value at least 0.99, 9 near-singular covariance matrices, 1 gradients above $10^{-4}$, and 77 fits in the union of numerical flags.

### Primary REML limited recovery and fit diagnostics

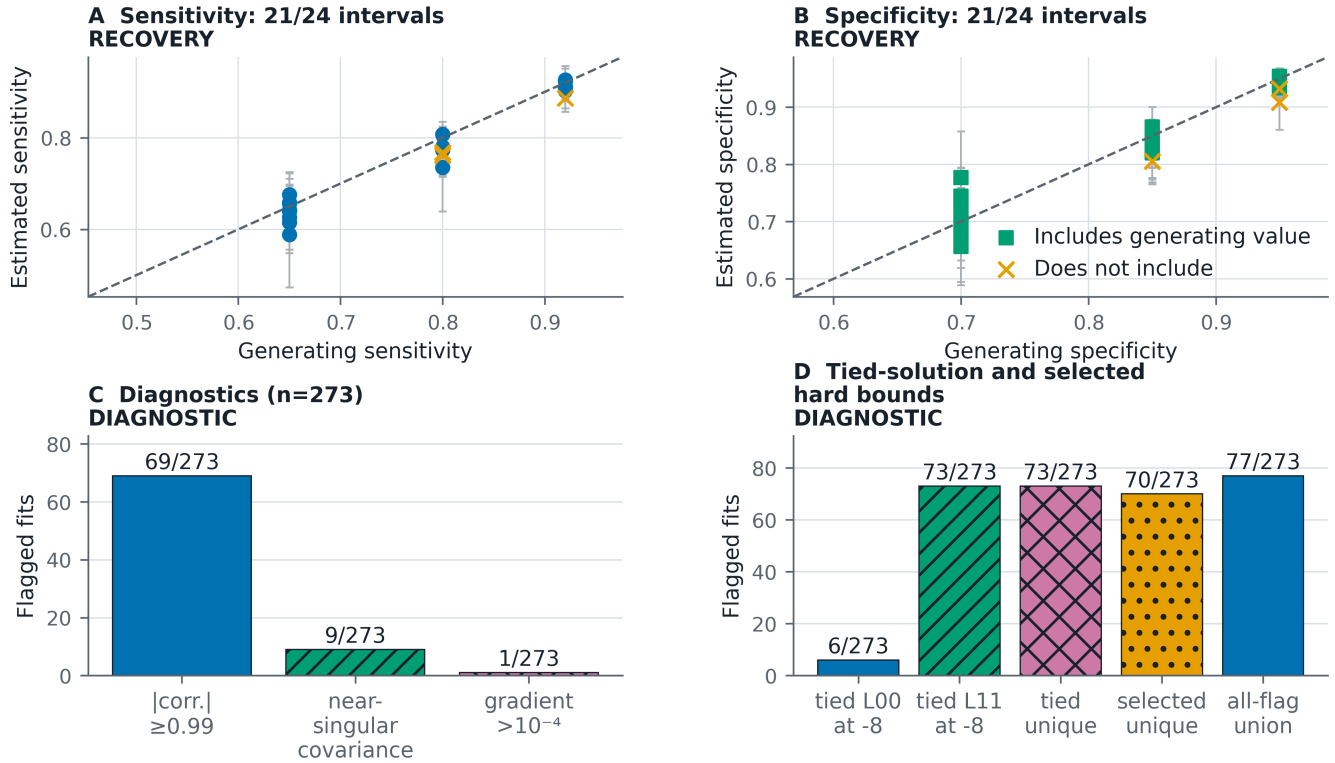

Se = sensitivity; Sp = specificity. 24 scenarios, one dataset each: limited numerical check, not repeated-sampling coverage. Approximate plug-in intervals condition on fitted heterogeneity; predictive ranges were not checked.

**Figure S4:** Limited recovery and numerical diagnostics for the primary continuity-corrected bivariate restricted maximum likelihood (REML) model. The recovery exercise is a limited numerical check, not evidence that the intervals have 95% repeated-sampling coverage. (A–B) In 24 scenarios with one dataset each, pooled-mean intervals included the generating sensitivity and specificity in 87.5% and 87.5%. These are approximate plug-in summaries conditional on fitted heterogeneity; heterogeneity-parameter uncertainty was not propagated and predictive ranges were not checked. (C) 69 fits had an absolute between-study correlation of at least 0.99, 9 had near-singular covariance estimates, and 1 had a raw optimiser gradient above $10^{-4}$. (D) Numerical ties included 6 sensitivity-heterogeneity and 73 specificity-heterogeneity lower-bound endpoints across 73 fits; selected endpoints included 6 and 70, respectively. A tied boundary signals fragility even when the selected endpoint is interior. Seven fits with tied boundary solutions were identified only by comparing optimisation endpoints, bringing the total with numerical flags to 77. Appendicitis had a tied specificity-heterogeneity boundary solution and selected correlation 1.000; its joint predictive ranges are fragile.

### S5. Estimator and correction sensitivity

We tested how estimator and continuity-correction choices affected the information values. The regularised direct-binomial model was used for sensitivity analysis. At the 20% anchor, like-for-like plug-in information differed from primary REML by median 0.033731 and maximum 0.195360 bits. The comparison combines likelihood family, regularisation or prior, and continuity-correction choices. It does not isolate a single causal modelling component.

Applying the continuity correction only to zero-containing studies moved at least one central quantity in 269 pooled estimates. Median and maximum information differences were 0.008617 and 0.028935 bits.

### Like-for-like plug-in central estimates and analysis-family sensitivity

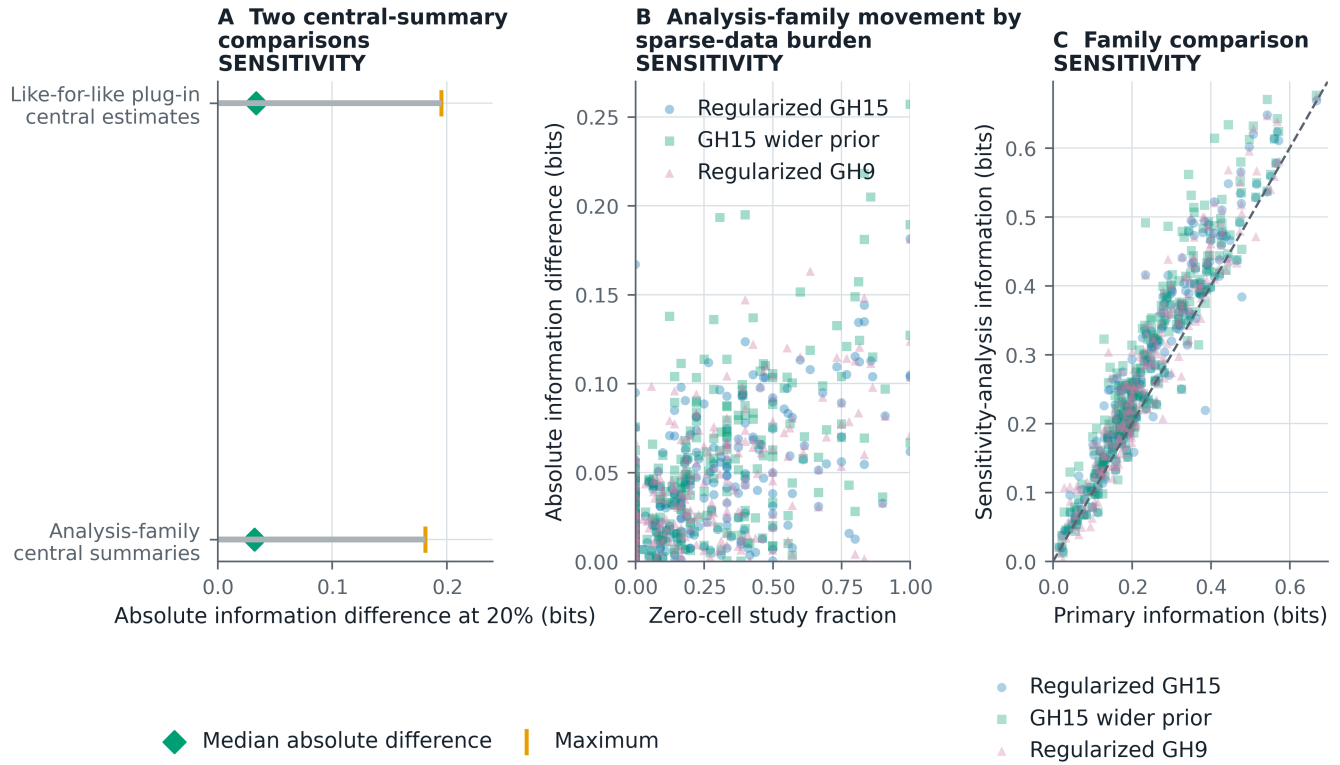

**Figure S5:** Sensitivity of expected information to analysis choices. These comparisons show movement across analysis families, not an effect attributable to any single modelling choice. (A) At 20%, plug-in expected information evaluated at each analysis's central sensitivity and specificity differed between REML and the 15-node Gauss-Hermite (GH15) direct-binomial fit by median 0.0337 and maximum 0.1954 bits. This comparison combines likelihood family, regularisation or prior, and continuity-correction choices. The comparison with the GH15 propagated median was 0.0321 and 0.1814 bits, but those are different central-summary estimands. (B–C) Point-level GH15, wider-prior GH15, and 9-node Gauss-Hermite values are sensitivity summaries, not primary-analysis intervals.

### S6. Exploratory within-review contrasts

To examine information differences between profiles from the same review, we evaluated all 49 preclassified within-review contrasts, including 32 sensitivity-specificity trade-offs. These comparisons are descriptive. Same-participant cells were established for 0 contrasts, cross-test covariance was available for 0, and paired inference was supported for 0. No paired interval or superiority probability was constructed.

### Exploratory within-review contrasts: magnitude and analysis-family movement

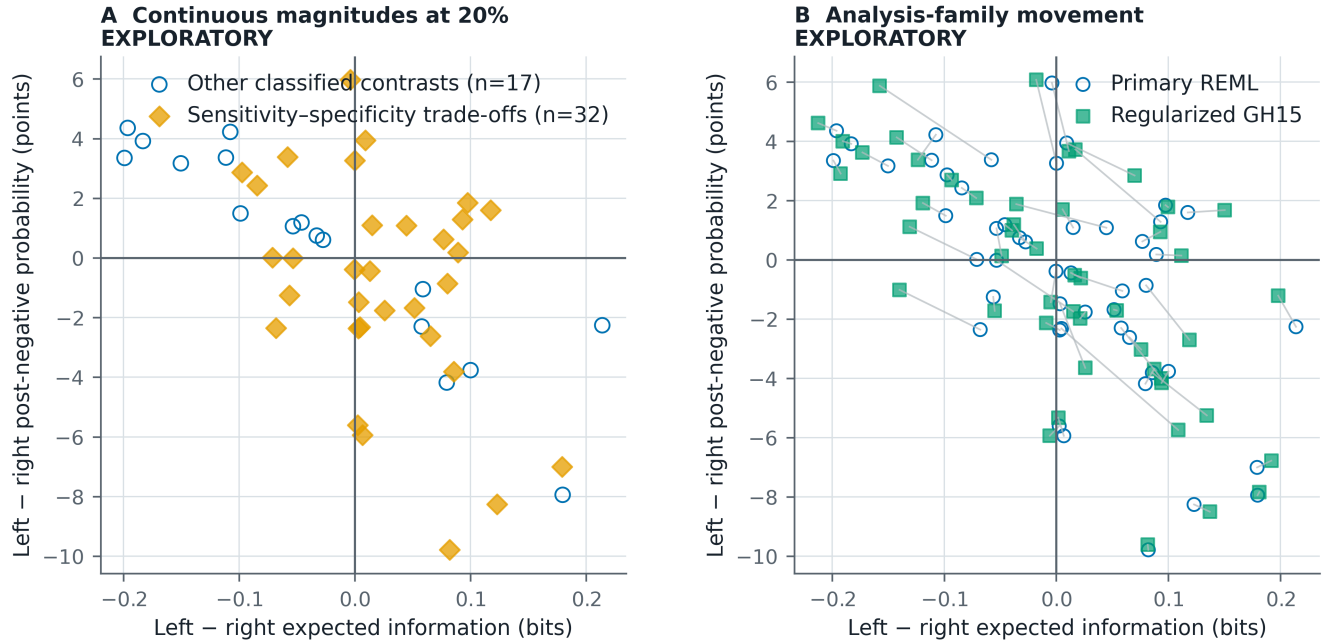

All 49 contrasts are descriptive: same-participant cells and cross-test covariance are unavailable, so no paired inference is shown.

**Figure S6:** Exploratory within-review magnitudes and analysis-family movement at the 20% anchor. (A) Signed differences in expected information and post-negative probability for all 49 preclassified contrasts, including 32 sensitivity-specificity trade-offs. (B) Movement from primary REML plug-in summaries to GH15 propagated-central summaries compares different estimands and must not be attributed purely to model family. Same-participant cells and cross-test covariance were available for 0 contrasts; no paired inference or superiority claim is shown.

#### S7. Near ties and study-composition restriction

We asked whether near ties in accuracy could conceal information differences, including after restricting comparisons by overlapping study identifiers. Near-tie thresholds are descriptive tolerances rather than clinical equivalence margins. Overlapping study identifiers do not establish common participants, paired outcomes, or covariance. The study-composition-restricted display contains 23 contrasts with at least three overlapping study identifiers.

### Near ties and study-composition-restricted audits

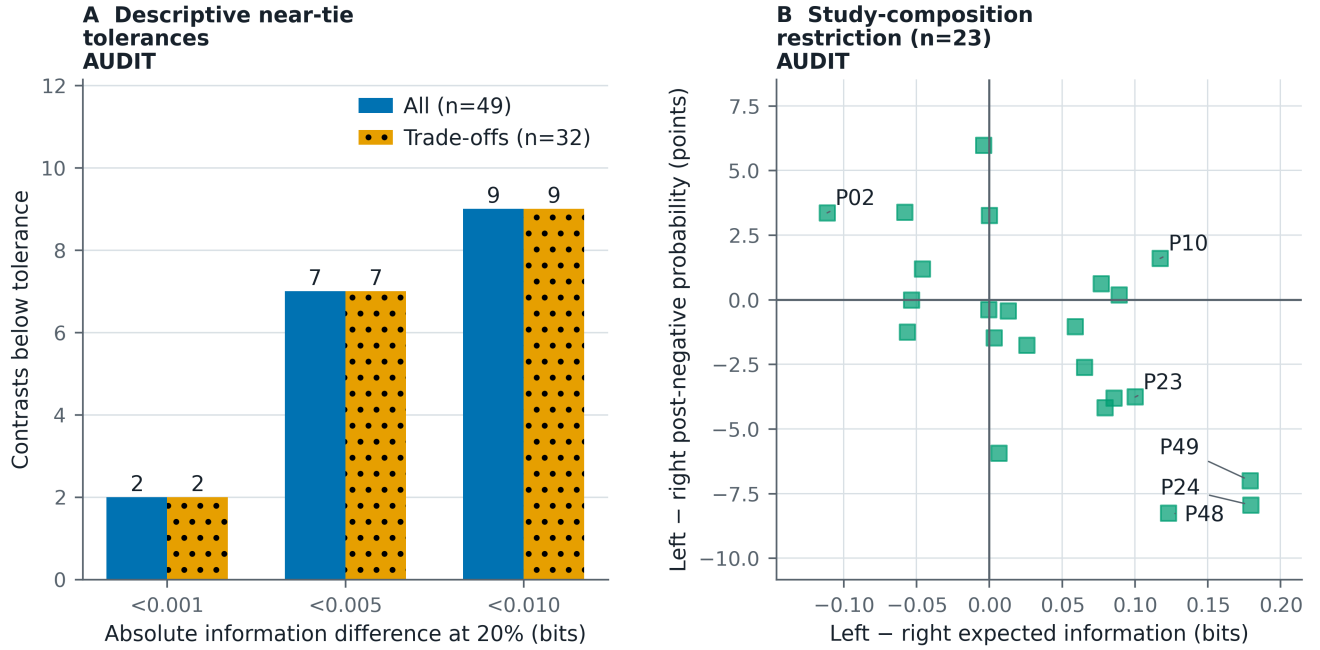

Near-tie thresholds are descriptive tolerances, not clinical equivalence margins. Overlapping study identifiers do not establish common participants or paired outcomes.

**Figure S7:** Near-tie and study-composition-restricted audits at the 20% anchor. (A) Counts below absolute information tolerances 0.001, 0.005, 0.010 bits among all 49 contrasts and the trade-off subset. These are descriptive tolerances, not clinical equivalence margins. (B) Continuous magnitudes for 23 contrasts with at least three overlapping study identifiers. Direct labels identify the six largest displayed magnitudes: P02, Typhidot versus TUBEX; P10, two frozen-section positivity definitions; P23, microhaematuria versus proteinuria; P24, microhaematuria versus leukocyturia; P48, head impulse versus test of skew; and P49, nystagmus versus test of skew. Identifier overlap does not establish common participants, paired outcomes, or cross-test covariance.

### S8. Expanded CEA and mathematical checks

We examined whether the CEA information crossing persisted under alternative synthesis choices. The CEA threshold profiles used the same review evidence.[w1] The primary REML curves crossed at 12.8% and the regularised direct-binomial curves crossed at 38.1%. Other model and numerical specifications are shown when a root existed. These probabilities describe model-specific expected-information ordering. They are not action thresholds or clinical-superiority estimates.

The exact mathematical checks gave an information difference of 0.010414 bits for equal-J diagnostic profiles, an information difference of 0.000090 bits with a 10.8 percentage-point post-negative difference for the near-equal-information pair, and a synthetic ordering crossing at 28.6%.

### Empirical CEA crossing sensitivity and additional theoretical checks

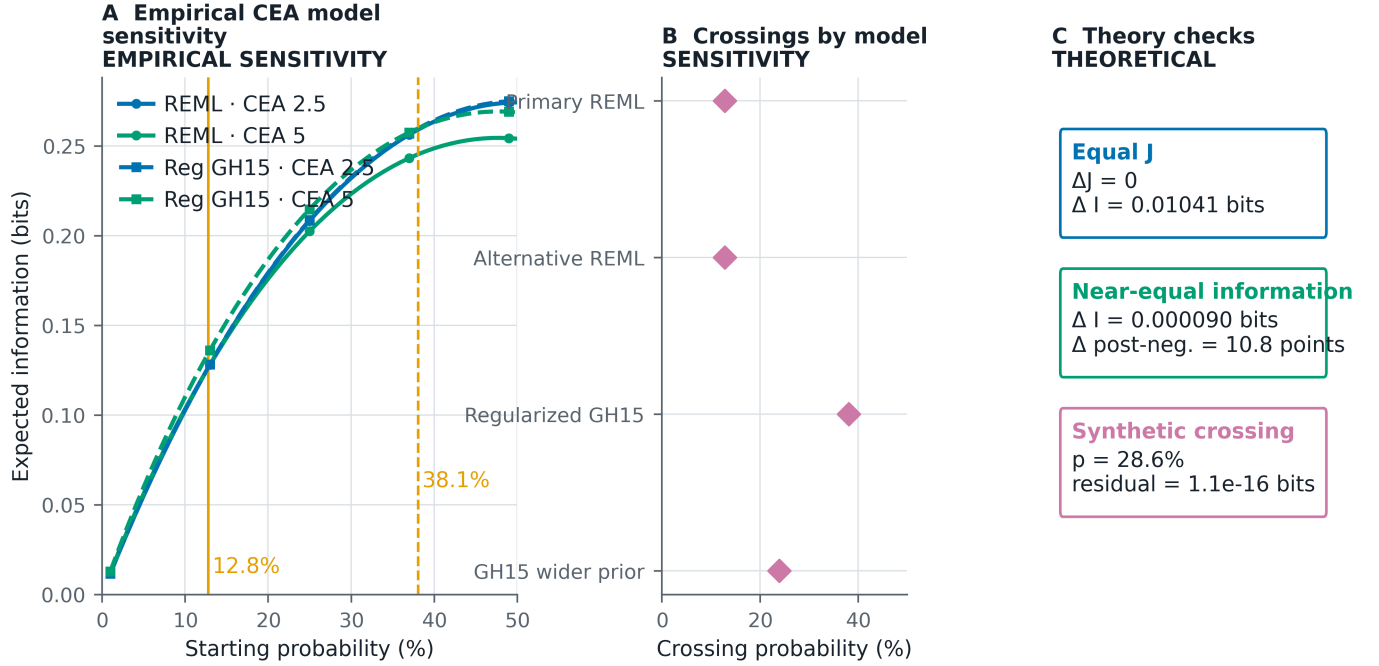

CEA crossings depend on the synthesis model. None of the displayed probabilities is an action threshold.

**Figure S8:** Empirical carcinoembryonic-antigen (CEA) crossing sensitivity and additional theoretical checks. (A–B) The empirical expected-information crossing was 12.8% under primary restricted maximum likelihood (REML) and 38.1% under the regularised direct-binomial model using 15-node Gauss-Hermite quadrature (GH15); other model and numerical sensitivities are shown where a root exists. (C) Independent recalculations verify equal-J, near-equal-information, and synthetic ordering-crossing examples. The CEA example uses published review data and is model specific. None of the displayed probabilities is an action threshold.

### S9. Result branches across all pooled diagnostic estimates

These analyses distinguish what a test is expected to teach from the uncertainty change after one result. At a 5% starting probability, a positive result increased binary uncertainty for 273/273 pooled estimates; at 20%, this occurred for 232/273. At 50%, the count was 0/273 because a result cannot move the probability closer to 50%. Negative results increased uncertainty for 0, 0, and 0 estimates at the three anchors. These branch changes describe whether uncertainty narrowed or widened after one result. For entropy reduction, defined as prior entropy minus posterior entropy, the probability-weighted average is mutual information. The tabulated and plotted entropy changes use posterior minus prior, whose weighted average is minus mutual information.

For disease state $D$, result $T = t$, and patient profile $X = x$, result-specific information gain is

$$G(t; x) = D_{KL} [P(D | T = t, X = x) \| P(D | X = x)].$$

Its probability-weighted average is the patient-conditioned mutual information:

$$I(D; T | X = x) = \sum_t P(T = t | X = x) G(t; x).$$

Entropy reduction $R(t; x) = H(D | X = x) - H(D | T = t, X = x)$ can be negative, whereas $G(t; x)$ is non-negative. Both average to $I(D; T | X = x)$. The tabulated entropy change is $-R(t; x)$. Thus a positive result can increase entropy while still providing positive Kullback-Leibler (KL) gain.

For the Ottawa ankle rule at the standardised 5% prior, the negative branch contributes 80.1% of the result-probability-weighted posterior-to-prior KL sum. This share uses $P(T = -)G(-)/I(D; T)$, not the entropy-reduction terms, whose signs can differ. Its likelihood ratios are 1.38 and 0.15; their base-2 logarithms, +0.47 and -2.77 bits, are signed changes in log odds in Good's sense.[w2] Positive values favour disease and negative values favour its absence. These are neither branch entropy reductions nor non-negative KL gains.

The same definitions apply to multilevel results by summing over all result categories. Conditioning on $X = x$ requires the prior and result distributions conditional on that context, including any previous test results; a marginal accuracy estimate is insufficient unless its transport is justified.

#### One result can increase uncertainty

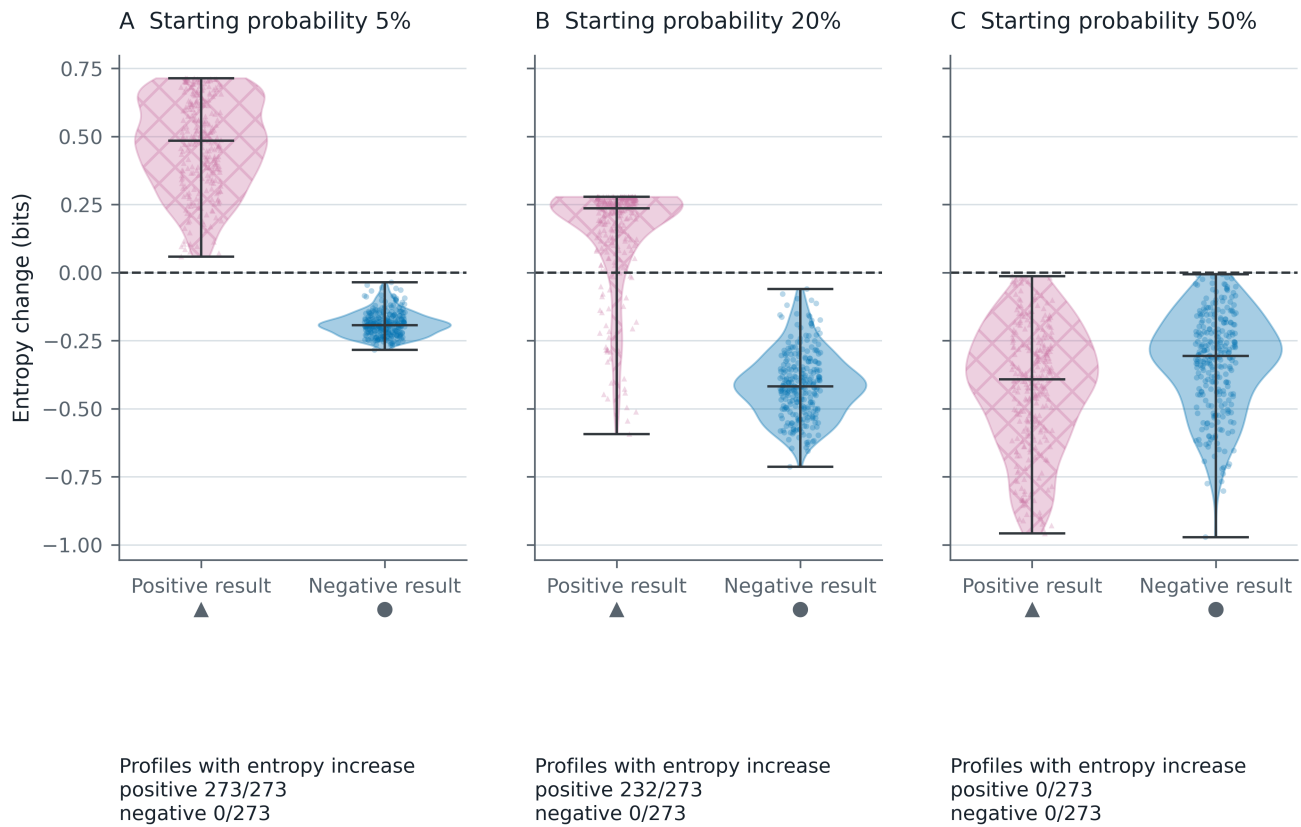

Entropy change = posterior minus prior. Each panel includes all 273 profiles.

**Figure S9:** Observed-result branch entropy across $n=273$ primary pooled diagnostic estimates at 5%, 20%, and 50% starting probabilities. Positive results increased binary entropy for 273/273, 232/273, and 0/273 points, respectively; negative results increased entropy for zero points at every anchor. The violin distributions include all points. Branch movement is not clinical benefit or harm, and expected mutual information remains non-negative. Primary estimator: continuity-corrected bivariate REML. Entropy change is posterior minus prior; negative values indicate uncertainty reduction. Its result-weighted mean is minus mutual information, whereas the weighted mean of posterior-to-prior KL gains is mutual information. KL denotes Kullback-Leibler divergence.

#### S10. Hypothetical entropy-curve illustration

Figure S10 uses the stipulated sensitivity of 90% and specificity of 95% from Box 1, at priors of 20% and 5%. These are teaching inputs, not estimated troponin or Ottawa performance. Bayes' rule supplies the result probabilities and posteriors; binary entropy gives each point's height. The prior equals the result-weighted mean posterior, placing the expected remaining entropy on the chord joining the two posterior points. The vertical gap from the prior on the curve to that average is mutual information.

| Prior | Positive result | Positive posterior | Negative posterior | Expected information | Percentage resolved |
| --- | --- | --- | --- | --- | --- |
| 20.00% | 22.00% | 81.82% | 2.56% | 0.437 bits | 60.57% |
| 5.00% | 9.25% | 48.65% | 0.55% | 0.149 bits | 52.11% |

At 5%, a positive result moves probability towards 50% and raises entropy, yet its posterior-to-prior KL gain is positive. Averaging over both results still reduces entropy. All values were computed at full precision; the plotted averages are not patient outcomes or additional posterior states.

#### Expected learning is the gap between the curve and the chord

Hypothetical test: sensitivity 90%, specificity 95%

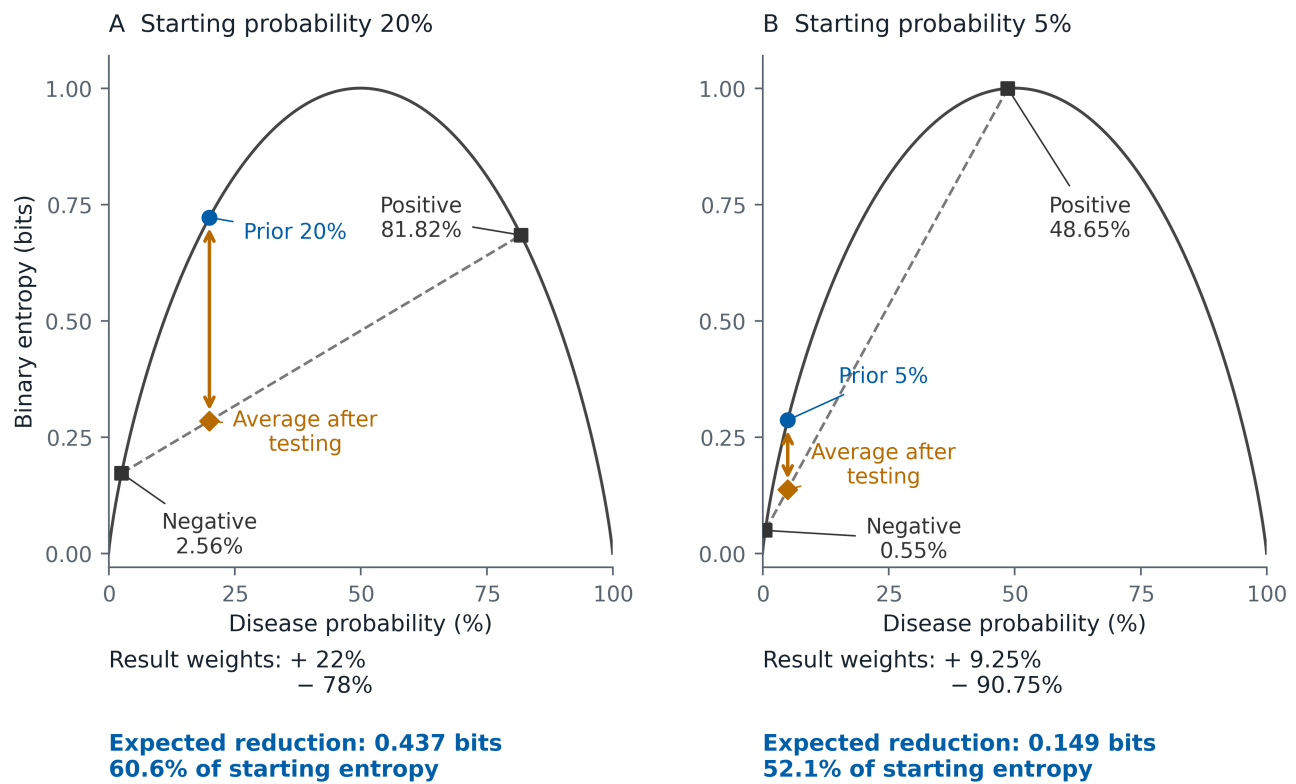

The diamond is a weighted average, not an additional possible result.

**Figure S10:** How expected diagnostic information appears on the binary entropy curve. An original hypothetical illustration uses stipulated sensitivity 90% and specificity 95%, with (A) a 20% prior and (B) a 5% prior. Squares mark posterior states after negative and positive results; the circle marks the prior. The diamond lies on the dashed chord joining the posterior points, at their result-probability-weighted mean entropy and mean probability (which equals the prior). It is an average, not a patient outcome. The vertical gap to the prior on the curve is mutual information: 0.437 bits (60.6%) at 20%, and 0.149 bits (52.1%) at 5%. In B, a positive result moves probability to 48.65%, increasing entropy despite positive KL gain and a positive expected entropy reduction. Axes are linear and identical; weights and values are calculated before rounding. These stipulated examples are distinct from the empirical Ottawa example in Figure 4. KL denotes Kullback-Leibler divergence.

### S11. Review influence and whole-review resampling

We asked whether reviews contributing several profiles materially influenced the reference median, and whether keeping profiles from the same review together changed its interval. Equal-review weighting changed the 20% median from 29.2% to 30.0%; whole-review resampling gave a similar interval (table S1). The median was stable in these checks. All three starting probabilities retained the same 273 profiles from 210 reviews: 186 reviews contributed one profile and 24 contributed several, with a maximum of ten.

For equal-review weighting, each profile received weight $1/n$, where $n$ was its review's number of retained profiles. Each review therefore had total weight one. We computed the weighted median across all profiles, taking the first value whose cumulative weight reached half the total and averaging adjacent supported values at an exact half-weight gap (absolute numerical tolerance $10^{-12}$). This differs from taking a median of review-specific medians.

For whole-review resampling, we sampled 210 reviews with replacement and retained all profiles from each selected review, including repeated copies when a review was selected more than once. Each replicate used the ordinary median across its sampled profiles; equal-review weights were not applied. We used 2,000 replicates, NumPy default\_rng seed 20260913, lexically sorted review identifiers and identical review draws at all three priors. The 95% intervals are the 2.5th and 97.5th percentiles with linear interpolation. The original profile-resampling intervals remain the primary analysis.

**Table S1. Review influence and uncertainty around the empirical reference medians.**

| Prior | Profile median, % | Review-weighted median, % | Difference, pp | Profile bootstrap 95% CI, % | Review bootstrap 95% CI, % |
| --- | --- | --- | --- | --- | --- |
| 5% | 23.3 | 23.6 | +0.3 | 22.0–26.7 | 21.8–27.3 |
| 20% | 29.2 | 30.0 | +0.7 | 27.6–32.7 | 27.5–33.5 |
| 50% | 31.4 | 33.0 | +1.6 | 29.5–34.9 | 29.4–35.2 |

*Table S1 footnote:* All medians and intervals describe the percentage of starting uncertainty resolved. Difference is the equal-review median minus the original equal-profile median, in percentage points (pp), calculated before rounding. Each review has total weight one only in the review-weighted median column. Both bootstrap intervals use ordinary profile medians; the resampling unit is a profile or an entire review, respectively. Whole-review resampling addresses shared review membership, but does not propagate within-profile estimation error or fully account for primary-study overlap between reviews. The included reviews are a reference collection, not a probability sample of all diagnostic medicine.

### S12. Reproducibility and protocol history

The protocol records an internal date of 16 August 2026 (CSR-DI-2026-001; https://doi.org/10.55157/csr.di.2026.001). The public record was verified on 13 September 2026. Evidence acquisition continued after the internal protocol date, with public-repository and PubMed Central searches documented on 17 August. During manuscript revision, we added the review-weighting and whole-review resampling analyses after completing the primary analysis.

Every included study result retains a pooled-estimate identifier and source identity. Group-level URLs were available for 181 pooled estimates; 92 were identified by their source archive or data package. The analysis-code supplement supplies executable methods, software requirements, derived analysis inputs, source-acquisition records and instructions for reproducing the results. Restricted original source packages must be obtained under their source terms.

Information is an expected reduction in binary uncertainty at a stated starting probability. It is not clinical utility, net benefit, or an action rule. The sample is not representative of all diagnostic medicine. Missing thresholds, settings, populations, reference standards, consequences, preferences, costs, and alternatives remain missing.

OpenAI Codex assisted with research code development, document and figure preparation, software testing, numerical consistency checks, source retrieval, and language editing. It did not independently select eligible evidence, adjudicate clinical content, or determine interpretation. SH reviewed all inputs, outputs, and AI-assisted text.

w2. Good IJ. *Probability and the Weighing of Evidence*. London: Charles Griffin; 1950.
