## Supplementary material for "The information in diagnostic tests": Executable analysis code, derived inputs, source-acquisition records, tests, and release metadata: README.md

# The information in diagnostic tests

## Analysis code and supporting data

This supplement provides the methods and derived data used to calculate diagnostic information for 273 pooled profiles from 210 reviews. It accompanies manuscript 1.13.3.

## Reproduce the results

Use Python 3.13. Extract the archive and open a terminal in its folder. Install the listed packages:

    python -m pip install -r requirements.txt

Then run:

    python -m scripts.reproduce_bmj_rmr

The command checks all 273 profiles and 4104 study-result appearances, recalculates information and post-test probabilities at 5%, 20% and 50% starting probabilities, reproduces both bootstrap analyses and the equal-review medians, and refits three primary models. It writes the results to the reproduced folder.

To refit all 273 primary models and reproduce all fourteen figures in colour and grayscale, run:

    python -m scripts.reproduce_bmj_rmr --refit all --figures --output reproduced-all

The alternative estimators and their numerical results are also included. The commands above verify the primary models and reported reference summaries; they do not refit every alternative estimator.

## Contents

- The scripts folder contains the calculations, source readers and figure code.
- The analysis folder contains the derived inputs, pooled estimates, source identifiers, search records and supplementary results.
- The styles folder contains figure settings.
- The source_licenses.csv file records the available source identities and licence information.
- The CITATION.cff file supplies the article title and author list for citation software.
- The MANIFEST.json file lists the size and SHA-256 checksum of each file.

The 4104 rows are study-result appearances. A study can contribute to more than one diagnostic profile. They are not a count of unique studies or participants. The acquisition-record.json file in analysis/bmj_rmr_v1131 contains the recorded search queries, dates and source coverage.

## Source access and reuse

The Cochrane DTA Reference Dataset is available under CC BY-NC 4.0 at https://zenodo.org/records/1303259. Other source materials retain their respective terms. Original restricted Cochrane packages and full article texts are not included; obtain them from the cited sources when rerunning source extraction. The supplied derived inputs support the reproduction commands above without those original packages. Redistribution of this supplement does not override the rights in third-party source material.

The registered protocol is <https://doi.org/10.55157/csr.di.2026.001>.

The interactive map and calculator are at <https://diagnosticuncertainty.org>.
