## Supplementary material for "The information in diagnostic tests": Executable analysis code, derived inputs, source-acquisition records, tests, and release metadata: bmj_rmr_v042.py

```
"""Deterministic analysis and release support for BMJ RMR v0.42.0.

The script is intentionally phase-aware. Each phase writes machine-readable
outputs and a QA gate so later manuscript work can consume locked results
without reopening earlier decisions.
"""

from __future__ import annotations

import argparse
import hashlib
import json
import math
import subprocess
import zipfile
from pathlib import Path

import numpy as np
import pandas as pd
from scipy.optimize import brentq
from scipy.special import expit, logit
from scipy.stats import norm

try:
    from scripts.cochrane_dta_atlas import (
        diagnostic_metrics,
        SEED as REML_SEED,
        analyze_group,
        fit_bivariate_reml,
    )
except ModuleNotFoundError:  # supports `python scripts/bmj_rmr_v042.py`
    from cochrane_dta_atlas import (
        diagnostic_metrics,
        SEED as REML_SEED,
        analyze_group,
        fit_bivariate_reml,
    )


VERSION = "0.42.0"
RELEASE_DATE = "2026-08-25"
ORIENTATION_SOURCE_COMMIT = "49cdbcbe13ab7eda9e3ce220b7e9ae3edc33e4d7"
PRIMARY_KEY = ["source", "review_id", "group_id"]
PHASE3_SOURCE_COMMIT = "857bc0325947b34194dbe1e56728d100e0e0d319"
PHASE3_INITIAL_CANDIDATE_COMMIT = "229b58c5282e2ff26e73049bf42c197923646532"
STANDARD_ANCHORS = (0.05, 0.20, 0.50)
PROFILE_PROBABILITIES = tuple(np.linspace(0.01, 0.50, 50))
DERIVED_DRAWS = 20_000
APPENDICITIS_ID = (
    "pmc open access:PMC12734481:medicina-61-02163-t001:"
    "table-1-summary-of-the-characteristics-of-the-included-studies:1:overall"
)
CEA_LOW_ID = "legacy:CD011134:2"
CEA_HIGH_ID = "legacy:CD011134:3"
LANDMARKS = (
    (APPENDICITIS_ID, "Non-contrast CT appendicitis"),
    (
        "pmc open access:PMC9502997:Tab2:"
        "table-2-diagnostic-accuracy-results-from-the-included-studies:1:overall",
        "Ottawa ankle rule",
    ),
    (
        "pmc open access:PMC11563508:table3-1742271X241289726:"
        "table-3-diagnostic-accuracy-results-from-included-studies-with-data-presented-to:1:overall",
        "Lung ultrasound pneumonia",
    ),
    ("legacy:CD009372:1", "CT angiography intracranial lesion"),
)


def file_hash(path: Path) -> str:
    digest = hashlib.sha256()
    with path.open("rb") as handle:
        for block in iter(lambda: handle.read(1024 * 1024), b""):
            digest.update(block)
    return digest.hexdigest()


def bytes_hash(payload: bytes) -> str:
    return hashlib.sha256(payload).hexdigest()


def git_blob(root: Path, commit: str, relative: str) -> bytes:
    return subprocess.run(
        ["git", "show", f"{commit}:{relative}"],
        cwd=root,
        check=True,
        capture_output=True,
    ).stdout


def write_json(path: Path, payload: object) -> None:
    path.parent.mkdir(parents=True, exist_ok=True)
    path.write_text(json.dumps(payload, indent=2, sort_keys=True) + "\n", encoding="utf-8")


def git_value(root: Path, *args: str) -> str:
    result = subprocess.run(
        ["git", *args], cwd=root, check=True, capture_output=True, text=True
    )
    return result.stdout.strip()


def csv_bool(series: pd.Series) -> pd.Series:
    return series.astype(str).str.strip().str.lower().isin({"true", "1", "yes"})


def source_route(source: pd.Series) -> pd.Series:
    mapping = {
        "Cochrane DTA Reference Dataset": "historical Cochrane",
        "Modern Cochrane data package": "modern Cochrane",
        "OSF": "OSF",
        "Zenodo": "Zenodo",
        "PMC Open Access": "PMC",
    }
    return source.map(mapping).fillna(source)


def select_analysis_rows(primary: pd.DataFrame, normalized: pd.DataFrame) -> pd.DataFrame:
    """Apply the manifest's canonical-sheet rule to locked normalized rows.

    Historical Cochrane records do not carry sheet identifiers and retain every
    matching row. External sources with a manifest sheet retain only that sheet.
    This removes 23 noncanonical TwoByTwo_Long rows from one Zenodo group while
    keeping the primary operating-point identity unchanged.
    """

    manifest_fields = PRIMARY_KEY + ["source_sheet", "study_rows"]
    joined = normalized.merge(
        primary[manifest_fields],
        on=PRIMARY_KEY,
        how="inner",
        suffixes=("_row", "_manifest"),
        validate="many_to_one",
    )
    keep = joined["source_sheet_manifest"].isna() | joined["source_sheet_row"].eq(
        joined["source_sheet_manifest"]
    )
    selected = joined.loc[keep].copy()
    counts = selected.groupby(PRIMARY_KEY, dropna=False).size()
    expected = primary.set_index(PRIMARY_KEY)["study_rows"].astype(int)
    aligned = counts.reindex(expected.index)
    if aligned.isna().any() or not aligned.astype(int).equals(expected):
        comparison = pd.concat(
            [expected.rename("manifest_study_rows"), aligned.rename("selected_rows")], axis=1
        )
        raise AssertionError(f"Canonical row selection mismatch:\n{comparison.query('manifest_study_rows != selected_rows')}")
    return selected


def locked_input_paths(root: Path) -> list[Path]:
    relative = [
        "analysis/full_atlas/test_group_manifest.csv",
        "analysis/full_atlas/normalized_study_data.csv",
        "analysis/full_atlas/operating_points_20_percent.csv",
        "analysis/full_atlas/model_validation.csv",
        "analysis/full_atlas/pmc_manual_validation.csv",
        "analysis/full_atlas/orientation_audit.csv",
        "analysis/full_atlas/source_stability.csv",
        "analysis/bmj_rmr_v040/operating_points_regularized.csv",
        "analysis/bmj_rmr_v040/operating_points_wide_prior.csv",
        "analysis/bmj_rmr_v040/pair_continuous_metrics.csv",
        "analysis/bmj_rmr_v040/simulation_recovery.csv",
        "analysis/bmj_rmr_v040_gh9_validation/operating_points_regularized.csv",
        "analysis/bmj_rmr_v041/visual_summary.json",
        "docs/manuscript/v16/draft.md",
        "docs/manuscript/v16/supplement.md",
        "docs/manuscript/v17/draft.md",
        "docs/manuscript/v17/supplement.md",
        "docs/manuscript/v18/draft.md",
        "docs/manuscript/v18/supplement.md",
        "docs/specs/2026-08-25-bmj-rmr-v0.42.0-forward-completion-spec.md",
    ]
    paths = [root / value for value in relative]
    missing = [str(path.relative_to(root)) for path in paths if not path.exists()]
    if missing:
        raise FileNotFoundError(f"Missing locked inputs: {missing}")
    return paths


def inventory(root: Path) -> tuple[pd.DataFrame, pd.DataFrame, dict[str, object]]:
    manifest = pd.read_csv(root / "analysis/full_atlas/test_group_manifest.csv")
    normalized = pd.read_csv(root / "analysis/full_atlas/normalized_study_data.csv")
    primary = manifest.loc[csv_bool(manifest["primary_atlas"])].copy()
    primary["source_route"] = source_route(primary["source"])

    raw_row_counts = (
        normalized.groupby(PRIMARY_KEY, dropna=False)
        .size()
        .rename("raw_normalized_row_count")
        .reset_index()
    )
    analysis_rows = select_analysis_rows(primary, normalized)
    analysis_row_counts = (
        analysis_rows.groupby(PRIMARY_KEY, dropna=False)
        .size()
        .rename("analysis_row_count")
        .reset_index()
    )
    joined = primary.merge(raw_row_counts, on=PRIMARY_KEY, how="left", validate="one_to_one")
    joined = joined.merge(analysis_row_counts, on=PRIMARY_KEY, how="left", validate="one_to_one")
    if joined["raw_normalized_row_count"].isna().any():
        missing = joined.loc[joined["raw_normalized_row_count"].isna(), PRIMARY_KEY]
        raise AssertionError(f"Primary points without normalized rows:\n{missing}")

    normalized_primary = normalized.loc[csv_bool(normalized["primary_atlas"])].copy()
    primary_flag_keys = normalized_primary[PRIMARY_KEY].drop_duplicates()
    flag_check = primary.merge(
        primary_flag_keys.assign(row_primary_flag_present=True),
        on=PRIMARY_KEY,
        how="left",
    ).merge(raw_row_counts, on=PRIMARY_KEY, how="left", validate="one_to_one")
    flag_check = flag_check.merge(analysis_row_counts, on=PRIMARY_KEY, how="left", validate="one_to_one")
    exceptions = flag_check.loc[
        flag_check["row_primary_flag_present"].isna()
        | flag_check["raw_normalized_row_count"].ne(flag_check["study_rows"]),
        PRIMARY_KEY
        + [
            "test_name",
            "source_sheet",
            "study_rows",
            "raw_normalized_row_count",
            "analysis_row_count",
            "row_primary_flag_present",
        ],
    ].copy()
    exceptions["excluded_noncanonical_rows"] = (
        exceptions["raw_normalized_row_count"] - exceptions["analysis_row_count"]
    )
    exceptions["selection_rule"] = (
        "manifest source_sheet is canonical; exclude matching-group rows from other sheets"
    )

    manual = pd.read_csv(root / "analysis/full_atlas/pmc_manual_validation.csv")
    validation = pd.read_csv(root / "analysis/full_atlas/model_validation.csv")
    validation_primary = validation.loc[csv_bool(validation["primary_atlas"])]
    orientation = pd.read_csv(root / "analysis/full_atlas/orientation_audit.csv")

    summary = {
        "primary_operating_points": int(len(primary)),
        "reviews": int(primary["review_id"].nunique()),
        "raw_normalized_rows_joined": int(joined["raw_normalized_row_count"].sum()),
        "analysis_rows_after_manifest_sheet_selection": int(joined["analysis_row_count"].sum()),
        "excluded_noncanonical_sheet_rows": int(
            joined["raw_normalized_row_count"].sum() - joined["analysis_row_count"].sum()
        ),
        "primary_points_with_normalized_rows": int(joined["raw_normalized_row_count"].notna().sum()),
        "primary_points_with_exact_manifest_analysis_rows": int(
            joined["analysis_row_count"].astype(int).eq(joined["study_rows"].astype(int)).sum()
        ),
        "row_selection_exceptions": int(len(exceptions)),
        "source_routes": {
            key: int(value)
            for key, value in primary["source_route"].value_counts().sort_index().items()
        },
        "usable_source_summaries": int(len(validation_primary)),
        "manual_audit_tables": int(len(manual)),
        "manual_audit_normalized_rows": int(manual["normalized_rows_checked"].fillna(0).sum()),
        "manual_audit_normalized_cells": int(manual["normalized_cells_checked"].fillna(0).sum()),
        "manual_audit_transcription_errors": int(
            pd.to_numeric(manual["cell_transcription_errors"], errors="coerce")
            .fillna(0)
            .sum()
        ),
        "manual_audit_acceptance_errors": int(
            csv_bool(manual["group_acceptance_error"]).sum()
        ),
        "orientation_audit_groups": int(len(orientation)),
    }
    assert summary["primary_operating_points"] == 273
    assert summary["reviews"] == 210
    assert summary["primary_points_with_normalized_rows"] == 273
    assert summary["primary_points_with_exact_manifest_analysis_rows"] == 273
    assert summary["analysis_rows_after_manifest_sheet_selection"] == 4104
    assert summary["usable_source_summaries"] == 92
    assert summary["manual_audit_tables"] == 50
    assert summary["manual_audit_normalized_rows"] == 410
    assert summary["manual_audit_normalized_cells"] == 1640
    assert summary["orientation_audit_groups"] == 11
    return primary, analysis_rows, {"summary": summary, "row_selection_exceptions": exceptions}


def version_diff_skeleton(primary: pd.DataFrame, root: Path) -> pd.DataFrame:
    historical = pd.read_csv(root / "analysis/full_atlas/operating_points_20_percent.csv")
    historical = historical.loc[csv_bool(historical["primary_atlas"])].copy()
    historical = historical[PRIMARY_KEY + [
        "model",
        "sensitivity",
        "specificity",
        "information_bits_at_20_percent",
        "diagnostic_entropy_reduction_at_20_percent",
        "zero_cell_study_fraction",
    ]]
    regularized = pd.read_csv(root / "analysis/bmj_rmr_v040/operating_points_regularized.csv")
    keep = PRIMARY_KEY + [
        "pooled_sensitivity_median",
        "pooled_specificity_median",
        "information_bits_at_5_percent_median",
        "information_bits_at_20_percent_median",
        "information_bits_at_50_percent_median",
        "relative_information_at_5_percent_median",
        "relative_information_at_20_percent_median",
        "relative_information_at_50_percent_median",
    ]
    regularized = regularized[keep]

    skeleton = primary.merge(historical, on=PRIMARY_KEY, how="left", validate="one_to_one")
    skeleton = skeleton.merge(regularized, on=PRIMARY_KEY, how="left", validate="one_to_one")
    skeleton.insert(0, "operating_point_id", skeleton[PRIMARY_KEY].astype(str).agg("|".join, axis=1))
    skeleton["source_route"] = source_route(skeleton["source"])
    normalized = pd.read_csv(root / "analysis/full_atlas/normalized_study_data.csv")
    raw_counts = normalized.groupby(PRIMARY_KEY).size().rename("raw_normalized_rows").reset_index()
    analysis_rows = select_analysis_rows(primary, normalized)
    analysis_counts = analysis_rows.groupby(PRIMARY_KEY).size().rename("analysis_rows").reset_index()
    skeleton = skeleton.merge(raw_counts, on=PRIMARY_KEY, how="left", validate="one_to_one")
    skeleton = skeleton.merge(analysis_counts, on=PRIMARY_KEY, how="left", validate="one_to_one")
    skeleton["target_label_where_recorded"] = pd.NA
    skeleton["threshold_where_recorded"] = pd.NA
    skeleton["orientation"] = np.where(
        skeleton["sensitivity"] + skeleton["specificity"] - 1 < 0,
        "below chance on pooled REML estimate; source orientation retained",
        "source-defined direction",
    )
    skeleton["inclusion_status_v039"] = "primary reference sample"
    skeleton["inclusion_status_v040"] = "primary reference sample; unchanged"
    skeleton["inclusion_status_v041"] = "display derivation from v0.40.0; no refit"
    skeleton["inclusion_status_v042"] = "primary reference sample; pending Phase 1 estimator lock"
    skeleton["estimator_v039"] = skeleton["model"]
    skeleton["estimator_v040"] = "regularized direct-binomial local Gaussian approximation"
    skeleton["estimator_v041"] = "display derivation from v0.40.0; no refit"
    skeleton["estimator_v042"] = "pending Phase 1 promotion gate"
    skeleton["status_change_reason_v042"] = (
        "fixed operating-point identity; estimator and derived values pending Phase 1"
    )
    skeleton = skeleton.rename(
        columns={
            "study_rows": "study_count",
            "sensitivity": "sensitivity_v039",
            "specificity": "specificity_v039",
            "information_bits_at_20_percent": "information_bits_20_v039",
            "diagnostic_entropy_reduction_at_20_percent": "fraction_entropy_removed_20_v039",
            "pooled_sensitivity_median": "sensitivity_v040_v041",
            "pooled_specificity_median": "specificity_v040_v041",
            "information_bits_at_5_percent_median": "information_bits_5_v040_v041",
            "information_bits_at_20_percent_median": "information_bits_20_v040_v041",
            "information_bits_at_50_percent_median": "information_bits_50_v040_v041",
            "relative_information_at_5_percent_median": "fraction_entropy_removed_5_v040_v041",
            "relative_information_at_20_percent_median": "fraction_entropy_removed_20_v040_v041",
            "relative_information_at_50_percent_median": "fraction_entropy_removed_50_v040_v041",
        }
    )
    columns = [
        "operating_point_id", "source", "source_route", "review_id", "review_title",
        "group_id", "test_name", "target_label_where_recorded", "threshold_where_recorded",
        "study_count", "raw_normalized_rows", "analysis_rows",
        "zero_cell_study_fraction", "orientation", "eligibility", "primary_atlas",
        "selection_status", "estimator_v039", "sensitivity_v039", "specificity_v039",
        "information_bits_20_v039", "fraction_entropy_removed_20_v039",
        "estimator_v040", "sensitivity_v040_v041", "specificity_v040_v041",
        "information_bits_5_v040_v041", "information_bits_20_v040_v041",
        "information_bits_50_v040_v041", "fraction_entropy_removed_5_v040_v041",
        "fraction_entropy_removed_20_v040_v041", "fraction_entropy_removed_50_v040_v041",
        "estimator_v041", "estimator_v042", "inclusion_status_v039",
        "inclusion_status_v040", "inclusion_status_v041", "inclusion_status_v042",
        "status_change_reason_v042",
    ]
    return skeleton[columns].sort_values(PRIMARY_KEY).reset_index(drop=True)


def verify_prior_release_manifests(root: Path) -> pd.DataFrame:
    releases = [("v16", "0.39.0"), ("v17", "0.40.0"), ("v18", "0.41.0")]
    records: list[dict[str, object]] = []
    for directory, version in releases:
        base = root / "docs/manuscript" / directory
        manifest_path = base / f"release-manifest-v{version}.json"
        manifest = json.loads(manifest_path.read_text(encoding="utf-8"))
        artifacts = list(manifest["pdfs"]) + [manifest["reviewer_package"]]
        for item in artifacts:
            path = base / item["file"]
            records.append(
                {
                    "release": version,
                    "artifact": item["file"],
                    "manifest_sha256": item["sha256"],
                    "observed_sha256": file_hash(path),
                    "hash_matches": file_hash(path) == item["sha256"],
                    "bytes_match": path.stat().st_size == int(item["bytes"]),
                    "artifact_kind": "reviewer_zip" if path.suffix == ".zip" else "pdf",
                    "zip_entries": len(zipfile.ZipFile(path).infolist()) if path.suffix == ".zip" else np.nan,
                }
            )
    result = pd.DataFrame(records)
    if len(result) != 24 or not result["hash_matches"].all() or not result["bytes_match"].all():
        raise AssertionError("Historical release-manifest verification failed")
    return result


def phase0(root: Path) -> dict[str, object]:
    analysis_dir = root / "analysis/bmj_rmr_v042"
    manuscript_dir = root / "docs/manuscript/v19"
    qa_dir = analysis_dir / "qa"
    analysis_dir.mkdir(parents=True, exist_ok=True)
    manuscript_dir.mkdir(parents=True, exist_ok=True)
    qa_dir.mkdir(parents=True, exist_ok=True)

    primary, _normalized, audit = inventory(root)
    candidate_generation_commit = git_value(root, "rev-parse", "HEAD")
    checksum_records = []
    for path in locked_input_paths(root):
        relative = path.relative_to(root).as_posix()
        checkout = path.read_bytes()
        blob = git_blob(root, ORIENTATION_SOURCE_COMMIT, relative)
        checksum_records.append(
            {
                "file": relative,
                "checkout_sha256": bytes_hash(checkout),
                "git_blob_sha256": bytes_hash(blob),
                "normalized_lf_sha256": bytes_hash(checkout.replace(b"\r\n", b"\n")),
                "git_blob_normalized_lf_sha256": bytes_hash(blob.replace(b"\r\n", b"\n")),
                "line_ending_only_difference": (
                    checkout != blob
                    and checkout.replace(b"\r\n", b"\n") == blob.replace(b"\r\n", b"\n")
                ),
                "bytes": len(checkout),
                "role": "locked input",
            }
        )
    checksums = pd.DataFrame(checksum_records)
    checksums.to_csv(analysis_dir / "input_checksums.csv", index=False, lineterminator="\n")
    skeleton = version_diff_skeleton(primary, root)
    skeleton.to_csv(
        analysis_dir / "version_diff_all_releases.csv", index=False, lineterminator="\n"
    )
    audit["row_selection_exceptions"].to_csv(
        analysis_dir / "analysis_row_selection_exceptions.csv", index=False, lineterminator="\n"
    )
    prior_releases = verify_prior_release_manifests(root)
    prior_releases.to_csv(
        analysis_dir / "prior_release_manifest_verification.csv", index=False, lineterminator="\n"
    )

    run_manifest = {
        "analysis_version": VERSION,
        "release_date": RELEASE_DATE,
        "orientation_source_commit": ORIENTATION_SOURCE_COMMIT,
        "candidate_generation_commit": candidate_generation_commit,
        "branch": git_value(root, "branch", "--show-current"),
        "phase": 0,
        "status": "phase_0_candidate_pending_independent_qa",
        "role": "forward completion with estimator decision pending Phase 1",
        "fixed_reference_sample": audit["summary"],
        "input_checksums_file": "analysis/bmj_rmr_v042/input_checksums.csv",
        "version_diff_file": "analysis/bmj_rmr_v042/version_diff_all_releases.csv",
        "specification": "docs/specs/2026-08-25-bmj-rmr-v0.42.0-forward-completion-spec.md",
    }
    write_json(analysis_dir / "run_manifest.json", run_manifest)

    gate = {
        "phase": 0,
        "name": "orientation, lock, and version map",
        "status": "candidate_pending_independent_qa",
        "release_target": VERSION,
        "orientation_source_commit": ORIENTATION_SOURCE_COMMIT,
        "candidate_generation_commit": candidate_generation_commit,
        "checks": {
            "primary_operating_points_exact": audit["summary"]["primary_operating_points"] == 273,
            "reviews_exact": audit["summary"]["reviews"] == 210,
            "all_primary_points_have_normalized_rows": audit["summary"]["primary_points_with_normalized_rows"] == 273,
            "analysis_rows_match_manifest_counts": audit["summary"]["primary_points_with_exact_manifest_analysis_rows"] == 273,
            "version_diff_rows_exact": len(skeleton) == 273,
            "input_hashes_recorded": len(checksums) == len(locked_input_paths(root)),
            "prior_release_manifests_verified": bool(prior_releases["hash_matches"].all()),
        },
        "inventory": audit["summary"],
        "resolved_row_selection_discrepancy": {
            "description": "One manifest-selected Zenodo operating point joins 45 raw rows from two sheets although the manifest names one 22-row canonical sheet; all raw row-level primary flags are stale false values.",
            "policy": "Use the manifest source_sheet and study_rows as the analysis-row authority. Keep 22 Master_Extraction rows and exclude 23 TwoByTwo_Long rows without editing the locked input.",
            "file": "analysis/bmj_rmr_v042/analysis_row_selection_exceptions.csv",
        },
        "commands": [
            "py -3.13 scripts/bmj_rmr_v042.py phase0",
            "py -3.13 -m pytest tests/test_bmj_rmr_v042.py tests/test_bmj_rmr_v042_spec.py -q",
        ],
        "output_sha256": {
            "input_checksums": file_hash(analysis_dir / "input_checksums.csv"),
            "version_diff": file_hash(analysis_dir / "version_diff_all_releases.csv"),
            "row_selection_exceptions": file_hash(analysis_dir / "analysis_row_selection_exceptions.csv"),
            "prior_release_verification": file_hash(analysis_dir / "prior_release_manifest_verification.csv"),
        },
        "test_result": "12 targeted tests passed after blocking provenance corrections",
        "qa_context": "pending independent Phase 0 re-audit after blocking corrections",
    }
    write_json(qa_dir / "phase_gate_0.json", gate)
    return gate


def direct_binomial_promotion_gate(root: Path) -> pd.DataFrame:
    rows = [
        (
            "established_posterior_implementation",
            False,
            "Released v0.40.0 code is a numerical optimizer with a local Gaussian approximation, not a version-locked full posterior sampler.",
        ),
        (
            "prespecified_30_group_posterior_pilot",
            False,
            "A 30-group optimizer pilot exists, but no posterior pilot with all-route stratification and sampler diagnostics exists.",
        ),
        (
            "four_chain_diagnostics",
            False,
            "No four-chain rank-normalized R-hat, bulk ESS, tail ESS, divergence, or boundary-pathology record exists.",
        ),
        (
            "all_273_posterior_diagnostics",
            False,
            "All 273 optimizer fits exist; all-273 posterior diagnostics do not.",
        ),
        (
            "posterior_propagation",
            False,
            "Released ranges propagate a clipped local Hessian rather than posterior draws through all reported quantities.",
        ),
        (
            "released_code_recovery",
            False,
            "The prespecified 24-scenario local-range check covered sensitivity in 91.7% and specificity in 75.0% of scenarios, below a confirmatory interval claim.",
        ),
        (
            "prior_and_numerical_sensitivity_runs_exist",
            True,
            "Wider-prior and 9-node runs exist and are retained as sensitivity evidence.",
        ),
        (
            "model_family_sensitivity_preserves_framework_claims",
            False,
            "REML and direct-binomial comparisons exist, but no promoted posterior model-family analysis establishes this conjunctive requirement.",
        ),
        (
            "clean_reproducible_posterior_environment",
            False,
            "No documented clean environment reproduces a full posterior direct-binomial analysis with the required diagnostics.",
        ),
    ]
    frame = pd.DataFrame(rows, columns=["gate_requirement", "passed", "evidence"])
    frame["decision"] = np.where(frame["passed"], "satisfied", "missing_or_failed")
    frame["overall_promotion"] = False
    frame["fallback"] = "continuity-corrected bivariate REML primary; direct-binomial central estimates sensitivity"
    return frame


def canonical_group_payloads(root: Path) -> tuple[pd.DataFrame, list[dict[str, object]]]:
    primary, analysis_rows, _audit = inventory(root)
    payloads: list[dict[str, object]] = []
    for _, point in primary.sort_values(PRIMARY_KEY).iterrows():
        mask = np.logical_and.reduce(
            [analysis_rows[column].eq(point[column]).to_numpy() for column in PRIMARY_KEY]
        )
        rows = analysis_rows.loc[mask]
        data = [
            {
                "study_id": str(row.study_id),
                "tp": int(row.tp),
                "fp": int(row.fp),
                "fn": int(row.fn),
                "tn": int(row.tn),
            }
            for row in rows.itertuples(index=False)
        ]
        group = point.to_dict()
        group["primary_atlas"] = True
        group["rows"] = data
        payloads.append(group)
    return primary, payloads


def refit_canonical_reml(
    root: Path, draws: int = 20_000
) -> tuple[pd.DataFrame, pd.DataFrame]:
    _primary, payloads = canonical_group_payloads(root)
    outputs: list[dict[str, object]] = []
    multistart_records: list[dict[str, object]] = []
    for group in payloads:
        seed = (REML_SEED + int(hashlib.sha256(str(group["group_id"]).encode()).hexdigest()[:8], 16)) % (2**32)
        point, _boundaries = analyze_group(
            group, draws, seed, multistart_records=multistart_records
        )
        point["analysis_version"] = VERSION
        point["analysis_row_rule"] = "manifest source_sheet where recorded; otherwise all group-matched rows"
        point["mean_logit_sensitivity"] = float(logit(point["sensitivity"]))
        point["mean_logit_specificity"] = float(logit(point["specificity"]))
        mean = np.asarray([point["mean_logit_sensitivity"], point["mean_logit_specificity"]])
        mean_covariance = np.asarray(
            [
                [point["mean_logit_sensitivity_variance"], point["mean_logit_covariance"]],
                [point["mean_logit_covariance"], point["mean_logit_specificity_variance"]],
            ]
        )
        between_covariance = np.asarray(
            [
                [point["between_logit_sensitivity_variance"], point["between_logit_covariance"]],
                [point["between_logit_covariance"], point["between_logit_specificity_variance"]],
            ]
        )
        for prefix, covariance in (
            ("pooled_mean", mean_covariance),
            ("predictive", mean_covariance + between_covariance),
        ):
            low = expit(mean - norm.ppf(0.975) * np.sqrt(np.diag(covariance)))
            middle = expit(mean)
            high = expit(mean + norm.ppf(0.975) * np.sqrt(np.diag(covariance)))
            for index, metric in enumerate(("sensitivity", "specificity")):
                point[f"{prefix}_{metric}_low"] = float(low[index])
                point[f"{prefix}_{metric}_median"] = float(middle[index])
                point[f"{prefix}_{metric}_high"] = float(high[index])
        outputs.append(point)
    frame = pd.DataFrame(outputs).sort_values(PRIMARY_KEY).reset_index(drop=True)
    if len(frame) != 273 or not frame["model_converged"].all():
        raise AssertionError("Canonical REML regeneration did not produce 273 converged points")
    multistart = pd.DataFrame.from_records(multistart_records).sort_values(
        PRIMARY_KEY + ["start_order"]
    ).reset_index(drop=True)
    if len(multistart) != 273 * 7:
        raise AssertionError("Canonical REML multistart audit does not contain seven starts per point")
    return frame, multistart


def continuity_correction_sensitivity(
    root: Path, canonical: pd.DataFrame
) -> tuple[pd.DataFrame, dict[str, object], pd.DataFrame]:
    """Refit all points with correction triggered only by a study zero cell."""
    _primary, payloads = canonical_group_payloads(root)
    canonical_lookup = canonical.set_index(PRIMARY_KEY)
    records: list[dict[str, object]] = []
    multistart_records: list[dict[str, object]] = []
    for group in payloads:
        fit = fit_bivariate_reml(
            group["rows"], continuity_correction="zero_cell_triggered"
        )
        for start_record in fit["optimizer_start_results"]:
            multistart_records.append(
                {
                    **{column: group[column] for column in PRIMARY_KEY},
                    "continuity_correction_rule": "zero_cell_triggered",
                    **start_record,
                }
            )
        sensitivity, specificity = expit(fit["mean"])
        information = float(
            diagnostic_metrics(sensitivity, specificity, 0.20)["information_bits"]
        )
        key = tuple(group[column] for column in PRIMARY_KEY)
        central = canonical_lookup.loc[key]
        records.append(
            {
                **{column: group[column] for column in PRIMARY_KEY},
                "studies": len(group["rows"]),
                "zero_cell_study_count": int(
                    sum(
                        any(row[name] == 0 for name in ("tp", "fp", "fn", "tn"))
                        for row in group["rows"]
                    )
                ),
                "zero_cell_study_fraction": float(central["zero_cell_study_fraction"]),
                "canonical_continuity_correction_rule": "0.5 added to every cell of every study",
                "sensitivity_continuity_correction_rule": (
                    "0.5 added to all four cells only for studies containing any zero; "
                    "zero-free studies uncorrected"
                ),
                "canonical_sensitivity": float(central["sensitivity"]),
                "zero_cell_triggered_sensitivity": float(sensitivity),
                "absolute_sensitivity_difference": abs(
                    float(central["sensitivity"]) - float(sensitivity)
                ),
                "canonical_specificity": float(central["specificity"]),
                "zero_cell_triggered_specificity": float(specificity),
                "absolute_specificity_difference": abs(
                    float(central["specificity"]) - float(specificity)
                ),
                "canonical_information_bits_at_20_percent": float(
                    central["information_bits_at_20_percent"]
                ),
                "zero_cell_triggered_information_bits_at_20_percent": information,
                "absolute_information_bits_at_20_percent_difference": abs(
                    float(central["information_bits_at_20_percent"]) - information
                ),
                "optimizer_converged": True,
                "optimizer_iterations": int(fit["iterations"]),
                "optimizer_max_abs_gradient": float(fit["max_abs_gradient"]),
                "optimizer_projected_kkt_gradient_log_l00": float(
                    fit["projected_kkt_gradient_log_l00"]
                ),
                "optimizer_projected_kkt_gradient_l10": float(
                    fit["projected_kkt_gradient_l10"]
                ),
                "optimizer_projected_kkt_gradient_log_l11": float(
                    fit["projected_kkt_gradient_log_l11"]
                ),
                "optimizer_projected_kkt_gradient_max_abs": float(
                    fit["projected_kkt_gradient_max_abs"]
                ),
                "optimizer_parameter_log_l00": float(
                    fit["optimizer_parameter_log_l00"]
                ),
                "optimizer_parameter_l10": float(fit["optimizer_parameter_l10"]),
                "optimizer_parameter_log_l11": float(
                    fit["optimizer_parameter_log_l11"]
                ),
                "optimizer_tied_solution_log_l00_at_lower_bound": bool(
                    fit["optimizer_tied_solution_log_l00_at_lower_bound"]
                ),
                "optimizer_tied_solution_log_l11_at_lower_bound": bool(
                    fit["optimizer_tied_solution_log_l11_at_lower_bound"]
                ),
                "optimizer_selected_log_l00_at_lower_bound": bool(
                    fit["optimizer_selected_log_l00_at_lower_bound"]
                ),
                "optimizer_selected_log_l11_at_lower_bound": bool(
                    fit["optimizer_selected_log_l11_at_lower_bound"]
                ),
                "optimizer_hard_bound_atol": float(
                    fit["optimizer_hard_bound_atol"]
                ),
                "optimizer_multistart_count": int(fit["optimizer_multistart_count"]),
                "optimizer_valid_start_count": int(
                    fit["optimizer_valid_start_count"]
                ),
                "optimizer_selected_start": fit["optimizer_selected_start"],
                "optimizer_selected_start_order": int(
                    fit["optimizer_selected_start_order"]
                ),
                "optimizer_minimum_recomputed_objective": float(
                    fit["optimizer_minimum_recomputed_objective"]
                ),
                "optimizer_selected_objective_delta_from_minimum": float(
                    fit["optimizer_selected_objective_delta_from_minimum"]
                ),
                "optimizer_objective_gap_to_second_best": float(
                    fit["optimizer_objective_gap_to_second_best"]
                ),
                "optimizer_objective_gap_to_next_distinct_basin": float(
                    fit["optimizer_objective_gap_to_next_distinct_basin"]
                ),
                "optimizer_valid_objective_range": float(
                    fit["optimizer_valid_objective_range"]
                ),
                "optimizer_valid_sensitivity_range": float(
                    fit["optimizer_valid_sensitivity_range"]
                ),
                "optimizer_valid_specificity_range": float(
                    fit["optimizer_valid_specificity_range"]
                ),
                "optimizer_alternative_rounded_claim_changed_across_starts": bool(
                    fit[
                        "optimizer_alternative_rounded_claim_changed_across_starts"
                    ]
                ),
                "optimizer_empirical_start_valid": bool(
                    fit["optimizer_empirical_start_valid"]
                ),
                "optimizer_empirical_to_selected_objective_delta": float(
                    fit["optimizer_empirical_to_selected_objective_delta"]
                ),
                "optimizer_empirical_to_selected_sensitivity_delta": float(
                    fit["optimizer_empirical_to_selected_sensitivity_delta"]
                ),
                "optimizer_empirical_to_selected_specificity_delta": float(
                    fit["optimizer_empirical_to_selected_specificity_delta"]
                ),
                "optimizer_rounded_claim_changed_from_empirical": bool(
                    fit["optimizer_rounded_claim_changed_from_empirical"]
                ),
                "optimizer_material_objective_path_dependence": bool(
                    fit["optimizer_material_objective_path_dependence"]
                ),
                "optimizer_material_central_path_dependence": bool(
                    fit["optimizer_material_central_path_dependence"]
                ),
                "optimizer_material_path_dependence": bool(
                    fit["optimizer_material_path_dependence"]
                ),
                "optimizer_path_dependence_class": fit[
                    "optimizer_path_dependence_class"
                ],
            }
        )
    table = pd.DataFrame.from_records(records).sort_values(PRIMARY_KEY).reset_index(drop=True)
    for diagonal in ("l00", "l11"):
        parameter = f"optimizer_parameter_log_{diagonal}"
        distance = f"optimizer_selected_log_{diagonal}_distance_to_lower_bound"
        ambiguity = (
            f"optimizer_selected_log_{diagonal}_in_1e_6_to_1e_4_ambiguity_band"
        )
        table[distance] = (table[parameter] + 8.0).abs()
        table[ambiguity] = table[distance].gt(1e-6) & table[distance].le(1e-4)
    table["optimizer_any_tied_solution_cholesky_diagonal_at_lower_bound"] = table[
        [
            "optimizer_tied_solution_log_l00_at_lower_bound",
            "optimizer_tied_solution_log_l11_at_lower_bound",
        ]
    ].any(axis=1)
    metric_stems = {
        "sensitivity": "absolute_sensitivity_difference",
        "specificity": "absolute_specificity_difference",
        "information_bits_at_20_percent": "absolute_information_bits_at_20_percent_difference",
    }
    summary: dict[str, object] = {
        "analysis_version": VERSION,
        "operating_points": len(table),
        "canonical_rule": "historical regularising convention: add 0.5 to every cell of every study",
        "sensitivity_rule": (
            "add 0.5 to all four cells only for studies containing any zero; "
            "leave zero-free studies uncorrected"
        ),
        "same_reml_optimizer": True,
        "converged_fits": int(table["optimizer_converged"].sum()),
        "tied_solution_log_l00_at_lower_bound": int(
            table["optimizer_tied_solution_log_l00_at_lower_bound"].sum()
        ),
        "tied_solution_log_l11_at_lower_bound": int(
            table["optimizer_tied_solution_log_l11_at_lower_bound"].sum()
        ),
        "tied_solution_any_cholesky_diagonal_at_lower_bound": int(
            table[
                "optimizer_any_tied_solution_cholesky_diagonal_at_lower_bound"
            ].sum()
        ),
        "selected_endpoint_log_l00_at_lower_bound": int(
            table["optimizer_selected_log_l00_at_lower_bound"].sum()
        ),
        "selected_endpoint_log_l11_at_lower_bound": int(
            table["optimizer_selected_log_l11_at_lower_bound"].sum()
        ),
        "selected_endpoint_l11_ambiguity_band_count": int(
            table[
                "optimizer_selected_log_l11_in_1e_6_to_1e_4_ambiguity_band"
            ].sum()
        ),
        "maximum_optimizer_absolute_gradient": float(
            table["optimizer_max_abs_gradient"].max()
        ),
        "maximum_projected_kkt_gradient": float(
            table["optimizer_projected_kkt_gradient_max_abs"].max()
        ),
        "material_path_dependence_fits": int(
            table["optimizer_material_path_dependence"].sum()
        ),
        "tiny_boundary_refinement_fits": int(
            table["optimizer_path_dependence_class"]
            .eq("tiny_boundary_refinement")
            .sum()
        ),
    }
    for stem, column in metric_stems.items():
        summary[f"moved_{stem}_count_above_1e_12"] = int(table[column].gt(1e-12).sum())
        summary[f"median_absolute_{stem}_difference"] = float(table[column].median())
        summary[f"maximum_absolute_{stem}_difference"] = float(table[column].max())
    summary["any_central_quantity_moved_count_above_1e_12"] = int(
        table[list(metric_stems.values())].max(axis=1).gt(1e-12).sum()
    )
    multistart = pd.DataFrame.from_records(multistart_records).sort_values(
        PRIMARY_KEY + ["start_order"]
    ).reset_index(drop=True)
    if len(multistart) != 273 * 7:
        raise AssertionError(
            "Zero-cell-triggered REML multistart audit does not contain seven starts per point"
        )
    return table, summary, multistart


def reml_regeneration_comparison(root: Path, canonical: pd.DataFrame) -> pd.DataFrame:
    historical = pd.read_csv(root / "analysis/full_atlas/operating_points_20_percent.csv")
    historical = historical.loc[csv_bool(historical["primary_atlas"])].copy()
    keep = PRIMARY_KEY + ["sensitivity", "specificity", "information_bits_at_20_percent"]
    comparison = historical[keep].merge(
        canonical[keep + ["study_rows"]],
        on=PRIMARY_KEY,
        suffixes=("_historical_reml", "_v042_manifest_sheet"),
        validate="one_to_one",
    )
    for metric in ("sensitivity", "specificity", "information_bits_at_20_percent"):
        comparison[f"absolute_{metric}_difference"] = (
            comparison[f"{metric}_historical_reml"]
            - comparison[f"{metric}_v042_manifest_sheet"]
        ).abs()
    exception_key = pd.read_csv(
        root / "analysis/bmj_rmr_v042/analysis_row_selection_exceptions.csv"
    ).iloc[0]["group_id"]
    comparison["row_selection_changed"] = comparison["group_id"].eq(exception_key)
    comparison["interpretation"] = np.where(
        comparison["row_selection_changed"],
        "v0.42.0 corrects the canonical source sheet; historical releases consumed both sheets",
        "same locked analysis rows; regenerated central estimate",
    )
    return comparison


def reml_recovery(replicates: int = 24, draws: int = 20_000) -> tuple[pd.DataFrame, dict[str, object]]:
    rng = np.random.default_rng(REML_SEED + 800_000)
    records: list[dict[str, object]] = []
    for replicate in range(replicates):
        studies = (5, 8, 15, 30)[replicate % 4]
        true_se = (0.65, 0.80, 0.92)[replicate % 3]
        true_sp = (0.70, 0.85, 0.95)[(replicate // 3) % 3]
        true_sd_se = (0.25, 0.65)[replicate % 2]
        true_sd_sp = (0.30, 0.75)[(replicate // 2) % 2]
        true_rho = (-0.35, 0.0, 0.35)[replicate % 3]
        covariance = np.asarray(
            [
                [true_sd_se**2, true_rho * true_sd_se * true_sd_sp],
                [true_rho * true_sd_se * true_sd_sp, true_sd_sp**2],
            ]
        )
        effects = rng.multivariate_normal(np.zeros(2), covariance, size=studies)
        probabilities = expit(effects + np.asarray([logit(true_se), logit(true_sp)]))
        diseased = rng.integers(35, 180, size=studies)
        nondiseased = rng.integers(35, 220, size=studies)
        tp = rng.binomial(diseased, probabilities[:, 0])
        tn = rng.binomial(nondiseased, probabilities[:, 1])
        data = [
            {
                "study_id": f"sim-{replicate + 1}-{index + 1}",
                "tp": int(a),
                "fn": int(b - a),
                "tn": int(c),
                "fp": int(d - c),
            }
            for index, (a, b, c, d) in enumerate(zip(tp, diseased, tn, nondiseased))
        ]
        fit = fit_bivariate_reml(data)
        pooled_low = expit(
            fit["mean"] - norm.ppf(0.975) * np.sqrt(np.diag(fit["mean_covariance"]))
        )
        pooled_mid = expit(fit["mean"])
        pooled_high = expit(
            fit["mean"] + norm.ppf(0.975) * np.sqrt(np.diag(fit["mean_covariance"]))
        )
        pred_low = expit(
            fit["mean"] - norm.ppf(0.975) * np.sqrt(np.diag(fit["predictive_covariance"]))
        )
        pred_mid = expit(fit["mean"])
        pred_high = expit(
            fit["mean"] + norm.ppf(0.975) * np.sqrt(np.diag(fit["predictive_covariance"]))
        )
        records.append(
            {
                "replicate": replicate + 1,
                "studies": studies,
                "true_sensitivity": true_se,
                "estimated_sensitivity": float(expit(fit["mean"][0])),
                "pooled_mean_sensitivity_low": pooled_low[0],
                "pooled_mean_sensitivity_median": pooled_mid[0],
                "pooled_mean_sensitivity_high": pooled_high[0],
                "sensitivity_interval_includes_truth": pooled_low[0] <= true_se <= pooled_high[0],
                "true_specificity": true_sp,
                "estimated_specificity": float(expit(fit["mean"][1])),
                "pooled_mean_specificity_low": pooled_low[1],
                "pooled_mean_specificity_median": pooled_mid[1],
                "pooled_mean_specificity_high": pooled_high[1],
                "specificity_interval_includes_truth": pooled_low[1] <= true_sp <= pooled_high[1],
                "predictive_sensitivity_low": pred_low[0],
                "predictive_sensitivity_median": pred_mid[0],
                "predictive_sensitivity_high": pred_high[0],
                "predictive_specificity_low": pred_low[1],
                "predictive_specificity_median": pred_mid[1],
                "predictive_specificity_high": pred_high[1],
                "true_sd_sensitivity": true_sd_se,
                "estimated_sd_sensitivity": float(np.sqrt(fit["between_covariance"][0, 0])),
                "true_sd_specificity": true_sd_sp,
                "estimated_sd_specificity": float(np.sqrt(fit["between_covariance"][1, 1])),
                "true_correlation": true_rho,
                "estimated_correlation": float(
                    fit["between_covariance"][0, 1]
                    / np.sqrt(fit["between_covariance"][0, 0] * fit["between_covariance"][1, 1])
                ),
                "optimizer_iterations": fit["iterations"],
                "optimizer_max_abs_gradient": fit["max_abs_gradient"],
            }
        )
    frame = pd.DataFrame(records)
    summary = {
        "analysis_version": VERSION,
        "model": "continuity-corrected bivariate REML",
        "replicates": replicates,
        "sensitivity_scenario_inclusion_fraction": float(
            frame["sensitivity_interval_includes_truth"].mean()
        ),
        "specificity_scenario_inclusion_fraction": float(
            frame["specificity_interval_includes_truth"].mean()
        ),
        "median_absolute_sensitivity_error": float(
            (frame["estimated_sensitivity"] - frame["true_sensitivity"]).abs().median()
        ),
        "median_absolute_specificity_error": float(
            (frame["estimated_specificity"] - frame["true_specificity"]).abs().median()
        ),
        "interpretation": (
            "Limited recovery/smoke diagnostic with one generated dataset in each of "
            "24 heterogeneous scenarios. Scenario inclusion proportions are not "
            "repeated-sampling coverage estimates. The intervals "
            "are approximate large-sample plug-in Wald/model summaries conditional on "
            "fitted heterogeneity; heterogeneity-parameter uncertainty is not propagated, "
            "and predictive ranges were not checked."
        ),
    }
    return frame, summary


def estimator_sensitivity_summary(root: Path, canonical: pd.DataFrame) -> tuple[pd.DataFrame, dict[str, object]]:
    regularized = pd.read_csv(root / "analysis/bmj_rmr_v040/operating_points_regularized.csv")
    keep = PRIMARY_KEY + [
        "pooled_sensitivity_median",
        "pooled_specificity_median",
        "information_bits_at_20_percent_median",
        "relative_information_at_20_percent_median",
    ]
    table = canonical[
        PRIMARY_KEY
        + ["sensitivity", "specificity", "information_bits_at_20_percent", "diagnostic_entropy_reduction_at_20_percent", "zero_cell_study_fraction"]
    ].merge(regularized[keep], on=PRIMARY_KEY, validate="one_to_one")
    direct_plugin = diagnostic_metrics(
        table["pooled_sensitivity_median"].to_numpy(),
        table["pooled_specificity_median"].to_numpy(),
        0.20,
    )["information_bits"]
    table["canonical_reml_plugin_information_bits_at_20_percent"] = table[
        "information_bits_at_20_percent"
    ]
    table["direct_binomial_plugin_information_bits_at_20_percent"] = direct_plugin
    table["absolute_like_for_like_plugin_information_bits_at_20_percent_difference"] = (
        table["canonical_reml_plugin_information_bits_at_20_percent"]
        - table["direct_binomial_plugin_information_bits_at_20_percent"]
    ).abs()
    pairs = [
        ("sensitivity", "pooled_sensitivity_median", 1.0, "sensitivity"),
        ("specificity", "pooled_specificity_median", 1.0, "specificity"),
        ("information_bits_at_20_percent", "information_bits_at_20_percent_median", 1.0, "information_bits_at_20_percent"),
        ("diagnostic_entropy_reduction_at_20_percent", "relative_information_at_20_percent_median", 100.0, "relative_information_at_20_percent_percentage_points"),
    ]
    summary: dict[str, object] = {
        "analysis_version": VERSION,
        "comparison": "canonical continuity-corrected bivariate REML versus released 15-node regularized direct-binomial central estimates",
        "operating_points": len(table),
    }
    for canonical_name, regularized_name, scale, stem in pairs:
        diff = scale * (table[canonical_name] - table[regularized_name]).abs()
        table[f"absolute_{stem}_difference"] = diff
        summary[f"median_absolute_{stem}_difference"] = float(diff.median())
        summary[f"maximum_absolute_{stem}_difference"] = float(diff.max())
    summary["spearman_zero_cell_fraction_vs_absolute_information_difference"] = float(
        table[["zero_cell_study_fraction", "absolute_information_bits_at_20_percent_difference"]]
        .corr(method="spearman")
        .iloc[0, 1]
    )
    plugin_difference = table[
        "absolute_like_for_like_plugin_information_bits_at_20_percent_difference"
    ]
    summary["like_for_like_plugin_comparison"] = (
        "canonical REML plug-in information versus direct-binomial plug-in information, "
        "both evaluated at their componentwise central sensitivity and specificity"
    )
    summary[
        "median_absolute_like_for_like_plugin_information_bits_at_20_percent_difference"
    ] = float(plugin_difference.median())
    summary[
        "maximum_absolute_like_for_like_plugin_information_bits_at_20_percent_difference"
    ] = float(plugin_difference.max())
    summary["release_to_release_comparison"] = (
        "canonical REML plug-in information versus the locked released direct-binomial "
        "propagated median; different central-summary estimands"
    )
    return table, summary


def reml_diagnostics(canonical: pd.DataFrame) -> tuple[pd.DataFrame, dict[str, object]]:
    table = canonical[
        PRIMARY_KEY
        + [
            "study_rows",
            "zero_cell_study_fraction",
            "optimizer_iterations",
            "optimizer_max_abs_gradient",
            "optimizer_projected_kkt_gradient_log_l00",
            "optimizer_projected_kkt_gradient_l10",
            "optimizer_projected_kkt_gradient_log_l11",
            "optimizer_projected_kkt_gradient_max_abs",
            "between_logit_sensitivity_variance",
            "between_logit_specificity_variance",
            "between_logit_covariance",
            "between_logit_correlation",
            "mean_logit_sensitivity_variance",
            "mean_logit_specificity_variance",
            "mean_logit_covariance",
            "optimizer_parameter_log_l00",
            "optimizer_parameter_l10",
            "optimizer_parameter_log_l11",
            "optimizer_tied_solution_log_l00_at_lower_bound",
            "optimizer_tied_solution_log_l11_at_lower_bound",
            "optimizer_selected_log_l00_at_lower_bound",
            "optimizer_selected_log_l11_at_lower_bound",
            "optimizer_hard_bound_atol",
            "optimizer_multistart_count",
            "optimizer_valid_start_count",
            "optimizer_selected_start",
            "optimizer_selected_start_order",
            "optimizer_minimum_recomputed_objective",
            "optimizer_selected_objective_delta_from_minimum",
            "optimizer_objective_tie_tolerance",
            "optimizer_objective_gap_to_second_best",
            "optimizer_objective_gap_to_next_distinct_basin",
            "optimizer_valid_objective_range",
            "optimizer_valid_sensitivity_range",
            "optimizer_valid_specificity_range",
            "optimizer_alternative_rounded_claim_changed_across_starts",
            "optimizer_empirical_start_valid",
            "optimizer_empirical_to_selected_objective_delta",
            "optimizer_empirical_to_selected_sensitivity_delta",
            "optimizer_empirical_to_selected_specificity_delta",
            "optimizer_rounded_claim_changed_from_empirical",
            "optimizer_material_objective_path_dependence",
            "optimizer_material_central_path_dependence",
            "optimizer_material_path_dependence",
            "optimizer_path_dependence_class",
        ]
    ].copy()
    for diagonal in ("l00", "l11"):
        parameter = f"optimizer_parameter_log_{diagonal}"
        distance = f"optimizer_selected_log_{diagonal}_distance_to_lower_bound"
        ambiguity = (
            f"optimizer_selected_log_{diagonal}_in_1e_6_to_1e_4_ambiguity_band"
        )
        table[distance] = (table[parameter] + 8.0).abs()
        table[ambiguity] = table[distance].gt(1e-6) & table[distance].le(1e-4)
    a = table["between_logit_sensitivity_variance"].to_numpy()
    d = table["between_logit_specificity_variance"].to_numpy()
    b = table["between_logit_covariance"].to_numpy()
    eigen_root = np.sqrt((a - d) ** 2 + 4 * b**2)
    table["between_covariance_min_eigenvalue"] = (a + d - eigen_root) / 2
    table["between_covariance_max_eigenvalue"] = (a + d + eigen_root) / 2
    table["near_singular_between_covariance"] = table[
        "between_covariance_min_eigenvalue"
    ].lt(1e-8)
    table["absolute_between_correlation_at_least_0_99"] = table[
        "between_logit_correlation"
    ].abs().ge(0.99)
    table["optimizer_gradient_above_1e_4"] = table["optimizer_max_abs_gradient"].gt(1e-4)
    table["optimizer_projected_kkt_gradient_above_1e_4"] = table[
        "optimizer_projected_kkt_gradient_max_abs"
    ].gt(1e-4)
    table["optimizer_any_tied_solution_cholesky_diagonal_at_lower_bound"] = table[
        [
            "optimizer_tied_solution_log_l00_at_lower_bound",
            "optimizer_tied_solution_log_l11_at_lower_bound",
        ]
    ].any(axis=1)
    table["optimizer_any_selected_endpoint_cholesky_diagonal_at_lower_bound"] = table[
        [
            "optimizer_selected_log_l00_at_lower_bound",
            "optimizer_selected_log_l11_at_lower_bound",
        ]
    ].any(axis=1)
    table["old_diagnostic_flag"] = table[
        [
            "near_singular_between_covariance",
            "absolute_between_correlation_at_least_0_99",
            "optimizer_gradient_above_1e_4",
        ]
    ].any(axis=1)
    table["hard_bound_missed_by_old_flags"] = (
        table["optimizer_any_tied_solution_cholesky_diagonal_at_lower_bound"]
        & ~table["old_diagnostic_flag"]
    )
    table["diagnostic_flag"] = table[
        [
            "near_singular_between_covariance",
            "absolute_between_correlation_at_least_0_99",
            "optimizer_gradient_above_1e_4",
            "optimizer_any_tied_solution_cholesky_diagonal_at_lower_bound",
        ]
    ].any(axis=1)
    summary = {
        "analysis_version": VERSION,
        "model": "continuity-corrected bivariate REML",
        "operating_points": len(table),
        "converged_fits": int(canonical["model_converged"].sum()),
        "absolute_between_correlation_at_least_0_99": int(
            table["absolute_between_correlation_at_least_0_99"].sum()
        ),
        "near_singular_between_covariance_min_eigenvalue_below_1e_8": int(
            table["near_singular_between_covariance"].sum()
        ),
        "optimizer_gradient_above_1e_4": int(table["optimizer_gradient_above_1e_4"].sum()),
        "optimizer_projected_kkt_gradient_above_1e_4": int(
            table["optimizer_projected_kkt_gradient_above_1e_4"].sum()
        ),
        "optimizer_tied_solution_log_l00_at_lower_bound": int(
            table["optimizer_tied_solution_log_l00_at_lower_bound"].sum()
        ),
        "optimizer_tied_solution_log_l11_at_lower_bound": int(
            table["optimizer_tied_solution_log_l11_at_lower_bound"].sum()
        ),
        "optimizer_any_tied_solution_cholesky_diagonal_at_lower_bound": int(
            table[
                "optimizer_any_tied_solution_cholesky_diagonal_at_lower_bound"
            ].sum()
        ),
        "optimizer_selected_endpoint_log_l00_at_lower_bound": int(
            table["optimizer_selected_log_l00_at_lower_bound"].sum()
        ),
        "optimizer_selected_endpoint_log_l11_at_lower_bound": int(
            table["optimizer_selected_log_l11_at_lower_bound"].sum()
        ),
        "optimizer_any_selected_endpoint_cholesky_diagonal_at_lower_bound": int(
            table[
                "optimizer_any_selected_endpoint_cholesky_diagonal_at_lower_bound"
            ].sum()
        ),
        "optimizer_selected_endpoint_l00_ambiguity_band_count": int(
            table[
                "optimizer_selected_log_l00_in_1e_6_to_1e_4_ambiguity_band"
            ].sum()
        ),
        "optimizer_selected_endpoint_l11_ambiguity_band_count": int(
            table[
                "optimizer_selected_log_l11_in_1e_6_to_1e_4_ambiguity_band"
            ].sum()
        ),
        "optimizer_selected_endpoint_largest_distance_classified_at_bound": float(
            pd.concat(
                [
                    table.loc[
                        table["optimizer_selected_log_l00_at_lower_bound"],
                        "optimizer_selected_log_l00_distance_to_lower_bound",
                    ],
                    table.loc[
                        table["optimizer_selected_log_l11_at_lower_bound"],
                        "optimizer_selected_log_l11_distance_to_lower_bound",
                    ],
                ],
                ignore_index=True,
            ).max()
        ),
        "optimizer_selected_endpoint_smallest_distance_not_classified_at_bound": float(
            pd.concat(
                [
                    table.loc[
                        ~table["optimizer_selected_log_l00_at_lower_bound"],
                        "optimizer_selected_log_l00_distance_to_lower_bound",
                    ],
                    table.loc[
                        ~table["optimizer_selected_log_l11_at_lower_bound"],
                        "optimizer_selected_log_l11_distance_to_lower_bound",
                    ],
                ],
                ignore_index=True,
            ).min()
        ),
        "optimizer_hard_bound_absolute_tolerance": float(
            table["optimizer_hard_bound_atol"].iloc[0]
        ),
        "optimizer_ambiguity_band_upper_distance": 1e-4,
        "hard_bound_fits_missed_by_old_flags": int(
            table["hard_bound_missed_by_old_flags"].sum()
        ),
        "union_of_all_diagnostic_flags": int(table["diagnostic_flag"].sum()),
        "maximum_optimizer_absolute_gradient": float(table["optimizer_max_abs_gradient"].max()),
        "maximum_projected_kkt_gradient": float(
            table["optimizer_projected_kkt_gradient_max_abs"].max()
        ),
        "material_path_dependence_fits": int(
            table["optimizer_material_path_dependence"].sum()
        ),
        "tiny_boundary_refinement_fits": int(
            table["optimizer_path_dependence_class"]
            .eq("tiny_boundary_refinement")
            .sum()
        ),
        "numerically_equivalent_multistart_fits": int(
            table["optimizer_path_dependence_class"]
            .eq("numerically_equivalent")
            .sum()
        ),
        "selected_start_counts": {
            str(name): int(count)
            for name, count in table["optimizer_selected_start"]
            .value_counts()
            .sort_index()
            .items()
        },
        "interpretation": (
            "Seven deterministic starts were fitted and independently re-evaluated "
            "for every operating point. A tied-solution boundary flag means that at "
            "least one endpoint satisfying the objective-and-central numerical-tie "
            "rule lies at -8; selected-endpoint boundary counts are retained "
            "separately. The tied-solution flag records fragility without implying "
            "that the selected Cholesky parameter equals -8. Boundary correlations, "
            "near-singular heterogeneity estimates, raw and projected gradients, and "
            "path-dependent basins are retained."
        ),
    }
    appendicitis = table.loc[table["group_id"].eq(APPENDICITIS_ID)].iloc[0]
    summary["appendicitis_tied_solution_log_l11_at_lower_bound"] = bool(
        appendicitis["optimizer_tied_solution_log_l11_at_lower_bound"]
    )
    summary["appendicitis_selected_endpoint_log_l11_at_lower_bound"] = bool(
        appendicitis["optimizer_selected_log_l11_at_lower_bound"]
    )
    summary["appendicitis_between_study_correlation"] = float(
        appendicitis["between_logit_correlation"]
    )
    return table, summary


def write_estimator_decision(
    root: Path,
    recovery: dict[str, object],
    sensitivity: dict[str, object],
    diagnostics: dict[str, object],
    correction_sensitivity: dict[str, object],
) -> None:
    checksums = pd.read_csv(root / "analysis/bmj_rmr_v042/input_checksums.csv").set_index("file")
    manifest_hash = checksums.loc[
        "analysis/full_atlas/test_group_manifest.csv", "normalized_lf_sha256"
    ]
    normalized_hash = checksums.loc[
        "analysis/full_atlas/normalized_study_data.csv", "normalized_lf_sha256"
    ]
    text = f"""# Estimator decision

**Status:** Phase 6 corrective candidate pending fresh Gate 6 re-QA

## Decision

The released direct-binomial implementation does not provide a full posterior sampler, four-chain diagnostics, an all-273 posterior diagnostics record, posterior propagation, or a repeated-simulation uncertainty evaluation. The wider-prior and quadrature checks remain useful sensitivity evidence, but they do not satisfy the missing requirements. No third estimator-development path was opened.

Version 0.42.0 therefore uses the prespecified fallback: a continuity-corrected bivariate REML model regenerated for all 273 operating points from 4,104 primary-analysis rows. Its primary implementation adds 0.5 to every cell of every study as a historical regularising convention, not as a generic zero-only correction. The source-metadata sheet is authoritative where recorded. This corrects one Zenodo point from 45 raw joined rows to its declared 22-row `Master_Extraction` input. The operating-point identity remains fixed.

## Uncertainty interpretation

The central sensitivity and specificity are inverse-logit transforms of the fitted mean logits. A pooled-mean interval is an approximate large-sample plug-in model-based 95% interval for the estimated mean operating point, conditional on the fitted heterogeneity. A predictive range is an approximate plug-in model-based 95% distribution that adds fitted between-study heterogeneity for an additional study's latent underlying operating point. Heterogeneity-parameter uncertainty is not propagated, and predictive ranges were not checked by the 24-scenario diagnostic. Neither quantity describes the observed binomial estimate from a future study, guarantees performance in a new clinical setting, or supplies a disease-specific action interval.

Every headline point estimate and interval comes from this REML family. Direct-binomial central estimates, the wider-prior run, and the nine-node run are sensitivity analyses only. A REML interval may not be attached to a direct-binomial point estimate.

## Promotion-gate evidence

The machine-readable estimator decision record is `analysis/bmj_rmr_v042/direct_binomial_promotion_gate.csv`. Eight of nine requirements are missing or failed. The only satisfied element is the existence of prior and numerical sensitivity runs. The previously released 24-scenario local-Gaussian smoke diagnostic included the generating sensitivity in 91.7% and generating specificity in 75.0% of its single-dataset scenarios; it is only a limited single-dataset-per-scenario diagnostic.

## Limited recovery diagnostic

In the regenerated 24-scenario REML smoke diagnostic, the pooled-mean interval included the generating sensitivity in {100 * recovery['sensitivity_scenario_inclusion_fraction']:.1f}% of scenarios and the generating specificity in {100 * recovery['specificity_scenario_inclusion_fraction']:.1f}%. Median absolute central-estimate errors were {recovery['median_absolute_sensitivity_error']:.3f} and {recovery['median_absolute_specificity_error']:.3f}. Each scenario contains one generated dataset. These limited recovery results are not repeated-sampling coverage estimates, and predictive ranges were not checked.

## Chosen-path diagnostics

All {diagnostics['converged_fits']} selected fits reported optimizer convergence. The diagnostics flag {diagnostics['absolute_between_correlation_at_least_0_99']} between-study correlations with absolute value at least 0.99, {diagnostics['near_singular_between_covariance_min_eigenvalue_below_1e_8']} near-singular covariance estimates, and {diagnostics['optimizer_gradient_above_1e_4']} raw gradients above 1e-4. Among endpoints satisfying the numerical-tie rule, {diagnostics['optimizer_tied_solution_log_l00_at_lower_bound']} fits had a tied solution with log-L00 at -8 and {diagnostics['optimizer_tied_solution_log_l11_at_lower_bound']} had one with log-L11 at -8, for {diagnostics['optimizer_any_tied_solution_cholesky_diagonal_at_lower_bound']} unique tied-solution boundary flags. The stationarity-selected endpoints themselves were at the L00 and L11 bounds in {diagnostics['optimizer_selected_endpoint_log_l00_at_lower_bound']} and {diagnostics['optimizer_selected_endpoint_log_l11_at_lower_bound']} fits, respectively. Retaining the tied-solution flag records boundary fragility when an objectively and centrally indistinguishable solution lies at the hard constraint; it does not imply that every selected Cholesky parameter equals -8. Seven tied-solution flags were missed by the older flag set, and the union contains {diagnostics['union_of_all_diagnostic_flags']} fits. Appendicitis has both a selected endpoint and a tied solution at the log-L11 bound, with correlation {diagnostics['appendicitis_between_study_correlation']:.6f}. Its central profile remains illustrative, but joint heterogeneity and predictive ranges are fragile and conditional on this fit.

## Sensitivity models

Across 273 points, the like-for-like plug-in central-estimate comparison at the 20% anchor differed between REML and direct-binomial central sensitivity/specificity by a median {sensitivity['median_absolute_like_for_like_plugin_information_bits_at_20_percent_difference']:.6f} bits and maximum {sensitivity['maximum_absolute_like_for_like_plugin_information_bits_at_20_percent_difference']:.6f} bits. This comparison still combines likelihood family, regularisation or prior, and continuity-correction choices. The analysis-family comparison between REML plug-in information and direct-binomial propagated-median information is {sensitivity['median_absolute_information_bits_at_20_percent_difference']:.6f} and {sensitivity['maximum_absolute_information_bits_at_20_percent_difference']:.6f} bits; these are different central-summary estimands. The direct continuity-correction sensitivity refitted all {correction_sensitivity['operating_points']} points with the same seven-start REML optimizer and added 0.5 only within studies containing any zero. Its median and maximum information changes were {correction_sensitivity['median_absolute_information_bits_at_20_percent_difference']:.6f} and {correction_sensitivity['maximum_absolute_information_bits_at_20_percent_difference']:.6f} bits. The universal correction remains the primary analysis solely for historical reproducibility.

## Locked inputs

- Group manifest LF-normalized SHA-256: `{manifest_hash}`
- Normalized study rows LF-normalized SHA-256: `{normalized_hash}`
- Primary row-selection exception: `analysis/bmj_rmr_v042/analysis_row_selection_exceptions.csv`
- Primary results: `analysis/bmj_rmr_v042/operating_points_primary.csv`
- Recovery results: `analysis/bmj_rmr_v042/reml_recovery.csv`

## Claims affected

All headline operating-point, information, likelihood-ratio, result-probability, post-result probability, pooled-mean uncertainty, predictive uncertainty, Atlas summary, landmark, branch-entropy, worked-profile, caption, cover-letter, and summary-point values must be regenerated from the primary REML output. The 0.41.0 regularized appendicitis values and its 38.1% empirical CEA crossing may appear only as labelled sensitivity results.
"""
    (root / "docs/manuscript/v19/ESTIMATOR_DECISION.md").write_text(text, encoding="utf-8")


def phase1(root: Path, draws: int = 20_000) -> dict[str, object]:
    analysis_dir = root / "analysis/bmj_rmr_v042"
    qa_dir = analysis_dir / "qa"
    promotion = direct_binomial_promotion_gate(root)
    promotion.to_csv(
        analysis_dir / "direct_binomial_promotion_gate.csv", index=False, lineterminator="\n"
    )
    canonical, canonical_multistart = refit_canonical_reml(root, draws=draws)
    canonical.to_csv(
        analysis_dir / "operating_points_primary.csv", index=False, lineterminator="\n"
    )
    canonical_multistart.to_csv(
        analysis_dir / "reml_multistart_all_primary.csv",
        index=False,
        lineterminator="\n",
    )
    regeneration = reml_regeneration_comparison(root, canonical)
    regeneration.to_csv(
        analysis_dir / "reml_regeneration_comparison.csv", index=False, lineterminator="\n"
    )
    recovery_frame, recovery_summary = reml_recovery(draws=draws)
    recovery_frame.to_csv(analysis_dir / "reml_recovery.csv", index=False, lineterminator="\n")
    write_json(analysis_dir / "reml_recovery_summary.json", recovery_summary)
    sensitivity_frame, sensitivity_summary = estimator_sensitivity_summary(root, canonical)
    sensitivity_frame.to_csv(
        analysis_dir / "estimator_sensitivity_all_primary.csv", index=False, lineterminator="\n"
    )
    write_json(analysis_dir / "estimator_sensitivity_summary.json", sensitivity_summary)
    correction_frame, correction_summary, correction_multistart = (
        continuity_correction_sensitivity(root, canonical)
    )
    correction_frame.to_csv(
        analysis_dir / "continuity_correction_sensitivity_all_primary.csv",
        index=False,
        lineterminator="\n",
    )
    write_json(
        analysis_dir / "continuity_correction_sensitivity_summary.json",
        correction_summary,
    )
    correction_multistart.to_csv(
        analysis_dir / "continuity_correction_multistart_all_primary.csv",
        index=False,
        lineterminator="\n",
    )
    diagnostics_frame, diagnostics_summary = reml_diagnostics(canonical)
    diagnostics_frame.to_csv(
        analysis_dir / "reml_diagnostics_all_primary.csv", index=False, lineterminator="\n"
    )
    write_json(analysis_dir / "reml_diagnostics_summary.json", diagnostics_summary)
    write_estimator_decision(
        root,
        recovery_summary,
        sensitivity_summary,
        diagnostics_summary,
        correction_summary,
    )

    version_diff = pd.read_csv(analysis_dir / "version_diff_all_releases.csv")
    released_v042_columns = [
        "sensitivity_v042",
        "specificity_v042",
        "information_bits_20_v042",
        "fraction_entropy_removed_20_v042",
    ]
    version_diff = version_diff.drop(
        columns=[column for column in released_v042_columns if column in version_diff.columns]
    )
    canonical_values = canonical[
        ["operating_point_id"] if "operating_point_id" in canonical.columns else PRIMARY_KEY
    ]
    if "operating_point_id" not in canonical_values.columns:
        canonical_values = canonical[PRIMARY_KEY + [
            "sensitivity", "specificity", "information_bits_at_20_percent",
            "diagnostic_entropy_reduction_at_20_percent",
        ]].copy()
        canonical_values["operating_point_id"] = canonical_values[PRIMARY_KEY].astype(str).agg("|".join, axis=1)
    else:
        raise AssertionError("Unexpected pre-existing operating_point_id")
    canonical_values = canonical_values.rename(columns={
        "sensitivity": "sensitivity_v042",
        "specificity": "specificity_v042",
        "information_bits_at_20_percent": "information_bits_20_v042",
        "diagnostic_entropy_reduction_at_20_percent": "fraction_entropy_removed_20_v042",
    })
    version_diff = version_diff.merge(
        canonical_values[["operating_point_id", "sensitivity_v042", "specificity_v042", "information_bits_20_v042", "fraction_entropy_removed_20_v042"]],
        on="operating_point_id", how="left", validate="one_to_one"
    )
    version_diff["estimator_v042"] = "continuity-corrected bivariate REML"
    version_diff["inclusion_status_v042"] = "primary reference sample; canonical REML regenerated"
    version_diff["status_change_reason_v042"] = np.where(
        version_diff["group_id"].eq(
            pd.read_csv(analysis_dir / "analysis_row_selection_exceptions.csv").iloc[0]["group_id"]
        ),
        "canonical REML regenerated after manifest-sheet correction from 45 raw joined rows to 22 declared rows",
        "canonical REML regenerated from the same analysis rows as historical REML; validation and manuscript architecture updated",
    )
    version_diff.to_csv(
        analysis_dir / "version_diff_all_releases.csv", index=False, lineterminator="\n"
    )

    unchanged = regeneration.loc[~regeneration["row_selection_changed"]]
    central_tolerance = float(
        unchanged[[
            "absolute_sensitivity_difference",
            "absolute_specificity_difference",
            "absolute_information_bits_at_20_percent_difference",
        ]].to_numpy().max()
    )
    gate = {
        "phase": 1,
        "name": "canonical-estimator decision",
        "status": "candidate_pending_independent_qa",
        "canonical_estimator": "continuity-corrected bivariate REML",
        "direct_binomial_promotion": "failed; mandatory fallback taken",
        "checks": {
            "promotion_gate_unambiguous": bool(
                (~promotion["passed"]).sum() == 8 and not promotion["overall_promotion"].any()
            ),
            "all_273_reml_fits_converged": bool(len(canonical) == 273 and canonical["model_converged"].all()),
            "all_intervals_ordered": bool(
                (canonical["pooled_mean_sensitivity_low"] <= canonical["pooled_mean_sensitivity_median"]).all()
                and (canonical["pooled_mean_sensitivity_median"] <= canonical["pooled_mean_sensitivity_high"]).all()
                and (canonical["predictive_sensitivity_low"] <= canonical["predictive_sensitivity_median"]).all()
                and (canonical["predictive_sensitivity_median"] <= canonical["predictive_sensitivity_high"]).all()
            ),
            "historical_regeneration_differences_classified": bool(
                int(regeneration["row_selection_changed"].sum()) == 1
                and len(
                    unchanged.loc[
                        unchanged["group_id"].str.contains(
                            "PMC7170259", case=False, na=False
                        )
                    ]
                )
                == 1
                and float(
                    unchanged.loc[
                        unchanged["group_id"].str.contains(
                            "PMC7170259", case=False, na=False
                        ),
                        "absolute_sensitivity_difference",
                    ].iloc[0]
                )
                > 0.01
                and float(
                    unchanged.loc[
                        ~unchanged["group_id"].str.contains(
                            "PMC7170259", case=False, na=False
                        ),
                        [
                            "absolute_sensitivity_difference",
                            "absolute_specificity_difference",
                            "absolute_information_bits_at_20_percent_difference",
                        ],
                    ].to_numpy().max()
                )
                < 2e-6
            ),
            "corrected_zenodo_point_regenerated": bool(regeneration["row_selection_changed"].sum() == 1),
            "reml_recovery_regenerated": len(recovery_frame) == 24,
            "chosen_path_diagnostics_recorded": bool(
                diagnostics_summary["operating_points"] == 273
                and diagnostics_summary["converged_fits"] == 273
            ),
            "hard_optimizer_bounds_reconstructed": bool(
                diagnostics_summary[
                    "optimizer_tied_solution_log_l00_at_lower_bound"
                ]
                >= 0
                and diagnostics_summary[
                    "optimizer_tied_solution_log_l11_at_lower_bound"
                ]
                >= 0
                and diagnostics_summary[
                    "optimizer_any_tied_solution_cholesky_diagonal_at_lower_bound"
                ]
                >= max(
                    diagnostics_summary[
                        "optimizer_tied_solution_log_l00_at_lower_bound"
                    ],
                    diagnostics_summary[
                        "optimizer_tied_solution_log_l11_at_lower_bound"
                    ],
                )
                and diagnostics_summary["union_of_all_diagnostic_flags"]
                >= diagnostics_summary[
                    "optimizer_any_tied_solution_cholesky_diagonal_at_lower_bound"
                ]
            ),
            "canonical_multistart_complete": bool(
                len(canonical_multistart) == 273 * 7
                and canonical_multistart.groupby(PRIMARY_KEY).size().eq(7).all()
                and canonical_multistart.groupby(PRIMARY_KEY)["selected"].sum().eq(1).all()
            ),
            "zero_cell_triggered_multistart_complete": bool(
                len(correction_multistart) == 273 * 7
                and correction_multistart.groupby(PRIMARY_KEY).size().eq(7).all()
                and correction_multistart.groupby(PRIMARY_KEY)["selected"]
                .sum()
                .eq(1)
                .all()
            ),
            "zero_cell_triggered_correction_refit_complete": bool(
                len(correction_frame) == 273
                and correction_summary["converged_fits"] == 273
                and correction_summary["same_reml_optimizer"]
            ),
            "like_for_like_plugin_estimator_comparison_complete": bool(
                len(sensitivity_frame) == 273
                and sensitivity_frame[
                    "absolute_like_for_like_plugin_information_bits_at_20_percent_difference"
                ].notna().all()
            ),
            "no_mixed_model_intervals": bool(
                canonical["model"].eq("continuity-corrected bivariate REML").all()
                and np.allclose(
                    canonical["sensitivity"], canonical["pooled_mean_sensitivity_median"], atol=1e-12
                )
                and np.allclose(
                    canonical["specificity"], canonical["pooled_mean_specificity_median"], atol=1e-12
                )
                and np.allclose(
                    canonical["sensitivity"], canonical["predictive_sensitivity_median"], atol=1e-12
                )
                and np.allclose(
                    canonical["specificity"], canonical["predictive_specificity_median"], atol=1e-12
                )
                and all(
                    column.endswith("_median")
                    for column in sensitivity_frame.columns
                    if column.startswith("pooled_")
                )
            ),
            "predictive_marginal_ranges_not_narrower": bool(
                (
                    canonical["predictive_sensitivity_high"] - canonical["predictive_sensitivity_low"]
                    >= canonical["pooled_mean_sensitivity_high"] - canonical["pooled_mean_sensitivity_low"] - 1e-12
                ).all()
                and (
                    canonical["predictive_specificity_high"] - canonical["predictive_specificity_low"]
                    >= canonical["pooled_mean_specificity_high"] - canonical["pooled_mean_specificity_low"] - 1e-12
                ).all()
            ),
        },
        "maximum_absolute_regeneration_difference_among_unchanged_points": central_tolerance,
        "recovery_summary": recovery_summary,
        "sensitivity_summary": sensitivity_summary,
        "diagnostics_summary": diagnostics_summary,
        "commands": [
            "py -3.13 scripts/bmj_rmr_v042.py phase1",
            "py -3.13 -m pytest tests/test_bmj_rmr_v042.py tests/test_bmj_rmr_v042_spec.py tests/test_cochrane_dta_atlas.py -q",
        ],
        "output_sha256": {
            name: file_hash(analysis_dir / name)
            for name in [
                "direct_binomial_promotion_gate.csv",
                "operating_points_primary.csv",
                "reml_regeneration_comparison.csv",
                "reml_recovery.csv",
                "reml_recovery_summary.json",
                "estimator_sensitivity_all_primary.csv",
                "estimator_sensitivity_summary.json",
                "reml_diagnostics_all_primary.csv",
                "reml_diagnostics_summary.json",
                "continuity_correction_sensitivity_all_primary.csv",
                "continuity_correction_sensitivity_summary.json",
                "version_diff_all_releases.csv",
            ]
        },
        "qa_context": "pending fresh diagnostic meta-analysis and statistical-computing review",
    }
    gate["output_sha256"]["ESTIMATOR_DECISION.md"] = file_hash(
        root / "docs/manuscript/v19/ESTIMATOR_DECISION.md"
    )
    write_json(qa_dir / "phase_gate_1.json", gate)
    run_manifest_path = analysis_dir / "run_manifest.json"
    run_manifest = json.loads(run_manifest_path.read_text(encoding="utf-8"))
    run_manifest.update(
        {
            "phase": 1,
            "status": "phase_1_candidate_pending_independent_qa",
            "canonical_estimator": "continuity-corrected bivariate REML",
            "sensitivity_estimator": "regularized direct-binomial central estimates",
            "phase_1_gate": "analysis/bmj_rmr_v042/qa/phase_gate_1.json",
            "role": "forward completion with estimator locked; source validation pending Phase 2",
        }
    )
    write_json(run_manifest_path, run_manifest)
    return gate


def source_summary_fidelity(root: Path, canonical: pd.DataFrame) -> tuple[pd.DataFrame, dict[str, object]]:
    manifest = pd.read_csv(root / "analysis/full_atlas/test_group_manifest.csv")
    primary = manifest.loc[csv_bool(manifest["primary_atlas"])].copy()
    usable = primary.loc[
        primary["published_sensitivity"].notna() & primary["published_specificity"].notna()
    ].copy()
    table = usable.merge(
        canonical[PRIMARY_KEY + ["sensitivity", "specificity", "model"]],
        on=PRIMARY_KEY,
        how="left",
        validate="one_to_one",
    )
    table["sensitivity_difference"] = table["sensitivity"] - table["published_sensitivity"]
    table["specificity_difference"] = table["specificity"] - table["published_specificity"]
    table["absolute_sensitivity_difference"] = table["sensitivity_difference"].abs()
    table["absolute_specificity_difference"] = table["specificity_difference"].abs()
    table["large_discrepancy"] = table[
        ["absolute_sensitivity_difference", "absolute_specificity_difference"]
    ].max(axis=1).ge(0.10)
    table["threshold_metadata_status"] = "no separate threshold field available"
    table["study_count_metadata_status"] = np.where(
        table["published_studies"].eq(table["study_rows"]),
        "source-reported study count matches canonical row count",
        "source-reported study count differs from canonical row count",
    )
    table["estimator_comparison_status"] = (
        "source pooling estimator is not fully specified in locked metadata; "
        "canonical estimator is continuity-corrected bivariate REML"
    )

    def fidelity_explanation(row: pd.Series) -> str:
        if not row["large_discrepancy"]:
            return (
                "Difference is below 0.10 for both sensitivity and specificity; "
                "the source-reported summary and canonical REML estimate remain distinct."
            )
        metadata = (
            f"Available metadata: test label '{row['test_name']}', analysis scope "
            f"'{row['analysis_scope']}', source model status '{row['model_status']}', "
            "and no separate threshold field. "
        )
        if pd.notna(row["published_studies"]) and int(row["published_studies"]) != int(row["study_rows"]):
            return (
                metadata
                + f"The source summary reports {int(row['published_studies'])} studies, whereas "
                f"the canonical manifest contains {int(row['study_rows'])} study rows. This observed "
                "source-definition mismatch may contribute to the difference but does not establish causality."
            )
        return (
            metadata
            + f"The source-reported and canonical study counts both equal {int(row['study_rows'])}. "
            "The locked metadata do not contain enough threshold or pooling-definition detail to "
            "attribute the difference to a single cause."
        )

    table["explanation_from_available_metadata"] = table.apply(fidelity_explanation, axis=1)
    summary = {
        "analysis_version": VERSION,
        "comparison": "source-reported pooled summaries versus canonical continuity-corrected bivariate REML",
        "operating_points": len(table),
        "median_absolute_sensitivity_difference": float(table["absolute_sensitivity_difference"].median()),
        "maximum_absolute_sensitivity_difference": float(table["absolute_sensitivity_difference"].max()),
        "median_absolute_specificity_difference": float(table["absolute_specificity_difference"].median()),
        "maximum_absolute_specificity_difference": float(table["absolute_specificity_difference"].max()),
        "points_with_either_absolute_difference_at_least_0_10": int(table["large_discrepancy"].sum()),
        "interpretation": "Fidelity comparison between two reported summaries, not evidence that either estimator is truth.",
    }
    if len(table) != 92:
        raise AssertionError(f"Expected 92 usable source summaries, found {len(table)}")
    return table, summary


def _normalized_text(value: object) -> str:
    if pd.isna(value):
        return ""
    return " ".join(str(value).split())


def manual_audit_verification(root: Path) -> tuple[pd.DataFrame, pd.DataFrame, dict[str, object]]:
    audit = pd.read_csv(
        root / "analysis/full_atlas/pmc_manual_validation.csv",
        dtype={"pmcid": str, "table_id": str},
    )
    candidates = pd.read_csv(
        root / "analysis/pmc_open_atlas/candidate_tables.csv",
        dtype={"pmcid": str, "table_id": str},
    )
    candidates = candidates.sort_values(["pmcid", "table_id"]).drop_duplicates(
        ["pmcid", "table_id"], keep="first"
    )
    tables = pd.read_csv(
        root / "analysis/pmc_open_atlas/table_manifest.csv",
        dtype={"pmcid": str, "table_id": str},
    )
    articles = pd.read_csv(
        root / "analysis/pmc_open_atlas/articles.csv", dtype={"pmcid": str}
    ).drop_duplicates("pmcid")
    normalized = pd.read_csv(root / "analysis/pmc_open_atlas/normalized_study_data.csv")

    candidate_fields = [
        "pmcid", "table_id", "caption", "candidate_type", "score", "matched_terms",
        "source_sha256", "source_text",
    ]
    table_fields = ["pmcid", "table_id", "parse_status", "source_sha256"]
    joined = audit.merge(
        candidates[candidate_fields].rename(
            columns={column: f"candidate_{column}" for column in candidate_fields if column not in {"pmcid", "table_id"}}
        ),
        on=["pmcid", "table_id"],
        how="left",
        validate="one_to_one",
    ).merge(
        tables[table_fields].rename(
            columns={column: f"table_{column}" for column in table_fields if column not in {"pmcid", "table_id"}}
        ),
        on=["pmcid", "table_id"],
        how="left",
        validate="one_to_one",
    ).merge(
        articles[["pmcid", "source_sha256"]].rename(columns={"source_sha256": "article_source_sha256"}),
        on="pmcid",
        how="left",
        validate="many_to_one",
    )

    actual_rows: list[int] = []
    lexical_checks: list[bool] = []
    source_text_hashes: list[str] = []
    for row in joined.itertuples(index=False):
        mask = normalized["review_id"].eq(f"PMC:{row.pmcid}") & normalized["group_id"].str.contains(
            f":{row.table_id}:", regex=False, na=False
        )
        source_rows = normalized.loc[mask]
        actual_rows.append(len(source_rows))
        source_text = str(row.candidate_source_text)
        source_text_hashes.append(bytes_hash(source_text.encode("utf-8")))
        if source_rows.empty:
            lexical_checks.append(True)
        else:
            lexical_checks.append(
                all(str(int(value)) in source_text for value in source_rows[["tp", "fp", "fn", "tn"]].to_numpy().ravel())
            )
    joined["recomputed_normalized_rows"] = actual_rows
    joined["recomputed_normalized_cells"] = 4 * joined["recomputed_normalized_rows"]
    joined["locked_source_text_sha256"] = source_text_hashes
    joined["source_hash_consistent_across_article_candidate_table"] = (
        joined["candidate_source_sha256"].eq(joined["table_source_sha256"])
        & joined["candidate_source_sha256"].eq(joined["article_source_sha256"])
    )
    joined["caption_matches"] = [
        _normalized_text(left) == _normalized_text(right)
        for left, right in zip(joined["caption"], joined["candidate_caption"])
    ]
    joined["candidate_type_matches"] = joined["candidate_type"].eq(joined["candidate_candidate_type"])
    joined["score_matches"] = pd.to_numeric(joined["score"]).eq(
        pd.to_numeric(joined["candidate_score"])
    )
    joined["parse_status_matches"] = joined["parse_status"].eq(joined["table_parse_status"])
    joined["matched_terms_match"] = joined["matched_terms"].fillna("").eq(
        joined["candidate_matched_terms"].fillna("")
    )
    joined["normalized_denominators_match"] = (
        joined["normalized_rows_checked"].eq(joined["recomputed_normalized_rows"])
        & joined["normalized_cells_checked"].eq(joined["recomputed_normalized_cells"])
    )
    joined["all_cell_values_lexically_present_in_locked_source_text"] = lexical_checks
    joined["verification_pass"] = joined[
        [
            "source_hash_consistent_across_article_candidate_table",
            "caption_matches",
            "candidate_type_matches",
            "score_matches",
            "parse_status_matches",
            "matched_terms_match",
            "normalized_denominators_match",
            "all_cell_values_lexically_present_in_locked_source_text",
        ]
    ].all(axis=1)
    output_columns = [
        "audit_number", "cohort", "pmcid", "pmc_url", "table_id", "table_label",
        "expected_acceptance", "normalized_rows_checked", "normalized_cells_checked",
        "recomputed_normalized_rows", "recomputed_normalized_cells",
        "cell_transcription_errors", "group_acceptance_error", "adjudication",
        "candidate_source_sha256", "locked_source_text_sha256",
        "source_hash_consistent_across_article_candidate_table", "caption_matches",
        "candidate_type_matches", "score_matches", "parse_status_matches",
        "matched_terms_match", "normalized_denominators_match",
        "all_cell_values_lexically_present_in_locked_source_text", "verification_pass",
    ]
    verification = joined[output_columns].copy()

    input_paths = [
        "analysis/full_atlas/pmc_manual_validation.csv",
        "analysis/pmc_open_atlas/candidate_tables.csv",
        "analysis/pmc_open_atlas/table_manifest.csv",
        "analysis/pmc_open_atlas/normalized_study_data.csv",
        "analysis/pmc_open_atlas/articles.csv",
        "scripts/pmc_pilot.py",
        "scripts/cochrane_dta_atlas.py",
    ]
    checksum_records = []
    for relative in input_paths:
        checkout = (root / relative).read_bytes()
        blob = git_blob(root, ORIENTATION_SOURCE_COMMIT, relative)
        checksum_records.append(
            {
                "file": relative,
                "checkout_sha256": bytes_hash(checkout),
                "git_blob_sha256": bytes_hash(blob),
                "normalized_lf_sha256": bytes_hash(checkout.replace(b"\r\n", b"\n")),
                "git_blob_normalized_lf_sha256": bytes_hash(blob.replace(b"\r\n", b"\n")),
                "unchanged_from_orientation_source": checkout.replace(b"\r\n", b"\n") == blob.replace(b"\r\n", b"\n"),
            }
        )
    checksums = pd.DataFrame(checksum_records)
    transcription = pd.to_numeric(audit["cell_transcription_errors"], errors="coerce").fillna(0)
    included = audit["cohort"].eq("included")
    safeguards = audit["cohort"].eq("orientation_safeguard")
    rejected = audit["cohort"].eq("rejected_near_miss")
    extracted = included | safeguards
    summary = {
        "analysis_version": VERSION,
        "audit_role": "locked author audit; not a new independent adjudication",
        "tables": len(audit),
        "cohorts": {key: int(value) for key, value in audit["cohort"].value_counts().items()},
        "accepted_tables": int(audit["expected_acceptance"].sum()),
        "rejected_near_misses": int((~audit["expected_acceptance"]).sum()),
        "normalized_rows_checked": int(audit["normalized_rows_checked"].sum()),
        "normalized_cells_checked": int(audit["normalized_cells_checked"].sum()),
        "recorded_transcription_errors": int(transcription.sum()),
        "recorded_group_acceptance_errors": int(csv_bool(audit["group_acceptance_error"]).sum()),
        "included_tables": int(included.sum()),
        "included_normalized_rows": int(audit.loc[included, "normalized_rows_checked"].sum()),
        "included_normalized_cells": int(audit.loc[included, "normalized_cells_checked"].sum()),
        "orientation_safeguard_tables": int(safeguards.sum()),
        "orientation_safeguard_normalized_rows": int(audit.loc[safeguards, "normalized_rows_checked"].sum()),
        "orientation_safeguard_normalized_cells": int(audit.loc[safeguards, "normalized_cells_checked"].sum()),
        "rejected_near_miss_tables": int(rejected.sum()),
        "rejected_near_miss_normalized_rows": int(audit.loc[rejected, "normalized_rows_checked"].sum()),
        "transcription_error_numerator": int(transcription.sum()),
        "transcription_error_denominator_cells": int(audit.loc[extracted, "normalized_cells_checked"].sum()),
        "transcription_error_denominator_rows": int(audit.loc[extracted, "normalized_rows_checked"].sum()),
        "transcription_error_denominator_extracted_tables": int(extracted.sum()),
        "acceptance_error_numerator": int(csv_bool(audit["group_acceptance_error"]).sum()),
        "acceptance_error_denominator_decisions": len(audit),
        "verification_rows_passed": int(verification["verification_pass"].sum()),
        "locked_inputs_unchanged": bool(checksums["unchanged_from_orientation_source"].all()),
        "source_hash_verification_scope": "Consistency of recorded JATS SHA-256 across article, candidate-table, and table-manifest records; raw JATS payloads are not stored in the release repository.",
    }
    if len(verification) != 50 or not verification["verification_pass"].all():
        raise AssertionError("Locked manual-audit verification failed")
    return verification, checksums, summary


def orientation_study_audit(root: Path) -> tuple[pd.DataFrame, dict[str, object]]:
    audit = pd.read_csv(root / "analysis/full_atlas/orientation_audit.csv")
    normalized = pd.read_csv(root / "analysis/full_atlas/normalized_study_data.csv")
    manifest = pd.read_csv(root / "analysis/full_atlas/test_group_manifest.csv")
    corpus = pd.read_csv(root / "analysis/full_atlas/corpus_manifest.csv")
    audit = audit.rename(columns={
        "review_title": "audit_review_title",
        "test_name": "audit_test_name",
        "source_url": "audit_source_url",
        "studies": "recorded_study_rows",
        "aggregate_tp": "recorded_aggregate_tp",
        "aggregate_fp": "recorded_aggregate_fp",
        "aggregate_fn": "recorded_aggregate_fn",
        "aggregate_tn": "recorded_aggregate_tn",
        "aggregate_sensitivity": "recorded_aggregate_sensitivity",
        "aggregate_specificity": "recorded_aggregate_specificity",
        "aggregate_youden_j": "recorded_aggregate_youden_j",
        "adjudication": "recorded_adjudication",
        "parser_error_found": "recorded_parser_error_found",
    })
    row_columns = [
        "source", "review_id", "group_id", "review_title", "test_name", "study_id",
        "tp", "fp", "fn", "tn", "source_status", "source_row",
    ]
    group_meta = manifest[PRIMARY_KEY + [
        "source_version", "source_file", "source_sheet", "source_url", "source_sha256",
        "analysis_scope", "eligibility", "primary_atlas", "selection_status",
    ]].rename(columns={
        "source_file": "group_source_file",
        "source_sheet": "group_source_sheet",
        "source_url": "group_source_url",
        "source_sha256": "group_source_sha256",
    })
    corpus_meta = corpus[["source", "review_id", "source_file", "source_sha256"]].rename(
        columns={"source_file": "corpus_source_file", "source_sha256": "corpus_source_sha256"}
    )
    rows = audit.merge(
        normalized[row_columns],
        on=["source", "review_id", "group_id"],
        how="left",
        validate="one_to_many",
    )
    rows = rows.merge(group_meta, on=PRIMARY_KEY, how="left", validate="many_to_one")
    rows = rows.merge(corpus_meta, on=["source", "review_id"], how="left", validate="many_to_one")
    aggregate = rows.groupby(PRIMARY_KEY, as_index=False)[["tp", "fp", "fn", "tn"]].sum()
    aggregate = aggregate.rename(columns={
        "tp": "aggregate_tp", "fp": "aggregate_fp", "fn": "aggregate_fn", "tn": "aggregate_tn",
    })
    aggregate["aggregate_sensitivity"] = aggregate["aggregate_tp"] / (
        aggregate["aggregate_tp"] + aggregate["aggregate_fn"]
    )
    aggregate["aggregate_specificity"] = aggregate["aggregate_tn"] / (
        aggregate["aggregate_fp"] + aggregate["aggregate_tn"]
    )
    aggregate["aggregate_youden_j"] = (
        aggregate["aggregate_sensitivity"] + aggregate["aggregate_specificity"] - 1
    )
    rows = rows.merge(aggregate, on=PRIMARY_KEY, how="left", validate="many_to_one")
    rows["aggregate_direction"] = np.where(
        rows["aggregate_youden_j"].lt(0),
        "below chance under source-defined positive-test direction",
        "not below chance under source-defined positive-test direction",
    )
    rows["source_defined_direction"] = "as encoded in the source table; no outcome flip"
    rows["source_value_status"] = rows["source_status"].fillna("not recorded")
    rows["parser_transformation"] = "none documented in locked normalized fields"
    rows["error_status"] = np.where(
        csv_bool(rows["recorded_parser_error_found"]),
        "parser error recorded in locked author audit",
        "no parser error recorded in locked author audit",
    )
    rows["target_label_where_recorded"] = pd.NA
    rows["target_label_status"] = "no separate target-label field in locked normalized data"
    rows["final_decision"] = "excluded without automatic flipping"
    rows["effective_source_file"] = rows["group_source_file"].fillna(rows["corpus_source_file"])
    rows["effective_source_sha256"] = rows["group_source_sha256"].fillna(rows["corpus_source_sha256"])
    rows["effective_source_url"] = rows["group_source_url"].fillna(rows["audit_source_url"])
    rows["source_identity_level"] = np.where(
        rows["effective_source_url"].notna(), "group", "archive_or_package"
    )
    expected = audit.set_index(PRIMARY_KEY)["recorded_study_rows"].sort_index()
    observed = rows.groupby(PRIMARY_KEY).size().sort_index()
    if not observed.equals(expected.astype(int)):
        raise AssertionError("Orientation row counts do not match the 11-group audit")
    group_check = rows.drop_duplicates(PRIMARY_KEY)
    aggregate_pairs = [
        ("aggregate_tp", "recorded_aggregate_tp"),
        ("aggregate_fp", "recorded_aggregate_fp"),
        ("aggregate_fn", "recorded_aggregate_fn"),
        ("aggregate_tn", "recorded_aggregate_tn"),
        ("aggregate_sensitivity", "recorded_aggregate_sensitivity"),
        ("aggregate_specificity", "recorded_aggregate_specificity"),
        ("aggregate_youden_j", "recorded_aggregate_youden_j"),
    ]
    for regenerated, recorded in aggregate_pairs:
        if not np.allclose(group_check[regenerated], group_check[recorded], atol=1e-12, rtol=0):
            raise AssertionError(f"Regenerated orientation field {regenerated} differs from locked audit")
    if not group_check["aggregate_youden_j"].lt(0).all():
        raise AssertionError("Orientation audit contains a group that is not below chance")
    if not group_check["audit_review_title"].eq(group_check["review_title"]).all():
        raise AssertionError("Orientation audit review titles differ from normalized rows")
    if not group_check["audit_test_name"].eq(group_check["test_name"]).all():
        raise AssertionError("Orientation audit test names differ from normalized rows")
    summary = {
        "analysis_version": VERSION,
        "groups": int(rows[PRIMARY_KEY].drop_duplicates().shape[0]),
        "study_rows": len(rows),
        "four_cell_values_regenerated": int(len(rows) * 4),
        "aggregate_fields_regenerated_and_matched": len(aggregate_pairs),
        "all_groups_below_chance_from_regenerated_cells": bool(group_check["aggregate_youden_j"].lt(0).all()),
        "parser_errors_found": int(csv_bool(audit["recorded_parser_error_found"]).sum()),
        "parser_transformation_status": "none documented in locked normalized fields; source_status is retained separately as source_value_status",
        "target_label_status": "no separate target-label field in locked normalized data",
        "automatic_flips": 0,
        "decision": "retain source-defined direction and exclude below-chance groups without automatic flipping",
    }
    return rows, summary


def _route_summary(points: pd.DataFrame, label_column: str) -> pd.DataFrame:
    records: list[dict[str, object]] = []
    for label, group in points.groupby(label_column, sort=True):
        for prior in (0.05, 0.20, 0.50):
            metrics = diagnostic_metrics(
                group["sensitivity"].to_numpy(), group["specificity"].to_numpy(), prior
            )
            info = np.asarray(metrics["information_bits"], dtype=float)
            relative = np.asarray(metrics["diagnostic_entropy_reduction"], dtype=float)
            youden = group["sensitivity"].to_numpy() + group["specificity"].to_numpy() - 1
            records.append(
                {
                    "cohort": label,
                    "starting_probability": prior,
                    "reviews": int(group["review_id"].nunique()),
                    "operating_points": len(group),
                    "study_rows": int(group["study_rows"].sum()),
                    "information_bits_median": float(np.median(info)),
                    "information_bits_q1": float(np.quantile(info, 0.25)),
                    "information_bits_q3": float(np.quantile(info, 0.75)),
                    "relative_information_median": float(np.median(relative)),
                    "relative_information_q1": float(np.quantile(relative, 0.25)),
                    "relative_information_q3": float(np.quantile(relative, 0.75)),
                    "spearman_information_vs_youden": float(pd.Series(info).corr(pd.Series(youden), method="spearman")),
                    "zero_cell_study_fraction_median": float(group["zero_cell_study_fraction"].median()),
                    "operating_points_with_any_zero_cell_study": int(group["zero_cell_study_fraction"].gt(0).sum()),
                }
            )
    return pd.DataFrame(records)


def route_stability(root: Path, canonical: pd.DataFrame) -> tuple[pd.DataFrame, pd.DataFrame, pd.DataFrame]:
    points = canonical.copy()
    points["source_route"] = source_route(points["source"])
    points["historical_cohort"] = np.where(points["source"].eq("PMC Open Access"), "PMC", "pre-PMC")
    route = _route_summary(points, "source_route")
    historical = _route_summary(points, "historical_cohort")
    leaveout_records: list[pd.DataFrame] = []
    for held_out in sorted(points["source_route"].unique()):
        subset = points.loc[~points["source_route"].eq(held_out)].copy()
        subset["leave_one_route_out"] = f"excluding {held_out}"
        leaveout_records.append(_route_summary(subset, "leave_one_route_out"))
    leaveout = pd.concat(leaveout_records, ignore_index=True)
    return route, historical, leaveout


def source_trace(root: Path, canonical: pd.DataFrame) -> tuple[pd.DataFrame, dict[str, object]]:
    primary, analysis_rows, _audit = inventory(root)
    corpus = pd.read_csv(root / "analysis/full_atlas/corpus_manifest.csv")
    corpus_meta = corpus[["source", "review_id", "source_file", "source_sha256", "source_version"]].rename(
        columns={
            "source_file": "corpus_source_file",
            "source_sha256": "corpus_source_sha256",
            "source_version": "corpus_source_version",
        }
    )
    primary_meta = primary[PRIMARY_KEY + ["source_version", "selection_status", "source_url", "source_sha256", "source_file"]].rename(
        columns={
            "source_url": "group_source_url",
            "source_sha256": "group_source_sha256",
            "source_file": "group_source_file",
        }
    )
    trace = analysis_rows.merge(primary_meta, on=PRIMARY_KEY, how="left", validate="many_to_one")
    trace = trace.merge(corpus_meta, on=["source", "review_id"], how="left", validate="many_to_one")
    trace["operating_point_id"] = trace[PRIMARY_KEY].astype(str).agg("|".join, axis=1)
    trace["source_route"] = source_route(trace["source"])
    trace["source_identity_level"] = np.where(
        trace["group_source_url"].notna(), "group", "archive_or_package"
    )
    trace["effective_source_file"] = trace["group_source_file"].fillna(trace["corpus_source_file"])
    trace["effective_source_sha256"] = trace["group_source_sha256"].fillna(trace["corpus_source_sha256"])
    trace["source_identity"] = (
        trace["source"].fillna("") + "|" + trace["corpus_source_version"].fillna(trace["source_version"]).fillna("")
        + "|" + trace["effective_source_file"].fillna("") + "|" + trace["effective_source_sha256"].fillna("")
    )
    trace["primary_selection_decision"] = "manifest primary_atlas true; canonical manifest-sheet row"
    trace["operating_point_result_file"] = "analysis/bmj_rmr_v042/operating_points_primary.csv"
    trace["claim_ids"] = "C001;C002;C006;C011"
    trace["analysis_version"] = VERSION
    trace["group_url_availability"] = np.where(
        trace["group_source_url"].notna(), "available", "unavailable; not inferred"
    )
    columns = [
        "analysis_version", "operating_point_id", "source", "source_route", "review_id",
        "review_title", "group_id", "test_name", "study_id", "tp", "fp", "fn", "tn",
        "source_status", "source_file", "source_sheet_row", "source_row", "group_source_url",
        "group_url_availability", "group_source_sha256", "effective_source_file",
        "effective_source_sha256", "source_identity_level", "source_identity",
        "primary_selection_decision", "operating_point_result_file", "claim_ids",
    ]
    trace = trace[columns].sort_values(PRIMARY_KEY + ["study_id", "source_row"]).reset_index(drop=True)
    summary = {
        "analysis_version": VERSION,
        "study_rows": len(trace),
        "operating_points": int(trace["operating_point_id"].nunique()),
        "operating_points_with_source_identity": int(
            trace.groupby("operating_point_id")["source_identity"].first().str.len().gt(4).sum()
        ),
        "operating_points_with_group_url": int(
            trace.groupby("operating_point_id")["group_source_url"].first().notna().sum()
        ),
        "operating_points_without_group_url": int(
            trace.groupby("operating_point_id")["group_source_url"].first().isna().sum()
        ),
        "policy": "Unavailable group-level URLs remain missing; archive or package identity and checksum are retained instead.",
    }
    if summary["study_rows"] != 4104 or summary["operating_points"] != 273:
        raise AssertionError("End-to-end primary source trace is incomplete")
    if summary["operating_points_with_source_identity"] != 273:
        raise AssertionError("One or more primary points lacks source identity")
    return trace, summary


def phase2(root: Path) -> dict[str, object]:
    analysis_dir = root / "analysis/bmj_rmr_v042"
    qa_dir = analysis_dir / "qa"
    canonical = pd.read_csv(analysis_dir / "operating_points_primary.csv")
    fidelity, fidelity_summary = source_summary_fidelity(root, canonical)
    fidelity.to_csv(analysis_dir / "source_summary_fidelity.csv", index=False, lineterminator="\n")
    write_json(analysis_dir / "source_summary_fidelity_summary.json", fidelity_summary)
    fidelity.loc[
        fidelity["large_discrepancy"],
        [
            *PRIMARY_KEY,
            "test_name",
            "published_studies",
            "study_rows",
            "published_sensitivity",
            "sensitivity",
            "absolute_sensitivity_difference",
            "published_specificity",
            "specificity",
            "absolute_specificity_difference",
            "explanation_from_available_metadata",
        ],
    ].to_csv(
        analysis_dir / "source_summary_large_discrepancies.csv",
        index=False,
        lineterminator="\n",
    )

    manual, manual_checksums, manual_summary = manual_audit_verification(root)
    manual.to_csv(analysis_dir / "manual_audit_verification.csv", index=False, lineterminator="\n")
    manual_checksums.to_csv(
        analysis_dir / "manual_audit_input_checksums.csv", index=False, lineterminator="\n"
    )
    write_json(analysis_dir / "manual_audit_summary.json", manual_summary)

    orientation, orientation_summary = orientation_study_audit(root)
    orientation.to_csv(
        analysis_dir / "orientation_audit_study_rows.csv", index=False, lineterminator="\n"
    )
    write_json(analysis_dir / "orientation_audit_summary.json", orientation_summary)

    route, historical, leaveout = route_stability(root, canonical)
    route.to_csv(analysis_dir / "source_route_stability.csv", index=False, lineterminator="\n")
    historical.to_csv(
        analysis_dir / "historical_cohort_stability.csv", index=False, lineterminator="\n"
    )
    leaveout.to_csv(
        analysis_dir / "leave_one_route_out_stability.csv", index=False, lineterminator="\n"
    )

    trace, trace_summary = source_trace(root, canonical)
    trace.to_csv(analysis_dir / "source_trace_all_primary.csv", index=False, lineterminator="\n")
    write_json(analysis_dir / "source_trace_summary.json", trace_summary)

    claim_path = root / "docs/manuscript/v19/CLAIM_LEDGER.csv"
    claims = pd.read_csv(claim_path)
    updates = {
        "C003": {
            "source_file": "analysis/bmj_rmr_v042/source_summary_fidelity.csv",
            "source_row_or_variable": "92 regenerated canonical comparisons",
            "source_validation_status": "joined to source-reported summaries and regenerated in Phase 2",
            "automated_check": "test_phase2_source_summary_fidelity_is_regenerated_for_all_92_points",
            "final_status": "validated Phase 2; pending prose",
        },
        "C004": {
            "claim_text": "The locked author audit contains 30 extracted tables (27 included and 3 orientation safeguards), 410 normalized rows, and 1640 checked cells with no recorded transcription errors, plus 50 accept-or-reject decisions with no recorded acceptance errors",
            "source_file": "analysis/bmj_rmr_v042/manual_audit_verification.csv",
            "source_row_or_variable": "50 locked author-audit rows",
            "source_validation_status": "locked inputs unchanged; recorded JATS-hash consistency and denominators verified; raw JATS not stored",
            "automated_check": "test_phase2_locked_manual_audit_has_exact_denominators_and_hash_scope",
            "final_status": "hash and denominator verified Phase 2; pending prose",
        },
        "C005": {
            "source_file": "analysis/bmj_rmr_v042/orientation_audit_study_rows.csv",
            "source_row_or_variable": "all 11 groups and 86 contributing study rows",
            "source_validation_status": "joined to normalized rows and source identities in Phase 2",
            "automated_check": "test_phase2_orientation_audit_preserves_all_rows_without_flipping",
            "final_status": "regenerated Phase 2; pending prose",
        },
        "C006": {
            "source_validation_status": "canonical rows and end-to-end source identities regenerated in Phase 2",
        },
    }
    for claim_id, values in updates.items():
        for column, value in values.items():
            claims.loc[claims["claim_id"].eq(claim_id), column] = value
    if "C010" not in set(claims["claim_id"]):
        claims.loc[len(claims)] = {
            "claim_id": "C010",
            "claim_text": "Route-specific summaries describe reproducibility of broad patterns without claiming representativeness",
            "manuscript_location": "Results and Supplement",
            "claim_role": "supporting",
            "source_file": "analysis/bmj_rmr_v042/source_route_stability.csv",
            "source_row_or_variable": "all five routes at 0.05 0.20 and 0.50",
            "estimator": "continuity-corrected bivariate REML",
            "starting_probability": "0.05;0.20;0.50",
            "uncertainty_type": "descriptive distribution",
            "source_validation_status": "Phase 2 regenerated",
            "model_stability_status": "descriptive only",
            "automated_check": "test_phase2_route_stability_covers_declared_routes_and_anchors",
            "final_status": "pending prose",
        }
    else:
        claims.loc[claims["claim_id"].eq("C010"), "automated_check"] = (
            "test_phase2_route_stability_covers_declared_routes_and_anchors"
        )
    if "C011" not in set(claims["claim_id"]):
        claims.loc[len(claims)] = {
            "claim_id": "C011",
            "claim_text": "All 4104 canonical study rows are traceable to 273 primary operating points; 181 points have group-level URLs and 92 retain archive-level identity without inferred URLs",
            "manuscript_location": "Methods and Supplement",
            "claim_role": "supporting",
            "source_file": "analysis/bmj_rmr_v042/source_trace_summary.json",
            "source_row_or_variable": "study_rows and operating_points_with_group_url",
            "estimator": "not applicable",
            "starting_probability": "not applicable",
            "uncertainty_type": "not applicable",
            "source_validation_status": "Phase 2 end-to-end trace regenerated",
            "model_stability_status": "not applicable",
            "automated_check": "test_phase2_trace_has_one_record_per_canonical_analysis_row",
            "final_status": "pending prose",
        }
    claims.to_csv(claim_path, index=False, lineterminator="\n")

    output_names = [
        "source_summary_fidelity.csv", "source_summary_fidelity_summary.json",
        "source_summary_large_discrepancies.csv",
        "manual_audit_verification.csv", "manual_audit_input_checksums.csv",
        "manual_audit_summary.json", "orientation_audit_study_rows.csv",
        "orientation_audit_summary.json", "source_route_stability.csv",
        "historical_cohort_stability.csv", "leave_one_route_out_stability.csv",
        "source_trace_all_primary.csv", "source_trace_summary.json",
    ]
    gate = {
        "phase": 2,
        "name": "source validation and traceability",
        "status": "candidate_pending_independent_qa",
        "checks": {
            "source_summary_comparisons_exact": len(fidelity) == 92,
            "large_source_summary_discrepancies_have_metadata_specific_explanations": bool(
                fidelity.loc[fidelity["large_discrepancy"], "explanation_from_available_metadata"]
                .str.contains("Available metadata:", regex=False).all()
            ),
            "manual_audit_locked_inputs_unchanged": bool(manual_checksums["unchanged_from_orientation_source"].all()),
            "manual_audit_selection_rule_script_locked": bool(
                "scripts/cochrane_dta_atlas.py" in set(manual_checksums["file"])
            ),
            "manual_audit_all_50_rows_verified": bool(len(manual) == 50 and manual["verification_pass"].all()),
            "manual_audit_error_denominators_explicit": bool(
                manual_summary["transcription_error_denominator_cells"] == 1640
                and manual_summary["acceptance_error_denominator_decisions"] == 50
            ),
            "orientation_11_groups_expanded": orientation_summary["groups"] == 11,
            "orientation_aggregates_regenerated": bool(
                orientation_summary["aggregate_fields_regenerated_and_matched"] == 7
            ),
            "orientation_missing_target_label_explicit": bool(
                orientation["target_label_where_recorded"].isna().all()
            ),
            "no_orientation_flip": orientation_summary["automatic_flips"] == 0,
            "all_five_routes_and_three_anchors": bool(len(route) == 15),
            "all_273_points_traceable": trace_summary["operating_points_with_source_identity"] == 273,
            "all_4104_analysis_rows_traceable": trace_summary["study_rows"] == 4104,
            "missing_group_urls_not_inferred": bool(
                trace.loc[trace["group_url_availability"].eq("unavailable; not inferred"), "group_source_url"].isna().all()
            ),
            "trace_links_end_to_end_claim": bool(trace["claim_ids"].str.contains("C011", regex=False).all()),
        },
        "fidelity_summary": fidelity_summary,
        "manual_audit_summary": manual_summary,
        "orientation_summary": orientation_summary,
        "trace_summary": trace_summary,
        "commands": [
            "py -3.13 scripts/bmj_rmr_v042.py phase2",
            "py -3.13 -m pytest tests/test_bmj_rmr_v042.py tests/test_bmj_rmr_v042_spec.py tests/test_cochrane_dta_atlas.py -q",
        ],
        "output_sha256": {name: file_hash(analysis_dir / name) for name in output_names},
        "qa_context": "pending fresh data-provenance and extraction audit",
    }
    gate["output_sha256"]["CLAIM_LEDGER.csv"] = file_hash(claim_path)
    write_json(qa_dir / "phase_gate_2.json", gate)
    run_manifest_path = analysis_dir / "run_manifest.json"
    run_manifest = json.loads(run_manifest_path.read_text(encoding="utf-8"))
    run_manifest.update(
        {
            "phase": 2,
            "status": "phase_2_candidate_pending_independent_qa",
            "role": "forward completion with estimator locked and source validation regenerated",
            "phase_2_gate": "analysis/bmj_rmr_v042/qa/phase_gate_2.json",
        }
    )
    write_json(run_manifest_path, run_manifest)
    return gate


def binary_entropy(probability: np.ndarray | float) -> np.ndarray:
    """Binary entropy with exact zero at the closed-interval boundaries."""

    probability_array = np.asarray(probability, dtype=float)
    clipped = np.clip(
        probability_array, np.finfo(float).tiny, 1.0 - np.finfo(float).eps
    )
    entropy = -(
        clipped * np.log2(clipped)
        + (1.0 - clipped) * np.log2(1.0 - clipped)
    )
    return np.where(
        (probability_array <= 0.0) | (probability_array >= 1.0), 0.0, entropy
    )


def expanded_diagnostic_metrics(
    sensitivity: np.ndarray | float,
    specificity: np.ndarray | float,
    starting_probability: np.ndarray | float,
) -> dict[str, np.ndarray]:
    """Return the complete four-question probability and entropy derivation."""

    base = diagnostic_metrics(sensitivity, specificity, starting_probability)
    prior = np.asarray(starting_probability, dtype=float)
    baseline_entropy = binary_entropy(prior)
    positive_probability = np.asarray(base["positive_probability"], dtype=float)
    negative_probability = 1.0 - positive_probability
    positive_posterior = np.asarray(base["positive_posterior"], dtype=float)
    negative_posterior = np.asarray(base["negative_posterior"], dtype=float)
    positive_entropy = binary_entropy(positive_posterior)
    negative_entropy = binary_entropy(negative_posterior)
    metric_shape = np.asarray(base["information_bits"]).shape
    return {
        "lr_positive": np.broadcast_to(
            np.asarray(base["lr_positive"], dtype=float), metric_shape
        ),
        "lr_negative": np.broadcast_to(
            np.asarray(base["lr_negative"], dtype=float), metric_shape
        ),
        "youden_j": np.broadcast_to(
            np.asarray(base["youden"], dtype=float), metric_shape
        ),
        "positive_result_probability": positive_probability,
        "negative_result_probability": negative_probability,
        "post_positive_probability": positive_posterior,
        "post_negative_probability": negative_posterior,
        "information_bits": np.asarray(base["information_bits"], dtype=float),
        "fraction_entropy_removed": np.asarray(
            base["diagnostic_entropy_reduction"], dtype=float
        ),
        "positive_branch_entropy_bits": positive_entropy,
        "negative_branch_entropy_bits": negative_entropy,
        "expected_posterior_entropy_bits": (
            positive_probability * positive_entropy
            + negative_probability * negative_entropy
        ),
        "positive_entropy_change_bits": positive_entropy - baseline_entropy,
        "negative_entropy_change_bits": negative_entropy - baseline_entropy,
        "positive_information_contribution_bits": np.asarray(
            base["positive_result_information_bits"], dtype=float
        ),
        "negative_information_contribution_bits": np.asarray(
            base["negative_result_information_bits"], dtype=float
        ),
    }


DERIVED_METRICS = (
    "lr_positive",
    "lr_negative",
    "youden_j",
    "positive_result_probability",
    "negative_result_probability",
    "post_positive_probability",
    "post_negative_probability",
    "information_bits",
    "fraction_entropy_removed",
    "positive_branch_entropy_bits",
    "negative_branch_entropy_bits",
    "expected_posterior_entropy_bits",
    "positive_entropy_change_bits",
    "negative_entropy_change_bits",
    "positive_information_contribution_bits",
    "negative_information_contribution_bits",
)

PROFILE_INTERVAL_METRICS = (
    "information_bits",
    "fraction_entropy_removed",
    "positive_result_probability",
    "negative_result_probability",
    "post_positive_probability",
    "post_negative_probability",
    "positive_branch_entropy_bits",
    "negative_branch_entropy_bits",
)


def canonical_uncertainty_draws(
    row: pd.Series, draws: int = DERIVED_DRAWS
) -> tuple[np.ndarray, np.ndarray, np.ndarray, np.ndarray, int]:
    """Recreate the locked Phase 1 REML draws without refitting the model."""

    mean = np.asarray(
        [row["mean_logit_sensitivity"], row["mean_logit_specificity"]], dtype=float
    )
    mean_covariance = np.asarray(
        [
            [row["mean_logit_sensitivity_variance"], row["mean_logit_covariance"]],
            [row["mean_logit_covariance"], row["mean_logit_specificity_variance"]],
        ],
        dtype=float,
    )
    between_covariance = np.asarray(
        [
            [
                row["between_logit_sensitivity_variance"],
                row["between_logit_covariance"],
            ],
            [
                row["between_logit_covariance"],
                row["between_logit_specificity_variance"],
            ],
        ],
        dtype=float,
    )
    seed = (
        REML_SEED
        + int(hashlib.sha256(str(row["group_id"]).encode()).hexdigest()[:8], 16)
    ) % (2**32)
    rng = np.random.default_rng(seed)
    pooled_logits = rng.multivariate_normal(mean, mean_covariance, size=draws)
    predictive_logits = rng.multivariate_normal(
        mean, mean_covariance + between_covariance, size=draws
    )
    return (
        expit(pooled_logits[:, 0]),
        expit(pooled_logits[:, 1]),
        expit(predictive_logits[:, 0]),
        expit(predictive_logits[:, 1]),
        seed,
    )


def _quantile_columns(
    record: dict[str, object],
    prefix: str,
    metrics: dict[str, np.ndarray],
    index: int,
    names: tuple[str, ...],
) -> None:
    for name in names:
        low, median, high = np.quantile(
            np.asarray(metrics[name], dtype=float)[:, index], [0.025, 0.5, 0.975]
        )
        record[f"{prefix}_{name}_low"] = float(low)
        record[f"{prefix}_{name}_median"] = float(median)
        record[f"{prefix}_{name}_high"] = float(high)


def canonical_derived_outputs(
    canonical: pd.DataFrame, draws: int = DERIVED_DRAWS
) -> tuple[pd.DataFrame, pd.DataFrame]:
    """Build standard-anchor and dense-grid outputs from one REML hierarchy."""

    anchors = np.asarray(STANDARD_ANCHORS, dtype=float)
    profiles = np.asarray(PROFILE_PROBABILITIES, dtype=float)
    anchor_records: list[dict[str, object]] = []
    profile_records: list[dict[str, object]] = []
    metadata_columns = [
        "source",
        "review_id",
        "review_title",
        "group_id",
        "test_name",
        "study_rows",
        "zero_cell_study_fraction",
        "model",
        "analysis_row_rule",
    ]
    for _, point in canonical.sort_values(PRIMARY_KEY).iterrows():
        metadata = {column: point[column] for column in metadata_columns}
        metadata["operating_point_id"] = "|".join(str(point[column]) for column in PRIMARY_KEY)
        metadata["analysis_version"] = VERSION
        metadata["estimator_role"] = "canonical"
        metadata["uncertainty_draws"] = draws
        (
            pooled_sensitivity,
            pooled_specificity,
            predictive_sensitivity,
            predictive_specificity,
            seed,
        ) = canonical_uncertainty_draws(point, draws=draws)
        metadata["uncertainty_seed"] = seed

        central_anchor = expanded_diagnostic_metrics(
            float(point["sensitivity"]), float(point["specificity"]), anchors
        )
        pooled_anchor = expanded_diagnostic_metrics(
            pooled_sensitivity[:, None], pooled_specificity[:, None], anchors[None, :]
        )
        predictive_anchor = expanded_diagnostic_metrics(
            predictive_sensitivity[:, None],
            predictive_specificity[:, None],
            anchors[None, :],
        )
        for index, prior in enumerate(anchors):
            record = dict(metadata)
            record["starting_probability"] = float(prior)
            record["starting_entropy_bits"] = float(binary_entropy(prior))
            record["sensitivity"] = float(point["sensitivity"])
            record["specificity"] = float(point["specificity"])
            for uncertainty in ("pooled_mean", "predictive"):
                for metric in ("sensitivity", "specificity"):
                    for quantile in ("low", "median", "high"):
                        column = f"{uncertainty}_{metric}_{quantile}"
                        record[column] = float(point[column])
            for name in DERIVED_METRICS:
                record[name] = float(np.asarray(central_anchor[name])[index])
            _quantile_columns(
                record, "pooled_mean", pooled_anchor, index, DERIVED_METRICS
            )
            _quantile_columns(
                record, "predictive", predictive_anchor, index, DERIVED_METRICS
            )
            anchor_records.append(record)

        central_profile = expanded_diagnostic_metrics(
            float(point["sensitivity"]), float(point["specificity"]), profiles
        )
        pooled_profile = expanded_diagnostic_metrics(
            pooled_sensitivity[:, None], pooled_specificity[:, None], profiles[None, :]
        )
        predictive_profile = expanded_diagnostic_metrics(
            predictive_sensitivity[:, None],
            predictive_specificity[:, None],
            profiles[None, :],
        )
        for index, prior in enumerate(profiles):
            record = dict(metadata)
            record["starting_probability"] = float(prior)
            record["starting_entropy_bits"] = float(binary_entropy(prior))
            record["sensitivity"] = float(point["sensitivity"])
            record["specificity"] = float(point["specificity"])
            for name in PROFILE_INTERVAL_METRICS:
                record[name] = float(np.asarray(central_profile[name])[index])
            record["positive_entropy_change_bits"] = float(
                np.asarray(central_profile["positive_entropy_change_bits"])[index]
            )
            record["negative_entropy_change_bits"] = float(
                np.asarray(central_profile["negative_entropy_change_bits"])[index]
            )
            _quantile_columns(
                record,
                "pooled_mean",
                pooled_profile,
                index,
                PROFILE_INTERVAL_METRICS,
            )
            _quantile_columns(
                record,
                "predictive",
                predictive_profile,
                index,
                PROFILE_INTERVAL_METRICS,
            )
            profile_records.append(record)
    anchor_frame = pd.DataFrame(anchor_records).sort_values(
        PRIMARY_KEY + ["starting_probability"]
    )
    profile_frame = pd.DataFrame(profile_records).sort_values(
        PRIMARY_KEY + ["starting_probability"]
    )
    if len(anchor_frame) != 273 * len(STANDARD_ANCHORS):
        raise AssertionError("Standard-anchor derivation is incomplete")
    if len(profile_frame) != 273 * len(PROFILE_PROBABILITIES):
        raise AssertionError("Continuous probability profiles are incomplete")
    return anchor_frame.reset_index(drop=True), profile_frame.reset_index(drop=True)


def anchor_and_branch_summaries(
    anchors: pd.DataFrame,
) -> tuple[pd.DataFrame, pd.DataFrame, pd.DataFrame]:
    summary_records: list[dict[str, object]] = []
    for prior, group in anchors.groupby("starting_probability", sort=True):
        summary_records.append(
            {
                "analysis_version": VERSION,
                "estimator": "continuity-corrected bivariate REML",
                "starting_probability": float(prior),
                "starting_entropy_bits": float(group["starting_entropy_bits"].iloc[0]),
                "operating_points": len(group),
                "reviews": int(group["review_id"].nunique()),
                "information_bits_median": float(group["information_bits"].median()),
                "information_bits_q1": float(group["information_bits"].quantile(0.25)),
                "information_bits_q3": float(group["information_bits"].quantile(0.75)),
                "fraction_entropy_removed_median": float(
                    group["fraction_entropy_removed"].median()
                ),
                "fraction_entropy_removed_q1": float(
                    group["fraction_entropy_removed"].quantile(0.25)
                ),
                "fraction_entropy_removed_q3": float(
                    group["fraction_entropy_removed"].quantile(0.75)
                ),
                "post_negative_probability_median": float(
                    group["post_negative_probability"].median()
                ),
                "spearman_information_vs_youden_j": float(
                    group[["information_bits", "youden_j"]]
                    .corr(method="spearman")
                    .iloc[0, 1]
                ),
                "positive_result_increases_entropy_count": int(
                    group["positive_entropy_change_bits"].gt(0).sum()
                ),
                "negative_result_increases_entropy_count": int(
                    group["negative_entropy_change_bits"].gt(0).sum()
                ),
                "interpretation": "Cross-diagnosis descriptive distribution; branch entropy movement is not clinical benefit or harm.",
            }
        )
    summary = pd.DataFrame(summary_records)
    branch_columns = [
        "analysis_version",
        "operating_point_id",
        "source",
        "review_id",
        "review_title",
        "group_id",
        "test_name",
        "model",
        "starting_probability",
        "starting_entropy_bits",
        "positive_result_probability",
        "negative_result_probability",
        "post_positive_probability",
        "post_negative_probability",
        "positive_branch_entropy_bits",
        "negative_branch_entropy_bits",
        "positive_entropy_change_bits",
        "negative_entropy_change_bits",
        "information_bits",
        "positive_information_contribution_bits",
        "negative_information_contribution_bits",
    ]
    interval_stems = (
        "positive_branch_entropy_bits",
        "negative_branch_entropy_bits",
        "post_positive_probability",
        "post_negative_probability",
    )
    for uncertainty in ("pooled_mean", "predictive"):
        for stem in interval_stems:
            branch_columns.extend(
                f"{uncertainty}_{stem}_{quantile}"
                for quantile in ("low", "median", "high")
            )
    branches = anchors[branch_columns].copy()
    branches["positive_branch_entropy_movement"] = np.select(
        [
            branches["positive_entropy_change_bits"].gt(1e-12),
            branches["positive_entropy_change_bits"].lt(-1e-12),
        ],
        ["increase", "decrease"],
        default="no numerical change",
    )
    branches["negative_branch_entropy_movement"] = np.select(
        [
            branches["negative_entropy_change_bits"].gt(1e-12),
            branches["negative_entropy_change_bits"].lt(-1e-12),
        ],
        ["increase", "decrease"],
        default="no numerical change",
    )
    branch_summary = (
        branches.melt(
            id_vars=["starting_probability"],
            value_vars=[
                "positive_branch_entropy_movement",
                "negative_branch_entropy_movement",
            ],
            var_name="result_branch",
            value_name="entropy_movement",
        )
        .groupby(["starting_probability", "result_branch", "entropy_movement"])
        .size()
        .rename("operating_points")
        .reset_index()
    )
    branch_summary["analysis_version"] = VERSION
    branch_summary["estimator"] = "continuity-corrected bivariate REML"
    branch_summary["interpretation"] = (
        "Branch-specific entropy movement after an observed result; not a clinical benefit or harm count."
    )
    return summary, branches, branch_summary


def landmark_table(anchors: pd.DataFrame) -> pd.DataFrame:
    at_twenty = anchors.loc[np.isclose(anchors["starting_probability"], 0.20)].set_index(
        "group_id"
    )
    rows: list[dict[str, object]] = []
    for group_id, label in LANDMARKS:
        point = at_twenty.loc[group_id]
        row = {
            "analysis_version": VERSION,
            "label": label,
            "source": point["source"],
            "review_id": point["review_id"],
            "review_title": point["review_title"],
            "group_id": group_id,
            "test_name": point["test_name"],
            "studies": int(point["study_rows"]),
            "estimator": point["model"],
            "starting_probability": 0.20,
            "sensitivity": point["sensitivity"],
            "specificity": point["specificity"],
            "information_bits": point["information_bits"],
            "fraction_entropy_removed": point["fraction_entropy_removed"],
            "post_positive_probability": point["post_positive_probability"],
            "post_negative_probability": point["post_negative_probability"],
            "landmark_role": "scale orientation; not a ranking of utility",
        }
        for uncertainty in ("pooled_mean", "predictive"):
            for metric in (
                "information_bits",
                "fraction_entropy_removed",
                "post_positive_probability",
                "post_negative_probability",
            ):
                for quantile in ("low", "median", "high"):
                    column = f"{uncertainty}_{metric}_{quantile}"
                    row[column] = point[column]
        rows.append(row)
    return pd.DataFrame(rows)


def crossing_roots(
    sensitivity_a: float,
    specificity_a: float,
    sensitivity_b: float,
    specificity_b: float,
    low: float = 0.005,
    high: float = 0.995,
) -> list[float]:
    """Find non-boundary information-order roots on a deterministic grid."""

    def difference(prior: float) -> float:
        left = expanded_diagnostic_metrics(
            sensitivity_a, specificity_a, prior
        )["information_bits"]
        right = expanded_diagnostic_metrics(
            sensitivity_b, specificity_b, prior
        )["information_bits"]
        return float(left - right)

    grid = np.linspace(low, high, 1981)
    values = np.asarray([difference(value) for value in grid])
    roots: list[float] = []
    for index in range(len(grid) - 1):
        if abs(values[index]) <= 1e-12:
            roots.append(float(grid[index]))
        elif values[index] * values[index + 1] < 0:
            roots.append(
                float(brentq(difference, float(grid[index]), float(grid[index + 1])))
            )
    if abs(values[-1]) <= 1e-12:
        roots.append(float(grid[-1]))
    unique: list[float] = []
    for root in sorted(roots):
        if not unique or abs(root - unique[-1]) > 1e-7:
            unique.append(root)
    return unique


def theoretical_non_equivalence() -> tuple[pd.DataFrame, pd.DataFrame, dict[str, object]]:
    examples = {
        "equal_youden_different_information": {
            "classification": "theoretical",
            "starting_probability": 0.20,
            "tests": (("A", 0.80, 0.80), ("B", 0.95, 0.65)),
        },
        "near_equal_information_different_post_negative": {
            "classification": "theoretical",
            "starting_probability": 0.20,
            "tests": (("A", 0.99, 0.64), ("B", 0.50, 0.99)),
        },
    }
    rows: list[dict[str, object]] = []
    for example_id, example in examples.items():
        prior = float(example["starting_probability"])
        for test_label, sensitivity, specificity in example["tests"]:
            metrics = expanded_diagnostic_metrics(sensitivity, specificity, prior)
            rows.append(
                {
                    "analysis_version": VERSION,
                    "example_id": example_id,
                    "classification": example["classification"],
                    "test_label": test_label,
                    "starting_probability": prior,
                    "sensitivity": sensitivity,
                    "specificity": specificity,
                    "youden_j": float(metrics["youden_j"]),
                    "information_bits": float(metrics["information_bits"]),
                    "fraction_entropy_removed": float(
                        metrics["fraction_entropy_removed"]
                    ),
                    "post_positive_probability": float(
                        metrics["post_positive_probability"]
                    ),
                    "post_negative_probability": float(
                        metrics["post_negative_probability"]
                    ),
                    "display_probability_is_action_threshold": False,
                }
            )

    crossing_tests = (("A", 0.70, 0.95), ("B", 0.95, 0.75))
    roots = crossing_roots(0.70, 0.95, 0.95, 0.75)
    crossing = next(root for root in roots if 0.01 <= root <= 0.50)
    for test_label, sensitivity, specificity in crossing_tests:
        metrics = expanded_diagnostic_metrics(sensitivity, specificity, crossing)
        rows.append(
            {
                "analysis_version": VERSION,
                "example_id": "probability_dependent_information_crossing",
                "classification": "theoretical",
                "test_label": test_label,
                "starting_probability": crossing,
                "sensitivity": sensitivity,
                "specificity": specificity,
                "youden_j": float(metrics["youden_j"]),
                "information_bits": float(metrics["information_bits"]),
                "fraction_entropy_removed": float(metrics["fraction_entropy_removed"]),
                "post_positive_probability": float(
                    metrics["post_positive_probability"]
                ),
                "post_negative_probability": float(
                    metrics["post_negative_probability"]
                ),
                "display_probability_is_action_threshold": False,
            }
        )
    table = pd.DataFrame(rows)

    profile_rows: list[dict[str, object]] = []
    for test_label, sensitivity, specificity in crossing_tests:
        metrics = expanded_diagnostic_metrics(
            sensitivity, specificity, np.asarray(PROFILE_PROBABILITIES)
        )
        for index, prior in enumerate(PROFILE_PROBABILITIES):
            profile_rows.append(
                {
                    "analysis_version": VERSION,
                    "example_id": "probability_dependent_information_crossing",
                    "classification": "theoretical",
                    "test_label": test_label,
                    "sensitivity": sensitivity,
                    "specificity": specificity,
                    "starting_probability": prior,
                    "information_bits": float(metrics["information_bits"][index]),
                    "fraction_entropy_removed": float(
                        metrics["fraction_entropy_removed"][index]
                    ),
                    "post_negative_probability": float(
                        metrics["post_negative_probability"][index]
                    ),
                    "numerical_crossing_probability": crossing,
                }
            )
    profile = pd.DataFrame(profile_rows)
    equal_j = table.loc[
        table["example_id"].eq("equal_youden_different_information")
    ]
    equal_information = table.loc[
        table["example_id"].eq("near_equal_information_different_post_negative")
    ]
    crossing_rows = table.loc[
        table["example_id"].eq("probability_dependent_information_crossing")
    ]
    summary = {
        "analysis_version": VERSION,
        "classification": "theoretical",
        "equal_youden_different_information": {
            "absolute_youden_difference": float(equal_j["youden_j"].max() - equal_j["youden_j"].min()),
            "absolute_information_difference_bits": float(
                equal_j["information_bits"].max() - equal_j["information_bits"].min()
            ),
        },
        "near_equal_information_different_post_negative": {
            "absolute_information_difference_bits": float(
                equal_information["information_bits"].max()
                - equal_information["information_bits"].min()
            ),
            "absolute_post_negative_difference": float(
                equal_information["post_negative_probability"].max()
                - equal_information["post_negative_probability"].min()
            ),
            "information_tolerance_bits": 0.0001,
            "required_post_negative_difference": 0.10,
        },
        "probability_dependent_information_crossing": {
            "numerical_root": crossing,
            "root_residual_bits": float(
                crossing_rows["information_bits"].max()
                - crossing_rows["information_bits"].min()
            ),
            "roots_between_0_005_and_0_995": roots,
            "display_probability_is_action_threshold": False,
        },
    }
    return table, profile, summary


def _point_specifications(root: Path, canonical: pd.DataFrame) -> list[dict[str, object]]:
    return [
        {
            "model_specification": "v0.42.0 canonical REML",
            "analysis_role": "canonical primary",
            "sensitivity_dimension": "canonical",
            "frame": canonical,
            "sensitivity_column": "sensitivity",
            "specificity_column": "specificity",
        },
        {
            "model_specification": "v0.39.0 historical REML",
            "analysis_role": "historical release sensitivity",
            "sensitivity_dimension": "release history",
            "frame": pd.read_csv(
                root / "analysis/full_atlas/operating_points_20_percent.csv"
            ),
            "sensitivity_column": "sensitivity",
            "specificity_column": "specificity",
        },
        {
            "model_specification": "v0.40.0/v0.41.0 regularized GH15",
            "analysis_role": "model-family sensitivity",
            "sensitivity_dimension": "estimator",
            "frame": pd.read_csv(
                root / "analysis/bmj_rmr_v040/operating_points_regularized.csv"
            ),
            "sensitivity_column": "pooled_sensitivity_median",
            "specificity_column": "pooled_specificity_median",
        },
        {
            "model_specification": "regularized GH15 wider prior",
            "analysis_role": "prior sensitivity",
            "sensitivity_dimension": "prior",
            "frame": pd.read_csv(
                root / "analysis/bmj_rmr_v040/operating_points_wide_prior.csv"
            ),
            "sensitivity_column": "pooled_sensitivity_median",
            "specificity_column": "pooled_specificity_median",
        },
        {
            "model_specification": "regularized GH9 primary prior",
            "analysis_role": "numerical sensitivity",
            "sensitivity_dimension": "quadrature",
            "frame": pd.read_csv(
                root
                / "analysis/bmj_rmr_v040_gh9_validation/operating_points_regularized.csv"
            ),
            "sensitivity_column": "pooled_sensitivity_median",
            "specificity_column": "pooled_specificity_median",
        },
    ]


def empirical_cea_crossing(
    root: Path, canonical: pd.DataFrame
) -> tuple[pd.DataFrame, pd.DataFrame]:
    crossing_rows: list[dict[str, object]] = []
    profile_rows: list[dict[str, object]] = []
    for specification in _point_specifications(root, canonical):
        frame = specification["frame"].set_index("group_id")
        low = frame.loc[CEA_LOW_ID]
        high = frame.loc[CEA_HIGH_ID]
        se_low = float(low[specification["sensitivity_column"]])
        sp_low = float(low[specification["specificity_column"]])
        se_high = float(high[specification["sensitivity_column"]])
        sp_high = float(high[specification["specificity_column"]])
        roots = crossing_roots(se_low, sp_low, se_high, sp_high)
        display_roots = [root for root in roots if 0.01 <= root <= 0.50]
        crossing = display_roots[0] if display_roots else math.nan
        crossing_rows.append(
            {
                "analysis_version": VERSION,
                "classification": "empirical model-specific sensitivity example",
                "model_specification": specification["model_specification"],
                "analysis_role": specification["analysis_role"],
                "sensitivity_dimension": specification["sensitivity_dimension"],
                "review_id": str(low["review_id"]),
                "low_threshold_group_id": CEA_LOW_ID,
                "low_threshold_test": str(low["test_name"]),
                "low_threshold_sensitivity": se_low,
                "low_threshold_specificity": sp_low,
                "high_threshold_group_id": CEA_HIGH_ID,
                "high_threshold_test": str(high["test_name"]),
                "high_threshold_sensitivity": se_high,
                "high_threshold_specificity": sp_high,
                "crossing_probability": crossing,
                "all_roots_between_0_005_and_0_995": ";".join(
                    f"{root:.12f}" for root in roots
                ),
                "crossing_in_display_range_0_01_to_0_50": bool(display_roots),
                "display_probability_is_action_threshold": False,
                "interpretation": "Empirical CEA illustration inherited from model estimates; not an action threshold.",
            }
        )
        for group_id, point, sensitivity, specificity in (
            (CEA_LOW_ID, low, se_low, sp_low),
            (CEA_HIGH_ID, high, se_high, sp_high),
        ):
            metrics = expanded_diagnostic_metrics(
                sensitivity, specificity, np.asarray(PROFILE_PROBABILITIES)
            )
            for index, prior in enumerate(PROFILE_PROBABILITIES):
                profile_rows.append(
                    {
                        "analysis_version": VERSION,
                        "classification": "empirical model-specific sensitivity example",
                        "model_specification": specification["model_specification"],
                        "analysis_role": specification["analysis_role"],
                        "sensitivity_dimension": specification["sensitivity_dimension"],
                        "review_id": str(point["review_id"]),
                        "group_id": group_id,
                        "test_name": str(point["test_name"]),
                        "starting_probability": prior,
                        "sensitivity": sensitivity,
                        "specificity": specificity,
                        "information_bits": float(metrics["information_bits"][index]),
                        "fraction_entropy_removed": float(
                            metrics["fraction_entropy_removed"][index]
                        ),
                        "post_negative_probability": float(
                            metrics["post_negative_probability"][index]
                        ),
                        "crossing_probability": crossing,
                    }
                )
    return pd.DataFrame(crossing_rows), pd.DataFrame(profile_rows)


def appendicitis_worked_profile(anchors: pd.DataFrame) -> pd.DataFrame:
    point = anchors.loc[
        anchors["group_id"].eq(APPENDICITIS_ID)
        & np.isclose(anchors["starting_probability"], 0.20)
    ].iloc[0]
    rows: list[dict[str, object]] = []

    def text_row(section: str, element: str, value_text: str, status: str) -> None:
        rows.append(
            {
                "analysis_version": VERSION,
                "profile_section": section,
                "element": element,
                "value": math.nan,
                "low": math.nan,
                "high": math.nan,
                "unit": "text",
                "uncertainty_type": "not applicable",
                "estimator": "not applicable",
                "starting_probability": math.nan,
                "value_text": value_text,
                "availability_status": status,
            }
        )

    def numeric_row(
        section: str,
        element: str,
        metric: str,
        unit: str,
        uncertainty: str,
        starting_probability: float | None = None,
    ) -> None:
        prefix = "pooled_mean" if uncertainty == "model-based 95% pooled-mean interval" else "predictive"
        rows.append(
            {
                "analysis_version": VERSION,
                "profile_section": section,
                "element": element,
                "value": float(point[metric]),
                "low": float(point[f"{prefix}_{metric}_low"]),
                "high": float(point[f"{prefix}_{metric}_high"]),
                "unit": unit,
                "uncertainty_type": uncertainty,
                "estimator": "continuity-corrected bivariate REML",
                "starting_probability": starting_probability,
                "value_text": "",
                "availability_status": "available",
            }
        )

    text_row(
        "Definition",
        "Index test context from review title",
        "Non-contrast CT",
        "review title identifies the index-test context",
    )
    text_row(
        "Definition",
        "Locked operating-point test_name",
        str(point["test_name"]),
        "recorded locked test_name; not a threshold label",
    )
    text_row(
        "Definition",
        "Target condition and setting",
        "Acute appendicitis in the emergency department",
        "recorded in source review title",
    )
    text_row(
        "Definition",
        "Reference standard",
        "",
        "not consistently encoded in the locked Atlas extract",
    )
    text_row(
        "Definition",
        "Index-test positivity or operating threshold",
        "",
        "no separate index-test positivity or operating-threshold field in the Atlas extract",
    )
    rows.append(
        {
            "analysis_version": VERSION,
            "profile_section": "Evidence",
            "element": "Study count",
            "value": int(point["study_rows"]),
            "low": math.nan,
            "high": math.nan,
            "unit": "studies",
            "uncertainty_type": "not applicable",
            "estimator": "not applicable",
            "starting_probability": math.nan,
            "value_text": "",
            "availability_status": "available",
        }
    )
    for metric, label, unit in (
        ("sensitivity", "Sensitivity", "probability"),
        ("specificity", "Specificity", "probability"),
        ("lr_positive", "Positive likelihood ratio", "ratio"),
        ("lr_negative", "Negative likelihood ratio", "ratio"),
    ):
        numeric_row(
            "Operating point",
            label,
            metric,
            unit,
            "model-based 95% pooled-mean interval",
        )
        numeric_row(
            "Operating point",
            f"{label} predictive",
            metric,
            unit,
            "model-based 95% predictive range",
        )
    rows.append(
        {
            "analysis_version": VERSION,
            "profile_section": "Starting probability",
            "element": "Illustrative starting probability",
            "value": 0.20,
            "low": math.nan,
            "high": math.nan,
            "unit": "probability",
            "uncertainty_type": "illustrative anchor; no interval",
            "estimator": "not applicable",
            "starting_probability": 0.20,
            "value_text": "not a disease-specific prevalence estimate",
            "availability_status": "illustrative",
        }
    )
    for metric, label, unit, section in (
        ("information_bits", "Expected information", "bits", "Expected information"),
        (
            "fraction_entropy_removed",
            "Fraction of starting entropy removed",
            "fraction",
            "Expected information",
        ),
        (
            "positive_result_probability",
            "Probability of a positive result",
            "probability",
            "Result branches",
        ),
        (
            "post_positive_probability",
            "Probability after a positive result",
            "probability",
            "Result branches",
        ),
        (
            "negative_result_probability",
            "Probability of a negative result",
            "probability",
            "Result branches",
        ),
        (
            "post_negative_probability",
            "Probability after a negative result",
            "probability",
            "Result branches",
        ),
    ):
        numeric_row(
            section,
            label,
            metric,
            unit,
            "model-based 95% pooled-mean interval",
            0.20,
        )
        numeric_row(
            section,
            f"{label} predictive",
            metric,
            unit,
            "model-based 95% predictive range",
            0.20,
        )
    text_row(
        "Action",
        "Clinical action",
        "",
        "not determined: no consequence model, action threshold, or patient-preference inputs were supplied",
    )
    result = pd.DataFrame(rows)
    result["source"] = point["source"]
    result["review_id"] = point["review_id"]
    result["group_id"] = point["group_id"]
    result["test_name"] = point["test_name"]
    return result


def estimator_prior_numerical_sensitivity(
    root: Path, canonical: pd.DataFrame, anchors: pd.DataFrame
) -> tuple[pd.DataFrame, pd.DataFrame]:
    canonical_metadata = canonical[
        PRIMARY_KEY
        + [
            "review_title",
            "test_name",
            "study_rows",
            "zero_cell_study_fraction",
        ]
    ].copy()
    rows: list[pd.DataFrame] = []
    for specification in _point_specifications(root, canonical):
        frame = specification["frame"].copy()
        if specification["model_specification"] == "v0.42.0 canonical REML":
            for prior in STANDARD_ANCHORS:
                subset = anchors.loc[
                    np.isclose(anchors["starting_probability"], prior)
                ].copy()
                output = subset[
                    PRIMARY_KEY
                    + [
                        "review_title",
                        "test_name",
                        "study_rows",
                        "zero_cell_study_fraction",
                        "sensitivity",
                        "specificity",
                        "pooled_mean_sensitivity_low",
                        "pooled_mean_sensitivity_median",
                        "pooled_mean_sensitivity_high",
                        "pooled_mean_specificity_low",
                        "pooled_mean_specificity_median",
                        "pooled_mean_specificity_high",
                        "information_bits",
                        "pooled_mean_information_bits_low",
                        "pooled_mean_information_bits_median",
                        "pooled_mean_information_bits_high",
                        "fraction_entropy_removed",
                        "pooled_mean_fraction_entropy_removed_low",
                        "pooled_mean_fraction_entropy_removed_median",
                        "pooled_mean_fraction_entropy_removed_high",
                        "post_negative_probability",
                        "pooled_mean_post_negative_probability_low",
                        "pooled_mean_post_negative_probability_median",
                        "pooled_mean_post_negative_probability_high",
                    ]
                ].copy()
                output["starting_probability"] = prior
                output = output.rename(
                    columns={
                        "pooled_mean_sensitivity_low": "sensitivity_low",
                        "pooled_mean_sensitivity_median": "sensitivity_interval_median",
                        "pooled_mean_sensitivity_high": "sensitivity_high",
                        "pooled_mean_specificity_low": "specificity_low",
                        "pooled_mean_specificity_median": "specificity_interval_median",
                        "pooled_mean_specificity_high": "specificity_high",
                        "pooled_mean_information_bits_low": "information_bits_low",
                        "pooled_mean_information_bits_median": "information_bits_interval_median",
                        "pooled_mean_information_bits_high": "information_bits_high",
                        "pooled_mean_fraction_entropy_removed_low": "fraction_entropy_removed_low",
                        "pooled_mean_fraction_entropy_removed_median": "fraction_entropy_removed_interval_median",
                        "pooled_mean_fraction_entropy_removed_high": "fraction_entropy_removed_high",
                        "pooled_mean_post_negative_probability_low": "post_negative_probability_low",
                        "pooled_mean_post_negative_probability_median": "post_negative_probability_interval_median",
                        "pooled_mean_post_negative_probability_high": "post_negative_probability_high",
                    }
                )
                if np.isclose(prior, 0.20):
                    locked_twenty = canonical.set_index("group_id")
                    output["information_bits"] = output["group_id"].map(
                        locked_twenty["information_bits_at_20_percent"]
                    )
                    output["fraction_entropy_removed"] = output["group_id"].map(
                        locked_twenty["diagnostic_entropy_reduction_at_20_percent"]
                    )
                    output["post_negative_probability"] = output["group_id"].map(
                        locked_twenty["negative_posterior_at_20_percent"]
                    )
                output["uncertainty_type"] = "model-based 95% pooled-mean interval"
                rows.append(output)
            continue

        is_regularized = specification["sensitivity_column"].startswith("pooled_")
        frame = frame.merge(
            canonical_metadata,
            on=PRIMARY_KEY,
            how="inner",
            suffixes=("_specification", "_canonical"),
            validate="one_to_one",
        )
        for prior in STANDARD_ANCHORS:
            label = int(round(100 * prior))
            output = frame[PRIMARY_KEY].copy()
            for metadata_column in (
                "review_title",
                "test_name",
                "study_rows",
                "zero_cell_study_fraction",
            ):
                canonical_name = f"{metadata_column}_canonical"
                output[metadata_column] = (
                    frame[canonical_name]
                    if canonical_name in frame
                    else frame[metadata_column]
                )
            sensitivity = frame[specification["sensitivity_column"]].astype(float)
            specificity = frame[specification["specificity_column"]].astype(float)
            output["sensitivity"] = sensitivity
            output["specificity"] = specificity
            output["starting_probability"] = prior
            if is_regularized:
                output["information_bits"] = frame[
                    f"information_bits_at_{label}_percent_median"
                ]
                output["fraction_entropy_removed"] = frame[
                    f"relative_information_at_{label}_percent_median"
                ]
                output["post_negative_probability"] = frame[
                    f"post_negative_at_{label}_percent_median"
                ]
                interval_columns = {
                    "sensitivity_low": "pooled_sensitivity_low",
                    "sensitivity_interval_median": "pooled_sensitivity_median",
                    "sensitivity_high": "pooled_sensitivity_high",
                    "specificity_low": "pooled_specificity_low",
                    "specificity_interval_median": "pooled_specificity_median",
                    "specificity_high": "pooled_specificity_high",
                    "information_bits_low": f"information_bits_at_{label}_percent_low",
                    "information_bits_interval_median": f"information_bits_at_{label}_percent_median",
                    "information_bits_high": f"information_bits_at_{label}_percent_high",
                    "fraction_entropy_removed_low": f"relative_information_at_{label}_percent_low",
                    "fraction_entropy_removed_interval_median": f"relative_information_at_{label}_percent_median",
                    "fraction_entropy_removed_high": f"relative_information_at_{label}_percent_high",
                    "post_negative_probability_low": f"post_negative_at_{label}_percent_low",
                    "post_negative_probability_interval_median": f"post_negative_at_{label}_percent_median",
                    "post_negative_probability_high": f"post_negative_at_{label}_percent_high",
                }
                for target, source_column in interval_columns.items():
                    output[target] = frame[source_column]
                output["uncertainty_type"] = (
                    "descriptive local-Gaussian sensitivity range; not canonical uncertainty"
                )
                output["central_quantity_estimand"] = (
                    "locked released propagated median for each reported quantity"
                )
            else:
                metrics = expanded_diagnostic_metrics(sensitivity, specificity, prior)
                output["information_bits"] = metrics["information_bits"]
                output["fraction_entropy_removed"] = metrics[
                    "fraction_entropy_removed"
                ]
                output["post_negative_probability"] = metrics[
                    "post_negative_probability"
                ]
                for target in (
                    "sensitivity_low",
                    "sensitivity_interval_median",
                    "sensitivity_high",
                    "specificity_low",
                    "specificity_interval_median",
                    "specificity_high",
                    "information_bits_low",
                    "information_bits_interval_median",
                    "information_bits_high",
                    "fraction_entropy_removed_low",
                    "fraction_entropy_removed_interval_median",
                    "fraction_entropy_removed_high",
                    "post_negative_probability_low",
                    "post_negative_probability_interval_median",
                    "post_negative_probability_high",
                ):
                    output[target] = math.nan
                output["uncertainty_type"] = (
                    "historical central estimate only in this sensitivity table"
                )
                output["central_quantity_estimand"] = (
                    "plug-in historical REML central operating point; no attached interval"
                )
            rows.append(output)

        specification_frame = rows[-1]
        del specification_frame

    combined_parts: list[pd.DataFrame] = []
    index = 0
    for specification in _point_specifications(root, canonical):
        for _prior in STANDARD_ANCHORS:
            frame = rows[index].copy()
            frame["analysis_version"] = VERSION
            frame["model_specification"] = specification["model_specification"]
            frame["analysis_role"] = specification["analysis_role"]
            frame["sensitivity_dimension"] = specification["sensitivity_dimension"]
            frame["central_estimator_mixed_with_canonical_interval"] = False
            if specification["analysis_role"] == "canonical primary":
                frame["central_quantity_estimand"] = (
                    "locked Phase 1 released central quantity with separately labeled pooled-mean interval"
                    if np.isclose(_prior, 0.20)
                    else "canonical plug-in REML mean operating point with separately labeled pooled-mean interval"
                )
            combined_parts.append(frame)
            index += 1
    combined = pd.concat(combined_parts, ignore_index=True)
    if len(combined) != 273 * len(STANDARD_ANCHORS) * 5:
        raise AssertionError("Estimator/prior/numerical sensitivity table is incomplete")
    regularized_rows = combined["model_specification"].str.startswith("regularized") | combined[
        "model_specification"
    ].eq("v0.40.0/v0.41.0 regularized GH15")
    direct_median_pairs = (
        ("sensitivity", "sensitivity_interval_median"),
        ("specificity", "specificity_interval_median"),
        ("information_bits", "information_bits_interval_median"),
        ("fraction_entropy_removed", "fraction_entropy_removed_interval_median"),
        ("post_negative_probability", "post_negative_probability_interval_median"),
    )
    combined["derived_central_matches_attached_interval_median"] = pd.NA
    direct_matches = np.logical_and.reduce(
        [
            np.isclose(
                combined.loc[regularized_rows, central],
                combined.loc[regularized_rows, median],
                atol=0,
                rtol=0,
            )
            for central, median in direct_median_pairs
        ]
    )
    combined.loc[
        regularized_rows, "derived_central_matches_attached_interval_median"
    ] = direct_matches
    if not bool(np.all(direct_matches)):
        raise AssertionError(
            "A direct-binomial sensitivity row mixes a propagated median with a plug-in central quantity"
        )

    canonical_values = combined.loc[
        combined["analysis_role"].eq("canonical primary"),
        PRIMARY_KEY
        + [
            "starting_probability",
            "sensitivity",
            "specificity",
            "information_bits",
            "fraction_entropy_removed",
            "post_negative_probability",
        ],
    ].rename(
        columns={
            metric: f"canonical_{metric}"
            for metric in (
                "sensitivity",
                "specificity",
                "information_bits",
                "fraction_entropy_removed",
                "post_negative_probability",
            )
        }
    )
    comparison = combined.merge(
        canonical_values,
        on=PRIMARY_KEY + ["starting_probability"],
        how="left",
        validate="many_to_one",
    )
    summary_records: list[dict[str, object]] = []
    for (model, role, dimension, prior), group in comparison.groupby(
        [
            "model_specification",
            "analysis_role",
            "sensitivity_dimension",
            "starting_probability",
        ],
        sort=True,
    ):
        record: dict[str, object] = {
            "analysis_version": VERSION,
            "model_specification": model,
            "analysis_role": role,
            "sensitivity_dimension": dimension,
            "starting_probability": prior,
            "operating_points": len(group),
        }
        for metric in (
            "sensitivity",
            "specificity",
            "information_bits",
            "fraction_entropy_removed",
            "post_negative_probability",
        ):
            difference = (group[metric] - group[f"canonical_{metric}"]).abs()
            record[f"median_absolute_{metric}_difference"] = float(difference.median())
            record[f"maximum_absolute_{metric}_difference"] = float(difference.max())
        record["spearman_zero_cell_fraction_vs_absolute_information_difference"] = float(
            pd.DataFrame(
                {
                    "zero_cell": group["zero_cell_study_fraction"],
                    "difference": (
                        group["information_bits"]
                        - group["canonical_information_bits"]
                    ).abs(),
                }
            )
            .corr(method="spearman")
            .iloc[0, 1]
        )
        summary_records.append(record)
    summary = pd.DataFrame(summary_records)
    summary["summary_value_source"] = (
        "calculated from the reconciled Phase 3 estimator sensitivity rows"
    )
    phase1_sensitivity = json.loads(
        (root / "analysis/bmj_rmr_v042/estimator_sensitivity_summary.json").read_text(
            encoding="utf-8"
        )
    )
    phase1_locked_row = summary["model_specification"].eq(
        "v0.40.0/v0.41.0 regularized GH15"
    ) & np.isclose(summary["starting_probability"], 0.20)
    summary.loc[
        phase1_locked_row, "median_absolute_information_bits_difference"
    ] = phase1_sensitivity[
        "median_absolute_information_bits_at_20_percent_difference"
    ]
    summary.loc[
        phase1_locked_row, "maximum_absolute_information_bits_difference"
    ] = phase1_sensitivity[
        "maximum_absolute_information_bits_at_20_percent_difference"
    ]
    summary.loc[phase1_locked_row, "summary_value_source"] = (
        "locked Phase 1 estimator_sensitivity_summary.json; avoids sub-ULP CSV reparse drift"
    )
    return combined.sort_values(
        ["model_specification"] + PRIMARY_KEY + ["starting_probability"]
    ).reset_index(drop=True), summary


def exploratory_within_review_contrasts(
    root: Path, anchors: pd.DataFrame
) -> tuple[pd.DataFrame, dict[str, object]]:
    directness = pd.read_csv(root / "analysis/full_atlas/v10/pair_directness_audit.csv")
    old_metrics = pd.read_csv(root / "analysis/full_atlas/v10/pair_model_metrics.csv")
    metadata = (
        old_metrics.sort_values(["pair_id", "prior_probability"])
        .drop_duplicates("pair_id")
        [
            [
                "pair_id",
                "review_id",
                "review_title",
                "left_group_id",
                "left_test",
                "right_group_id",
                "right_test",
                "comparison_type",
                "clinical_comparability_note",
                "baseline_tradeoff_set",
            ]
        ]
    )
    metadata = metadata.merge(
        directness[
            [
                "pair_id",
                "overlapping_study_identifier_count",
                "overlapping_study_identifiers",
                "directness_classification",
                "same_participants_established",
                "cross_test_cells_available",
                "cross_test_covariance_available",
                "interpretation",
            ]
        ],
        on="pair_id",
        how="left",
        validate="one_to_one",
    )
    lookup = anchors.set_index(["group_id", "starting_probability"])
    records: list[dict[str, object]] = []
    for pair in metadata.itertuples(index=False):
        for prior in STANDARD_ANCHORS:
            left = lookup.loc[(pair.left_group_id, prior)]
            right = lookup.loc[(pair.right_group_id, prior)]
            information_difference = float(left["information_bits"] - right["information_bits"])
            relative_difference = float(
                100
                * (
                    left["fraction_entropy_removed"]
                    - right["fraction_entropy_removed"]
                )
            )
            youden_difference = float(left["youden_j"] - right["youden_j"])
            post_negative_difference = float(
                100
                * (
                    left["post_negative_probability"]
                    - right["post_negative_probability"]
                )
            )
            records.append(
                {
                    "analysis_version": VERSION,
                    "pair_id": pair.pair_id,
                    "review_id": pair.review_id,
                    "review_title": pair.review_title,
                    "left_group_id": pair.left_group_id,
                    "left_test": pair.left_test,
                    "right_group_id": pair.right_group_id,
                    "right_test": pair.right_test,
                    "comparison_type": int(pair.comparison_type),
                    "clinical_comparability_note": pair.clinical_comparability_note,
                    "baseline_tradeoff_set": bool(pair.baseline_tradeoff_set),
                    "starting_probability": prior,
                    "directness_classification": pair.directness_classification,
                    "overlapping_study_identifier_count": int(
                        pair.overlapping_study_identifier_count
                    ),
                    "overlapping_study_identifiers": pair.overlapping_study_identifiers,
                    "same_participants_established": bool(
                        pair.same_participants_established
                    ),
                    "cross_test_cells_available": bool(pair.cross_test_cells_available),
                    "cross_test_covariance_available": bool(
                        pair.cross_test_covariance_available
                    ),
                    "paired_inference_supported": False,
                    "left_information_bits": float(left["information_bits"]),
                    "right_information_bits": float(right["information_bits"]),
                    "signed_information_bits_difference": information_difference,
                    "absolute_information_bits_difference": abs(
                        information_difference
                    ),
                    "signed_fraction_entropy_removed_difference_percentage_points": relative_difference,
                    "absolute_fraction_entropy_removed_difference_percentage_points": abs(
                        relative_difference
                    ),
                    "signed_youden_j_difference": youden_difference,
                    "absolute_youden_j_difference": abs(youden_difference),
                    "left_post_negative_probability": float(
                        left["post_negative_probability"]
                    ),
                    "right_post_negative_probability": float(
                        right["post_negative_probability"]
                    ),
                    "signed_post_negative_probability_difference_percentage_points": post_negative_difference,
                    "absolute_post_negative_probability_difference_percentage_points": abs(
                        post_negative_difference
                    ),
                    "youden_information_direction_differs": bool(
                        np.sign(information_difference) != 0
                        and np.sign(youden_difference) != 0
                        and np.sign(information_difference) != np.sign(youden_difference)
                    ),
                    "information_negative_result_direction_differs": bool(
                        np.sign(information_difference) != 0
                        and np.sign(post_negative_difference) != 0
                        and np.sign(information_difference)
                        == np.sign(post_negative_difference)
                    ),
                    "uncertainty_status": "not calculated: same-participant cross-test cells and covariance are unavailable",
                    "inference_status": "exploratory within-review descriptive contrast only; not paired comparative inference",
                    "source_interpretation": pair.interpretation,
                    "estimator": "continuity-corrected bivariate REML central estimates",
                }
            )
    table = pd.DataFrame(records)
    at_twenty = table.loc[np.isclose(table["starting_probability"], 0.20)]
    tradeoffs = at_twenty.loc[at_twenty["baseline_tradeoff_set"]]
    summary = {
        "analysis_version": VERSION,
        "role": "exploratory within-review descriptive contrasts; not paired comparative inference",
        "classified_contrasts": int(table["pair_id"].nunique()),
        "standard_anchor_rows": len(table),
        "tradeoff_contrasts": int(tradeoffs["pair_id"].nunique()),
        "same_participants_established_contrasts": int(
            table.drop_duplicates("pair_id")["same_participants_established"].sum()
        ),
        "cross_test_covariance_available_contrasts": int(
            table.drop_duplicates("pair_id")["cross_test_covariance_available"].sum()
        ),
        "paired_inference_supported_contrasts": int(
            table.drop_duplicates("pair_id")["paired_inference_supported"].sum()
        ),
        "twenty_percent_descriptive_median_absolute_information_difference_bits": float(
            tradeoffs["absolute_information_bits_difference"].median()
        ),
        "twenty_percent_descriptive_median_absolute_post_negative_difference_percentage_points": float(
            tradeoffs[
                "absolute_post_negative_probability_difference_percentage_points"
            ].median()
        ),
        "interpretation": "Continuous magnitudes are retained for the supplement. Sign counts are exploratory and do not support a primary claim.",
    }
    return table, summary


def complete_version_diff(
    root: Path, version_diff: pd.DataFrame, anchors: pd.DataFrame
) -> pd.DataFrame:
    table = version_diff.copy()
    obsolete = [
        column
        for column in table.columns
        if column.endswith("_v042")
        and column
        not in {
            "estimator_v042",
            "inclusion_status_v042",
            "status_change_reason_v042",
        }
    ]
    table = table.drop(columns=obsolete, errors="ignore")
    canonical_wide = anchors.pivot(
        index="operating_point_id",
        columns="starting_probability",
        values=["information_bits", "fraction_entropy_removed"],
    )
    canonical_wide.columns = [
        f"{metric}_{int(round(100 * prior))}_v042"
        for metric, prior in canonical_wide.columns
    ]
    canonical_wide = canonical_wide.reset_index()
    central = anchors.loc[np.isclose(anchors["starting_probability"], 0.20)][
        ["operating_point_id", "sensitivity", "specificity"]
    ].rename(
        columns={"sensitivity": "sensitivity_v042", "specificity": "specificity_v042"}
    )
    canonical_wide = canonical_wide.merge(
        central, on="operating_point_id", validate="one_to_one"
    )
    table = table.merge(canonical_wide, on="operating_point_id", validate="one_to_one")

    curves = pd.read_csv(root / "analysis/full_atlas/operating_point_curves.csv")
    historical_wide = curves.loc[
        curves["pretest_probability"].isin(STANDARD_ANCHORS)
    ].pivot(
        index="group_id",
        columns="pretest_probability",
        values=["information_bits", "diagnostic_entropy_reduction"],
    )
    historical_wide.columns = [
        (
            f"information_bits_{int(round(100 * prior))}_v039"
            if metric == "information_bits"
            else f"fraction_entropy_removed_{int(round(100 * prior))}_v039"
        )
        for metric, prior in historical_wide.columns
    ]
    historical_wide = historical_wide.reset_index()
    historical_derived_columns = [
        f"{metric}_{prior}_v039"
        for metric in ("information_bits", "fraction_entropy_removed")
        for prior in (5, 20, 50)
    ]
    table = table.drop(columns=historical_derived_columns, errors="ignore").merge(
        historical_wide, on="group_id", validate="one_to_one"
    )
    table = table.rename(
        columns={
            "information_bits_5_v040_v041": "information_bits_5_v040_v041",
            "information_bits_20_v040_v041": "information_bits_20_v040_v041",
            "information_bits_50_v040_v041": "information_bits_50_v040_v041",
        }
    )
    table["target_label_status"] = (
        "no separate target-label field in locked normalized data"
    )
    table["threshold_status"] = "no separate threshold field in locked metadata"
    table["zero_cell_burden_definition"] = (
        "fraction of canonical study rows with any zero among TP FP FN TN"
    )
    table["value_change_reason_v039"] = (
        "historical continuity-corrected bivariate REML baseline release"
    )
    table["value_change_reason_v040"] = (
        "regularized direct-binomial GH15 refit replaced historical REML central values"
    )
    table["value_change_reason_v041"] = (
        "display derivation from v0.40.0; no refit and no operating-point value change"
    )
    exception_group = pd.read_csv(
        root / "analysis/bmj_rmr_v042/analysis_row_selection_exceptions.csv"
    ).iloc[0]["group_id"]
    table["value_change_reason_v042"] = np.where(
        table["group_id"].eq(exception_group),
        "canonical REML regenerated after manifest-sheet correction from 45 raw joined rows to 22 declared rows",
        "canonical REML regenerated from the same rows as historical REML; v0.41.0 direct-binomial display values replaced",
    )
    table["transition_reason_v039_to_v040"] = (
        "fixed sample retained; estimator changed from continuity-corrected REML to regularized direct-binomial GH15"
    )
    table["transition_reason_v040_to_v041"] = (
        "fixed sample and values retained; v0.41.0 changed display and narrative architecture only"
    )
    table["transition_reason_v041_to_v042"] = np.where(
        table["group_id"].eq(exception_group),
        "fixed operating-point identity retained; canonical hierarchy returned to REML and corrected one declared source sheet",
        "fixed sample retained; canonical hierarchy returned to regenerated REML while validation and manuscript architecture remain new",
    )
    table["primary_status_v039"] = "primary reference sample"
    table["primary_status_v040"] = "primary reference sample"
    table["primary_status_v041"] = "primary reference sample"
    table["primary_status_v042"] = "primary reference sample"
    table["analysis_status_v042"] = "complete canonical Phase 3 derivation"
    table["status_change_reason_v042"] = table["value_change_reason_v042"]
    required_values = [
        f"{metric}_{prior}_{release}"
        for metric in ("information_bits", "fraction_entropy_removed")
        for prior in (5, 20, 50)
        for release in ("v039", "v040_v041", "v042")
    ]
    if table[required_values].isna().any().any():
        raise AssertionError("Version progression has missing standard-anchor values")
    if len(table) != 273:
        raise AssertionError("Version progression did not preserve 273 operating points")
    return table.sort_values(PRIMARY_KEY).reset_index(drop=True)


def information_identity_checks(anchors: pd.DataFrame) -> dict[str, object]:
    probability_error = (
        anchors["positive_result_probability"]
        + anchors["negative_result_probability"]
        - 1.0
    ).abs()
    entropy_error = (
        anchors["information_bits"]
        - (
            anchors["starting_entropy_bits"]
            - anchors["expected_posterior_entropy_bits"]
        )
    ).abs()
    contribution_error = (
        anchors["information_bits"]
        - anchors["positive_information_contribution_bits"]
        - anchors["negative_information_contribution_bits"]
    ).abs()
    relative_error = (
        anchors["fraction_entropy_removed"]
        - anchors["information_bits"] / anchors["starting_entropy_bits"]
    ).abs()
    return {
        "maximum_result_probability_sum_error": float(probability_error.max()),
        "maximum_entropy_reduction_identity_error_bits": float(entropy_error.max()),
        "maximum_branch_kl_decomposition_error_bits": float(
            contribution_error.max()
        ),
        "maximum_fraction_entropy_removed_identity_error": float(
            relative_error.max()
        ),
        "minimum_information_bits": float(anchors["information_bits"].min()),
        "maximum_information_minus_starting_entropy_bits": float(
            (anchors["information_bits"] - anchors["starting_entropy_bits"]).max()
        ),
        "all_probabilities_in_closed_unit_interval": bool(
            anchors[
                [
                    "positive_result_probability",
                    "negative_result_probability",
                    "post_positive_probability",
                    "post_negative_probability",
                ]
            ]
            .ge(0)
            .all()
            .all()
            and anchors[
                [
                    "positive_result_probability",
                    "negative_result_probability",
                    "post_positive_probability",
                    "post_negative_probability",
                ]
            ]
            .le(1)
            .all()
            .all()
        ),
    }


def update_phase3_claim_ledger(
    root: Path,
    anchor_summary: pd.DataFrame,
    branch_summary: pd.DataFrame,
    theory: dict[str, object],
    cea: pd.DataFrame,
    profile: pd.DataFrame,
    sensitivity_summary: pd.DataFrame,
    contrasts_summary: dict[str, object],
    draws: int,
) -> pd.DataFrame:
    claim_path = root / "docs/manuscript/v19/CLAIM_LEDGER.csv"
    claims = pd.read_csv(claim_path)
    claims = claims.loc[
        ~claims["claim_id"].isin([f"C{number:03d}" for number in range(12, 30)])
    ].copy()
    anchor_by_prior = anchor_summary.set_index("starting_probability")
    cea_lookup = cea.set_index("model_specification")
    profile_lookup = profile.set_index("element")
    direct_sensitivity = sensitivity_summary.loc[
        sensitivity_summary["model_specification"].eq(
            "v0.40.0/v0.41.0 regularized GH15"
        )
        & np.isclose(sensitivity_summary["starting_probability"], 0.20)
    ].iloc[0]
    estimator_effect = json.loads(
        (root / "analysis/bmj_rmr_v042/estimator_sensitivity_summary.json").read_text(
            encoding="utf-8"
        )
    )
    correction_effect = json.loads(
        (
            root
            / "analysis/bmj_rmr_v042/continuity_correction_sensitivity_summary.json"
        ).read_text(encoding="utf-8")
    )
    diagnostics = json.loads(
        (root / "analysis/bmj_rmr_v042/reml_diagnostics_summary.json").read_text(
            encoding="utf-8"
        )
    )
    fidelity_effect = json.loads(
        (root / "analysis/bmj_rmr_v042/source_summary_fidelity_summary.json").read_text(
            encoding="utf-8"
        )
    )

    def profile_quantity(element: str, digits: int) -> str:
        pooled = profile_lookup.loc[element]
        predictive = profile_lookup.loc[f"{element} predictive"]
        return (
            f"{pooled['value']:.{digits}f} "
            f"(pooled-mean 95% interval {pooled['low']:.{digits}f} to {pooled['high']:.{digits}f}; "
            f"predictive 95% range {predictive['low']:.{digits}f} to {predictive['high']:.{digits}f})"
        )

    branch_counts = {
        (float(row.starting_probability), str(row.result_branch), str(row.entropy_movement)): int(
            row.operating_points
        )
        for row in branch_summary.itertuples(index=False)
    }
    new_claims = [
        {
            "claim_id": "C012",
            "claim_text": (
                "Median expected information across 273 primary operating points is "
                f"{anchor_by_prior.loc[0.05, 'information_bits_median']:.3f}, "
                f"{anchor_by_prior.loc[0.20, 'information_bits_median']:.3f}, and "
                f"{anchor_by_prior.loc[0.50, 'information_bits_median']:.3f} bits at 5%, 20%, and 50% starting probabilities"
            ),
            "manuscript_location": "Results and Supplement",
            "claim_role": "primary",
            "source_file": "analysis/bmj_rmr_v042/standard_anchor_summary.csv",
            "source_row_or_variable": "information_bits_median at 0.05 0.20 0.50",
            "estimator": "continuity-corrected bivariate REML",
            "starting_probability": "0.05;0.20;0.50",
            "uncertainty_type": "cross-diagnosis descriptive distribution",
            "source_validation_status": "all 273 points traceable in Phase 2",
            "model_stability_status": "primary Phase 3 derivation",
            "automated_check": "test_phase3_anchor_outputs_are_complete_and_information_identities_hold",
            "final_status": "validated Phase 3; pending prose",
        },
        {
            "claim_id": "C013",
            "claim_text": (
                "Within the fixed 273-point sample, expected information and Youden's J show descriptive empirical rank concordance at the standard anchors "
                f"(Spearman rho {anchor_by_prior.loc[0.05, 'spearman_information_vs_youden_j']:.6f}, "
                f"{anchor_by_prior.loc[0.20, 'spearman_information_vs_youden_j']:.6f}, and "
                f"{anchor_by_prior.loc[0.50, 'spearman_information_vs_youden_j']:.6f} at 5%, 20%, and 50%)"
            ),
            "manuscript_location": "Results; Figure 2; Figure S3",
            "claim_role": "primary",
            "source_file": "analysis/bmj_rmr_v042/standard_anchor_summary.csv",
            "source_row_or_variable": "spearman_information_vs_youden_j at starting_probability 0.05 0.20 0.50",
            "estimator": "continuity-corrected bivariate REML",
            "starting_probability": "0.05;0.20;0.50",
            "uncertainty_type": "cross-diagnosis rank concordance",
            "source_validation_status": "all 273 points traceable in Phase 2",
            "model_stability_status": "fixed-sample empirical description; shared sensitivity and specificity inputs do not guarantee concordance",
            "automated_check": "test_phase3_claim_ledger_separates_empirical_concordance_from_theory",
            "final_status": "validated Phase 3; pending prose",
        },
        {
            "claim_id": "C014",
            "claim_text": (
                "Mathematical equivalence does not follow from empirical concordance: the specified exact equal-Youden example has identical J but expected-information values differing by "
                f"{theory['equal_youden_different_information']['absolute_information_difference_bits']:.6f} bits at 20%"
            ),
            "manuscript_location": "Results; Figure 3",
            "claim_role": "primary",
            "source_file": "analysis/bmj_rmr_v042/theoretical_non_equivalence_summary.json",
            "source_row_or_variable": "equal_youden_different_information.absolute_youden_difference and absolute_information_difference_bits",
            "estimator": "theoretical calculation",
            "starting_probability": "0.20",
            "uncertainty_type": "not applicable",
            "source_validation_status": "not applicable; theoretical",
            "model_stability_status": "exact equality and information difference numerically verified",
            "automated_check": "test_phase3_theoretical_non_equivalence_examples_are_recalculated",
            "final_status": "validated Phase 3; pending prose",
        },
        {
            "claim_id": "C015",
            "claim_text": (
                "At 20%, the specified near-equal-information example differs by "
                f"{theory['near_equal_information_different_post_negative']['absolute_information_difference_bits']:.6f} bits but by "
                f"{100 * theory['near_equal_information_different_post_negative']['absolute_post_negative_difference']:.1f} percentage points after a negative result"
            ),
            "manuscript_location": "Results; Figure 3",
            "claim_role": "primary",
            "source_file": "analysis/bmj_rmr_v042/theoretical_non_equivalence_summary.json",
            "source_row_or_variable": "near_equal_information_different_post_negative",
            "estimator": "theoretical calculation",
            "starting_probability": "0.20",
            "uncertainty_type": "not applicable",
            "source_validation_status": "not applicable; theoretical",
            "model_stability_status": "specified tolerance independently verified",
            "automated_check": "test_phase3_theoretical_non_equivalence_examples_are_recalculated",
            "final_status": "validated Phase 3; pending prose",
        },
        {
            "claim_id": "C016",
            "claim_text": (
                "In the observed primary branch profiles, positive results increase binary entropy for "
                f"{branch_counts[(0.05, 'positive_branch_entropy_movement', 'increase')]}/273 points at 5% and "
                f"{branch_counts[(0.20, 'positive_branch_entropy_movement', 'increase')]}/273 at 20%, but 0/273 at 50%; "
                "negative results increase entropy for 0/273 points at every standard anchor. Expected mutual information remains non-negative"
            ),
            "manuscript_location": "Results; Figure 4",
            "claim_role": "primary",
            "source_file": "analysis/bmj_rmr_v042/branch_entropy_summary.csv",
            "source_row_or_variable": "all result_branch x entropy_movement counts at starting_probability 0.05 0.20 0.50",
            "estimator": "continuity-corrected bivariate REML",
            "starting_probability": "0.05;0.20;0.50",
            "uncertainty_type": "observed primary central-estimate branch counts",
            "source_validation_status": "all 273 points traceable in Phase 2",
            "model_stability_status": "entropy identities numerically verified",
            "automated_check": "test_phase3_branch_claim_is_narrowed_to_observed_canonical_pattern",
            "final_status": "validated Phase 3; pending prose",
        },
        {
            "claim_id": "C017",
            "claim_text": (
                "The empirical CEA information crossing is "
                f"{100 * cea_lookup.loc['v0.42.0 canonical REML', 'crossing_probability']:.1f}% under primary REML and "
                f"{100 * cea_lookup.loc['v0.40.0/v0.41.0 regularized GH15', 'crossing_probability']:.1f}% under the historical headline sensitivity model"
            ),
            "manuscript_location": "Supplement",
            "claim_role": "supporting",
            "source_file": "analysis/bmj_rmr_v042/empirical_cea_crossing_sensitivity.csv",
            "source_row_or_variable": "primary REML and regularized GH15 crossing_probability",
            "estimator": "primary REML with model-specific sensitivities",
            "starting_probability": "numerical roots",
            "uncertainty_type": "model-specific central-estimate sensitivity",
            "source_validation_status": "empirical source points traceable",
            "model_stability_status": "model dependence explicit",
            "automated_check": "test_phase3_empirical_cea_crossing_is_canonical_with_history_explicit",
            "final_status": "validated Phase 3; pending prose",
        },
        {
            "claim_id": "C018",
            "claim_text": (
                "For the 11-study appendicitis profile, whose review title identifies non-contrast CT while the recorded test_name is 'Overall structured 2 x 2 data', primary REML gives "
                f"sensitivity {profile_quantity('Sensitivity', 4)}, specificity {profile_quantity('Specificity', 4)}, and expected information "
                f"{profile_quantity('Expected information', 4)} bits at the illustrative 20% starting probability"
            ),
            "manuscript_location": "Results; Figure 1; Table 2",
            "claim_role": "primary",
            "source_file": "analysis/bmj_rmr_v042/appendicitis_worked_profile.csv",
            "source_row_or_variable": "Sensitivity; Sensitivity predictive; Specificity; Specificity predictive; Expected information; Expected information predictive; Index test context from review title; Locked operating-point test_name",
            "estimator": "continuity-corrected bivariate REML",
            "starting_probability": "0.20 illustrative",
            "uncertainty_type": "model-based pooled-mean interval and predictive range",
            "source_validation_status": "operating point traceable in Phase 2",
            "model_stability_status": "primary values and intervals regenerated",
            "automated_check": "test_phase3_appendicitis_claims_have_exact_values_intervals_and_provenance",
            "final_status": "validated Phase 3; pending prose",
        },
        {
            "claim_id": "C019",
            "claim_text": (
                "At 20%, the like-for-like plug-in central-estimate comparison between primary REML and regularized GH15 has an expected-information difference with median "
                f"{estimator_effect['median_absolute_like_for_like_plugin_information_bits_at_20_percent_difference']!r} bits and maximum "
                f"{estimator_effect['maximum_absolute_like_for_like_plugin_information_bits_at_20_percent_difference']!r} bits. The analysis-family propagated-central movement is "
                f"{float(direct_sensitivity['median_absolute_information_bits_difference'])!r} and "
                f"{float(direct_sensitivity['maximum_absolute_information_bits_difference'])!r} bits and is a comparison of different central-summary estimands"
            ),
            "manuscript_location": "Results and Supplement",
            "claim_role": "supporting",
            "source_file": "analysis/bmj_rmr_v042/estimator_sensitivity_summary.json",
            "source_row_or_variable": "like-for-like plug-in and analysis-family information-difference summaries",
            "estimator": "REML versus regularized direct-binomial GH15",
            "starting_probability": "0.20",
            "uncertainty_type": "like-for-like plug-in central-estimate comparison; analysis-family movement separately labelled as different estimands",
            "source_validation_status": "fixed model inputs",
            "model_stability_status": "model dependence quantified",
            "automated_check": "test_phase6_estimator_effect_and_release_movement_are_distinct",
            "final_status": "validated Phase 3; pending prose",
        },
        {
            "claim_id": "C020",
            "claim_text": (
                f"The {contrasts_summary['classified_contrasts']} classified within-review contrasts are exploratory descriptions; none has established same-participant data or cross-test covariance for paired inference"
            ),
            "manuscript_location": "Supplement",
            "claim_role": "exploratory",
            "source_file": "analysis/bmj_rmr_v042/exploratory_within_review_contrasts.csv",
            "source_row_or_variable": "all pair_id rows and inference_status",
            "estimator": "continuity-corrected bivariate REML central estimates",
            "starting_probability": "0.05;0.20;0.50",
            "uncertainty_type": "none; covariance unavailable",
            "source_validation_status": "locked author classification",
            "model_stability_status": "descriptive only",
            "automated_check": "test_phase3_exploratory_contrasts_never_claim_paired_inference",
            "final_status": "validated Phase 3; pending prose",
        },
        {
            "claim_id": "C021",
            "claim_text": "Clinical action remains undetermined in the appendicitis profile because no consequence model, action threshold, or patient-preference inputs were supplied",
            "manuscript_location": "Figure 1; Table 2",
            "claim_role": "primary",
            "source_file": "analysis/bmj_rmr_v042/appendicitis_worked_profile.csv",
            "source_row_or_variable": "Clinical action availability_status",
            "estimator": "not applicable",
            "starting_probability": "not applicable",
            "uncertainty_type": "not applicable",
            "source_validation_status": "missing inputs preserved as missing",
            "model_stability_status": "not applicable",
            "automated_check": "test_phase3_appendicitis_profile_has_required_values_intervals_and_open_action",
            "final_status": "validated Phase 3; pending prose",
        },
        {
            "claim_id": "C022",
            "claim_text": (
                "The theoretical probability-dependent ordering example crosses at starting probability "
                f"{theory['probability_dependent_information_crossing']['numerical_root']:.12f}; this numerical root is a display quantity, not an action threshold"
            ),
            "manuscript_location": "Results; Figure 3; Figure S8",
            "claim_role": "primary",
            "source_file": "analysis/bmj_rmr_v042/theoretical_non_equivalence_summary.json",
            "source_row_or_variable": "probability_dependent_information_crossing.numerical_root root_residual_bits display_probability_is_action_threshold",
            "estimator": "theoretical calculation",
            "starting_probability": str(theory["probability_dependent_information_crossing"]["numerical_root"]),
            "uncertainty_type": "numerical root",
            "source_validation_status": "not applicable; theoretical",
            "model_stability_status": "root residual tested",
            "automated_check": "test_phase3_theoretical_non_equivalence_examples_are_recalculated",
            "final_status": "validated Phase 3; pending prose",
        },
        {
            "claim_id": "C023",
            "claim_text": (
                "In the appendicitis worked profile, LR+ is "
                f"{profile_quantity('Positive likelihood ratio', 4)} and LR- is "
                f"{profile_quantity('Negative likelihood ratio', 4)}"
            ),
            "manuscript_location": "Figure 1; Table 2",
            "claim_role": "primary",
            "source_file": "analysis/bmj_rmr_v042/appendicitis_worked_profile.csv",
            "source_row_or_variable": "Positive likelihood ratio; Positive likelihood ratio predictive; Negative likelihood ratio; Negative likelihood ratio predictive",
            "estimator": "continuity-corrected bivariate REML",
            "starting_probability": "not applicable",
            "uncertainty_type": "model-based pooled-mean interval and predictive range",
            "source_validation_status": "operating point traceable in Phase 2",
            "model_stability_status": "canonical values and intervals regenerated",
            "automated_check": "test_phase3_appendicitis_claims_have_exact_values_intervals_and_provenance",
            "final_status": "validated Phase 3; pending prose",
        },
        {
            "claim_id": "C024",
            "claim_text": (
                "At the illustrative 20% anchor, the appendicitis profile gives probability of a positive result "
                f"{profile_quantity('Probability of a positive result', 4)}, probability after a positive result "
                f"{profile_quantity('Probability after a positive result', 4)}, probability of a negative result "
                f"{profile_quantity('Probability of a negative result', 4)}, and probability after a negative result "
                f"{profile_quantity('Probability after a negative result', 4)}"
            ),
            "manuscript_location": "Figure 1; Table 2",
            "claim_role": "primary",
            "source_file": "analysis/bmj_rmr_v042/appendicitis_worked_profile.csv",
            "source_row_or_variable": "Probability of a positive result; Probability of a positive result predictive; Probability after a positive result; Probability after a positive result predictive; Probability of a negative result; Probability of a negative result predictive; Probability after a negative result; Probability after a negative result predictive",
            "estimator": "continuity-corrected bivariate REML",
            "starting_probability": "0.20 illustrative",
            "uncertainty_type": "model-based pooled-mean interval and predictive range",
            "source_validation_status": "operating point traceable in Phase 2",
            "model_stability_status": "canonical values and intervals regenerated",
            "automated_check": "test_phase3_appendicitis_claims_have_exact_values_intervals_and_provenance",
            "final_status": "validated Phase 3; pending prose",
        },
        {
            "claim_id": "C025",
            "claim_text": "The author-proposed Diagnostic Information Profile is an organising and reporting template for four linked questions: what is the test, how much expected uncertainty it removes, where positive and negative results lead, and what action follows when a consequence model and preferences are available",
            "manuscript_location": "Table 1",
            "claim_role": "primary",
            "source_file": "analysis/bmj_rmr_v042/table_1_reporting_architecture.csv",
            "source_row_or_variable": "all four question rows and five reporting columns",
            "estimator": "reporting architecture; not an estimator",
            "starting_probability": "test-dependent",
            "uncertainty_type": "required uncertainty or context column",
            "source_validation_status": "specified in the durable v0.42.0 forward-completion specification",
            "model_stability_status": "not applicable",
            "automated_check": "test_phase3_figure_table_map_sources_and_claim_coverage_are_complete",
            "final_status": "validated Phase 3; pending prose",
        },
        {
            "claim_id": "C026",
            "claim_text": (
                "Across 92 fidelity comparisons, median absolute sensitivity and specificity differences were "
                f"{fidelity_effect['median_absolute_sensitivity_difference']:.6f} and {fidelity_effect['median_absolute_specificity_difference']:.6f}, and maxima were "
                f"{fidelity_effect['maximum_absolute_sensitivity_difference']:.6f} and {fidelity_effect['maximum_absolute_specificity_difference']:.6f}. Three large discrepancies were reader-audited: the CD007394 6-versus-7 row mismatch may contribute to one, while the other two cannot be attributed because threshold and pooling metadata are incomplete"
            ),
            "manuscript_location": "Methods; Results; Supplement; Figure S2",
            "claim_role": "supporting",
            "source_file": "analysis/bmj_rmr_v042/source_summary_fidelity_summary.json; analysis/bmj_rmr_v042/source_summary_large_discrepancies.csv",
            "source_row_or_variable": "fidelity median/maximum fields; all three large-discrepancy rows and explanation_from_available_metadata",
            "estimator": "source-reported summary versus primary bivariate REML",
            "starting_probability": "not applicable",
            "uncertainty_type": "fidelity discrepancy; no truth standard",
            "source_validation_status": "all three discrepancies traced to source and primary-analysis records",
            "model_stability_status": "causes remain unassigned where metadata are incomplete",
            "automated_check": "test_phase6_large_fidelity_discrepancies_are_reader_facing",
            "final_status": "Phase 6 corrective candidate pending fresh re-QA",
        },
        {
            "claim_id": "C027",
            "claim_text": (
                "A direct all-273 sensitivity that added 0.5 to all four cells only for studies containing any zero, while leaving zero-free studies uncorrected, changed 20% expected information by median "
                f"{correction_effect['median_absolute_information_bits_at_20_percent_difference']:.6f} bits and maximum "
                f"{correction_effect['maximum_absolute_information_bits_at_20_percent_difference']:.6f} bits; "
                f"{correction_effect['moved_information_bits_at_20_percent_count_above_1e_12']}/273 points moved above 1e-12"
            ),
            "manuscript_location": "Methods; Supplement; Limitations",
            "claim_role": "supporting",
            "source_file": "analysis/bmj_rmr_v042/continuity_correction_sensitivity_summary.json",
            "source_row_or_variable": "all central movement and optimizer diagnostic fields",
            "estimator": "bivariate REML under zero-cell-triggered continuity correction",
            "starting_probability": "0.20",
            "uncertainty_type": "central-estimate sensitivity",
            "source_validation_status": "same fixed 273 points and 4104 primary-analysis rows",
            "model_stability_status": "framework conclusions retained; universal correction retained as the primary historical convention for reproducibility",
            "automated_check": "test_phase6_zero_cell_triggered_continuity_sensitivity",
            "final_status": "Phase 6 corrective candidate pending fresh re-QA",
        },
        {
            "claim_id": "C028",
            "claim_text": (
                f"Among numerically tied REML solutions, {diagnostics['optimizer_tied_solution_log_l00_at_lower_bound']} fits had log-L00 and {diagnostics['optimizer_tied_solution_log_l11_at_lower_bound']} had log-L11 at -8, {diagnostics['optimizer_any_tied_solution_cholesky_diagonal_at_lower_bound']} unique; the stationarity-selected endpoints were at those bounds in {diagnostics['optimizer_selected_endpoint_log_l00_at_lower_bound']} and {diagnostics['optimizer_selected_endpoint_log_l11_at_lower_bound']} fits. Seven tied-solution flags were missed by older flags and the diagnostic union contains {diagnostics['union_of_all_diagnostic_flags']} fits. Appendicitis has both a selected endpoint and tied solution at log-L11 -8, with between-study correlation {diagnostics['appendicitis_between_study_correlation']:.6f}"
            ),
            "manuscript_location": "Methods; Supplement; Figure S4; Limitations",
            "claim_role": "supporting",
            "source_file": "analysis/bmj_rmr_v042/reml_diagnostics_summary.json",
            "source_row_or_variable": "hard-bound, union, and appendicitis diagnostic fields",
            "estimator": "continuity-corrected bivariate REML",
            "starting_probability": "not applicable",
            "uncertainty_type": "optimizer and heterogeneity diagnostics",
            "source_validation_status": "all 273 primary fits",
            "model_stability_status": "central profile retained; joint heterogeneity and predictive ranges conditional and fragile in affected fits",
            "automated_check": "test_phase6_hard_bound_diagnostics_are_reconstructed",
            "final_status": "Phase 6 corrective candidate pending fresh re-QA",
        },
        {
            "claim_id": "C029",
            "claim_text": (
                f"Pooled-mean interval and predictive-range summaries use {draws:,} deterministic draws per operating point from the fitted bivariate mean covariance and the covariance with added fitted heterogeneity, respectively"
            ),
            "manuscript_location": "Methods",
            "claim_role": "supporting",
            "source_file": "analysis/bmj_rmr_v042/phase3_summary.json",
            "source_row_or_variable": "uncertainty_draws_per_operating_point",
            "estimator": "continuity-corrected bivariate REML",
            "starting_probability": "not applicable",
            "uncertainty_type": "deterministic plug-in model distributions for pooled-mean intervals and predictive ranges, conditional on fitted heterogeneity",
            "source_validation_status": "draw count and covariance sources recorded in Phase 3 outputs",
            "model_stability_status": "heterogeneity-parameter uncertainty not propagated; predictive ranges not recovery-checked",
            "automated_check": "test_phase6_claim_governance_for_uncertainty_draws_and_historical_regeneration",
            "final_status": "Phase 6 corrective candidate pending fresh re-QA",
        },
    ]
    claims = pd.concat([claims, pd.DataFrame(new_claims)], ignore_index=True)
    claims.loc[claims["claim_id"].eq("C006"), "claim_text"] = (
        "The prespecified primary estimator is bivariate REML using the historical regularising convention that adds 0.5 to every cell of every study"
    )
    claims.loc[claims["claim_id"].eq("C008"), "claim_text"] = (
        "The 24 single-dataset REML scenarios are a limited recovery/smoke diagnostic: pooled-mean intervals included the generating sensitivity and specificity in 87.5% of scenarios, not a repeated-sampling coverage evaluation"
    )
    claims.loc[claims["claim_id"].eq("C009"), "claim_text"] = (
        f"All 273 selected REML fits converged; numerical ties included {diagnostics['optimizer_any_tied_solution_cholesky_diagonal_at_lower_bound']} fits with a Cholesky-diagonal endpoint at -8, selected endpoints were at a bound in {diagnostics['optimizer_any_selected_endpoint_cholesky_diagonal_at_lower_bound']} fits, and the full diagnostic union contains {diagnostics['union_of_all_diagnostic_flags']} fits"
    )
    claims.loc[claims["claim_id"].eq("C010"), "claim_text"] = (
        "Route-specific summaries describe fixed-sample variation only and do not establish reproducibility, representativeness, or transportability"
    )
    claims["final_status"] = (
        "0.42.0 scientific release passed; 0.42.1 reader/figure patch pending North Star re-audit"
    )
    claims.to_csv(claim_path, index=False, lineterminator="\n")
    return claims


def reporting_architecture_table() -> pd.DataFrame:
    return pd.DataFrame(
        [
            {
                "question": "What exactly is the test?",
                "quantity": "Index test, target condition, setting, index-test positivity or operating threshold, and evidence base",
                "when_known": "Before interpreting derived information",
                "required_uncertainty_or_context": "Preserve missing labels and thresholds as missing; distinguish review-title context from the locked test_name",
                "common_interpretation_error": "Treating a review title, test family, or pooled label as a recorded threshold",
            },
            {
                "question": "How much uncertainty does it remove on average?",
                "quantity": "Expected mutual information and fraction of starting entropy removed",
                "when_known": "After specifying a starting probability and an operating-point model",
                "required_uncertainty_or_context": "Report the starting probability and keep pooled-mean intervals separate from predictive ranges",
                "common_interpretation_error": "Reading expected information as the effect of every observed result",
            },
            {
                "question": "Where do positive and negative results lead?",
                "quantity": "Result probabilities, post-result probabilities, and branch-specific entropy",
                "when_known": "After specifying the starting probability and likelihood ratios",
                "required_uncertainty_or_context": "Report positive and negative branches separately with their propagated uncertainty",
                "common_interpretation_error": "Equating branch entropy movement with clinical benefit or harm",
            },
            {
                "question": "What action follows?",
                "quantity": "Action selected under explicit consequences, action thresholds, costs, alternatives, and patient preferences",
                "when_known": "Only when an applicable consequence model and preference inputs are supplied",
                "required_uncertainty_or_context": "State the action as undetermined when required decision inputs are unavailable",
                "common_interpretation_error": "Treating an information crossing or illustrative starting probability as an action threshold",
            },
        ]
    )


def write_phase3_figure_table_map(root: Path) -> Path:
    map_path = root / "docs/manuscript/v19/FIGURE_TABLE_MAP.md"
    rows = [
        ("Figure 1", "Four-question framework and primary appendicitis profile", "analysis/bmj_rmr_v042/appendicitis_worked_profile.csv; analysis/bmj_rmr_v042/table_1_reporting_architecture.csv", "C018; C021; C023; C024; C025", "Gate 5 passed; Phase 6 corrective candidate pending fresh Gate 6 re-QA"),
        ("Figure 2", "Complete 273-point signature Atlas", "analysis/bmj_rmr_v042/derived_operating_points_standard_anchors.csv; analysis/bmj_rmr_v042/atlas_landmarks.csv", "C001; C012; C013", "Phase 3 sources complete; Phase 4 will compose the display"),
        ("Figure 3", "Three mathematical non-equivalences", "analysis/bmj_rmr_v042/theoretical_non_equivalence_examples.csv; analysis/bmj_rmr_v042/theoretical_crossing_probability_profile.csv; analysis/bmj_rmr_v042/theoretical_non_equivalence_summary.json", "C014; C015; C022", "Phase 3 sources complete; Phase 4 will compose the display"),
        ("Figure 4", "Result-branch uncertainty at 5%, 20%, and 50%", "analysis/bmj_rmr_v042/branch_entropy_all_primary.csv; analysis/bmj_rmr_v042/branch_entropy_summary.csv", "C016", "Phase 3 sources complete; Phase 4 will compose the display"),
        ("Table 1", "Author-proposed four-question reporting template", "analysis/bmj_rmr_v042/table_1_reporting_architecture.csv", "C025", "Phase 3 source complete; Phase 4 will compose the table"),
        ("Table 2", "Worked Diagnostic Information Profile", "analysis/bmj_rmr_v042/appendicitis_worked_profile.csv", "C018; C021; C023; C024", "Phase 3 source complete; Phase 4 will compose the table"),
        ("Figure S1", "Corpus and selection flow", "analysis/bmj_rmr_v042/run_manifest.json; analysis/bmj_rmr_v042/source_trace_summary.json; analysis/bmj_rmr_v042/analysis_row_selection_exceptions.csv", "C001; C002; C011", "Gate 5 passed; Phase 6 corrective candidate pending fresh Gate 6 re-QA"),
        ("Figure S2", "Source fidelity, audit denominators, and fixed-sample route variation", "analysis/bmj_rmr_v042/source_summary_fidelity.csv; analysis/bmj_rmr_v042/manual_audit_summary.json; analysis/bmj_rmr_v042/source_route_stability.csv; analysis/bmj_rmr_v042/source_summary_large_discrepancies.csv", "C003; C004; C010; C026", "Gate 5 passed; Phase 6 corrective candidate pending fresh Gate 6 re-QA"),
        ("Figure S3", "Information distributions and Youden correlations", "analysis/bmj_rmr_v042/derived_operating_points_standard_anchors.csv; analysis/bmj_rmr_v042/standard_anchor_summary.csv", "C012; C013", "Phase 3 sources complete; Phase 4 will compose the display"),
        ("Figure S4", "Limited recovery/smoke diagnostic and optimizer diagnostics", "analysis/bmj_rmr_v042/reml_recovery.csv; analysis/bmj_rmr_v042/reml_recovery_summary.json; analysis/bmj_rmr_v042/reml_diagnostics_all_primary.csv; analysis/bmj_rmr_v042/reml_diagnostics_summary.json", "C006; C008; C009; C028", "Phase 6 corrected sources complete; pending fresh Gate 6 re-QA"),
        ("Figure S5", "Like-for-like plug-in central estimates and analysis-family sensitivity", "analysis/bmj_rmr_v042/estimator_prior_numerical_sensitivity_all_primary.csv; analysis/bmj_rmr_v042/estimator_prior_numerical_sensitivity_summary.csv; analysis/bmj_rmr_v042/estimator_sensitivity_summary.json", "C019", "Phase 6 corrected sources complete; pending fresh Gate 6 re-QA"),
        ("Figure S6", "Exploratory within-review magnitudes and analysis-family movement", "analysis/bmj_rmr_v042/exploratory_within_review_contrasts.csv; analysis/bmj_rmr_v042/exploratory_within_review_contrasts_summary.json; analysis/bmj_rmr_v042/estimator_prior_numerical_sensitivity_all_primary.csv", "C020", "Gate 5 passed; Phase 6 corrective candidate pending fresh Gate 6 re-QA"),
        ("Figure S7", "Near-tie and restricted-composition audits", "analysis/bmj_rmr_v042/exploratory_within_review_contrasts.csv; analysis/bmj_rmr_v042/derived_operating_points_standard_anchors.csv", "C020", "Phase 3 source inputs complete; Phase 4 will compose the audit display"),
        ("Figure S8", "Empirical CEA crossing sensitivity and theoretical checks", "analysis/bmj_rmr_v042/empirical_cea_crossing_sensitivity.csv; analysis/bmj_rmr_v042/empirical_cea_probability_profiles.csv; analysis/bmj_rmr_v042/theoretical_non_equivalence_summary.json", "C014; C015; C017; C022", "Phase 3 sources complete; Phase 4 will compose the display"),
    ]
    rows = [
        (
            display,
            role,
            sources,
            claim_ids,
            "Gate 5 passed; Phase 6 corrective candidate pending fresh Gate 6 re-QA",
        )
        for display, role, sources, claim_ids, _status in rows
    ]
    lines = [
        "# Figure and table map",
        "",
        "Every path below is repository-root relative and identifies an existing machine-readable source. Gate 5 passed for the inherited package; the corrected Phase 6 display package is a candidate pending fresh Gate 6 re-QA.",
        "Method-only claims C007 and C029 govern the supplementary historical-regeneration sentence and the main uncertainty-draw statement, respectively; neither is a display claim.",
        "",
        "| Display | Scientific role | Required source files | Claim IDs | Source status |",
        "|---|---|---|---|---|",
    ]
    for display, role, sources, claim_ids, status in rows:
        linked_sources = "; ".join(f"`{source}`" for source in sources.split("; "))
        lines.append(f"| {display} | {role} | {linked_sources} | {claim_ids} | {status} |")
    map_path.write_text("\n".join(lines) + "\n", encoding="utf-8")
    return map_path


def phase3(
    root: Path, draws: int = DERIVED_DRAWS, reuse_derived: bool = False
) -> dict[str, object]:
    analysis_dir = root / "analysis/bmj_rmr_v042"
    qa_dir = analysis_dir / "qa"
    canonical = pd.read_csv(analysis_dir / "operating_points_primary.csv")
    if len(canonical) != 273 or not canonical["model"].eq(
        "continuity-corrected bivariate REML"
    ).all():
        raise AssertionError("Phase 3 requires the locked 273-point canonical REML output")

    anchor_path = analysis_dir / "derived_operating_points_standard_anchors.csv"
    profile_path = analysis_dir / "derived_operating_points_probability_profiles.csv"
    if reuse_derived:
        anchors = pd.read_csv(anchor_path)
        probability_profiles = pd.read_csv(profile_path)
        if (
            len(anchors) != 819
            or len(probability_profiles) != 13_650
            or not anchors["uncertainty_draws"].eq(draws).all()
            or not probability_profiles["uncertainty_draws"].eq(draws).all()
        ):
            raise AssertionError(
                "--reuse-derived requires complete outputs generated with the requested draw count"
            )
    else:
        anchors, probability_profiles = canonical_derived_outputs(
            canonical, draws=draws
        )
    if not reuse_derived:
        anchors.to_csv(
            anchor_path,
            index=False,
            lineterminator="\n",
        )
        probability_profiles.to_csv(
            profile_path,
            index=False,
            lineterminator="\n",
        )
    # Standardize all downstream calculations on the exact released CSV values so
    # a full regeneration and --reuse-derived finalization are byte-identical.
    anchors = pd.read_csv(anchor_path)
    probability_profiles = pd.read_csv(profile_path)
    anchor_summary, branches, branch_summary = anchor_and_branch_summaries(anchors)
    anchor_summary.to_csv(
        analysis_dir / "standard_anchor_summary.csv", index=False, lineterminator="\n"
    )
    branches.to_csv(
        analysis_dir / "branch_entropy_all_primary.csv", index=False, lineterminator="\n"
    )
    branch_summary.to_csv(
        analysis_dir / "branch_entropy_summary.csv", index=False, lineterminator="\n"
    )

    landmarks = landmark_table(anchors)
    landmarks.to_csv(
        analysis_dir / "atlas_landmarks.csv", index=False, lineterminator="\n"
    )
    theory_table, theory_profile, theory_summary = theoretical_non_equivalence()
    theory_table.to_csv(
        analysis_dir / "theoretical_non_equivalence_examples.csv",
        index=False,
        lineterminator="\n",
    )
    theory_profile.to_csv(
        analysis_dir / "theoretical_crossing_probability_profile.csv",
        index=False,
        lineterminator="\n",
    )
    write_json(
        analysis_dir / "theoretical_non_equivalence_summary.json", theory_summary
    )

    cea_crossings, cea_profiles = empirical_cea_crossing(root, canonical)
    cea_crossings.to_csv(
        analysis_dir / "empirical_cea_crossing_sensitivity.csv",
        index=False,
        lineterminator="\n",
    )
    cea_profiles.to_csv(
        analysis_dir / "empirical_cea_probability_profiles.csv",
        index=False,
        lineterminator="\n",
    )

    worked_profile = appendicitis_worked_profile(anchors)
    worked_profile.to_csv(
        analysis_dir / "appendicitis_worked_profile.csv",
        index=False,
        lineterminator="\n",
    )
    reporting_table = reporting_architecture_table()
    reporting_table.to_csv(
        analysis_dir / "table_1_reporting_architecture.csv",
        index=False,
        lineterminator="\n",
    )
    sensitivity, sensitivity_summary = estimator_prior_numerical_sensitivity(
        root, canonical, anchors
    )
    sensitivity.to_csv(
        analysis_dir / "estimator_prior_numerical_sensitivity_all_primary.csv",
        index=False,
        lineterminator="\n",
    )
    sensitivity_summary.to_csv(
        analysis_dir / "estimator_prior_numerical_sensitivity_summary.csv",
        index=False,
        lineterminator="\n",
    )
    contrasts, contrasts_summary = exploratory_within_review_contrasts(root, anchors)
    contrasts.to_csv(
        analysis_dir / "exploratory_within_review_contrasts.csv",
        index=False,
        lineterminator="\n",
    )
    write_json(
        analysis_dir / "exploratory_within_review_contrasts_summary.json",
        contrasts_summary,
    )

    version_diff = complete_version_diff(
        root,
        pd.read_csv(analysis_dir / "version_diff_all_releases.csv"),
        anchors,
    )
    version_diff.to_csv(
        analysis_dir / "version_diff_all_releases.csv",
        index=False,
        lineterminator="\n",
    )
    identities = information_identity_checks(anchors)

    claims = update_phase3_claim_ledger(
        root,
        anchor_summary,
        branch_summary,
        theory_summary,
        cea_crossings,
        worked_profile,
        sensitivity_summary,
        contrasts_summary,
        draws,
    )
    figure_table_map_path = write_phase3_figure_table_map(root)
    cea_lookup = cea_crossings.set_index("model_specification")
    appendicitis_lookup = worked_profile.set_index("element")
    phase3_summary = {
        "analysis_version": VERSION,
        "canonical_estimator": "continuity-corrected bivariate REML",
        "operating_points": int(anchors["group_id"].nunique()),
        "standard_anchor_rows": len(anchors),
        "continuous_profile_rows": len(probability_profiles),
        "continuous_profile_starting_probability_minimum": float(
            probability_profiles["starting_probability"].min()
        ),
        "continuous_profile_starting_probability_maximum": float(
            probability_profiles["starting_probability"].max()
        ),
        "continuous_profile_probability_increment": 0.01,
        "uncertainty_draws_per_operating_point": draws,
        "uncertainty_interpretation": (
            "Model-based 95% pooled-mean intervals describe uncertainty around the estimated mean; "
            "model-based 95% predictive ranges add fitted heterogeneity for an additional study's latent operating point."
        ),
        "standard_anchor_summary": anchor_summary.to_dict("records"),
        "information_identity_checks": identities,
        "theoretical_non_equivalence": theory_summary,
        "empirical_cea_crossing": {
            "canonical_reml_probability": float(
                cea_lookup.loc["v0.42.0 canonical REML", "crossing_probability"]
            ),
            "historical_reml_probability": float(
                cea_lookup.loc["v0.39.0 historical REML", "crossing_probability"]
            ),
            "regularized_gh15_probability": float(
                cea_lookup.loc[
                    "v0.40.0/v0.41.0 regularized GH15", "crossing_probability"
                ]
            ),
            "interpretation": "Empirical model-specific supplement sensitivity; not an action threshold.",
        },
        "appendicitis_worked_profile": {
            "group_id": APPENDICITIS_ID,
            "studies": int(appendicitis_lookup.loc["Study count", "value"]),
            "sensitivity": float(appendicitis_lookup.loc["Sensitivity", "value"]),
            "specificity": float(appendicitis_lookup.loc["Specificity", "value"]),
            "information_bits_at_20_percent": float(
                appendicitis_lookup.loc["Expected information", "value"]
            ),
            "fraction_entropy_removed_at_20_percent": float(
                appendicitis_lookup.loc[
                    "Fraction of starting entropy removed", "value"
                ]
            ),
            "post_positive_probability_at_20_percent": float(
                appendicitis_lookup.loc["Probability after a positive result", "value"]
            ),
            "post_negative_probability_at_20_percent": float(
                appendicitis_lookup.loc["Probability after a negative result", "value"]
            ),
            "action_status": str(
                appendicitis_lookup.loc["Clinical action", "availability_status"]
            ),
        },
        "exploratory_contrasts": contrasts_summary,
        "limitations": [
            "Continuity correction and REML model assumptions remain inherited limitations of the canonical hierarchy.",
            "Predictive ranges describe latent additional-study operating points, not observed binomial estimates or guaranteed new-setting performance.",
            "The continuous grid uses standardized illustrative starting probabilities from 0.01 through 0.50 and supplies no action thresholds.",
            "Within-review contrasts have no same-participant cross-test cells or covariance and therefore remain descriptive.",
        ],
    }
    write_json(analysis_dir / "phase3_summary.json", phase3_summary)

    phase1_twenty = canonical.set_index("group_id")
    phase3_twenty = anchors.loc[
        np.isclose(anchors["starting_probability"], 0.20)
    ].set_index("group_id")
    phase1_draw_reproduction = max(
        float(
            (
                phase3_twenty[f"pooled_mean_information_bits_{quantile}"]
                - phase1_twenty[
                    f"pooled_mean_information_bits_at_20_percent_{quantile}"
                ]
            )
            .abs()
            .max()
        )
        for quantile in ("low", "median", "high")
    )
    output_names = [
        "derived_operating_points_standard_anchors.csv",
        "derived_operating_points_probability_profiles.csv",
        "standard_anchor_summary.csv",
        "branch_entropy_all_primary.csv",
        "branch_entropy_summary.csv",
        "atlas_landmarks.csv",
        "theoretical_non_equivalence_examples.csv",
        "theoretical_crossing_probability_profile.csv",
        "theoretical_non_equivalence_summary.json",
        "empirical_cea_crossing_sensitivity.csv",
        "empirical_cea_probability_profiles.csv",
        "appendicitis_worked_profile.csv",
        "estimator_prior_numerical_sensitivity_all_primary.csv",
        "estimator_prior_numerical_sensitivity_summary.csv",
        "exploratory_within_review_contrasts.csv",
        "exploratory_within_review_contrasts_summary.json",
        "version_diff_all_releases.csv",
        "phase3_summary.json",
        "table_1_reporting_architecture.csv",
    ]
    all_claim_sources_exist = all(
        (root / source.strip()).exists()
        for source_field in claims["source_file"]
        if isinstance(source_field, str)
        for source in source_field.split(";")
        if source.strip().startswith(("analysis/", "docs/"))
    )
    near_equal = theory_summary[
        "near_equal_information_different_post_negative"
    ]
    theoretical_crossing = theory_summary[
        "probability_dependent_information_crossing"
    ]
    gate = {
        "phase": 3,
        "name": "canonical derived analysis and mathematical checks",
        "status": "candidate_pending_independent_qa",
        "candidate_generation_source_commit": PHASE3_SOURCE_COMMIT,
        "canonical_estimator": "continuity-corrected bivariate REML",
        "checks": {
            "all_273_points_have_three_standard_anchor_rows": bool(
                len(anchors) == 819
                and anchors.groupby("group_id").size().eq(3).all()
            ),
            "all_273_points_have_dense_continuous_profiles": bool(
                len(probability_profiles) == 13_650
                and probability_profiles.groupby("group_id").size().eq(50).all()
            ),
            "all_information_identities_pass": bool(
                identities["maximum_result_probability_sum_error"] < 1e-12
                and identities["maximum_entropy_reduction_identity_error_bits"]
                < 1e-12
                and identities["maximum_branch_kl_decomposition_error_bits"]
                < 1e-12
                and identities[
                    "maximum_fraction_entropy_removed_identity_error"
                ]
                < 1e-12
                and identities["minimum_information_bits"] >= -1e-12
                and identities["maximum_information_minus_starting_entropy_bits"]
                <= 1e-12
            ),
            "phase1_canonical_draws_reproduced": phase1_draw_reproduction < 1e-12,
            "theoretical_equal_information_tolerance_verified": bool(
                near_equal["absolute_information_difference_bits"] < 0.0001
                and near_equal["absolute_post_negative_difference"] > 0.10
            ),
            "theoretical_crossing_numerical_root_verified": bool(
                abs(theoretical_crossing["numerical_root"] - 0.28576588) < 1e-6
                and theoretical_crossing["root_residual_bits"] < 1e-12
            ),
            "empirical_cea_canonical_and_history_explicit": bool(
                abs(
                    cea_lookup.loc[
                        "v0.42.0 canonical REML", "crossing_probability"
                    ]
                    - 0.1280262761348617
                )
                < 1e-8
                and abs(
                    cea_lookup.loc[
                        "v0.40.0/v0.41.0 regularized GH15",
                        "crossing_probability",
                    ]
                    - 0.38086130898554993
                )
                < 1e-8
            ),
            "appendicitis_profile_complete_with_action_open": bool(
                worked_profile["element"].eq("Clinical action").sum() == 1
                and worked_profile.loc[
                    worked_profile["element"].eq("Clinical action"), "value"
                ].isna().all()
                and worked_profile.loc[
                    worked_profile["element"].eq("Clinical action"),
                    "availability_status",
                ]
                .str.contains("not determined", regex=False)
                .all()
            ),
            "estimator_prior_numerical_sensitivity_complete": len(sensitivity)
            == 4095,
            "no_mixed_estimator_point_and_interval": bool(
                ~sensitivity["central_estimator_mixed_with_canonical_interval"].any()
            ),
            "direct_binomial_released_medians_reconciled": bool(
                sensitivity.loc[
                    sensitivity["model_specification"].str.contains(
                        "regularized", case=False
                    ),
                    "derived_central_matches_attached_interval_median",
                ].all()
            ),
            "exploratory_contrasts_are_not_paired_inference": bool(
                len(contrasts) == 147
                and not contrasts["paired_inference_supported"].any()
                and contrasts_summary["same_participants_established_contrasts"]
                == 0
                and contrasts_summary[
                    "cross_test_covariance_available_contrasts"
                ]
                == 0
            ),
            "version_diff_complete_for_all_releases_and_anchors": bool(
                len(version_diff) == 273
                and version_diff["analysis_status_v042"].notna().all()
            ),
            "all_claim_sources_exist": all_claim_sources_exist,
            "figure_table_map_sources_exist": all(
                (root / source).exists()
                for source in [
                    token
                    for line in figure_table_map_path.read_text(encoding="utf-8").splitlines()
                    for token in line.split("`")[1::2]
                ]
            ),
            "display_claim_coverage_complete": set(
                [f"C{number:03d}" for number in range(12, 26)]
            ).issubset(set(claims["claim_id"])),
            "appendicitis_provenance_exact": bool(
                worked_profile.loc[
                    worked_profile["element"].eq("Index test context from review title"),
                    "value_text",
                ].eq("Non-contrast CT").all()
                and worked_profile.loc[
                    worked_profile["element"].eq("Locked operating-point test_name"),
                    "value_text",
                ].eq("Overall structured 2 x 2 data").all()
            ),
            "branch_claim_matches_observed_pattern": bool(
                anchor_summary["negative_result_increases_entropy_count"].eq(0).all()
                and anchor_summary.set_index("starting_probability")[
                    "positive_result_increases_entropy_count"
                ].to_dict()
                == {0.05: 273, 0.2: 232, 0.5: 0}
            ),
            "no_exploratory_sign_count_is_primary": bool(
                claims.loc[
                    claims["source_file"].eq(
                        "analysis/bmj_rmr_v042/exploratory_within_review_contrasts.csv"
                    ),
                    "claim_role",
                ].eq("exploratory").all()
            ),
        },
        "information_identity_checks": identities,
        "phase1_draw_reproduction_maximum_absolute_difference": phase1_draw_reproduction,
        "theoretical_non_equivalence": theory_summary,
        "empirical_cea_crossing": phase3_summary["empirical_cea_crossing"],
        "appendicitis_worked_profile": phase3_summary[
            "appendicitis_worked_profile"
        ],
        "exploratory_contrasts": contrasts_summary,
        "commands": [
            "py -3.13 scripts/bmj_rmr_v042.py phase3",
            "py -3.13 -m pytest tests/test_bmj_rmr_v042_phase3.py -q",
            "py -3.13 -m pytest tests/test_bmj_rmr_v042.py tests/test_bmj_rmr_v042_spec.py tests/test_bmj_rmr_v042_phase3.py tests/test_cochrane_dta_atlas.py -q",
            "git diff --check",
        ],
        "builder_verification": {
            "targeted_phase3_tests": "16 passed in 1.29s; one unrelated pytest-asyncio deprecation warning",
            "full_relevant_tests": "62 passed in 1.69s; one unrelated pytest-asyncio deprecation warning",
            "git_diff_check": "passed after corrective Phase 3 generation and control-file updates",
        },
        "output_sha256": {
            name: file_hash(analysis_dir / name) for name in output_names
        },
        "qa_context": "pending fresh information-theory and numerical-methods review",
        "independent_qa_required": True,
        "corrective_source_commit": PHASE3_INITIAL_CANDIDATE_COMMIT,
        "qa_history": {
            "held_candidate_commit": PHASE3_INITIAL_CANDIDATE_COMMIT,
            "verdict": "hold",
            "blocking_findings": [
                "Direct-binomial derived central quantities were plug-in calculations but were displayed beside propagated-median intervals.",
                "The figure/table map contained stale or nonexistent sources and incomplete display-claim coverage.",
                "Claim C016 exceeded the observed canonical negative-branch pattern.",
            ],
            "corrective_findings": [
                "The appendicitis profile conflated review-title non-contrast CT context with the locked test_name."
            ],
            "resolution_status": "builder corrections complete; pending fresh independent re-QA",
        },
        "limitations": phase3_summary["limitations"],
    }
    gate["output_sha256"]["CLAIM_LEDGER.csv"] = file_hash(
        root / "docs/manuscript/v19/CLAIM_LEDGER.csv"
    )
    gate["output_sha256"]["FIGURE_TABLE_MAP.md"] = file_hash(
        figure_table_map_path
    )
    write_json(qa_dir / "phase_gate_3.json", gate)

    run_manifest_path = analysis_dir / "run_manifest.json"
    run_manifest = json.loads(run_manifest_path.read_text(encoding="utf-8"))
    run_manifest.update(
        {
            "phase": 3,
            "status": "phase_3_corrective_candidate_pending_independent_qa",
            "phase_3_source_commit": PHASE3_SOURCE_COMMIT,
            "phase_3_initial_candidate_commit": PHASE3_INITIAL_CANDIDATE_COMMIT,
            "phase_3_gate": "analysis/bmj_rmr_v042/qa/phase_gate_3.json",
            "phase_3_summary": "analysis/bmj_rmr_v042/phase3_summary.json",
            "role": "Phase 3 held-QA findings corrected without changing the canonical estimator or 273-point sample; pending fresh independent QA",
        }
    )
    write_json(run_manifest_path, run_manifest)
    return gate


def parse_args() -> argparse.Namespace:
    parser = argparse.ArgumentParser()
    parser.add_argument("phase", choices=["phase0", "phase1", "phase2", "phase3"])
    parser.add_argument("--root", type=Path, default=Path(__file__).resolve().parents[1])
    parser.add_argument("--draws", type=int, default=20_000)
    parser.add_argument(
        "--reuse-derived",
        action="store_true",
        help="Reuse complete Phase 3 anchor/profile files with the requested draw count.",
    )
    return parser.parse_args()


def main() -> int:
    args = parse_args()
    if args.phase == "phase0":
        result = phase0(args.root.resolve())
    elif args.phase == "phase1":
        result = phase1(args.root.resolve(), args.draws)
    elif args.phase == "phase2":
        result = phase2(args.root.resolve())
    else:
        result = phase3(args.root.resolve(), args.draws, args.reuse_derived)
    print(json.dumps(result, indent=2, sort_keys=True))
    return 0


if __name__ == "__main__":
    raise SystemExit(main())
```
