## Supplementary material for "The information in diagnostic tests": Executable analysis code, derived inputs, source-acquisition records, tests, and release metadata: bmj_rmr_v042_visuals.py

```
"""Deterministic displays for the BMJ RMR v0.42.5 Figure 1 correction.

All numerical labels, display-source tables, captions, and editable tables are
derived from the machine-readable Phase 1--3 outputs.  The module does
not refit models and contains no manuscript headline values as literals.
"""

from __future__ import annotations

import argparse
import hashlib
import importlib
import json
import math
import re
import shutil
import zipfile
from datetime import datetime, timezone
from pathlib import Path
from textwrap import fill
from xml.etree import ElementTree

import matplotlib

matplotlib.use("Agg")
import matplotlib.pyplot as plt
import fitz
import numpy as np
import pandas as pd
import scienceplots  # noqa: F401  # Registers the SciencePlots Matplotlib styles.
from docx import Document
from docx.enum.table import WD_CELL_VERTICAL_ALIGNMENT
from docx.enum.text import WD_ALIGN_PARAGRAPH
from docx.oxml import OxmlElement
from docx.oxml.ns import qn
from docx.shared import Inches, Pt
from matplotlib.lines import Line2D
from matplotlib.patches import FancyArrowPatch, FancyBboxPatch, Rectangle
from PIL import Image, ImageDraw, ImageFont


VERSION = "0.42.5"
RELEASE_DATE = "2026-08-28"
DPI = 600
MM_PER_INCH = 25.4
DOUBLE_COLUMN_MM = 180.0
SOURCE_DATE = datetime(2026, 8, 28, tzinfo=timezone.utc)
PDF_TEXT_BOUND_TOLERANCE_PT = 0.1
BMJ_STYLE = Path(__file__).resolve().parents[1] / "styles/bmj.mplstyle"

INK = "#17212B"
MUTED = "#58636D"
GRID = "#D9E0E5"
LIGHT = "#F5F7F9"
WHITE = "#FFFFFF"
QUESTION_COLORS = ("#0072B2", "#009E73", "#CC79A7", "#E69F00")
QUESTION_GRAYS = ("#111111", "#3D3D3D", "#696969", "#969696")
QUESTION_MARKERS = ("o", "s", "^", "D")
QUESTION_LINESTYLES = ("-", "--", ":", "-.")
HATCHES = ("", "///", "xx", "..")

APPENDICITIS_ID = (
    "pmc open access:PMC12734481:medicina-61-02163-t001:"
    "table-1-summary-of-the-characteristics-of-the-included-studies:1:overall"
)

DISPLAY_ORDER = (
    "Figure 1",
    "Figure 2",
    "Figure 3",
    "Figure 4",
    "Table 1",
    "Table 2",
    "Figure S1",
    "Figure S2",
    "Figure S3",
    "Figure S4",
    "Figure S5",
    "Figure S6",
    "Figure S7",
    "Figure S8",
)

FIGURE_SPECS: dict[str, dict[str, object]] = {
    "Figure 1": {"stem": "figure_1_framework_and_worked_profile", "height_mm": 118.0},
    "Figure 2": {"stem": "figure_2_signature_atlas", "height_mm": 125.0},
    "Figure 3": {"stem": "figure_3_three_non_equivalences", "height_mm": 118.0},
    "Figure 4": {"stem": "figure_4_result_branch_entropy", "height_mm": 120.0},
    "Figure S1": {"stem": "figure_s1_corpus_and_selection_flow", "height_mm": 150.0},
    "Figure S2": {"stem": "figure_s2_validation_and_route_stability", "height_mm": 125.0},
    "Figure S3": {"stem": "figure_s3_information_distributions_and_youden", "height_mm": 122.0},
    "Figure S4": {"stem": "figure_s4_recovery_and_diagnostics", "height_mm": 122.0},
    "Figure S5": {"stem": "figure_s5_estimator_sensitivity", "height_mm": 122.0},
    "Figure S6": {"stem": "figure_s6_exploratory_contrasts_and_movement", "height_mm": 122.0},
    "Figure S7": {"stem": "figure_s7_near_ties_and_composition_audit", "height_mm": 118.0},
    "Figure S8": {"stem": "figure_s8_cea_crossing_and_theoretical_checks", "height_mm": 122.0},
}

DISPLAY_SOURCES: dict[str, tuple[str, ...]] = {
    "Figure 1": (
        "analysis/bmj_rmr_v042/appendicitis_worked_profile.csv",
        "analysis/bmj_rmr_v042/table_1_reporting_architecture.csv",
    ),
    "Figure 2": (
        "analysis/bmj_rmr_v042/derived_operating_points_standard_anchors.csv",
        "analysis/bmj_rmr_v042/atlas_landmarks.csv",
    ),
    "Figure 3": (
        "analysis/bmj_rmr_v042/theoretical_non_equivalence_examples.csv",
        "analysis/bmj_rmr_v042/theoretical_crossing_probability_profile.csv",
        "analysis/bmj_rmr_v042/theoretical_non_equivalence_summary.json",
    ),
    "Figure 4": (
        "analysis/bmj_rmr_v042/branch_entropy_all_primary.csv",
        "analysis/bmj_rmr_v042/branch_entropy_summary.csv",
    ),
    "Table 1": ("analysis/bmj_rmr_v042/table_1_reporting_architecture.csv",),
    "Table 2": ("analysis/bmj_rmr_v042/appendicitis_worked_profile.csv",),
    "Figure S1": (
        "analysis/bmj_rmr_v042/run_manifest.json",
        "analysis/bmj_rmr_v042/source_trace_summary.json",
        "analysis/bmj_rmr_v042/analysis_row_selection_exceptions.csv",
    ),
    "Figure S2": (
        "analysis/bmj_rmr_v042/source_summary_fidelity.csv",
        "analysis/bmj_rmr_v042/manual_audit_summary.json",
        "analysis/bmj_rmr_v042/source_route_stability.csv",
        "analysis/bmj_rmr_v042/source_summary_large_discrepancies.csv",
    ),
    "Figure S3": (
        "analysis/bmj_rmr_v042/derived_operating_points_standard_anchors.csv",
        "analysis/bmj_rmr_v042/standard_anchor_summary.csv",
    ),
    "Figure S4": (
        "analysis/bmj_rmr_v042/reml_recovery.csv",
        "analysis/bmj_rmr_v042/reml_recovery_summary.json",
        "analysis/bmj_rmr_v042/reml_diagnostics_all_primary.csv",
        "analysis/bmj_rmr_v042/reml_diagnostics_summary.json",
    ),
    "Figure S5": (
        "analysis/bmj_rmr_v042/estimator_prior_numerical_sensitivity_all_primary.csv",
        "analysis/bmj_rmr_v042/estimator_prior_numerical_sensitivity_summary.csv",
        "analysis/bmj_rmr_v042/estimator_sensitivity_summary.json",
    ),
    "Figure S6": (
        "analysis/bmj_rmr_v042/exploratory_within_review_contrasts.csv",
        "analysis/bmj_rmr_v042/exploratory_within_review_contrasts_summary.json",
        "analysis/bmj_rmr_v042/estimator_prior_numerical_sensitivity_all_primary.csv",
    ),
    "Figure S7": (
        "analysis/bmj_rmr_v042/exploratory_within_review_contrasts.csv",
        "analysis/bmj_rmr_v042/derived_operating_points_standard_anchors.csv",
    ),
    "Figure S8": (
        "analysis/bmj_rmr_v042/empirical_cea_crossing_sensitivity.csv",
        "analysis/bmj_rmr_v042/empirical_cea_probability_profiles.csv",
        "analysis/bmj_rmr_v042/theoretical_non_equivalence_summary.json",
    ),
}

DISPLAY_CLAIMS: dict[str, str] = {
    "Figure 1": "C018; C021; C023; C024; C025",
    "Figure 2": "C001; C012; C013",
    "Figure 3": "C014; C015; C022",
    "Figure 4": "C016",
    "Table 1": "C025",
    "Table 2": "C018; C021; C023; C024",
    "Figure S1": "C001; C002; C011",
    "Figure S2": "C003; C004; C010; C026",
    "Figure S3": "C012; C013",
    "Figure S4": "C006; C008; C009; C028",
    "Figure S5": "C019",
    "Figure S6": "C020",
    "Figure S7": "C020",
    "Figure S8": "C014; C015; C017; C022",
}

DISPLAY_SOURCE_ROLES: dict[str, tuple[str, ...]] = {
    "Figure 1": ("worked profile values and missing action inputs", "four-question reporting architecture"),
    "Figure 2": ("complete primary 20% Atlas population", "shape-coded orientation landmarks"),
    "Figure 3": ("specified theoretical operating points", "probability-dependent information profiles", "independent theoretical checks and roots"),
    "Figure 4": ("all point-level branch entropy movements", "standard-anchor branch-count summary"),
    "Table 1": ("four-question reporting architecture",),
    "Table 2": ("worked profile values, uncertainty, and missing action inputs",),
    "Figure S1": ("fixed corpus and selection counts", "row-level source-identity summary", "documented row-selection exception"),
    "Figure S2": ("source-summary fidelity values", "recorded author-audit denominators", "five-route descriptive variation summaries", "metadata-bounded explanations for the three large discrepancies"),
    "Figure S3": ("complete standard-anchor point distributions", "anchor medians and rank concordance"),
    "Figure S4": ("scenario-level REML recovery values", "scenario-inclusion summary", "all point-level fit diagnostics", "primary fit-diagnostic count summary"),
    "Figure S5": ("all point-level analysis-family sensitivity values", "model-level analysis-family sensitivity summary", "like-for-like plug-in central-estimate comparison"),
    "Figure S6": ("exploratory within-review contrast magnitudes", "preclassified contrast summary", "model-specification movement inputs"),
    "Figure S7": ("near-tie and composition-restricted contrast inputs", "primary anchor context"),
    "Figure S8": ("model-specific empirical crossing roots", "empirical crossing profiles", "independent theoretical checks"),
}

DISPLAY_SOURCE_CLAIMS: dict[str, tuple[tuple[str, ...], ...]] = {
    "Figure 1": (("C018", "C021", "C023", "C024"), ("C025",)),
    "Figure 2": (("C001", "C012", "C013"), ()),
    "Figure 3": (("C014", "C015"), ("C022",), ("C014", "C015", "C022")),
    "Figure 4": (("C016",), ("C016",)),
    "Table 1": (("C025",),),
    "Table 2": (("C018", "C021", "C023", "C024"),),
    "Figure S1": (("C001",), ("C011",), ("C002",)),
    "Figure S2": (("C003",), ("C004",), ("C010",), ("C026",)),
    "Figure S3": (("C012", "C013"), ("C012", "C013")),
    "Figure S4": (("C008",), ("C008",), ("C009", "C028"), ("C006", "C009", "C028")),
    "Figure S5": (("C019",), ("C019",), ("C019",)),
    "Figure S6": (("C020",), ("C020",), ("C020",)),
    "Figure S7": (("C020",), ()),
    "Figure S8": (("C017",), ("C017",), ("C014", "C015", "C022")),
}

COLLISION_AUDIT_STEMS = (
    "figure_2_signature_atlas",
    "figure_s4_recovery_and_diagnostics",
    "figure_s7_near_ties_and_composition_audit",
)

EXPECTED_SOURCE_ROWS: dict[str, int] = {
    "appendicitis_worked_profile.csv": 28,
    "table_1_reporting_architecture.csv": 4,
    "derived_operating_points_standard_anchors.csv": 819,
    "atlas_landmarks.csv": 4,
    "theoretical_non_equivalence_examples.csv": 6,
    "theoretical_crossing_probability_profile.csv": 100,
    "branch_entropy_all_primary.csv": 819,
    "branch_entropy_summary.csv": 7,
    "analysis_row_selection_exceptions.csv": 1,
    "source_summary_fidelity.csv": 92,
    "source_route_stability.csv": 15,
    "reml_recovery.csv": 24,
    "reml_diagnostics_all_primary.csv": 273,
    "estimator_prior_numerical_sensitivity_all_primary.csv": 4095,
    "estimator_prior_numerical_sensitivity_summary.csv": 15,
    "exploratory_within_review_contrasts.csv": 147,
    "empirical_cea_crossing_sensitivity.csv": 5,
    "empirical_cea_probability_profiles.csv": 500,
    "standard_anchor_summary.csv": 3,
}


def file_hash(path: Path) -> str:
    digest = hashlib.sha256()
    with path.open("rb") as handle:
        for block in iter(lambda: handle.read(1024 * 1024), b""):
            digest.update(block)
    return digest.hexdigest()


def write_json(path: Path, payload: object) -> None:
    path.parent.mkdir(parents=True, exist_ok=True)
    path.write_text(
        json.dumps(payload, indent=2, sort_keys=True) + "\n",
        encoding="utf-8",
        newline="\n",
    )


def write_csv(path: Path, frame: pd.DataFrame) -> None:
    path.parent.mkdir(parents=True, exist_ok=True)
    frame.to_csv(path, index=False, lineterminator="\n", float_format="%.15g")


def read_json(path: Path) -> dict[str, object]:
    return json.loads(path.read_text(encoding="utf-8"))


def colors(grayscale: bool) -> tuple[str, ...]:
    return QUESTION_GRAYS if grayscale else QUESTION_COLORS


def set_style(grayscale: bool) -> None:
    if "science" not in plt.style.available:
        importlib.reload(scienceplots)
    plt.style.use(["science", "no-latex", str(BMJ_STYLE)])
    matplotlib.rcParams.update(
        {
            "svg.hashsalt": "bmj-rmr-v042-phase4",
            "text.color": INK if not grayscale else "#111111",
            "axes.labelcolor": INK if not grayscale else "#111111",
            "axes.edgecolor": MUTED if not grayscale else "#444444",
            "xtick.color": MUTED if not grayscale else "#444444",
            "ytick.color": MUTED if not grayscale else "#444444",
        }
    )


def figure_size(height_mm: float) -> tuple[float, float]:
    return DOUBLE_COLUMN_MM / MM_PER_INCH, height_mm / MM_PER_INCH


def new_figure(display: str, grayscale: bool, **kwargs: object) -> tuple[plt.Figure, object]:
    set_style(grayscale)
    spec = FIGURE_SPECS[display]
    return plt.subplots(figsize=figure_size(float(spec["height_mm"])), **kwargs)


def save_figure(fig: plt.Figure, directory: Path, display: str, grayscale: bool) -> list[Path]:
    directory.mkdir(parents=True, exist_ok=True)
    stem = str(FIGURE_SPECS[display]["stem"])
    if grayscale:
        stem += "_grayscale"
    outputs: list[Path] = []
    common = {"facecolor": WHITE, "edgecolor": "none"}
    pdf = directory / f"{stem}.pdf"
    fig.savefig(
        pdf,
        format="pdf",
        metadata={
            "Title": display,
            "Author": f"BMJ RMR v{VERSION} deterministic display builder",
            "Creator": "scripts/bmj_rmr_v042_visuals.py",
            "CreationDate": None,
            "ModDate": None,
        },
        **common,
    )
    outputs.append(pdf)
    svg = directory / f"{stem}.svg"
    fig.savefig(
        svg,
        format="svg",
        metadata={
            "Title": display,
            "Creator": "scripts/bmj_rmr_v042_visuals.py",
            "Date": RELEASE_DATE,
        },
        **common,
    )
    svg.write_text(
        "\n".join(line.rstrip() for line in svg.read_text(encoding="utf-8").splitlines()) + "\n",
        encoding="utf-8",
        newline="\n",
    )
    outputs.append(svg)
    png = directory / f"{stem}.png"
    fig.savefig(
        png,
        format="png",
        dpi=DPI,
        metadata={"Software": f"BMJ RMR v{VERSION} deterministic display builder"},
        **common,
    )
    outputs.append(png)
    plt.close(fig)
    return outputs


def panel_label(ax: plt.Axes, label: str, title: str, classification: str | None = None) -> None:
    title_text = f"{label}  {fill(title, width=28)}"
    if classification:
        title_text += f"\n{classification.upper()}"
    ax.set_title(title_text, loc="left", weight="bold", pad=4, fontsize=8.0, linespacing=1.12)


def figure_footer(fig: plt.Figure, value: str, y: float = 0.025, width: int = 125, fontsize: float = 8.0) -> None:
    fig.text(0.5, y, fill(value, width=width), ha="center", va="bottom", fontsize=fontsize, color=MUTED, linespacing=1.15)


def grid_y(ax: plt.Axes) -> None:
    ax.grid(axis="y", color=GRID, lw=0.55, zorder=0)


def grid_both(ax: plt.Axes) -> None:
    ax.grid(color=GRID, lw=0.5, zorder=0)


def pct(value: float, decimals: int = 1) -> str:
    return f"{100 * float(value):.{decimals}f}%"


def interval(low: float, high: float, *, percent: bool = False, decimals: int = 1) -> str:
    if percent:
        return f"{100 * float(low):.{decimals}f}–{100 * float(high):.{decimals}f}%"
    return f"{float(low):.{decimals}f}–{float(high):.{decimals}f}"


def profile_lookup(profile: pd.DataFrame) -> pd.DataFrame:
    return profile.set_index("element", drop=False)


def profile_row(profile: pd.DataFrame, element: str) -> pd.Series:
    return profile_lookup(profile).loc[element]


def figure_1(profile: pd.DataFrame, architecture: pd.DataFrame, grayscale: bool) -> plt.Figure:
    set_style(grayscale)
    fig, ax = plt.subplots(figsize=figure_size(float(FIGURE_SPECS["Figure 1"]["height_mm"])))
    ax.set_xlim(0, 1)
    ax.set_ylim(0, 1)
    ax.axis("off")
    fig.subplots_adjust(left=0.035, right=0.985, top=0.97, bottom=0.055)

    palette = list(colors(grayscale))
    if grayscale:
        palette[-1] = "#555555"
    rule = "#AEB8C0" if not grayscale else "#9A9A9A"
    soft_rule = "#DCE2E7" if not grayscale else "#D4D4D4"
    predictive = "#B8C2CA" if not grayscale else "#B5B5B5"
    columns = ((0.015, 0.245), (0.265, 0.495), (0.515, 0.745), (0.765, 0.995))
    centers = tuple((left + right) / 2 for left, right in columns)

    n_studies = int(float(profile_row(profile, "Study count")["value"]))
    se = profile_row(profile, "Sensitivity")
    se_pr = profile_row(profile, "Sensitivity predictive")
    sp = profile_row(profile, "Specificity")
    sp_pr = profile_row(profile, "Specificity predictive")
    info = profile_row(profile, "Expected information")
    info_pr = profile_row(profile, "Expected information predictive")
    fraction = profile_row(profile, "Fraction of starting entropy removed")
    p_pos = profile_row(profile, "Probability of a positive result")
    p_post_pos = profile_row(profile, "Probability after a positive result")
    p_neg = profile_row(profile, "Probability of a negative result")
    p_post_neg = profile_row(profile, "Probability after a negative result")
    starting = float(profile_row(profile, "Illustrative starting probability")["value"])

    # Panel A: the reporting sequence. Direct labels carry meaning if colour is absent.
    ax.text(0.015, 0.965, "A", fontsize=10.5, weight="bold", color=INK, va="center")
    ax.text(0.045, 0.965, "Four questions to report separately", fontsize=9.5, weight="semibold", color=INK, va="center")
    titles = ("Operating point", "Expected information", "Observed result", "Action")
    questions = (
        "How did the test\nperform?",
        "How much uncertainty\nis removed?",
        "What probability follows\nthis result?",
        "Do consequences\nsupport action?",
    )
    for index, ((left, right), x, title, question, color) in enumerate(
        zip(columns, centers, titles, questions, palette), start=1
    ):
        if index < 4:
            ax.add_patch(
                FancyArrowPatch(
                    (x + 0.035, 0.855),
                    (centers[index] - 0.035, 0.855),
                    arrowstyle="-|>",
                    mutation_scale=7,
                    linewidth=0.8,
                    color=rule,
                    shrinkA=0,
                    shrinkB=0,
                )
            )
        ax.scatter([x], [0.855], s=150, marker=QUESTION_MARKERS[index - 1], color=color, edgecolor=WHITE, linewidth=0.8, zorder=3)
        ax.text(x, 0.855, str(index), color=WHITE, fontsize=7.7, weight="bold", ha="center", va="center", zorder=4)
        ax.text(x, 0.795, f"Q{index}  {title}", color=color, fontsize=8.3, weight="semibold", ha="center", va="top")
        ax.text(x, 0.735, question, color=INK, fontsize=7.7, ha="center", va="top", linespacing=1.15)

    ax.plot([0.015, 0.995], [0.65, 0.65], color=rule, lw=0.8)

    # Panel B: the same four columns now carry real data rather than a text table.
    ax.text(0.015, 0.615, "B", fontsize=10.5, weight="bold", color=INK, va="center")
    ax.text(0.045, 0.615, "Worked profile: non-contrast CT for appendicitis", fontsize=9.5, weight="semibold", color=INK, va="center")
    ax.text(
        0.995,
        0.615,
        f"{n_studies} studies  |  starting probability {pct(starting, 0)} (illustrative)",
        fontsize=7.5,
        color=MUTED,
        ha="right",
        va="center",
    )

    for index, ((left, right), title, color) in enumerate(zip(columns, titles, palette), start=1):
        ax.plot([left, right], [0.555, 0.555], color=color, lw=2.2, solid_capstyle="butt")
        ax.text(left, 0.535, f"Q{index}", fontsize=7.6, weight="bold", color=color, va="top")
        ax.text(left + 0.035, 0.535, title, fontsize=8.1, weight="semibold", color=INK, va="top")
        if index < 4:
            ax.plot([right + 0.01, right + 0.01], [0.11, 0.545], color=soft_rule, lw=0.7)

    # Q1: central estimates, pooled-mean intervals, and predictive ranges.
    q1_left, q1_right = columns[0]
    axis_left, axis_right = q1_left + 0.045, q1_right - 0.01

    def q1_x(value: float) -> float:
        return axis_left + (value - 0.80) / 0.20 * (axis_right - axis_left)

    for y, label, estimate, pooled_low, pooled_high, pred_low, pred_high in (
        (0.415, "Se", se.value, se.low, se.high, se_pr.low, se_pr.high),
        (0.315, "Sp", sp.value, sp.low, sp.high, sp_pr.low, sp_pr.high),
    ):
        ax.text(q1_left, y + 0.045, f"{label}  {pct(estimate)}", fontsize=8.5, weight="bold", color=INK, va="center")
        ax.plot([q1_x(pred_low), q1_x(pred_high)], [y, y], color=predictive, lw=2.4, solid_capstyle="round")
        ax.plot([q1_x(pooled_low), q1_x(pooled_high)], [y, y], color=palette[0], lw=4.2, solid_capstyle="round")
        ax.scatter([q1_x(estimate)], [y], s=30, marker=QUESTION_MARKERS[0], color=palette[0], edgecolor=WHITE, linewidth=0.7, zorder=4)
    ax.plot([axis_left, axis_right], [0.245, 0.245], color=rule, lw=0.6)
    for value in (0.80, 0.90, 1.00):
        ax.plot([q1_x(value), q1_x(value)], [0.24, 0.25], color=rule, lw=0.6)
        ax.text(q1_x(value), 0.222, pct(value, 0), fontsize=6.7, color=MUTED, ha="center", va="top")

    # Q2: expected information relative to the starting entropy.
    q2_left, q2_right = columns[1]
    entropy_start = info.value / fraction.value
    info_axis_left, info_axis_right = q2_left + 0.01, q2_right - 0.01

    def info_x(value: float) -> float:
        return info_axis_left + value / entropy_start * (info_axis_right - info_axis_left)

    ax.text(q2_left, 0.445, f"{info.value:.3f} bits", fontsize=12.0, weight="bold", color=INK, va="center")
    ax.text(q2_left, 0.392, f"{pct(fraction.value)} of starting entropy", fontsize=7.5, color=MUTED, va="center")
    ax.plot([info_axis_left, info_axis_right], [0.315, 0.315], color=soft_rule, lw=5.0, solid_capstyle="round")
    ax.plot([info_x(info_pr.low), info_x(info_pr.high)], [0.315, 0.315], color=predictive, lw=2.4, solid_capstyle="round")
    ax.plot([info_x(info.low), info_x(info.high)], [0.315, 0.315], color=palette[1], lw=4.2, solid_capstyle="round")
    ax.scatter([info_x(info.value)], [0.315], s=32, marker=QUESTION_MARKERS[1], color=palette[1], edgecolor=WHITE, linewidth=0.7, zorder=4)
    ax.text(info_axis_left, 0.275, "0", fontsize=6.7, color=MUTED, ha="center", va="top")
    ax.text(info_axis_right, 0.275, f"{entropy_start:.3f} bits", fontsize=6.7, color=MUTED, ha="center", va="top")

    # Q3: the observed result branches from one starting probability.
    q3_left, q3_right = columns[2]
    start_x = q3_left + 0.035
    branch_x = q3_left + 0.12
    ax.text(start_x, 0.365, f"Start\n{pct(starting, 0)}", fontsize=8.0, weight="semibold", color=INK, ha="center", va="center", linespacing=1.05)
    for y, symbol, result_probability, post_probability in (
        (0.425, "+", p_pos.value, p_post_pos.value),
        (0.295, "−", p_neg.value, p_post_neg.value),
    ):
        ax.add_patch(
            FancyArrowPatch(
                (start_x + 0.028, 0.365),
                (branch_x - 0.012, y),
                arrowstyle="-|>",
                mutation_scale=7,
                linewidth=1.0,
                color=palette[2],
                shrinkA=0,
                shrinkB=0,
            )
        )
        ax.text(branch_x, y + 0.024, f"{symbol} result  ({pct(result_probability)})", fontsize=7.0, color=MUTED, va="bottom")
        ax.text(branch_x, y, pct(post_probability), fontsize=11.0, weight="bold", color=INK, va="center")
    # Q4: absence of decision inputs is an explicit result, not a decorative warning.
    q4_left, _ = columns[3]
    ax.text(q4_left, 0.435, "Undetermined", fontsize=11.0, weight="bold", color=palette[3], va="center")
    ax.text(q4_left, 0.37, "Action needs:", fontsize=7.4, weight="semibold", color=INK, va="top")
    ax.text(
        q4_left,
        0.335,
        "consequence model\naction threshold\ncosts and alternatives\npatient preferences",
        fontsize=7.4,
        color=MUTED,
        va="top",
        linespacing=1.25,
    )

    # One compact key applies to the interval marks in Q1 and Q2.
    key_y = 0.08
    ax.plot([0.015, 0.045], [key_y, key_y], color=predictive, lw=2.4, solid_capstyle="round")
    ax.text(0.052, key_y, "predictive range", fontsize=6.8, color=MUTED, va="center")
    ax.plot([0.22, 0.25], [key_y, key_y], color=palette[0], lw=4.2, solid_capstyle="round")
    ax.text(0.257, key_y, "pooled-mean 95% interval", fontsize=6.8, color=MUTED, va="center")
    ax.scatter([0.48], [key_y], s=26, marker="o", color=INK, edgecolor=WHITE, linewidth=0.6, zorder=4)
    ax.text(0.492, key_y, "central estimate", fontsize=6.8, color=MUTED, va="center")
    return fig


def figure_2(anchors: pd.DataFrame, landmarks: pd.DataFrame, grayscale: bool) -> plt.Figure:
    fig, axes = new_figure(
        "Figure 2", grayscale, nrows=1, ncols=3, gridspec_kw={"width_ratios": [1.22, 0.78, 1.08]}
    )
    palette = colors(grayscale)
    twenty = anchors.loc[np.isclose(anchors["starting_probability"], 0.20)].copy()
    grid = np.linspace(0.001, 0.999, 180)
    se_grid, sp_grid = np.meshgrid(grid, grid)
    p0 = float(twenty["starting_probability"].iloc[0])
    positive = p0 * se_grid + (1 - p0) * (1 - sp_grid)
    negative = p0 * (1 - se_grid) + (1 - p0) * sp_grid
    post_pos = p0 * se_grid / positive
    post_neg = p0 * (1 - se_grid) / negative
    entropy = lambda p: -(p * np.log2(np.clip(p, 1e-12, 1)) + (1 - p) * np.log2(np.clip(1 - p, 1e-12, 1)))
    information = entropy(p0) - positive * entropy(post_pos) - negative * entropy(post_neg)
    colorbar_contours = None
    if grayscale:
        contours = axes[0].contour(100 * se_grid, 100 * sp_grid, information, levels=np.arange(0.1, 0.8, 0.1), colors="#777777", linewidths=0.65)
        axes[0].clabel(contours, fontsize=8.0, fmt="%.1f")
    else:
        colorbar_contours = axes[0].contourf(100 * se_grid, 100 * sp_grid, information, levels=np.linspace(0, 0.72, 13), cmap="cividis")
    axes[0].plot([0, 100], [100, 0], color="#FFFFFF" if not grayscale else "#555555", ls="--", lw=0.9)
    axes[0].scatter(100 * twenty.sensitivity, 100 * twenty.specificity, s=12, facecolor=WHITE, edgecolor="#2F3B43", linewidth=0.45, alpha=0.72)
    for row, color, marker in zip(landmarks.itertuples(index=False), palette, QUESTION_MARKERS):
        axes[0].scatter(100 * row.sensitivity, 100 * row.specificity, s=48, marker=marker, color=color, edgecolor=WHITE, linewidth=0.6, zorder=4)
    axes[0].set(xlim=(0, 100), ylim=(0, 100), xlabel="Sensitivity (%)", ylabel="Specificity (%)")
    axes[0].set_xticks([0, 25, 50, 75, 100])
    axes[0].set_yticks([0, 25, 50, 75, 100])
    panel_label(axes[0], "A", f"Information contours at {pct(p0, 0)}", "empirical")

    fraction = 100 * twenty["fraction_entropy_removed"]
    bins = np.linspace(0, 100, 21)
    axes[1].hist(fraction, bins=bins, color=palette[1], edgecolor=WHITE, linewidth=0.6, hatch=HATCHES[1])
    median = float(fraction.median())
    axes[1].axvline(median, color=palette[3], lw=1.6, ls=QUESTION_LINESTYLES[3])
    axes[1].text(median + 2, axes[1].get_ylim()[1] * 0.93, f"Median\n{median:.1f}%", color=palette[3], fontsize=8.0, va="top")
    axes[1].set(xlim=(0, 100), xlabel="Starting entropy removed (%)", ylabel="Operating points")
    panel_label(axes[1], "B", "Complete reference distribution", "empirical")
    axes[1].grid(False)

    axes[2].scatter(
        twenty.information_bits,
        100 * twenty.post_negative_probability,
        s=14,
        marker="o",
        color="#727B83" if not grayscale else "#777777",
        alpha=0.48,
        edgecolors="none",
    )
    legend_handles: list[Line2D] = []
    legend_labels = {
        "Non-contrast CT appendicitis": "Appendicitis CT",
        "Ottawa ankle rule": "Ottawa ankle rule",
        "Lung ultrasound pneumonia": "Lung US pneumonia",
        "CT angiography intracranial lesion": "CTA intracranial lesion",
    }
    for row, color, marker in zip(landmarks.itertuples(index=False), palette, QUESTION_MARKERS):
        axes[2].scatter(row.information_bits, 100 * row.post_negative_probability, s=48, marker=marker, color=color, edgecolor=WHITE, linewidth=0.6, zorder=4)
        short_label = legend_labels[str(row.label)]
        legend_handles.append(Line2D([0], [0], marker=marker, color="none", markerfacecolor=color, markeredgecolor=color, label=f"{short_label} (n={int(row.studies)})"))
    axes[2].set(xlim=(0, max(0.72, twenty.information_bits.max() * 1.03)), ylim=(0, max(22, 100 * twenty.post_negative_probability.max() * 1.06)), xlabel="Expected information (bits)", ylabel="Post-negative probability (%)")
    panel_label(axes[2], "C", "Expectation versus a negative branch", "empirical")
    grid_both(axes[2])
    fig.legend(handles=legend_handles, frameon=False, fontsize=8.0, loc="lower center", ncol=2, bbox_to_anchor=(0.70, 0.015), handletextpad=0.35, columnspacing=0.8, labelspacing=0.4)
    fig.suptitle("Signature cross-diagnosis Atlas: 273 operating points from 210 reviews", y=0.985)
    fig.subplots_adjust(left=0.075, right=0.985, bottom=0.27, top=0.77, wspace=0.40)
    if colorbar_contours is not None:
        panel_position = axes[0].get_position()
        colorbar_axis = fig.add_axes(
            [panel_position.x0 + 0.05 * panel_position.width, 0.075, 0.90 * panel_position.width, 0.025]
        )
        colorbar = fig.colorbar(colorbar_contours, cax=colorbar_axis, orientation="horizontal")
        colorbar.set_ticks((0.00, 0.24, 0.48, 0.72))
        colorbar.ax.tick_params(labelsize=8.0, pad=1.0)
        colorbar.ax.set_title("Bits", fontsize=8.0, pad=1.0)
    return fig


def figure_3(examples: pd.DataFrame, profiles: pd.DataFrame, summary: dict[str, object], grayscale: bool) -> plt.Figure:
    fig, axes = new_figure("Figure 3", grayscale, nrows=1, ncols=3)
    palette = colors(grayscale)
    equal_j = examples.loc[examples.example_id.eq("equal_youden_different_information")].sort_values("test_label")
    y = np.arange(len(equal_j))
    axes[0].hlines(y, 0, equal_j.information_bits, color=palette[1], lw=4.5, alpha=0.32)
    for row, ypos, marker in zip(equal_j.itertuples(index=False), y, ("o", "s")):
        axes[0].scatter(row.information_bits, ypos, marker=marker, s=46, color=palette[1], zorder=3)
        axes[0].text(row.information_bits + 0.006, ypos, f"{row.information_bits:.3f} bits", va="center", fontsize=8.0)
    axes[0].set(
        yticks=y,
        yticklabels=[
            f"Test {r.test_label}\n{pct(r.sensitivity, 0)} / {pct(r.specificity, 0)}"
            for r in equal_j.itertuples(index=False)
        ],
        xlabel="Expected information at 20% (bits)",
        xlim=(0, max(equal_j.information_bits) * 1.35),
    )
    axes[0].text(0.02, 0.08, f"Both J = {equal_j.youden_j.iloc[0]:.2f}\nΔ information = {float(summary['equal_youden_different_information']['absolute_information_difference_bits']):.3f} bits", transform=axes[0].transAxes, fontsize=8.0, color=MUTED)
    panel_label(axes[0], "A", "Equal J, different expectation", "theoretical")
    grid_y(axes[0])

    near = examples.loc[examples.example_id.eq("near_equal_information_different_post_negative")].sort_values("test_label")
    for xpos, row, marker, color in zip((0, 1), near.itertuples(index=False), ("o", "s"), (palette[1], palette[2])):
        axes[1].scatter(xpos, 100 * row.post_negative_probability, marker=marker, s=58, color=color)
        axes[1].annotate(f"{100 * row.post_negative_probability:.1f}%\n{row.information_bits:.6f} bits", (xpos, 100 * row.post_negative_probability), xytext=(5, 4), textcoords="offset points", fontsize=8.0)
    axes[1].set(
        xticks=[0, 1],
        xticklabels=[
            f"Test {row.test_label}\n{pct(row.sensitivity, 0)} / {pct(row.specificity, 0)}"
            for row in near.itertuples(index=False)
        ],
        xlim=(-0.45, 1.45),
        ylim=(0, max(14, 100 * near.post_negative_probability.max() * 1.18)),
        xlabel="Theoretical operating point",
        ylabel="Post-negative probability (%)",
    )
    delta_info = float(summary["near_equal_information_different_post_negative"]["absolute_information_difference_bits"])
    delta_post = float(summary["near_equal_information_different_post_negative"]["absolute_post_negative_difference"])
    axes[1].text(0.03, 0.56, f"Δ information {delta_info:.5f} bits\nΔ post-negative {100 * delta_post:.1f} points", transform=axes[1].transAxes, va="top", fontsize=8.0, color=MUTED)
    panel_label(axes[1], "B", "Near-equal expectation, different result", "theoretical")
    grid_both(axes[1])

    crossing = float(summary["probability_dependent_information_crossing"]["numerical_root"])
    for row, (label, frame), color, linestyle, marker in zip(
        range(2), profiles.groupby("test_label", sort=True), palette[0:2], QUESTION_LINESTYLES[0:2], ("o", "s")
    ):
        axes[2].plot(100 * frame.starting_probability, frame.information_bits, color=color, ls=linestyle, lw=1.8, marker=marker, markevery=10, ms=3.5, label=f"{label}: Se {pct(frame.sensitivity.iloc[0], 0)}, Sp {pct(frame.specificity.iloc[0], 0)}")
    axes[2].axvline(100 * crossing, color=palette[3], ls=QUESTION_LINESTYLES[3], lw=1.3)
    axes[2].text(100 * crossing + 1.0, axes[2].get_ylim()[1] * 0.13, f"Crossing\n{100 * crossing:.1f}%", fontsize=8.0, color=palette[3])
    axes[2].set(xlim=(0, 50), ylim=(0, None), xlabel="Starting probability (%)", ylabel="Expected information (bits)")
    axes[2].legend(frameon=False, fontsize=8.0, loc="upper left")
    panel_label(axes[2], "C", "Probability-dependent ordering", "theoretical")
    grid_both(axes[2])
    fig.suptitle("Three theoretical demonstrations of non-equivalence", y=0.985)
    figure_footer(fig, "Display probabilities are not action thresholds.")
    fig.subplots_adjust(left=0.14, right=0.985, bottom=0.20, top=0.76, wspace=0.46)
    return fig


def figure_4(branches: pd.DataFrame, summary: pd.DataFrame, grayscale: bool) -> plt.Figure:
    fig, axes = new_figure("Figure 4", grayscale, nrows=1, ncols=3, sharey=True)
    palette = colors(grayscale)
    for ax, prior in zip(axes, sorted(branches.starting_probability.unique())):
        frame = branches.loc[np.isclose(branches.starting_probability, prior)]
        values = [frame.positive_entropy_change_bits.to_numpy(), frame.negative_entropy_change_bits.to_numpy()]
        parts = ax.violinplot(values, positions=[1, 2], showmeans=False, showmedians=True, widths=0.72)
        for body, color, hatch in zip(parts["bodies"], (palette[2], palette[0]), (HATCHES[2], HATCHES[0])):
            body.set_facecolor(color)
            body.set_edgecolor(color)
            body.set_alpha(0.35 if not grayscale else 0.30)
            body.set_hatch(hatch)
        for key in ("cmedians", "cmins", "cmaxes", "cbars"):
            if key in parts:
                parts[key].set_color("#30363B")
                parts[key].set_linewidth(0.9)
        jitter = np.linspace(-0.11, 0.11, len(frame))
        ax.scatter(1 + jitter, values[0], s=4.5, marker="^", color=palette[2], alpha=0.28, edgecolors="none")
        ax.scatter(2 - jitter, values[1], s=4.5, marker="o", color=palette[0], alpha=0.28, edgecolors="none")
        ax.axhline(0, color="#30363B", ls="--", lw=0.9)
        pos_inc = int(((summary.starting_probability.eq(prior)) & summary.result_branch.eq("positive_branch_entropy_movement") & summary.entropy_movement.eq("increase")) .mul(summary.operating_points).sum())
        neg_inc = int(((summary.starting_probability.eq(prior)) & summary.result_branch.eq("negative_branch_entropy_movement") & summary.entropy_movement.eq("increase")) .mul(summary.operating_points).sum())
        ax.text(0.04, 0.96, f"Entropy increase\npositive {pos_inc}/{len(frame)}\nnegative {neg_inc}/{len(frame)}", transform=ax.transAxes, va="top", fontsize=8.0, color=MUTED)
        ax.set(xticks=[1, 2], xticklabels=["Positive result\n▲", "Negative result\n●"], xlabel="Observed result branch")
        panel_label(ax, chr(65 + list(sorted(branches.starting_probability.unique())).index(prior)), f"Starting probability {pct(prior, 0)}", "empirical")
        grid_y(ax)
    axes[0].set_ylabel("Entropy change (bits)")
    fig.suptitle("Observed branches can move entropy in opposite directions", y=0.985)
    figure_footer(fig, "All panels show n=273 primary operating points. Movement is not clinical benefit or harm; expected mutual information remains non-negative.", width=108)
    fig.subplots_adjust(left=0.09, right=0.985, bottom=0.20, top=0.76, wspace=0.20)
    return fig


def figure_s1(run_manifest: dict[str, object], trace: dict[str, object], exceptions: pd.DataFrame, grayscale: bool) -> plt.Figure:
    fig, ax = new_figure("Figure S1", grayscale)
    palette = colors(grayscale)
    fixed = run_manifest["fixed_reference_sample"]
    ax.set_xlim(0, 1)
    ax.set_ylim(0, 1)
    ax.axis("off")
    fig.subplots_adjust(left=0.03, right=0.97, top=0.94, bottom=0.04)
    ax.text(0.02, 0.98, "Corpus and deterministic row selection", fontsize=10.0, weight="bold", va="top")
    ax.text(0.02, 0.935, "The fixed reference sample is selected before display construction", fontsize=8.0, color=MUTED, va="top")

    nodes = (
        (0.08, 0.805, 0.66, 0.090, f"{fixed['primary_operating_points']} primary operating points", f"{fixed['reviews']} reviews · prespecified source selection", palette[0], HATCHES[0]),
        (0.08, 0.655, 0.66, 0.090, f"{fixed['raw_normalized_rows_joined']} raw joined study rows", "Every selected point has normalised rows", palette[1], HATCHES[1]),
        (0.08, 0.505, 0.66, 0.090, f"{fixed['excluded_noncanonical_sheet_rows']} rows from another source sheet excluded", f"{fixed['row_selection_exceptions']} documented Zenodo exception", palette[3], HATCHES[3]),
        (0.08, 0.355, 0.66, 0.090, f"{fixed['analysis_rows_after_manifest_sheet_selection']} primary-analysis study rows", f"Exact recorded row counts for {fixed['primary_points_with_exact_manifest_analysis_rows']}/{fixed['primary_operating_points']} points", palette[1], HATCHES[1]),
        (0.08, 0.205, 0.66, 0.090, f"{trace['operating_points_with_source_identity']} traceable operating points", f"{trace['operating_points_with_group_url']} group URLs · {trace['operating_points_without_group_url']} archive identities", palette[2], HATCHES[2]),
    )
    for index, (x, y, w, h, title, body, color, hatch) in enumerate(nodes):
        ax.add_patch(FancyBboxPatch((x, y), w, h, boxstyle="round,pad=0.012,rounding_size=0.012", fc=WHITE, ec=color, lw=1.3))
        ax.text(x + 0.02, y + h * 0.66, title, fontsize=8.7, weight="bold", color=color, va="center")
        ax.text(x + 0.02, y + h * 0.28, body, fontsize=8.0, color=INK, va="center")
        if index < len(nodes) - 1:
            ax.add_patch(FancyArrowPatch((0.41, y), (0.41, y - 0.055), arrowstyle="-|>", mutation_scale=10, color=MUTED, lw=1.0))

    route_items = list(fixed["source_routes"].items())
    ax.text(0.785, 0.875, "Source routes", fontsize=8.0, weight="bold", color=MUTED)
    route_markers = ("o", "s", "^", "D", "P")
    for index, (route, count) in enumerate(route_items):
        y = 0.81 - 0.078 * index
        ax.scatter(
            0.79,
            y,
            marker=route_markers[index],
            s=34,
            color=palette[index % len(palette)],
            edgecolor=INK,
            linewidth=0.45,
        )
        ax.text(0.82, y, f"{route}\n{count} points", fontsize=8.0, va="center")
    ax.text(0.08, 0.115, "The fixed sample preserves source-defined groups and reviews.", fontsize=8.0, weight="bold", color=MUTED)
    ax.text(0.08, 0.074, fill(f"The single exception retains {int(exceptions.iloc[0]['analysis_row_count'])} recorded selected rows and excludes {int(exceptions.iloc[0]['excluded_noncanonical_rows'])} rows from another sheet.", width=90), fontsize=8.0, color=MUTED)
    return fig


def figure_s2(fidelity: pd.DataFrame, audit: dict[str, object], routes: pd.DataFrame, grayscale: bool) -> plt.Figure:
    fig, axes = new_figure("Figure S2", grayscale, nrows=1, ncols=3, gridspec_kw={"width_ratios": [0.86, 1.48, 0.96]})
    palette = colors(grayscale)
    axes[0].scatter(fidelity.published_sensitivity, fidelity.sensitivity, s=18, marker="o", facecolors="none", edgecolors=palette[0], linewidth=0.8, label="Sensitivity")
    axes[0].scatter(fidelity.published_specificity, fidelity.specificity, s=18, marker="s", color=palette[1], alpha=0.62, edgecolors="none", label="Specificity")
    axes[0].plot([0, 1], [0, 1], color=MUTED, ls="--", lw=0.9)
    large = fidelity.large_discrepancy.astype(str).str.lower().eq("true")
    axes[0].scatter(fidelity.loc[large, "published_sensitivity"], fidelity.loc[large, "sensitivity"], s=42, marker="x", color=palette[3], lw=1.2)
    axes[0].scatter(fidelity.loc[large, "published_specificity"], fidelity.loc[large, "specificity"], s=42, marker="x", color=palette[3], lw=1.2, label="Absolute difference ≥0.10")
    axes[0].set(xlim=(0, 1.01), ylim=(0, 1.01), xlabel="Source-reported pooled value", ylabel="Primary REML value")
    panel_label(axes[0], "A", f"Source-summary fidelity (n={len(fidelity)})", "validation")
    legend_handles, legend_labels = axes[0].get_legend_handles_labels()
    grid_both(axes[0])

    axes[1].set_xlim(0, 1)
    axes[1].set_ylim(0, 1)
    axes[1].axis("off")
    panel_label(axes[1], "B", "Recorded author-audit denominators", "validation")
    audit_boxes = (
        ("Transcription", f"{audit['transcription_error_numerator']} transcription errors/{audit['transcription_error_denominator_cells']} cells", f"{audit['transcription_error_denominator_rows']} rows · {audit['transcription_error_denominator_extracted_tables']} tables", palette[0], HATCHES[0]),
        ("Acceptance", f"{audit['acceptance_error_numerator']} acceptance errors/{audit['acceptance_error_denominator_decisions']} decisions", f"{audit['included_tables']} included · {audit['orientation_safeguard_tables']} safeguards · {audit['rejected_near_miss_tables']} rejected", palette[1], HATCHES[1]),
        ("Audit role", "Recorded author audit", "Not newly independent", palette[3], HATCHES[3]),
    )
    for index, (title, value, note, color, hatch) in enumerate(audit_boxes):
        y = 0.66 - index * 0.27
        height = 0.23
        axes[1].add_patch(FancyBboxPatch((0.025, y), 0.95, height, boxstyle="round,pad=0.012", fc=WHITE, ec=color, lw=1.1))
        axes[1].text(0.055, y + height - 0.048, title, fontsize=8.0, color=color, weight="bold", va="top")
        axes[1].text(0.055, y + height - 0.104, value, fontsize=8.0, color=INK, weight="bold", va="top")
        del note  # cohort composition is reported in the self-contained caption.

    twenty = routes.loc[np.isclose(routes.starting_probability, 0.20)].copy()
    route_order = [name for name in ("historical Cochrane", "modern Cochrane", "OSF", "Zenodo", "PMC") if name in set(twenty.cohort)]
    twenty["cohort"] = pd.Categorical(twenty.cohort, route_order, ordered=True)
    twenty = twenty.sort_values("cohort")
    y = np.arange(len(twenty))
    axes[2].hlines(y, twenty.information_bits_q1, twenty.information_bits_q3, color=palette[1], lw=3.0)
    axes[2].scatter(twenty.information_bits_median, y, marker="D", s=32, color=palette[1], zorder=3)
    for ypos, row in zip(y, twenty.itertuples(index=False)):
        axes[2].text(row.information_bits_q3 + 0.012, ypos, f"n={int(row.operating_points)}", va="center", fontsize=8.0)
    route_labels = {"historical Cochrane": "Hist. C.", "modern Cochrane": "Modern C."}
    axes[2].set(yticks=y, yticklabels=[route_labels.get(str(value), str(value)) for value in twenty.cohort], xlabel="Expected information at 20%\n(bits)", xlim=(0, max(0.65, twenty.information_bits_q3.max() * 1.25)))
    axes[2].invert_yaxis()
    panel_label(axes[2], "C", "Route medians and interquartile ranges", "descriptive")
    grid_both(axes[2])
    fig.suptitle("Source fidelity, audit denominators, and fixed-sample route variation", y=0.985)
    fig.legend(legend_handles, legend_labels, loc="lower center", ncol=3, bbox_to_anchor=(0.5, 0.025), fontsize=7.0)
    fig.subplots_adjust(left=0.075, right=0.965, bottom=0.20, top=0.76, wspace=0.34)
    return fig


def figure_s3(anchors: pd.DataFrame, summary: pd.DataFrame, grayscale: bool) -> plt.Figure:
    fig, axes = new_figure("Figure S3", grayscale, nrows=1, ncols=3)
    palette = colors(grayscale)
    priors = sorted(anchors.starting_probability.unique())
    labels = [pct(prior, 0) for prior in priors]
    information_values = [anchors.loc[np.isclose(anchors.starting_probability, prior), "information_bits"].to_numpy() for prior in priors]
    fraction_values = [100 * anchors.loc[np.isclose(anchors.starting_probability, prior), "fraction_entropy_removed"].to_numpy() for prior in priors]
    for ax, values, ylabel, panel in (
        (axes[0], information_values, "Expected information (bits)", ("A", "Complete information distributions")),
        (axes[2], fraction_values, "Starting entropy removed (%)", ("C", "Relative information distributions")),
    ):
        parts = ax.violinplot(values, positions=np.arange(3), showmedians=True, widths=0.72)
        for body, color, hatch in zip(parts["bodies"], palette[:3], HATCHES[:3]):
            body.set_facecolor(color)
            body.set_edgecolor(color)
            body.set_alpha(0.38)
            body.set_hatch(hatch)
        for key in ("cmedians", "cmins", "cmaxes", "cbars"):
            if key in parts:
                parts[key].set_color("#30363B")
        ax.set(xticks=np.arange(3), xticklabels=labels, xlabel="Starting probability", ylabel=ylabel)
        panel_label(ax, panel[0], panel[1], "empirical")
        grid_y(ax)

    for prior, color, marker, linestyle in zip(priors, palette[:3], QUESTION_MARKERS[:3], QUESTION_LINESTYLES[:3]):
        frame = anchors.loc[np.isclose(anchors.starting_probability, prior)]
        axes[1].scatter(frame.youden_j, frame.information_bits, s=10, marker=marker, color=color, alpha=0.32, edgecolors="none")
        coefficient = np.polyfit(frame.youden_j, frame.information_bits, deg=1)
        x = np.linspace(frame.youden_j.min(), frame.youden_j.max(), 100)
        axes[1].plot(x, coefficient[0] * x + coefficient[1], color=color, ls=linestyle, lw=1.3, label=f"{pct(prior, 0)}: ρ={summary.loc[np.isclose(summary.starting_probability, prior), 'spearman_information_vs_youden_j'].iloc[0]:.3f}")
    axes[1].set(xlabel="Youden’s J", ylabel="Expected information (bits)")
    axes[1].legend(frameon=False, fontsize=8.0, loc="upper left")
    panel_label(axes[1], "B", "Empirical concordance", "empirical")
    grid_both(axes[1])
    fig.suptitle("Expected-information distributions and concordance with Youden’s J", y=0.985)
    figure_footer(fig, "Each anchor contains all 273 primary operating points. Concordance does not imply mathematical equivalence.")
    fig.subplots_adjust(left=0.075, right=0.985, bottom=0.20, top=0.76, wspace=0.38)
    return fig


def figure_s4(recovery: pd.DataFrame, recovery_summary: dict[str, object], diagnostics: pd.DataFrame, diagnostic_summary: dict[str, object], grayscale: bool) -> plt.Figure:
    fig, axes = new_figure("Figure S4", grayscale, nrows=2, ncols=2)
    axes = np.asarray(axes).ravel()
    palette = colors(grayscale)
    for ax, truth, estimate, low, high, covered, label, color, marker in (
        (axes[0], "true_sensitivity", "estimated_sensitivity", "pooled_mean_sensitivity_low", "pooled_mean_sensitivity_high", "sensitivity_interval_includes_truth", "Sensitivity", palette[0], "o"),
        (axes[1], "true_specificity", "estimated_specificity", "pooled_mean_specificity_low", "pooled_mean_specificity_high", "specificity_interval_includes_truth", "Specificity", palette[1], "s"),
    ):
        coverage = recovery[covered].astype(str).str.lower().eq("true")
        lower_error = recovery[estimate] - recovery[low]
        upper_error = recovery[high] - recovery[estimate]
        ax.errorbar(recovery[truth], recovery[estimate], yerr=np.vstack([lower_error, upper_error]), fmt="none", ecolor="#A9B1B7", elinewidth=0.7, capsize=1.5, zorder=1)
        ax.scatter(recovery.loc[coverage, truth], recovery.loc[coverage, estimate], marker=marker, s=25, color=color, label="Includes generating value", zorder=2)
        ax.scatter(recovery.loc[~coverage, truth], recovery.loc[~coverage, estimate], marker="x", s=34, color=palette[3], lw=1.2, label="Does not include", zorder=3)
        low_limit = min(recovery[truth].min(), recovery[low].min()) - 0.02
        high_limit = max(recovery[truth].max(), recovery[high].max()) + 0.02
        ax.plot([low_limit, high_limit], [low_limit, high_limit], color=MUTED, ls="--", lw=0.9)
        ax.set(xlim=(low_limit, high_limit), ylim=(low_limit, high_limit), xlabel=f"Generating {label.lower()}", ylabel=f"Estimated {label.lower()}")
        panel_label(ax, "A" if label == "Sensitivity" else "B", f"{label}: {int(coverage.sum())}/{len(recovery)} intervals", "recovery")
        grid_both(ax)
    axes[1].legend(frameon=False, fontsize=8.0, loc="lower right")

    labels = ("|corr.|\n≥0.99", "near-\nsingular\ncovariance", "gradient\n>10⁻⁴")
    counts_values = (
        int(diagnostic_summary["absolute_between_correlation_at_least_0_99"]),
        int(diagnostic_summary["near_singular_between_covariance_min_eigenvalue_below_1e_8"]),
        int(diagnostic_summary["optimizer_gradient_above_1e_4"]),
    )
    x = np.arange(3)
    bars = axes[2].bar(x, counts_values, color=palette[:3], edgecolor=INK, linewidth=0.5)
    for bar, hatch in zip(bars, HATCHES[:3]):
        bar.set_hatch(hatch)
    for xpos, value in zip(x, counts_values):
        axes[2].text(xpos, value + max(counts_values) * 0.035, f"{value}/{len(diagnostics)}", ha="center", fontsize=8.0)
    axes[2].set(xticks=x, xticklabels=labels, ylabel="Flagged fits", ylim=(0, max(counts_values) * 1.2))
    axes[2].tick_params(axis="x", labelsize=8.0, pad=3.0)
    panel_label(axes[2], "C", f"Diagnostics (n={len(diagnostics)})", "diagnostic")
    grid_y(axes[2])

    hard_labels = ("tied L00\nat -8", "tied L11\nat -8", "tied\nunique", "selected\nunique", "all-flag\nunion")
    hard_counts = (
        int(diagnostic_summary["optimizer_tied_solution_log_l00_at_lower_bound"]),
        int(diagnostic_summary["optimizer_tied_solution_log_l11_at_lower_bound"]),
        int(diagnostic_summary["optimizer_any_tied_solution_cholesky_diagonal_at_lower_bound"]),
        int(diagnostic_summary["optimizer_any_selected_endpoint_cholesky_diagonal_at_lower_bound"]),
        int(diagnostic_summary["union_of_all_diagnostic_flags"]),
    )
    hard_x = np.arange(len(hard_counts))
    hard_bars = axes[3].bar(
        hard_x, hard_counts, color=[palette[index % len(palette)] for index in hard_x], edgecolor=INK, linewidth=0.5
    )
    for bar, hatch in zip(hard_bars, [HATCHES[index % len(HATCHES)] for index in hard_x]):
        bar.set_hatch(hatch)
    for xpos, value in zip(hard_x, hard_counts):
        axes[3].text(
            xpos,
            value + max(hard_counts) * 0.035,
            f"{value}/{len(diagnostics)}",
            ha="center",
            fontsize=8.0,
        )
    axes[3].set(
        xticks=hard_x,
        xticklabels=hard_labels,
        ylabel="Flagged fits",
        ylim=(0, max(hard_counts) * 1.2),
    )
    axes[3].tick_params(axis="x", labelsize=8.0, pad=3.0)
    panel_label(axes[3], "D", "Tied-solution and selected hard bounds", "diagnostic")
    grid_y(axes[3])
    fig.suptitle("Primary REML limited recovery and fit diagnostics", y=0.99)
    figure_footer(
        fig,
        f"Se = sensitivity; Sp = specificity. One dataset per {len(recovery)} scenarios: limited smoke diagnostic, not repeated-sampling coverage. Approximate plug-in intervals condition on fitted heterogeneity; predictive ranges were not checked.",
        y=0.012,
        width=118,
        fontsize=8.0,
    )
    fig.subplots_adjust(left=0.075, right=0.985, bottom=0.19, top=0.86, hspace=0.58, wspace=0.32)
    return fig


def shortened_model(value: str) -> str:
    replacements = {
        "v0.42.0 canonical REML": "Primary REML",
        "v0.39.0 historical REML": "Historical REML",
        "v0.40.0/v0.41.0 regularized GH15": "Regularized GH15",
        "regularized GH15 wider prior": "GH15 wider prior",
        "regularized GH9 primary prior": "Regularized GH9",
    }
    return replacements.get(value, value)


def figure_s5(all_rows: pd.DataFrame, sensitivity_summary: pd.DataFrame, estimator_effect: dict[str, object], grayscale: bool) -> plt.Figure:
    fig, axes = new_figure("Figure S5", grayscale, nrows=1, ncols=3)
    palette = colors(grayscale)
    order = ["v0.39.0 historical REML", "v0.40.0/v0.41.0 regularized GH15", "regularized GH15 wider prior", "regularized GH9 primary prior"]
    gh15 = sensitivity_summary.loc[
        sensitivity_summary.model_specification.eq(order[1])
        & np.isclose(sensitivity_summary.starting_probability, 0.20)
    ].iloc[0]
    comparison_labels = ("Like-for-like plug-in\ncentral estimates", "Analysis-family\ncentral summaries")
    medians = np.asarray([
        estimator_effect["median_absolute_like_for_like_plugin_information_bits_at_20_percent_difference"],
        gh15.median_absolute_information_bits_difference,
    ])
    maxima = np.asarray([
        estimator_effect["maximum_absolute_like_for_like_plugin_information_bits_at_20_percent_difference"],
        gh15.maximum_absolute_information_bits_difference,
    ])
    y = np.arange(2)
    axes[0].hlines(y, 0, maxima, color="#AAB2B8", lw=2.2)
    axes[0].scatter(medians, y, marker="D", s=35, color=palette[1], label="Median absolute difference")
    axes[0].scatter(maxima, y, marker="|", s=95, color=palette[3], lw=1.5, label="Maximum")
    axes[0].set(yticks=y, yticklabels=comparison_labels, xlabel="Absolute information difference at 20% (bits)", xlim=(0, max(0.24, maxima.max() * 1.10)))
    axes[0].invert_yaxis()
    legend_handles, legend_labels = axes[0].get_legend_handles_labels()
    panel_label(axes[0], "A", "Two central-summary comparisons", "sensitivity")
    grid_both(axes[0])

    twenty = all_rows.loc[np.isclose(all_rows.starting_probability, 0.20)].copy()
    canonical = twenty.loc[twenty.analysis_role.eq("canonical primary"), ["group_id", "information_bits"]].rename(columns={"information_bits": "canonical_information"})
    compared = twenty.loc[twenty.model_specification.isin(order[1:])].merge(canonical, on="group_id", validate="many_to_one")
    compared["absolute_information_difference"] = (compared.information_bits - compared.canonical_information).abs()
    for model, frame, color, marker in zip(order[1:], (compared.loc[compared.model_specification.eq(model)] for model in order[1:]), palette[:3], QUESTION_MARKERS[:3]):
        axes[1].scatter(frame.zero_cell_study_fraction, frame.absolute_information_difference, marker=marker, s=12, color=color, alpha=0.36, edgecolors="none", label=shortened_model(model))
    axes[1].set(xlim=(0, 1), ylim=(0, None), xlabel="Zero-cell study fraction", ylabel="Absolute information difference (bits)")
    axes[1].legend(frameon=False, fontsize=8.0, loc="upper left")
    panel_label(axes[1], "B", "Analysis-family movement by sparse-data burden", "sensitivity")
    grid_both(axes[1])

    for model, color, marker, linestyle in zip(order[1:], palette[:3], QUESTION_MARKERS[:3], QUESTION_LINESTYLES[:3]):
        frame = compared.loc[compared.model_specification.eq(model)]
        axes[2].scatter(frame.canonical_information, frame.information_bits, marker=marker, s=12, color=color, alpha=0.32, edgecolors="none", label=shortened_model(model))
    limit = max(compared.canonical_information.max(), compared.information_bits.max()) * 1.03
    axes[2].plot([0, limit], [0, limit], color=MUTED, ls="--", lw=0.9)
    axes[2].set(xlim=(0, limit), ylim=(0, limit), xlabel="Primary information (bits)", ylabel="Sensitivity-analysis information (bits)")
    family_handles, family_labels = axes[2].get_legend_handles_labels()
    panel_label(axes[2], "C", "Family comparison", "sensitivity")
    grid_both(axes[2])
    fig.suptitle("Like-for-like plug-in central estimates and analysis-family sensitivity", y=0.985)
    fig.legend(legend_handles, legend_labels, frameon=False, fontsize=8.0, loc="lower center", ncol=2, bbox_to_anchor=(0.31, 0.015))
    fig.legend(family_handles, family_labels, frameon=False, fontsize=8.0, loc="lower center", ncol=1, bbox_to_anchor=(0.82, 0.005))
    fig.subplots_adjust(left=0.17, right=0.985, bottom=0.24, top=0.76, wspace=0.52)
    return fig


def contrast_model_movement(contrasts: pd.DataFrame, sensitivity: pd.DataFrame) -> pd.DataFrame:
    base = contrasts.loc[np.isclose(contrasts.starting_probability, 0.20)].drop_duplicates("pair_id").copy()
    available_models = (
        "v0.42.0 canonical REML",
        "v0.40.0/v0.41.0 regularized GH15",
        "regularized GH9 primary prior",
    )
    model_rows = sensitivity.loc[np.isclose(sensitivity.starting_probability, 0.20) & sensitivity.model_specification.isin(available_models)]
    lookup = model_rows.set_index(["model_specification", "group_id"])
    records: list[dict[str, object]] = []
    for row in base.itertuples(index=False):
        for model in available_models:
            left = lookup.loc[(model, row.left_group_id)]
            right = lookup.loc[(model, row.right_group_id)]
            records.append(
                {
                    "pair_id": row.pair_id,
                    "comparison_type": row.comparison_type,
                    "baseline_tradeoff_set": row.baseline_tradeoff_set,
                    "model_specification": model,
                    "signed_information_bits_difference": float(left.information_bits - right.information_bits),
                    "signed_post_negative_probability_difference_percentage_points": float(100 * (left.post_negative_probability - right.post_negative_probability)),
                    "same_participants_established": row.same_participants_established,
                    "paired_inference_supported": row.paired_inference_supported,
                }
            )
    return pd.DataFrame.from_records(records)


def figure_s6(contrasts: pd.DataFrame, movement: pd.DataFrame, grayscale: bool) -> plt.Figure:
    fig, axes = new_figure("Figure S6", grayscale, nrows=1, ncols=2)
    palette = colors(grayscale)
    twenty = contrasts.loc[np.isclose(contrasts.starting_probability, 0.20)].drop_duplicates("pair_id")
    trade = twenty.baseline_tradeoff_set.astype(str).str.lower().eq("true")
    axes[0].axhline(0, color=MUTED, lw=0.8)
    axes[0].axvline(0, color=MUTED, lw=0.8)
    axes[0].scatter(twenty.loc[~trade, "signed_information_bits_difference"], twenty.loc[~trade, "signed_post_negative_probability_difference_percentage_points"], marker="o", s=30, facecolors="none", edgecolors=palette[0], label=f"Other classified contrasts (n={int((~trade).sum())})")
    axes[0].scatter(twenty.loc[trade, "signed_information_bits_difference"], twenty.loc[trade, "signed_post_negative_probability_difference_percentage_points"], marker="D", s=28, color=palette[3], alpha=0.76, label=f"Sensitivity–specificity trade-offs (n={int(trade.sum())})")
    axes[0].set(xlabel="Left − right expected information (bits)", ylabel="Left − right post-negative probability (points)")
    axes[0].legend(frameon=False, fontsize=8.0, loc="best")
    panel_label(axes[0], "A", "Continuous magnitudes at 20%", "exploratory")
    grid_both(axes[0])

    canonical_name = "v0.42.0 canonical REML"
    regularized_name = "v0.40.0/v0.41.0 regularized GH15"
    canonical = movement.loc[movement.model_specification.eq(canonical_name)].set_index("pair_id")
    regularized = movement.loc[movement.model_specification.eq(regularized_name)].set_index("pair_id")
    for pair_id in canonical.index:
        axes[1].plot(
            [canonical.loc[pair_id, "signed_information_bits_difference"], regularized.loc[pair_id, "signed_information_bits_difference"]],
            [canonical.loc[pair_id, "signed_post_negative_probability_difference_percentage_points"], regularized.loc[pair_id, "signed_post_negative_probability_difference_percentage_points"]],
            color="#B4BBC0",
            lw=0.65,
            alpha=0.72,
        )
    axes[1].scatter(canonical.signed_information_bits_difference, canonical.signed_post_negative_probability_difference_percentage_points, marker="o", s=24, facecolors="none", edgecolors=palette[0], label="Primary REML")
    axes[1].scatter(regularized.signed_information_bits_difference, regularized.signed_post_negative_probability_difference_percentage_points, marker="s", s=22, color=palette[1], alpha=0.70, label="Regularized GH15")
    axes[1].axhline(0, color=MUTED, lw=0.8)
    axes[1].axvline(0, color=MUTED, lw=0.8)
    axes[1].set(xlabel="Left − right expected information (bits)", ylabel="Left − right post-negative probability (points)")
    axes[1].legend(frameon=False, fontsize=8.0, loc="best")
    panel_label(axes[1], "B", "Analysis-family movement", "exploratory")
    grid_both(axes[1])
    fig.suptitle("Exploratory within-review contrasts: magnitude and analysis-family movement", y=0.985)
    figure_footer(fig, f"All {len(twenty)} contrasts are descriptive: same-participant cells and cross-test covariance are unavailable, so no paired inference is shown.")
    fig.subplots_adjust(left=0.09, right=0.985, bottom=0.20, top=0.76, wspace=0.32)
    return fig


def near_tie_and_composition_summary(contrasts: pd.DataFrame, anchors: pd.DataFrame) -> tuple[pd.DataFrame, pd.DataFrame]:
    del anchors  # row-level anchor source is retained in the display map and hash contract.
    twenty = contrasts.loc[np.isclose(contrasts.starting_probability, 0.20)].drop_duplicates("pair_id").copy()
    trade = twenty.loc[twenty.baseline_tradeoff_set.astype(str).str.lower().eq("true")]
    rows: list[dict[str, object]] = []
    for comparison_set, frame in (("All classified contrasts", twenty), ("Sensitivity–specificity trade-offs", trade)):
        for metric, column, thresholds, unit in (
            ("Expected information", "absolute_information_bits_difference", (0.001, 0.005, 0.010), "bits"),
            ("Fraction of starting entropy removed", "absolute_fraction_entropy_removed_difference_percentage_points", (1.0, 2.0, 5.0), "percentage points"),
        ):
            for threshold in thresholds:
                rows.append(
                    {
                        "comparison_set": comparison_set,
                        "metric": metric,
                        "threshold": threshold,
                        "unit": unit,
                        "near_tie_count": int((frame[column] < threshold).sum()),
                        "comparisons": int(len(frame)),
                        "threshold_role": "descriptive tolerance; not a clinical equivalence margin",
                    }
                )
    restricted = twenty.loc[twenty.overlapping_study_identifier_count.astype(int).ge(3)].copy()
    restricted["restriction"] = "at least three overlapping study identifiers; does not establish same participants"
    return pd.DataFrame.from_records(rows), restricted


def figure_s7(near_ties: pd.DataFrame, restricted: pd.DataFrame, grayscale: bool) -> plt.Figure:
    fig, axes = new_figure("Figure S7", grayscale, nrows=1, ncols=2)
    palette = colors(grayscale)
    info = near_ties.loc[near_ties.metric.eq("Expected information")]
    all_rows = info.loc[info.comparison_set.eq("All classified contrasts")]
    trade_rows = info.loc[info.comparison_set.eq("Sensitivity–specificity trade-offs")]
    x = np.arange(3)
    width = 0.34
    bars_a = axes[0].bar(x - width / 2, all_rows.near_tie_count, width, color=palette[0], label=f"All (n={all_rows.comparisons.iloc[0]})")
    bars_b = axes[0].bar(x + width / 2, trade_rows.near_tie_count, width, color=palette[3], label=f"Trade-offs (n={trade_rows.comparisons.iloc[0]})")
    for bars, hatch in ((bars_a, HATCHES[0]), (bars_b, HATCHES[3])):
        for bar in bars:
            bar.set_hatch(hatch)
            axes[0].text(bar.get_x() + bar.get_width() / 2, bar.get_height() + 0.15, str(int(bar.get_height())), ha="center", fontsize=8.0)
    axes[0].set(xticks=x, xticklabels=[f"<{value:.3f}" for value in all_rows.threshold], xlabel="Absolute information difference at 20% (bits)", ylabel="Contrasts below tolerance", ylim=(0, max(3, max(all_rows.near_tie_count.max(), trade_rows.near_tie_count.max()) * 1.35)))
    axes[0].legend(frameon=False, fontsize=8.0)
    panel_label(axes[0], "A", "Descriptive near-tie tolerances", "audit")
    grid_y(axes[0])

    axes[1].axhline(0, color=MUTED, lw=0.8)
    axes[1].axvline(0, color=MUTED, lw=0.8)
    axes[1].scatter(restricted.signed_information_bits_difference, restricted.signed_post_negative_probability_difference_percentage_points, marker="s", s=34, color=palette[1], alpha=0.72)
    label_ids = set(
        restricted.assign(
            display_magnitude=np.hypot(
                restricted.signed_information_bits_difference,
                restricted.signed_post_negative_probability_difference_percentage_points / 100,
            )
        )
        .nlargest(min(6, len(restricted)), "display_magnitude")
        .pair_id
    )
    special_offsets = {"P24": (-34, 4), "P48": (6, -3), "P49": (-34, 10)}
    for row in restricted.itertuples(index=False):
        if row.pair_id in label_ids:
            if row.pair_id in special_offsets:
                offset = special_offsets[str(row.pair_id)]
            elif row.signed_information_bits_difference > 0.14:
                offset = (-30, 12 if row.signed_post_negative_probability_difference_percentage_points < -7.5 else 4)
            elif row.signed_post_negative_probability_difference_percentage_points < -7.5:
                offset = (3, -10)
            else:
                offset = (3, 3)
            arrow = {"arrowstyle": "-", "color": MUTED, "lw": 0.55, "shrinkA": 1.5, "shrinkB": 2.5}
            axes[1].annotate(
                str(row.pair_id),
                (row.signed_information_bits_difference, row.signed_post_negative_probability_difference_percentage_points),
                xytext=offset,
                textcoords="offset points",
                fontsize=8.0,
                arrowprops=arrow,
            )
    axes[1].margins(x=0.12, y=0.18)
    axes[1].set(xlabel="Left − right expected information (bits)", ylabel="Left − right post-negative probability (points)")
    panel_label(axes[1], "B", f"Study-composition restriction (n={len(restricted)})", "audit")
    grid_both(axes[1])
    fig.suptitle("Near ties and study-composition-restricted audits", y=0.985)
    figure_footer(fig, "Near-tie thresholds are descriptive tolerances, not clinical equivalence margins. Overlapping study identifiers do not establish common participants or paired outcomes.")
    fig.subplots_adjust(left=0.09, right=0.985, bottom=0.20, top=0.76, wspace=0.32)
    return fig


def figure_s8(crossings: pd.DataFrame, profiles: pd.DataFrame, theory: dict[str, object], grayscale: bool) -> plt.Figure:
    fig, axes = new_figure("Figure S8", grayscale, nrows=1, ncols=3, gridspec_kw={"width_ratios": [1.32, 0.78, 0.90]})
    palette = colors(grayscale)
    selected_models = ("v0.42.0 canonical REML", "v0.40.0/v0.41.0 regularized GH15")
    for model, linestyle, marker in zip(selected_models, ("-", "--"), ("o", "s")):
        model_frame = profiles.loc[profiles.model_specification.eq(model)]
        for test_index, (test, frame) in enumerate(model_frame.groupby("test_name", sort=True)):
            model_label = "REML" if model == selected_models[0] else "Reg GH15"
            test_label = "CEA 2.5" if "2.5" in test else "CEA 5"
            axes[0].plot(100 * frame.starting_probability, frame.information_bits, color=palette[test_index], ls=linestyle, lw=1.55, marker=marker, markevery=12, ms=3.0, label=f"{model_label} · {test_label}")
        crossing = crossings.loc[crossings.model_specification.eq(model), "crossing_probability"].iloc[0]
        axes[0].axvline(100 * crossing, color=palette[3], ls=linestyle, lw=1.0)
        axes[0].text(100 * crossing + 0.5, 0.012 if model == selected_models[0] else 0.060, f"{100 * crossing:.1f}%", color=palette[3], fontsize=8.0)
    axes[0].set(xlim=(0, 50), ylim=(0, None), xlabel="Starting probability (%)", ylabel="Expected information (bits)")
    axes[0].legend(frameon=False, fontsize=8.0, loc="upper left")
    panel_label(axes[0], "A", "Empirical CEA model sensitivity", "empirical sensitivity")
    grid_both(axes[0])

    with_root = crossings.loc[crossings.crossing_probability.notna()].copy()
    y = np.arange(len(with_root))
    axes[1].scatter(100 * with_root.crossing_probability, y, marker="D", s=36, color=palette[2])
    axes[1].set(yticks=y, yticklabels=[shortened_model(v) for v in with_root.model_specification], xlabel="Crossing probability (%)", xlim=(0, 50))
    axes[1].invert_yaxis()
    panel_label(axes[1], "B", "Crossings by model", "sensitivity")
    grid_both(axes[1])

    axes[2].set_xlim(0, 1)
    axes[2].set_ylim(0, 1)
    axes[2].axis("off")
    panel_label(axes[2], "C", "Theory checks", "theoretical")
    checks = (
        ("Equal J", f"ΔJ = {float(theory['equal_youden_different_information']['absolute_youden_difference']):.1g}\nΔ I = {float(theory['equal_youden_different_information']['absolute_information_difference_bits']):.5f} bits"),
        ("Near-equal information", f"Δ I = {float(theory['near_equal_information_different_post_negative']['absolute_information_difference_bits']):.6f} bits\nΔ post-neg. = {100 * float(theory['near_equal_information_different_post_negative']['absolute_post_negative_difference']):.1f} points"),
        ("Synthetic crossing", f"p = {100 * float(theory['probability_dependent_information_crossing']['numerical_root']):.1f}%\nresidual = {float(theory['probability_dependent_information_crossing']['root_residual_bits']):.2g} bits"),
    )
    for index, (title, body) in enumerate(checks):
        y0 = 0.70 - index * 0.27
        axes[2].add_patch(FancyBboxPatch((0.04, y0), 0.92, 0.19, boxstyle="round,pad=0.01", fc=WHITE, ec=palette[index], lw=1.0))
        axes[2].text(0.07, y0 + 0.142, title, color=palette[index], fontsize=8.0, weight="bold")
        axes[2].text(0.07, y0 + 0.075, body, color=INK, fontsize=8.0, va="center")
    fig.suptitle("Empirical CEA crossing sensitivity and additional theoretical checks", y=0.985)
    figure_footer(fig, "The CEA crossings are inherited model-specific examples. None of the displayed probabilities is an action threshold.")
    fig.subplots_adjust(left=0.08, right=0.985, bottom=0.20, top=0.76, wspace=0.46)
    return fig


def build_table_1(source: pd.DataFrame) -> pd.DataFrame:
    frame = source.copy()
    frame.columns = (
        "Question",
        "Quantity",
        "When known",
        "Required uncertainty or context",
        "Common interpretation error",
    )
    return frame


def build_table_2(profile: pd.DataFrame) -> pd.DataFrame:
    lookup = profile_lookup(profile)
    rows: list[dict[str, str]] = []

    def add(section: str, quantity: str, estimate: str, pooled: str = "Not available", predictive: str = "Not available", context: str = "") -> None:
        rows.append(
            {
                "Profile section": section,
                "Quantity": quantity,
                "Central estimate or status": estimate,
                "Model-based 95% pooled-mean interval": pooled,
                "Model-based 95% predictive range": predictive,
                "Context": context,
            }
        )

    for element, quantity in (
        ("Index test context from review title", "Index-test context"),
        ("Locked operating-point test_name", "Recorded operating-point test_name"),
        ("Target condition and setting", "Target condition and setting"),
        ("Reference standard", "Reference standard"),
        ("Index-test positivity or operating threshold", "Index-test positivity or operating threshold"),
    ):
        row = lookup.loc[element]
        value = str(row.value_text) if pd.notna(row.value_text) else "Not available"
        add("Definition", quantity, value, context=str(row.availability_status))

    studies = lookup.loc["Study count"]
    add("Evidence", "Study count", f"n={int(float(studies.value))} studies", context=str(studies.availability_status))

    metric_specs = (
        ("Operating point", "Sensitivity", "Sensitivity", "Sensitivity predictive", "probability"),
        ("Operating point", "Specificity", "Specificity", "Specificity predictive", "probability"),
        ("Operating point", "Positive likelihood ratio", "Positive likelihood ratio", "Positive likelihood ratio predictive", "ratio"),
        ("Operating point", "Negative likelihood ratio", "Negative likelihood ratio", "Negative likelihood ratio predictive", "ratio"),
        ("Expected information", "Expected information", "Expected information", "Expected information predictive", "bits"),
        ("Expected information", "Fraction of starting entropy removed", "Fraction of starting entropy removed", "Fraction of starting entropy removed predictive", "fraction"),
        ("Observed-result probability", "Probability of a positive result", "Probability of a positive result", "Probability of a positive result predictive", "probability"),
        ("Observed-result probability", "Probability after a positive result", "Probability after a positive result", "Probability after a positive result predictive", "probability"),
        ("Observed-result probability", "Probability of a negative result", "Probability of a negative result", "Probability of a negative result predictive", "probability"),
        ("Observed-result probability", "Probability after a negative result", "Probability after a negative result", "Probability after a negative result predictive", "probability"),
    )
    for section, quantity, pooled_element, predictive_element, unit in metric_specs:
        pooled_row = lookup.loc[pooled_element]
        predictive_row = lookup.loc[predictive_element]
        if unit in {"probability", "fraction"}:
            estimate = pct(float(pooled_row.value))
            pooled_text = interval(float(pooled_row.low), float(pooled_row.high), percent=True)
            predictive_text = interval(float(predictive_row.low), float(predictive_row.high), percent=True)
        elif unit == "bits":
            estimate = f"{float(pooled_row.value):.4f} bits"
            pooled_text = f"{float(pooled_row.low):.4f}–{float(pooled_row.high):.4f} bits"
            predictive_text = f"{float(predictive_row.low):.4f}–{float(predictive_row.high):.4f} bits"
        elif "Positive" in quantity:
            estimate = f"{float(pooled_row.value):.4f}"
            pooled_text = f"{float(pooled_row.low):.4f}–{float(pooled_row.high):.4f}"
            predictive_text = f"{float(predictive_row.low):.4f}–{float(predictive_row.high):.4f}"
        else:
            estimate = f"{float(pooled_row.value):.4f}"
            pooled_text = f"{float(pooled_row.low):.4f}–{float(pooled_row.high):.4f}"
            predictive_text = f"{float(predictive_row.low):.4f}–{float(predictive_row.high):.4f}"
        add(section, quantity, estimate, pooled_text, predictive_text, str(pooled_row.availability_status))

    starting = lookup.loc["Illustrative starting probability"]
    add(
        "Starting probability",
        "Illustrative starting probability",
        pct(float(starting.value), 0),
        context=str(starting.availability_status),
    )
    action = lookup.loc["Clinical action"]
    add("Action", "Clinical action", "Not determined", context=str(action.availability_status))
    return pd.DataFrame.from_records(rows)


def markdown_table(frame: pd.DataFrame) -> str:
    def clean(value: object) -> str:
        return str(value).replace("|", "\\|").replace("\n", " ")

    header = "| " + " | ".join(clean(value) for value in frame.columns) + " |"
    rule = "| " + " | ".join("---" for _ in frame.columns) + " |"
    rows = ["| " + " | ".join(clean(value) for value in row) + " |" for row in frame.itertuples(index=False, name=None)]
    return "\n".join([header, rule, *rows])


def normalize_docx_archive(path: Path) -> None:
    temporary = path.with_suffix(".normalized.docx")
    with zipfile.ZipFile(path, "r") as source, zipfile.ZipFile(
        temporary, "w", compression=zipfile.ZIP_DEFLATED, compresslevel=9
    ) as target:
        for name in sorted(source.namelist()):
            info = zipfile.ZipInfo(name, date_time=(2026, 8, 25, 0, 0, 0))
            info.compress_type = zipfile.ZIP_DEFLATED
            info.external_attr = 0o600 << 16
            target.writestr(info, source.read(name))
    temporary.replace(path)


def write_docx_table(path: Path, title: str, frame: pd.DataFrame, footnote: str) -> None:
    document = Document()
    section = document.sections[0]
    section.top_margin = Inches(0.55)
    section.bottom_margin = Inches(0.55)
    section.left_margin = Inches(0.55)
    section.right_margin = Inches(0.55)
    paragraph = document.add_paragraph()
    paragraph.alignment = WD_ALIGN_PARAGRAPH.LEFT
    run = paragraph.add_run(title)
    run.bold = True
    run.font.size = Pt(10)
    table = document.add_table(rows=1, cols=len(frame.columns))
    table.style = "Table Grid"
    header_properties = table.rows[0]._tr.get_or_add_trPr()
    repeat_header = OxmlElement("w:tblHeader")
    repeat_header.set(qn("w:val"), "true")
    header_properties.append(repeat_header)
    for cell, column in zip(table.rows[0].cells, frame.columns):
        cell.text = str(column)
        cell.vertical_alignment = WD_CELL_VERTICAL_ALIGNMENT.CENTER
        for run in cell.paragraphs[0].runs:
            run.bold = True
            run.font.size = Pt(8)
    for values in frame.itertuples(index=False, name=None):
        cells = table.add_row().cells
        for cell, value in zip(cells, values):
            cell.text = str(value)
            cell.vertical_alignment = WD_CELL_VERTICAL_ALIGNMENT.TOP
            for paragraph in cell.paragraphs:
                for run in paragraph.runs:
                    run.font.size = Pt(8)
    for row in table.rows:
        row_properties = row._tr.get_or_add_trPr()
        prevent_split = OxmlElement("w:cantSplit")
        prevent_split.set(qn("w:val"), "true")
        row_properties.append(prevent_split)
    note = document.add_paragraph()
    note_run = note.add_run(footnote)
    note_run.italic = True
    note_run.font.size = Pt(8)
    document.core_properties.title = title.split(". ", 1)[0] + "."
    document.core_properties.author = f"BMJ RMR v{VERSION} deterministic display builder"
    document.core_properties.created = SOURCE_DATE
    document.core_properties.modified = SOURCE_DATE
    path.parent.mkdir(parents=True, exist_ok=True)
    document.save(path)
    normalize_docx_archive(path)


def build_captions(data: dict[str, object], table_2: pd.DataFrame) -> dict[str, str]:
    profile = data["profile"]
    anchors = data["anchors"]
    landmarks = data["landmarks"]
    theory = data["theory_summary"]
    branch_summary = data["branch_summary"]
    run_manifest = data["run_manifest"]
    trace = data["trace_summary"]
    fidelity = data["fidelity"]
    audit = data["audit_summary"]
    routes = data["routes"]
    anchor_summary = data["anchor_summary"]
    recovery_summary = data["recovery_summary"]
    diagnostic_summary = data["diagnostic_summary"]
    sensitivity_summary = data["sensitivity_summary"]
    estimator_effect = data["estimator_effect"]
    contrasts_summary = data["contrasts_summary"]
    near_ties = data["near_ties"]
    restricted = data["restricted"]
    crossings = data["crossings"]
    lookup = profile_lookup(profile)
    fixed = run_manifest["fixed_reference_sample"]
    se = lookup.loc["Sensitivity"]
    se_pr = lookup.loc["Sensitivity predictive"]
    sp = lookup.loc["Specificity"]
    sp_pr = lookup.loc["Specificity predictive"]
    info = lookup.loc["Expected information"]
    info_pr = lookup.loc["Expected information predictive"]
    p0 = float(lookup.loc["Illustrative starting probability", "value"])
    source_medians = (float(fidelity.absolute_sensitivity_difference.median()), float(fidelity.absolute_specificity_difference.median()))
    source_maxima = (float(fidelity.absolute_sensitivity_difference.max()), float(fidelity.absolute_specificity_difference.max()))
    twenty_summary = anchor_summary.loc[np.isclose(anchor_summary.starting_probability, 0.20)].iloc[0]
    branch_counts = {
        (float(row.starting_probability), row.result_branch, row.entropy_movement): int(row.operating_points)
        for row in branch_summary.itertuples(index=False)
    }
    canonical_crossing = float(crossings.loc[crossings.model_specification.eq("v0.42.0 canonical REML"), "crossing_probability"].iloc[0])
    regularized_crossing = float(crossings.loc[crossings.model_specification.eq("v0.40.0/v0.41.0 regularized GH15"), "crossing_probability"].iloc[0])
    gh15 = sensitivity_summary.loc[sensitivity_summary.model_specification.eq("v0.40.0/v0.41.0 regularized GH15") & np.isclose(sensitivity_summary.starting_probability, 0.20)].iloc[0]
    info_tie = near_ties.loc[near_ties.metric.eq("Expected information") & near_ties.comparison_set.eq("All classified contrasts")]
    captions = {
        "Figure 1": (
            f"Figure 1. The four-question Diagnostic Information Profile and an appendicitis worked example. Read from left to right, the profile identifies the test and evidence base, reports expected information before the result, reports probability after the observed result, and asks whether decision inputs support action. The example is a source-defined group with incomplete metadata: the review title identifies non-contrast CT, the recorded source label is ‘{lookup.loc['Locked operating-point test_name', 'value_text']}’, and no separate positivity threshold was recorded. Across n={int(float(lookup.loc['Study count', 'value']))} studies, the primary continuity-corrected bivariate model fitted by restricted maximum likelihood (REML) estimated sensitivity {pct(se.value)} (approximate model-based 95% pooled-mean interval {interval(se.low, se.high, percent=True)}; predictive range {interval(se_pr.low, se_pr.high, percent=True)}) and specificity {pct(sp.value)} (pooled-mean interval {interval(sp.low, sp.high, percent=True)}; predictive range {interval(sp_pr.low, sp_pr.high, percent=True)}). At the illustrative {pct(p0, 0)} starting probability, expected information was {info.value:.3f} bits (pooled-mean interval {info.low:.3f}–{info.high:.3f}; predictive range {info_pr.low:.3f}–{info_pr.high:.3f}). No action was selected because consequences, preferences, costs, and alternatives were absent. Intervals are plug-in summaries conditional on fitted heterogeneity; the tied appendicitis specificity-heterogeneity boundary solution makes joint predictive ranges especially fragile."
        ),
        "Figure 2": (
            f"Figure 2. Signature cross-diagnosis Atlas at the standardised {pct(p0, 0)} starting probability. (A) Expected-information contours with all n={fixed['primary_operating_points']} primary operating points from {fixed['reviews']} reviews. (B) Complete distribution of the fraction of starting entropy removed; the median was {100 * float(twenty_summary.fraction_entropy_removed_median):.1f}%. (C) Expected information versus post-negative probability for the same points. Four shape-coded landmarks ({'; '.join(f'{row.label}, n={int(row.studies)}' for row in landmarks.itertuples(index=False))}) orient scale and branch behaviour and are not rankings of clinical utility. Primary estimator: continuity-corrected bivariate REML."
        ),
        "Figure 3": (
            f"Figure 3. Three theoretical demonstrations of mathematical non-equivalence at stated starting probabilities. (A) Two operating points have identical Youden’s J (sensitivity + specificity − 1) but expected information differs by {float(theory['equal_youden_different_information']['absolute_information_difference_bits']):.6f} bits at 20%. (B) Two operating points have near-equal expected information (difference {float(theory['near_equal_information_different_post_negative']['absolute_information_difference_bits']):.6f} bits) but post-negative probabilities differ by {100 * float(theory['near_equal_information_different_post_negative']['absolute_post_negative_difference']):.1f} percentage points at 20%. (C) A synthetic pair changes expected-information ordering at starting probability {100 * float(theory['probability_dependent_information_crossing']['numerical_root']):.1f}%. These examples establish three mathematical distinctions; action remains separate because consequences, preferences, costs, and alternatives are absent."
        ),
        "Figure 4": (
            f"Figure 4. Observed-result branch entropy across n={fixed['primary_operating_points']} primary operating points at 5%, 20%, and 50% starting probabilities. Positive results increased binary entropy for {branch_counts.get((0.05, 'positive_branch_entropy_movement', 'increase'), 0)}/{fixed['primary_operating_points']}, {branch_counts.get((0.20, 'positive_branch_entropy_movement', 'increase'), 0)}/{fixed['primary_operating_points']}, and {branch_counts.get((0.50, 'positive_branch_entropy_movement', 'increase'), 0)}/{fixed['primary_operating_points']} points, respectively; negative results increased entropy for zero points at every anchor. The violin distributions include all points. Branch movement is not clinical benefit or harm, and expected mutual information remains non-negative. Primary estimator: continuity-corrected bivariate REML."
        ),
        "Table 1": "Table 1. Reusable four-question reporting architecture for the Diagnostic Information Profile. The four rows distinguish definition and operating-point performance, expected information before a result, observed-result probability after each branch, and action under explicit consequences and preferences. Missing threshold, reference-standard, consequence, or preference inputs remain visibly missing rather than inferred.",
        "Table 2": (
            f"Table 2. Worked Diagnostic Information Profile for the appendicitis example. This source-defined meta-analytic test-group record has incomplete defining metadata, including no separate index-test positivity threshold. Central estimates, approximate model-based 95% pooled-mean intervals, and predictive ranges are separately labeled and conditional on fitted heterogeneity. The {pct(p0, 0)} starting probability is illustrative. LR = likelihood ratio; n = number of studies. No action was selected because consequences, preferences, costs, and alternatives were not supplied. The editable table contains {len(table_2)} rows."
        ),
        "Figure S1": (
            f"Figure S1. Corpus and deterministic selection flow. Prespecified source selection retained {fixed['primary_operating_points']} operating points from {fixed['reviews']} reviews. They joined to {fixed['raw_normalized_rows_joined']} raw study rows; the recorded source-sheet restriction excluded {fixed['excluded_noncanonical_sheet_rows']} rows from one documented Zenodo exception and retained {fixed['analysis_rows_after_manifest_sheet_selection']} primary-analysis rows. All {trace['operating_points_with_source_identity']} points have source identity; {trace['operating_points_with_group_url']} have group-level URLs and {trace['operating_points_without_group_url']} retain archive-level identity without inferred URLs."
        ),
        "Figure S2": (
            f"Figure S2. Source fidelity, author-audit denominators, and fixed-sample route variation. (A) Source-reported versus primary-analysis pooled sensitivity and specificity for n={len(fidelity)} source-defined test groups; median absolute differences were {source_medians[0]:.4f} and {source_medians[1]:.4f}, maxima were {source_maxima[0]:.4f} and {source_maxima[1]:.4f}, and 3 points differed by at least 0.10. For CD007394 a 6-versus-7 row mismatch may contribute; the other two cannot be attributed because threshold and pooling metadata are incomplete. This is a fidelity check, not a truth standard. (B) The recorded author audit found 0 transcription errors/1640 cells and 0 acceptance errors/50 decisions; it is not independent adjudication. (C) Route-specific descriptive distributions at 20%, with operating-point n for each of {routes.cohort.nunique()} routes. Route summaries describe this sample only, not reproducibility or representativeness."
        ),
        "Figure S3": (
            f"Figure S3. Fixed-sample expected-information distributions and empirical concordance with Youden’s J across 5%, 20%, and 50% starting probabilities. Each anchor contains n={fixed['primary_operating_points']} source-defined meta-analytic test groups. Median expected information was {', '.join(f'{float(row.information_bits_median):.3f}' for row in anchor_summary.itertuples(index=False))} bits, and Spearman ρ was {', '.join(f'{float(row.spearman_information_vs_youden_j):.3f}' for row in anchor_summary.itertuples(index=False))}. Correlations are descriptive and sample-composition dependent; shared sensitivity and specificity inputs do not guarantee them."
        ),
        "Figure S4": (
            f"Figure S4. Limited recovery and numerical diagnostics for the primary continuity-corrected bivariate restricted maximum likelihood (REML) model. The recovery exercise is a smoke check, not evidence that the intervals have 95% repeated-sampling coverage. (A–B) In {recovery_summary['replicates']} scenarios with one dataset each, pooled-mean intervals included the generating sensitivity and specificity in {100 * float(recovery_summary['sensitivity_scenario_inclusion_fraction']):.1f}% and {100 * float(recovery_summary['specificity_scenario_inclusion_fraction']):.1f}%. These are approximate plug-in summaries conditional on fitted heterogeneity; heterogeneity-parameter uncertainty was not propagated and predictive ranges were not checked. (C) {diagnostic_summary['absolute_between_correlation_at_least_0_99']} fits had an absolute between-study correlation of at least 0.99, {diagnostic_summary['near_singular_between_covariance_min_eigenvalue_below_1e_8']} had near-singular covariance estimates, and {diagnostic_summary['optimizer_gradient_above_1e_4']} had a raw optimiser gradient above 10^-4. (D) Numerical ties included {diagnostic_summary['optimizer_tied_solution_log_l00_at_lower_bound']} sensitivity-heterogeneity and {diagnostic_summary['optimizer_tied_solution_log_l11_at_lower_bound']} specificity-heterogeneity lower-bound endpoints across {diagnostic_summary['optimizer_any_tied_solution_cholesky_diagonal_at_lower_bound']} fits; selected endpoints included {diagnostic_summary['optimizer_selected_endpoint_log_l00_at_lower_bound']} and {diagnostic_summary['optimizer_selected_endpoint_log_l11_at_lower_bound']}, respectively. A tied boundary signals fragility even when the selected endpoint is interior. Seven tied-boundary fits were missed by older flags, and the overall union was {diagnostic_summary['union_of_all_diagnostic_flags']}. Appendicitis had a tied specificity-heterogeneity boundary solution and selected correlation {diagnostic_summary['appendicitis_between_study_correlation']:.3f}; its joint predictive ranges are fragile."
        ),
        "Figure S5": (
            f"Figure S5. Sensitivity of expected information to analysis choices. These comparisons show movement across analysis families, not an effect attributable to any single modelling choice. (A) At 20%, plug-in expected information evaluated at each analysis’s central sensitivity and specificity differed between REML and the 15-node Gauss-Hermite (GH15) direct-binomial fit by median {float(estimator_effect['median_absolute_like_for_like_plugin_information_bits_at_20_percent_difference']):.4f} and maximum {float(estimator_effect['maximum_absolute_like_for_like_plugin_information_bits_at_20_percent_difference']):.4f} bits. This comparison combines likelihood family, regularisation or prior, and continuity-correction choices. The comparison with the GH15 propagated median was {float(gh15.median_absolute_information_bits_difference):.4f} and {float(gh15.maximum_absolute_information_bits_difference):.4f} bits, but those are different central-summary estimands. (B–C) Point-level GH15, wider-prior GH15, and 9-node Gauss-Hermite values are sensitivity summaries, not primary-analysis intervals."
        ),
        "Figure S6": (
            f"Figure S6. Exploratory within-review magnitudes and analysis-family movement at the 20% anchor. (A) Signed differences in expected information and post-negative probability for all {contrasts_summary['classified_contrasts']} preclassified contrasts, including {contrasts_summary['tradeoff_contrasts']} sensitivity–specificity trade-offs. (B) Movement from primary REML plug-in summaries to GH15 propagated-central summaries compares different estimands and must not be attributed purely to model family. Same-participant cells and cross-test covariance were available for 0 contrasts; no paired inference or superiority claim is shown."
        ),
        "Figure S7": (
            f"Figure S7. Near-tie and study-composition-restricted audits at the 20% anchor. (A) Counts below absolute information tolerances {', '.join(f'{float(v):.3f}' for v in info_tie.threshold)} bits among all {int(info_tie.comparisons.iloc[0])} contrasts and the trade-off subset. These are descriptive tolerances, not clinical equivalence margins. (B) Continuous magnitudes for {len(restricted)} contrasts with at least three overlapping study identifiers. Direct labels identify the six largest displayed magnitudes: P02, Typhidot versus TUBEX; P10, two frozen-section positivity definitions; P23, microhaematuria versus proteinuria; P24, microhaematuria versus leukocyturia; P48, head impulse versus test of skew; and P49, nystagmus versus test of skew. Identifier overlap does not establish common participants, paired outcomes, or cross-test covariance."
        ),
        "Figure S8": (
            f"Figure S8. Empirical carcinoembryonic-antigen (CEA) crossing sensitivity and additional theoretical checks. (A–B) The empirical expected-information crossing was {100 * canonical_crossing:.1f}% under primary restricted maximum likelihood (REML) and {100 * regularized_crossing:.1f}% under the historical regularised direct-binomial model using 15-node Gauss-Hermite quadrature (GH15); other model and numerical sensitivities are shown where a root exists. (C) Independent recalculations verify equal-J, near-equal-information, and synthetic ordering-crossing examples. The CEA example is inherited and model specific. None of the displayed probabilities is an action threshold."
        ),
    }
    return captions


def load_data(root: Path) -> dict[str, object]:
    analysis = root / "analysis/bmj_rmr_v042"
    data: dict[str, object] = {
        "profile": pd.read_csv(analysis / "appendicitis_worked_profile.csv"),
        "architecture": pd.read_csv(analysis / "table_1_reporting_architecture.csv"),
        "anchors": pd.read_csv(analysis / "derived_operating_points_standard_anchors.csv"),
        "landmarks": pd.read_csv(analysis / "atlas_landmarks.csv"),
        "theory_examples": pd.read_csv(analysis / "theoretical_non_equivalence_examples.csv"),
        "theory_profiles": pd.read_csv(analysis / "theoretical_crossing_probability_profile.csv"),
        "theory_summary": read_json(analysis / "theoretical_non_equivalence_summary.json"),
        "branches": pd.read_csv(analysis / "branch_entropy_all_primary.csv"),
        "branch_summary": pd.read_csv(analysis / "branch_entropy_summary.csv"),
        "run_manifest": read_json(analysis / "run_manifest.json"),
        "trace_summary": read_json(analysis / "source_trace_summary.json"),
        "exceptions": pd.read_csv(analysis / "analysis_row_selection_exceptions.csv"),
        "fidelity": pd.read_csv(analysis / "source_summary_fidelity.csv"),
        "audit_summary": read_json(analysis / "manual_audit_summary.json"),
        "routes": pd.read_csv(analysis / "source_route_stability.csv"),
        "anchor_summary": pd.read_csv(analysis / "standard_anchor_summary.csv"),
        "recovery": pd.read_csv(analysis / "reml_recovery.csv"),
        "recovery_summary": read_json(analysis / "reml_recovery_summary.json"),
        "diagnostics": pd.read_csv(analysis / "reml_diagnostics_all_primary.csv"),
        "diagnostic_summary": read_json(analysis / "reml_diagnostics_summary.json"),
        "sensitivity": pd.read_csv(analysis / "estimator_prior_numerical_sensitivity_all_primary.csv"),
        "sensitivity_summary": pd.read_csv(analysis / "estimator_prior_numerical_sensitivity_summary.csv"),
        "estimator_effect": read_json(analysis / "estimator_sensitivity_summary.json"),
        "contrasts": pd.read_csv(analysis / "exploratory_within_review_contrasts.csv"),
        "contrasts_summary": read_json(analysis / "exploratory_within_review_contrasts_summary.json"),
        "crossings": pd.read_csv(analysis / "empirical_cea_crossing_sensitivity.csv"),
        "cea_profiles": pd.read_csv(analysis / "empirical_cea_probability_profiles.csv"),
    }
    movement = contrast_model_movement(data["contrasts"], data["sensitivity"])
    near_ties, restricted = near_tie_and_composition_summary(data["contrasts"], data["anchors"])
    data["movement"] = movement
    data["near_ties"] = near_ties
    data["restricted"] = restricted
    return data


def validate_source_rows(root: Path) -> None:
    mapped = sorted({source for sources in DISPLAY_SOURCES.values() for source in sources})
    missing = [source for source in mapped if not (root / source).exists()]
    if missing:
        raise FileNotFoundError(f"Mapped display sources are missing: {missing}")
    for source in mapped:
        path = root / source
        expected = EXPECTED_SOURCE_ROWS.get(path.name)
        if expected is None or path.suffix.lower() != ".csv":
            continue
        observed = len(pd.read_csv(path))
        if observed != expected:
            raise AssertionError(f"Unexpected row count for {source}: {observed} != {expected}")


def validate_display_claim_alignment(root: Path) -> None:
    """Require every declared display claim to be assigned to a mapped source role."""

    claim_ledger = pd.read_csv(root / "docs/manuscript/v19/CLAIM_LEDGER.csv")
    ledger_claims = set(claim_ledger["claim_id"])
    for display in DISPLAY_ORDER:
        sources = DISPLAY_SOURCES[display]
        roles = DISPLAY_SOURCE_ROLES[display]
        source_claims = DISPLAY_SOURCE_CLAIMS[display]
        if not (len(sources) == len(roles) == len(source_claims)):
            raise AssertionError(f"Display source-role cardinality mismatch for {display}")
        declared = {value.strip() for value in DISPLAY_CLAIMS[display].split(";")}
        aligned = {claim for claims in source_claims for claim in claims}
        if declared != aligned:
            raise AssertionError(f"Display claim-source alignment mismatch for {display}: {declared} != {aligned}")
        if not declared <= ledger_claims:
            raise AssertionError(f"Display {display} references missing ledger claims: {declared - ledger_claims}")


def update_run_manifest(root: Path) -> None:
    path = root / "analysis/bmj_rmr_v042/run_manifest.json"
    payload = read_json(path)
    payload.update(
        {
            "phase": 4,
            "phase_4_builder": "scripts/bmj_rmr_v042_visuals.py",
            "phase_4_gate": "analysis/bmj_rmr_v042/qa/phase_gate_4.json",
            "phase_4_display_manifest": "analysis/bmj_rmr_v042/phase4_display_manifest.csv",
            "role": "Phase 4 deterministic display candidate complete; independent visual and production QA pending",
            "status": "phase_4_candidate_pending_independent_qa",
        }
    )
    write_json(path, payload)


def write_display_sources(output_analysis: Path, data: dict[str, object]) -> list[Path]:
    directory = output_analysis / "display_sources"
    frames: dict[str, pd.DataFrame] = {
        "figure_1_appendicitis_profile.csv": data["profile"],
        "figure_2_atlas_20_percent.csv": data["anchors"].loc[np.isclose(data["anchors"].starting_probability, 0.20)],
        "figure_2_landmarks.csv": data["landmarks"],
        "figure_3_theoretical_examples.csv": data["theory_examples"],
        "figure_3_theoretical_crossing_profile.csv": data["theory_profiles"],
        "figure_4_branch_entropy.csv": data["branches"],
        "figure_s2_source_fidelity.csv": data["fidelity"],
        "figure_s2_route_stability.csv": data["routes"],
        "figure_s3_anchor_summary.csv": data["anchor_summary"],
        "figure_s4_recovery.csv": data["recovery"],
        "figure_s4_diagnostics.csv": data["diagnostics"],
        "figure_s5_sensitivity_summary.csv": data["sensitivity_summary"],
        "figure_s6_contrast_movement.csv": data["movement"],
        "figure_s7_near_tie_summary.csv": data["near_ties"],
        "figure_s7_composition_restricted.csv": data["restricted"],
        "figure_s8_crossings.csv": data["crossings"],
    }
    fixed = data["run_manifest"]["fixed_reference_sample"]
    trace = data["trace_summary"]
    flow = pd.DataFrame.from_records(
        [
            {"order": 1, "stage": "Prespecified source selection", "count": fixed["primary_operating_points"], "unit": "operating points", "context": f"{fixed['reviews']} reviews"},
            {"order": 2, "stage": "Raw normalized rows joined", "count": fixed["raw_normalized_rows_joined"], "unit": "study rows", "context": "all selected operating points linked"},
            {"order": 3, "stage": "Rows from another source sheet excluded", "count": fixed["excluded_noncanonical_sheet_rows"], "unit": "study rows", "context": f"{fixed['row_selection_exceptions']} documented exception"},
            {"order": 4, "stage": "Primary-analysis rows", "count": fixed["analysis_rows_after_manifest_sheet_selection"], "unit": "study rows", "context": "recorded source-sheet restriction"},
            {"order": 5, "stage": "Points with source identity", "count": trace["operating_points_with_source_identity"], "unit": "operating points", "context": f"{trace['operating_points_with_group_url']} URLs; {trace['operating_points_without_group_url']} archive identities"},
        ]
    )
    frames["figure_s1_flow.csv"] = flow
    paths: list[Path] = []
    for name, frame in frames.items():
        path = directory / name
        write_csv(path, frame)
        paths.append(path)
    return paths


def write_editable_tables(manuscript_dir: Path, table_1: pd.DataFrame, table_2: pd.DataFrame, captions: dict[str, str]) -> list[Path]:
    directory = manuscript_dir / "tables" / f"v{VERSION}"
    directory.mkdir(parents=True, exist_ok=True)
    outputs: list[Path] = []
    table_specs = (
        (
            "table_1_reporting_architecture",
            table_1,
            captions["Table 1"],
            "All quantities require a defined operating point. Expected information and observed-result probability are conditioned on a stated starting probability; action additionally requires consequences and preferences.",
        ),
        (
            "table_2_appendicitis_worked_profile",
            table_2,
            captions["Table 2"],
            "PMI = model-based 95% pooled-mean interval; predictive range = model-based 95% range for an additional study’s latent operating point; LR = likelihood ratio. The 20% starting probability is illustrative, not an action threshold.",
        ),
    )
    for stem, frame, title, footnote in table_specs:
        csv_path = directory / f"{stem}.csv"
        write_csv(csv_path, frame)
        outputs.append(csv_path)
        md_path = directory / f"{stem}.md"
        md_path.write_text(f"{title}\n\n{markdown_table(frame)}\n\n*{footnote}*\n", encoding="utf-8", newline="\n")
        outputs.append(md_path)
        docx_path = directory / f"{stem}.docx"
        write_docx_table(docx_path, title, frame, footnote)
        outputs.append(docx_path)
    captions_path = directory / "TABLE_CAPTIONS.md"
    captions_path.write_text(
        "# Table captions\n\n" + "\n\n".join(captions[name] for name in ("Table 1", "Table 2")) + "\n",
        encoding="utf-8",
        newline="\n",
    )
    outputs.append(captions_path)
    return outputs


def write_caption_files(manuscript_dir: Path, output_analysis: Path, captions: dict[str, str]) -> list[Path]:
    figures_dir = manuscript_dir / "figures" / f"v{VERSION}"
    figures_dir.mkdir(parents=True, exist_ok=True)
    markdown = ["# Figure captions"]
    records: list[dict[str, str]] = []
    for display in DISPLAY_ORDER:
        if not display.startswith("Figure"):
            continue
        markdown.extend(
            [
                "",
                f"## {display}",
                "",
                captions[display],
                "",
                f"Sources: {'; '.join(DISPLAY_SOURCES[display])}",
                "",
                f"Claims: {DISPLAY_CLAIMS[display]}",
            ]
        )
        records.append(
            {
                "display": display,
                "caption": captions[display],
                "source_files": "; ".join(DISPLAY_SOURCES[display]),
                "claim_ids": DISPLAY_CLAIMS[display],
            }
        )
    md_path = figures_dir / "FIGURE_CAPTIONS.md"
    md_path.write_text("\n".join(markdown) + "\n", encoding="utf-8", newline="\n")
    csv_path = output_analysis / "phase4_figure_captions.csv"
    write_csv(csv_path, pd.DataFrame.from_records(records))
    return [md_path, csv_path]


def render_pdf_previews(figures_dir: Path, qa_dir: Path) -> list[Path]:
    render_dir = qa_dir / "phase4" / "pdf_renders"
    render_dir.mkdir(parents=True, exist_ok=True)
    outputs: list[Path] = []
    for path in sorted(figures_dir.glob("*.pdf")):
        document = fitz.open(path)
        if len(document) != 1:
            raise AssertionError(f"Expected one page in {path}")
        pixmap = document[0].get_pixmap(matrix=fitz.Matrix(2, 2), alpha=False)
        output = render_dir / f"{path.stem}.png"
        pixmap.save(output)
        outputs.append(output)
        document.close()
    return outputs


def audit_pdf_text_bounds(figures_dir: Path, qa_dir: Path) -> list[Path]:
    """Audit every non-empty PDF text span against its one-page media box.

    The 0.1-point tolerance is one seven-hundred-and-twentieth of an inch and
    protects the diagnostic from harmless floating-point representation noise.
    The persisted strict count separately proves that the final masters contain
    no span with any actual out-of-page coordinate.
    """

    summary_records: list[dict[str, object]] = []
    violation_records: list[dict[str, object]] = []
    for path in sorted(figures_dir.glob("*.pdf")):
        document = fitz.open(path)
        if document.page_count != 1:
            document.close()
            raise AssertionError(f"Expected one page in {path}")
        page = document[0]
        page_rect = page.rect
        span_bounds: list[tuple[float, float, float, float]] = []
        strict_count = 0
        tolerance_count = 0
        worst_overrun = 0.0
        for block in page.get_text("dict").get("blocks", []):
            if block.get("type") != 0:
                continue
            for line in block.get("lines", []):
                for span in line.get("spans", []):
                    text_value = str(span.get("text", ""))
                    if not text_value.strip():
                        continue
                    x0, y0, x1, y1 = (float(value) for value in span["bbox"])
                    span_bounds.append((x0, y0, x1, y1))
                    left = max(0.0, -x0)
                    top = max(0.0, -y0)
                    right = max(0.0, x1 - float(page_rect.width))
                    bottom = max(0.0, y1 - float(page_rect.height))
                    overrun = max(left, top, right, bottom)
                    if overrun > 0:
                        strict_count += 1
                    if overrun > PDF_TEXT_BOUND_TOLERANCE_PT:
                        tolerance_count += 1
                        violation_records.append(
                            {
                                "pdf_file": path.name,
                                "text": text_value,
                                "x0_pt": x0,
                                "y0_pt": y0,
                                "x1_pt": x1,
                                "y1_pt": y1,
                                "left_overrun_pt": left,
                                "top_overrun_pt": top,
                                "right_overrun_pt": right,
                                "bottom_overrun_pt": bottom,
                                "maximum_overrun_pt": overrun,
                            }
                        )
                    worst_overrun = max(worst_overrun, overrun)
        if not span_bounds:
            document.close()
            raise AssertionError(f"No text spans found in {path}")
        min_x0 = min(value[0] for value in span_bounds)
        min_y0 = min(value[1] for value in span_bounds)
        max_x1 = max(value[2] for value in span_bounds)
        max_y1 = max(value[3] for value in span_bounds)
        summary_records.append(
            {
                "pdf_file": path.name,
                "page_width_pt": float(page_rect.width),
                "page_height_pt": float(page_rect.height),
                "text_span_count": len(span_bounds),
                "minimum_x0_pt": min_x0,
                "minimum_y0_pt": min_y0,
                "maximum_x1_pt": max_x1,
                "maximum_y1_pt": max_y1,
                "minimum_left_margin_pt": min_x0,
                "minimum_top_margin_pt": min_y0,
                "minimum_right_margin_pt": float(page_rect.width) - max_x1,
                "minimum_bottom_margin_pt": float(page_rect.height) - max_y1,
                "strict_out_of_page_span_count": strict_count,
                "beyond_tolerance_span_count": tolerance_count,
                "worst_overrun_pt": worst_overrun,
                "tolerance_pt": PDF_TEXT_BOUND_TOLERANCE_PT,
                "status": "pass" if tolerance_count == 0 else "fail",
            }
        )
        document.close()

    audit_dir = qa_dir / "phase4"
    summary_path = audit_dir / "pdf_text_bounds_audit.csv"
    violations_path = audit_dir / "pdf_text_bounds_violations.csv"
    json_path = audit_dir / "pdf_text_bounds_audit.json"
    summary_frame = pd.DataFrame.from_records(summary_records)
    violation_columns = (
        "pdf_file",
        "text",
        "x0_pt",
        "y0_pt",
        "x1_pt",
        "y1_pt",
        "left_overrun_pt",
        "top_overrun_pt",
        "right_overrun_pt",
        "bottom_overrun_pt",
        "maximum_overrun_pt",
    )
    violation_frame = pd.DataFrame.from_records(violation_records, columns=violation_columns)
    write_csv(summary_path, summary_frame)
    write_csv(violations_path, violation_frame)
    write_json(
        json_path,
        {
            "analysis_version": VERSION,
            "status": "pass" if not violation_records else "fail",
            "tolerance_pt": PDF_TEXT_BOUND_TOLERANCE_PT,
            "pdf_count": len(summary_frame),
            "text_span_count": int(summary_frame["text_span_count"].sum()),
            "strict_out_of_page_span_count": int(summary_frame["strict_out_of_page_span_count"].sum()),
            "beyond_tolerance_span_count": int(summary_frame["beyond_tolerance_span_count"].sum()),
            "worst_overrun_pt": float(summary_frame["worst_overrun_pt"].max()),
            "minimum_text_margin_pt": float(
                summary_frame[
                    [
                        "minimum_left_margin_pt",
                        "minimum_top_margin_pt",
                        "minimum_right_margin_pt",
                        "minimum_bottom_margin_pt",
                    ]
                ].min().min()
            ),
            "method": "PyMuPDF non-empty text-span bounding boxes checked against each one-page PDF media box.",
        },
    )
    if violation_records:
        details = "; ".join(
            f"{row['pdf_file']}: {row['text']!r} ({row['maximum_overrun_pt']:.3f} pt)"
            for row in violation_records
        )
        raise AssertionError(f"PDF text-bound audit failed beyond {PDF_TEXT_BOUND_TOLERANCE_PT} pt: {details}")
    return [summary_path, violations_path, json_path]


def pdf_text_spans(path: Path) -> list[dict[str, object]]:
    document = fitz.open(path)
    if document.page_count != 1:
        document.close()
        raise AssertionError(f"Expected one page in {path}")
    records: list[dict[str, object]] = []
    for block in document[0].get_text("dict").get("blocks", []):
        if block.get("type") != 0:
            continue
        for line in block.get("lines", []):
            for span in line.get("spans", []):
                text_value = str(span.get("text", "")).strip()
                if text_value:
                    records.append({"text": text_value, "rect": fitz.Rect(span["bbox"])})
    document.close()
    return records


def union_text_rect(spans: list[dict[str, object]], texts: tuple[str, ...]) -> fitz.Rect:
    selected = [record["rect"] for text_value in texts for record in spans if record["text"] == text_value]
    if len(selected) != len(texts):
        found = [record["text"] for record in spans if record["text"] in texts]
        raise AssertionError(f"Expected one span for each of {texts}; found {found}")
    rectangle = fitz.Rect(selected[0])
    for item in selected[1:]:
        rectangle |= item
    return rectangle


def collision_metrics(left: fitz.Rect, right: fitz.Rect) -> dict[str, float | bool]:
    overlap_width = max(0.0, min(left.x1, right.x1) - max(left.x0, right.x0))
    overlap_height = max(0.0, min(left.y1, right.y1) - max(left.y0, right.y0))
    horizontal_gap = -overlap_width if overlap_width > 0 else max(right.x0 - left.x1, left.x0 - right.x1)
    vertical_gap = -overlap_height if overlap_height > 0 else max(right.y0 - left.y1, left.y0 - right.y1)
    collision = overlap_width > 0 and overlap_height > 0
    separation = math.hypot(max(0.0, horizontal_gap), max(0.0, vertical_gap))
    return {
        "horizontal_gap_pt": float(horizontal_gap),
        "vertical_gap_pt": float(vertical_gap),
        "overlap_width_pt": float(overlap_width),
        "overlap_height_pt": float(overlap_height),
        "overlap_area_pt2": float(overlap_width * overlap_height),
        "minimum_separating_gap_pt": float(separation),
        "collision": collision,
    }


def audit_targeted_internal_collisions(figures_dir: Path, qa_dir: Path) -> list[Path]:
    """Persist regression checks for the three independently reported internal collisions."""

    records: list[dict[str, object]] = []
    for suffix, variant in (("", "color"), ("_grayscale", "grayscale")):
        f2_name = f"figure_2_signature_atlas{suffix}.pdf"
        f2_spans = pdf_text_spans(figures_dir / f2_name)
        f2_target_record = next(record for record in f2_spans if record["text"] == "Operating points")
        comparisons = []
        for record in f2_spans:
            if record is f2_target_record:
                continue
            metrics = collision_metrics(f2_target_record["rect"], record["rect"])
            comparisons.append((metrics, record))
        collisions = [item for item in comparisons if item[0]["collision"]]
        chosen_metrics, chosen_record = (
            max(collisions, key=lambda item: item[0]["overlap_area_pt2"])
            if collisions
            else min(comparisons, key=lambda item: item[0]["minimum_separating_gap_pt"])
        )
        records.append(
            {
                "pdf_file": f2_name,
                "variant": variant,
                "check_id": "f2_panel_b_y_label_vs_other_text",
                "left_text": "Operating points",
                "right_text": chosen_record["text"],
                **chosen_metrics,
                "status": "fail" if collisions else "pass",
            }
        )

        s4_name = f"figure_s4_recovery_and_diagnostics{suffix}.pdf"
        s4_spans = pdf_text_spans(figures_dir / s4_name)
        diagnostic_groups = (
            (("|corr.|", "≥0.99"), "absolute correlation flag"),
            (("near-", "singular", "covariance"), "near-singular covariance flag"),
            (("gradient", ">10⁻⁴"), "optimizer gradient flag"),
        )
        group_rectangles = [union_text_rect(s4_spans, texts) for texts, _ in diagnostic_groups]
        for index in range(2):
            metrics = collision_metrics(group_rectangles[index], group_rectangles[index + 1])
            records.append(
                {
                    "pdf_file": s4_name,
                    "variant": variant,
                    "check_id": f"s4_diagnostic_tick_group_{index + 1}_vs_{index + 2}",
                    "left_text": diagnostic_groups[index][1],
                    "right_text": diagnostic_groups[index + 1][1],
                    **metrics,
                    "status": "fail" if metrics["collision"] else "pass",
                }
            )

        s7_name = f"figure_s7_near_ties_and_composition_audit{suffix}.pdf"
        s7_spans = pdf_text_spans(figures_dir / s7_name)
        p49 = union_text_rect(s7_spans, ("P49",))
        p24 = union_text_rect(s7_spans, ("P24",))
        metrics = collision_metrics(p49, p24)
        records.append(
            {
                "pdf_file": s7_name,
                "variant": variant,
                "check_id": "s7_p49_vs_p24_direct_labels",
                "left_text": "P49",
                "right_text": "P24",
                **metrics,
                "status": "fail" if metrics["collision"] else "pass",
            }
        )
        direct_labels = [record for record in s7_spans if re.fullmatch(r"P\d{2}", str(record["text"]))]
        label_comparisons = []
        for left_index, left_record in enumerate(direct_labels):
            for right_record in direct_labels[left_index + 1 :]:
                label_comparisons.append(
                    (collision_metrics(left_record["rect"], right_record["rect"]), left_record, right_record)
                )
        label_collisions = [item for item in label_comparisons if item[0]["collision"]]
        selected_metrics, selected_left, selected_right = (
            max(label_collisions, key=lambda item: item[0]["overlap_area_pt2"])
            if label_collisions
            else min(label_comparisons, key=lambda item: item[0]["minimum_separating_gap_pt"])
        )
        records.append(
            {
                "pdf_file": s7_name,
                "variant": variant,
                "check_id": "s7_all_direct_labels_pairwise",
                "left_text": selected_left["text"],
                "right_text": selected_right["text"],
                **selected_metrics,
                "status": "fail" if label_collisions else "pass",
            }
        )

    frame = pd.DataFrame.from_records(records)
    audit_dir = qa_dir / "phase4"
    csv_path = audit_dir / "internal_text_collision_audit.csv"
    json_path = audit_dir / "internal_text_collision_audit.json"
    write_csv(csv_path, frame)
    collision_count = int(frame["collision"].sum())
    write_json(
        json_path,
        {
            "analysis_version": VERSION,
            "status": "pass" if collision_count == 0 else "fail",
            "method": "Targeted PyMuPDF text-span and multiline-label union geometry checks in the affected color and grayscale PDF masters.",
            "pdf_count": 6,
            "check_count": len(frame),
            "collision_count": collision_count,
            "minimum_separating_gap_pt": float(frame["minimum_separating_gap_pt"].min()),
            "records": frame.to_dict(orient="records"),
        },
    )
    if collision_count:
        failed = "; ".join(f"{row.pdf_file}: {row.check_id}" for row in frame.loc[frame.collision].itertuples())
        raise AssertionError(f"Targeted internal-text collision audit failed: {failed}")
    return [csv_path, json_path]


def render_collision_previews_600dpi(figures_dir: Path, qa_dir: Path) -> list[Path]:
    render_dir = qa_dir / "phase4" / "collision_renders_600dpi"
    render_dir.mkdir(parents=True, exist_ok=True)
    outputs: list[Path] = []
    for stem in COLLISION_AUDIT_STEMS:
        for suffix in ("", "_grayscale"):
            path = figures_dir / f"{stem}{suffix}.pdf"
            document = fitz.open(path)
            pixmap = document[0].get_pixmap(matrix=fitz.Matrix(DPI / 72, DPI / 72), alpha=False)
            pixmap.set_dpi(DPI, DPI)
            output = render_dir / f"{path.stem}.png"
            pixmap.save(output)
            outputs.append(output)
            document.close()
    return outputs


def contact_sheet(
    paths: list[Path], output: Path, title: str, *, dpi: int = 150, label_variant: bool = True
) -> None:
    columns = 3
    tile_width = 880
    tile_height = 680
    header_height = 74
    rows = math.ceil(len(paths) / columns)
    canvas = Image.new("RGB", (columns * tile_width, header_height + rows * tile_height), "white")
    draw = ImageDraw.Draw(canvas)
    font_path = matplotlib.font_manager.findfont("DejaVu Sans")
    title_font = ImageFont.truetype(font_path, 32)
    label_font = ImageFont.truetype(font_path, 22)
    draw.text((24, 18), title, fill="black", font=title_font)
    for index, path in enumerate(paths):
        image = Image.open(path).convert("RGB")
        image.thumbnail((tile_width - 24, tile_height - 58), Image.Resampling.LANCZOS)
        x0 = (index % columns) * tile_width
        y0 = header_height + (index // columns) * tile_height
        x = x0 + (tile_width - image.width) // 2
        y = y0 + 40 + (tile_height - 50 - image.height) // 2
        canvas.paste(image, (x, y))
        base_stem = path.stem.removesuffix("_grayscale")
        display = next((name for name, spec in FIGURE_SPECS.items() if spec["stem"] == base_stem), base_stem)
        label = (
            f"{display}: {'grayscale' if path.stem.endswith('_grayscale') else 'color'}"
            if label_variant
            else display
        )
        draw.text((x0 + 12, y0 + 7), label, fill="black", font=label_font)
        draw.rectangle((x0, y0, x0 + tile_width - 1, y0 + tile_height - 1), outline="#A0A0A0", width=2)
    output.parent.mkdir(parents=True, exist_ok=True)
    canvas.save(output, format="PNG", optimize=False, dpi=(dpi, dpi))


def write_contact_sheets(previews: list[Path], qa_dir: Path) -> list[Path]:
    color = [path for path in previews if not path.stem.endswith("_grayscale")]
    gray = [path for path in previews if path.stem.endswith("_grayscale")]
    outputs = [qa_dir / "phase4" / "contact_sheet_color.png", qa_dir / "phase4" / "contact_sheet_grayscale.png"]
    contact_sheet(color, outputs[0], f"BMJ RMR v{VERSION}: color PDF renders")
    contact_sheet(gray, outputs[1], f"BMJ RMR v{VERSION}: grayscale PDF renders")
    return outputs


def write_collision_contact_sheets(previews: list[Path], qa_dir: Path) -> list[Path]:
    color = [path for path in previews if not path.stem.endswith("_grayscale")]
    gray = [path for path in previews if path.stem.endswith("_grayscale")]
    outputs = [
        qa_dir / "phase4" / "collision_contact_sheet_color_600dpi.png",
        qa_dir / "phase4" / "collision_contact_sheet_grayscale_600dpi.png",
    ]
    contact_sheet(
        color,
        outputs[0],
        "Phase 4 collision corrections: color 600-dpi PDF renders",
        dpi=DPI,
        label_variant=False,
    )
    contact_sheet(
        gray,
        outputs[1],
        "Phase 4 collision corrections: grayscale 600-dpi PDF renders",
        dpi=DPI,
        label_variant=False,
    )
    return outputs


def artifact_rows(root: Path, paths: list[Path]) -> pd.DataFrame:
    records = []
    for path in sorted(set(paths)):
        if not path.exists() or not path.is_file():
            continue
        suffix = path.suffix.lower().lstrip(".")
        width_px: int | None = None
        height_px: int | None = None
        dpi: float | None = None
        width_mm: float | None = None
        height_mm: float | None = None
        if suffix == "png":
            with Image.open(path) as image:
                width_px, height_px = image.size
                dpi_value = image.info.get("dpi")
                if dpi_value:
                    dpi = float(dpi_value[0])
        elif suffix == "pdf" and "figures" in path.parts:
            document = fitz.open(path)
            rectangle = document[0].rect
            width_mm = float(rectangle.width / 72 * MM_PER_INCH)
            height_mm = float(rectangle.height / 72 * MM_PER_INCH)
            document.close()
        records.append(
            {
                "path": path.relative_to(root).as_posix() if path.is_relative_to(root) else str(path),
                "format": suffix,
                "bytes": path.stat().st_size,
                "sha256": file_hash(path),
                "width_px": width_px,
                "height_px": height_px,
                "dpi": dpi,
                "width_mm": width_mm,
                "height_mm": height_mm,
            }
        )
    return pd.DataFrame.from_records(records)


def source_hash_rows(root: Path) -> pd.DataFrame:
    records: list[dict[str, object]] = []
    for source in sorted({item for values in DISPLAY_SOURCES.values() for item in values}):
        path = root / source
        rows: int | None = None
        if path.suffix.lower() == ".csv":
            rows = len(pd.read_csv(path))
        records.append(
            {
                "source_file": source,
                "sha256": file_hash(path),
                "rows": rows,
                "mapped_displays": "; ".join(display for display, sources in DISPLAY_SOURCES.items() if source in sources),
            }
        )
    return pd.DataFrame.from_records(records)


def display_manifest(captions: dict[str, str]) -> pd.DataFrame:
    records = []
    for order, display in enumerate(DISPLAY_ORDER, start=1):
        if display.startswith("Figure"):
            spec = FIGURE_SPECS[display]
            outputs = "; ".join(
                f"docs/manuscript/v19/figures/v{VERSION}/{spec['stem']}{gray}.{extension}"
                for gray in ("", "_grayscale")
                for extension in ("pdf", "svg", "png")
            )
            dimensions = f"{DOUBLE_COLUMN_MM:.0f} × {float(spec['height_mm']):.0f} mm"
        else:
            table_number = display.split()[-1]
            stem = "table_1_reporting_architecture" if table_number == "1" else "table_2_appendicitis_worked_profile"
            outputs = "; ".join(f"docs/manuscript/v19/tables/v{VERSION}/{stem}.{extension}" for extension in ("csv", "md", "docx"))
            dimensions = "editable table; not rasterized"
        records.append(
            {
                "order": order,
                "display": display,
                "source_files": "; ".join(DISPLAY_SOURCES[display]),
                "source_roles": " | ".join(DISPLAY_SOURCE_ROLES[display]),
                "claim_ids": DISPLAY_CLAIMS[display],
                "claim_source_alignment": " | ".join(
                    f"{source} -> {','.join(claims) if claims else 'context only'}"
                    for source, claims in zip(DISPLAY_SOURCES[display], DISPLAY_SOURCE_CLAIMS[display])
                ),
                "outputs": outputs,
                "final_dimensions": dimensions,
                "caption": captions[display],
                "classification_encoding": "empirical/theoretical/illustrative labels embedded where applicable",
                "redundant_encoding": "four-question colors plus markers, line styles, hatches, and direct labels",
            }
        )
    return pd.DataFrame.from_records(records)


def initial_visual_log(qa_dir: Path, previews: list[Path]) -> Path:
    path = qa_dir / "phase4" / "visual_inspection_log.json"
    if path.exists():
        return path
    records = [
        {
            "display": path_item.stem.replace("_grayscale", ""),
            "variant": "grayscale" if path_item.stem.endswith("_grayscale") else "color",
            "render_path": path_item.as_posix(),
            "inspection_status": "pending_builder_visual_inspection",
            "defects": [],
            "fixes": [],
        }
        for path_item in previews
    ]
    write_json(
        path,
        {
            "analysis_version": VERSION,
            "inspection_method": "Every one-page PDF rendered at 144 dpi; color and grayscale contact sheets plus individual renders inspected with view_image.",
            "minimum_effective_text_points": 8.0,
            "records": records,
            "overall_status": "pending_builder_visual_inspection",
        },
    )
    return path


def write_visual_markdown(qa_dir: Path) -> Path:
    json_path = qa_dir / "phase4" / "visual_inspection_log.json"
    payload = read_json(json_path)
    lines = [
        "# Phase 4 visual inspection log",
        "",
        f"Overall status: `{payload['overall_status']}`",
        "",
        str(payload["inspection_method"]),
        "",
        "| Display render | Variant | Status | Defects found | Fixes applied |",
        "|---|---|---|---|---|",
    ]
    for record in payload["records"]:
        defects = "; ".join(record.get("defects", [])) or "None"
        fixes = "; ".join(record.get("fixes", [])) or "None"
        lines.append(f"| `{record['display']}` | {record['variant']} | {record['inspection_status']} | {defects} | {fixes} |")
    lines.extend(
        [
            "",
            "Contact sheets:",
            "",
            "- `analysis/bmj_rmr_v042/qa/phase4/contact_sheet_color.png`",
            "- `analysis/bmj_rmr_v042/qa/phase4/contact_sheet_grayscale.png`",
            "- `analysis/bmj_rmr_v042/qa/phase4/collision_contact_sheet_color_600dpi.png`",
            "- `analysis/bmj_rmr_v042/qa/phase4/collision_contact_sheet_grayscale_600dpi.png`",
        ]
    )
    if "targeted_collision_reinspection" in payload:
        targeted = payload["targeted_collision_reinspection"]
        lines.extend(
            [
                "",
                f"Targeted 600-dpi collision reinspection: `{targeted['status']}`; "
                f"{targeted['individual_600dpi_renders']} individual renders, "
                f"{targeted['collision_contact_sheets_reinspected']} collision contact sheets, and "
                f"{targeted['full_contact_sheets_reinspected']} full contact sheets inspected.",
                "",
                str(targeted["secondary_defect_found_and_fixed"]),
            ]
        )
    path = qa_dir / "phase4" / "visual_inspection_log.md"
    path.write_text("\n".join(lines) + "\n", encoding="utf-8", newline="\n")
    return path


def write_phase_gate(root: Path, output_analysis: Path, qa_dir: Path) -> Path:
    artifact_manifest_path = output_analysis / "phase4_artifact_manifest.csv"
    source_manifest_path = output_analysis / "phase4_source_hash_manifest.csv"
    artifacts = pd.read_csv(artifact_manifest_path)
    sources = pd.read_csv(source_manifest_path)
    inspection_path = qa_dir / "phase4" / "visual_inspection_log.json"
    inspection = read_json(inspection_path) if inspection_path.exists() else {"overall_status": "missing"}
    verification_path = qa_dir / "phase4" / "builder_verification.json"
    verification = read_json(verification_path) if verification_path.exists() else {"status": "pending final test run"}
    bounds_path = qa_dir / "phase4" / "pdf_text_bounds_audit.json"
    bounds_audit = read_json(bounds_path) if bounds_path.exists() else {"status": "missing"}
    collision_path = qa_dir / "phase4" / "internal_text_collision_audit.json"
    collision_audit = read_json(collision_path) if collision_path.exists() else {"status": "missing"}
    docx_path = qa_dir / "phase4" / "docx_inspection_log.json"
    docx_inspection = read_json(docx_path) if docx_path.exists() else {"status": "missing"}
    run_manifest = read_json(root / "analysis/bmj_rmr_v042/run_manifest.json")
    payload = {
        "analysis_version": VERSION,
        "phase": 4,
        "name": "deterministic figures and editable tables",
        "status": "candidate_pending_independent_qa",
        "candidate_commit": run_manifest.get("phase_4_artifact_candidate_commit", "pending_commit_at_builder_handoff"),
        "independent_qa_required": True,
        "canonical_estimator": "continuity-corrected bivariate REML",
        "checks": {
            "all_mapped_displays_built": bool(len(artifacts) > 0),
            "color_and_grayscale_variants_built": True,
            "pdf_svg_and_600_dpi_png_built": True,
            "editable_tables_built": True,
            "captions_source_aligned": True,
            "redundant_encoding_declared": True,
            "builder_visual_inspection_complete": inspection.get("overall_status") == "pass_after_fixes",
            "pdf_text_bounds_audit_complete_and_passed": bounds_audit.get("status") == "pass",
            "targeted_internal_collision_audit_complete_and_passed": collision_audit.get("status") == "pass",
            "docx_production_inspection_complete": docx_inspection.get("status") == "pass",
            "gate_4_passed": False,
        },
        "commands": [
            "py -3.13 scripts/bmj_rmr_v042_visuals.py build",
            "py -3.13 -m pytest tests/test_bmj_rmr_v042_phase4.py -q",
            "py -3.13 -m pytest tests/test_bmj_rmr_v042.py tests/test_bmj_rmr_v042_spec.py tests/test_bmj_rmr_v042_phase3.py tests/test_bmj_rmr_v042_phase4.py tests/test_cochrane_dta_atlas.py -q",
            "git diff --check",
        ],
        "source_sha256": {row.source_file: row.sha256 for row in sources.itertuples(index=False)},
        "output_sha256": {row.path: row.sha256 for row in artifacts.itertuples(index=False)},
        "visual_inspection": inspection,
        "pdf_text_bounds_audit": bounds_audit,
        "targeted_internal_collision_audit": collision_audit,
        "docx_production_inspection": docx_inspection,
        "builder_verification": verification,
        "limitations": [
            "The figures display model-based summaries from the primary REML hierarchy; no figure supplies disease-specific action thresholds.",
            "The CEA crossing, estimator sensitivity, and within-review contrasts remain supplementary and model-dependent or exploratory.",
            "Independent clinical-reader and journal-production QA remains required before Gate 4 can pass.",
        ],
    }
    path = qa_dir / "phase_gate_4.json"
    write_json(path, payload)
    return path


def build(root: Path, output_analysis: Path, manuscript_dir: Path, qa_dir: Path, *, update_control: bool) -> None:
    if update_control:
        update_run_manifest(root)
    validate_source_rows(root)
    validate_display_claim_alignment(root)
    data = load_data(root)
    figures_dir = manuscript_dir / "figures" / f"v{VERSION}"
    figure_builders = {
        "Figure 1": lambda gray: figure_1(data["profile"], data["architecture"], gray),
        "Figure 2": lambda gray: figure_2(data["anchors"], data["landmarks"], gray),
        "Figure 3": lambda gray: figure_3(data["theory_examples"], data["theory_profiles"], data["theory_summary"], gray),
        "Figure 4": lambda gray: figure_4(data["branches"], data["branch_summary"], gray),
        "Figure S1": lambda gray: figure_s1(data["run_manifest"], data["trace_summary"], data["exceptions"], gray),
        "Figure S2": lambda gray: figure_s2(data["fidelity"], data["audit_summary"], data["routes"], gray),
        "Figure S3": lambda gray: figure_s3(data["anchors"], data["anchor_summary"], gray),
        "Figure S4": lambda gray: figure_s4(data["recovery"], data["recovery_summary"], data["diagnostics"], data["diagnostic_summary"], gray),
        "Figure S5": lambda gray: figure_s5(data["sensitivity"], data["sensitivity_summary"], data["estimator_effect"], gray),
        "Figure S6": lambda gray: figure_s6(data["contrasts"], data["movement"], gray),
        "Figure S7": lambda gray: figure_s7(data["near_ties"], data["restricted"], gray),
        "Figure S8": lambda gray: figure_s8(data["crossings"], data["cea_profiles"], data["theory_summary"], gray),
    }
    figure_paths: list[Path] = []
    for display in FIGURE_SPECS:
        for grayscale in (False, True):
            figure_paths.extend(save_figure(figure_builders[display](grayscale), figures_dir, display, grayscale))

    table_1 = build_table_1(data["architecture"])
    table_2 = build_table_2(data["profile"])
    captions = build_captions(data, table_2)
    display_source_paths = write_display_sources(output_analysis, data)
    table_paths = write_editable_tables(manuscript_dir, table_1, table_2, captions)
    caption_paths = write_caption_files(manuscript_dir, output_analysis, captions)
    manifest = display_manifest(captions)
    display_manifest_path = output_analysis / "phase4_display_manifest.csv"
    write_csv(display_manifest_path, manifest)
    encoding_path = output_analysis / "phase4_encoding_manifest.csv"
    write_csv(
        encoding_path,
        pd.DataFrame(
            {
                "question": [1, 2, 3, 4],
                "role": ["operating point", "expected information", "observed result", "action"],
                "color_hex": QUESTION_COLORS,
                "grayscale_hex": QUESTION_GRAYS,
                "marker": QUESTION_MARKERS,
                "line_style": QUESTION_LINESTYLES,
                "hatch": HATCHES,
            }
        ),
    )

    pdf_text_audit_paths = audit_pdf_text_bounds(figures_dir, qa_dir)
    internal_collision_audit_paths = audit_targeted_internal_collisions(figures_dir, qa_dir)
    previews = render_pdf_previews(figures_dir, qa_dir)
    collision_previews = render_collision_previews_600dpi(figures_dir, qa_dir)
    contact_paths = write_contact_sheets(previews, qa_dir)
    collision_contact_paths = write_collision_contact_sheets(collision_previews, qa_dir)
    visual_json = initial_visual_log(qa_dir, previews)
    visual_md = write_visual_markdown(qa_dir)
    source_manifest_path = output_analysis / "phase4_source_hash_manifest.csv"
    write_csv(source_manifest_path, source_hash_rows(root))
    artifact_paths = [
        *figure_paths,
        *display_source_paths,
        *table_paths,
        *caption_paths,
        display_manifest_path,
        encoding_path,
        root / "docs/manuscript/v19/FIGURE_TABLE_MAP.md",
        root / "docs/manuscript/v19/CLAIM_LEDGER.csv",
        *pdf_text_audit_paths,
        *internal_collision_audit_paths,
        *previews,
        *collision_previews,
        *contact_paths,
        *collision_contact_paths,
        visual_json,
        visual_md,
    ]
    builder_verification_path = qa_dir / "phase4" / "builder_verification.json"
    if builder_verification_path.exists():
        artifact_paths.append(builder_verification_path)
    docx_qa_paths = sorted((qa_dir / "phase4" / "docx_renders").glob("*.png"))
    for optional_path in (
        qa_dir / "phase4" / "docx_inspection_log.json",
        qa_dir / "phase4" / "docx_inspection_log.md",
    ):
        if optional_path.exists():
            docx_qa_paths.append(optional_path)
    artifact_paths.extend(docx_qa_paths)
    artifact_manifest_path = output_analysis / "phase4_artifact_manifest.csv"
    write_csv(artifact_manifest_path, artifact_rows(root, artifact_paths))
    if update_control:
        write_phase_gate(root, output_analysis, qa_dir)


def parse_args() -> argparse.Namespace:
    parser = argparse.ArgumentParser(description=__doc__)
    parser.add_argument("command", choices=("build", "gate"), nargs="?", default="build")
    parser.add_argument("--root", type=Path, default=Path(__file__).resolve().parents[1])
    parser.add_argument("--output-analysis", type=Path)
    parser.add_argument("--manuscript-dir", type=Path)
    parser.add_argument("--qa-dir", type=Path)
    parser.add_argument("--no-control-update", action="store_true")
    return parser.parse_args()


def main() -> None:
    args = parse_args()
    root = args.root.resolve()
    output_analysis = (args.output_analysis or root / "analysis/bmj_rmr_v042").resolve()
    manuscript_dir = (args.manuscript_dir or root / "docs/manuscript/v19").resolve()
    qa_dir = (args.qa_dir or root / "analysis/bmj_rmr_v042/qa").resolve()
    if args.command == "build":
        build(root, output_analysis, manuscript_dir, qa_dir, update_control=not args.no_control_update)
    else:
        write_visual_markdown(qa_dir)
        write_phase_gate(root, output_analysis, qa_dir)


if __name__ == "__main__":
    main()
```
