## Supplementary material for "The information in diagnostic tests": Executable analysis code, derived inputs, source-acquisition records, tests, and release metadata: bmj_rmr_v100_visuals.py

```
"""Build the Information EBM 1.0.0 figures from the locked v0.42 analysis."""

from __future__ import annotations

import argparse
from pathlib import Path

import matplotlib.pyplot as plt
import numpy as np
import pandas as pd
from matplotlib.lines import Line2D

from scripts import bmj_rmr_v042_visuals as base


VERSION = "1.0.0"
RELEASE_DATE = "2026-08-28"

FIGURE_SPECS: dict[str, dict[str, object]] = {
    "Figure 1": {"stem": "figure_1_information_atlas", "height_mm": 125.0},
    "Figure 2": {"stem": "figure_2_information_and_accuracy", "height_mm": 112.0},
    "Figure 3": {"stem": "figure_3_cea_starting_probability", "height_mm": 112.0},
    "Figure 4": {"stem": "figure_4_result_branch_entropy", "height_mm": 120.0},
    "Figure S1": {"stem": "figure_s1_corpus_and_selection_flow", "height_mm": 150.0},
    "Figure S2": {"stem": "figure_s2_validation_and_route_stability", "height_mm": 125.0},
    "Figure S3": {"stem": "figure_s3_information_distributions_and_youden", "height_mm": 122.0},
    "Figure S4": {"stem": "figure_s4_recovery_and_diagnostics", "height_mm": 122.0},
    "Figure S5": {"stem": "figure_s5_estimator_sensitivity", "height_mm": 122.0},
    "Figure S6": {"stem": "figure_s6_exploratory_contrasts_and_movement", "height_mm": 122.0},
    "Figure S7": {"stem": "figure_s7_near_ties_and_composition_audit", "height_mm": 118.0},
    "Figure S8": {"stem": "figure_s8_cea_crossing_and_theoretical_checks", "height_mm": 122.0},
}

DISPLAY_ORDER = (
    "Figure 1",
    "Figure 2",
    "Figure 3",
    "Figure 4",
    "Table 1",
    "Figure S1",
    "Figure S2",
    "Figure S3",
    "Figure S4",
    "Figure S5",
    "Figure S6",
    "Figure S7",
    "Figure S8",
)

DISPLAY_SOURCES: dict[str, tuple[str, ...]] = {
    "Figure 1": (
        "analysis/bmj_rmr_v042/derived_operating_points_standard_anchors.csv",
        "analysis/bmj_rmr_v042/atlas_landmarks.csv",
    ),
    "Figure 2": (
        "analysis/bmj_rmr_v042/derived_operating_points_standard_anchors.csv",
        "analysis/bmj_rmr_v042/theoretical_non_equivalence_examples.csv",
        "analysis/bmj_rmr_v042/theoretical_non_equivalence_summary.json",
    ),
    "Figure 3": (
        "analysis/bmj_rmr_v042/empirical_cea_probability_profiles.csv",
        "analysis/bmj_rmr_v042/empirical_cea_crossing_sensitivity.csv",
    ),
    "Figure 4": (
        "analysis/bmj_rmr_v042/branch_entropy_all_primary.csv",
        "analysis/bmj_rmr_v042/branch_entropy_summary.csv",
    ),
    "Table 1": ("analysis/bmj_rmr_v042/standard_anchor_summary.csv",),
    **{key: value for key, value in base.DISPLAY_SOURCES.items() if key.startswith("Figure S")},
}

DISPLAY_CLAIMS = {
    "Figure 1": "C001; C012; C013",
    "Figure 2": "C013; C014",
    "Figure 3": "C017",
    "Figure 4": "C016",
    "Table 1": "C012; C013; C016",
    **{key: value for key, value in base.DISPLAY_CLAIMS.items() if key.startswith("Figure S")},
}


def configure_base() -> None:
    base.VERSION = VERSION
    base.RELEASE_DATE = RELEASE_DATE
    base.FIGURE_SPECS = FIGURE_SPECS
    base.DISPLAY_ORDER = DISPLAY_ORDER
    base.DISPLAY_SOURCES = DISPLAY_SOURCES
    base.DISPLAY_CLAIMS = DISPLAY_CLAIMS


def figure_1(data: dict[str, object], grayscale: bool) -> plt.Figure:
    """Promote the validated v19 Atlas composition to Figure 1."""

    figure = base.figure_2(data["anchors"], data["landmarks"], grayscale)
    figure._suptitle.set_text("The information in diagnostic tests")
    return figure


def figure_2(data: dict[str, object], grayscale: bool) -> plt.Figure:
    fig, axes = base.new_figure(
        "Figure 2", grayscale, nrows=1, ncols=2, gridspec_kw={"width_ratios": [1.18, 0.82]}
    )
    palette = base.colors(grayscale)
    twenty = data["anchors"].loc[np.isclose(data["anchors"].starting_probability, 0.20)]
    axes[0].scatter(
        twenty.youden_j,
        twenty.information_bits,
        s=18,
        color=palette[0],
        edgecolor=base.WHITE,
        linewidth=0.35,
        alpha=0.70,
    )
    rho = float(data["anchor_summary"].loc[np.isclose(data["anchor_summary"].starting_probability, 0.20), "spearman_information_vs_youden_j"].iloc[0])
    axes[0].text(
        0.04,
        0.94,
        f"Spearman ρ = {rho:.3f}\nn = {len(twenty)} operating points",
        transform=axes[0].transAxes,
        va="top",
        fontsize=8.5,
        color=base.INK,
    )
    axes[0].set(
        xlabel="Youden's J",
        ylabel="Expected information at 20% (bits)",
        xlim=(0, 1.02),
        ylim=(0, max(0.74, float(twenty.information_bits.max()) * 1.04)),
    )
    base.panel_label(axes[0], "A", "Strong empirical association", "empirical")
    base.grid_both(axes[0])

    equal_j = data["theory_examples"].loc[
        data["theory_examples"].example_id.eq("equal_youden_different_information")
    ].sort_values("test_label")
    ypos = np.arange(len(equal_j))
    for index, (row, marker) in enumerate(zip(equal_j.itertuples(index=False), ("o", "s"))):
        axes[1].hlines(ypos[index], 0, row.information_bits, color=palette[index + 1], lw=5.0, alpha=0.28)
        axes[1].scatter(row.information_bits, ypos[index], marker=marker, s=56, color=palette[index + 1], zorder=3)
        axes[1].text(
            row.information_bits - 0.004,
            ypos[index],
            f"{row.information_bits:.3f} bits",
            va="center",
            ha="right",
            fontsize=8.5,
        )
    delta = float(data["theory_summary"]["equal_youden_different_information"]["absolute_information_difference_bits"])
    axes[1].set(
        yticks=ypos,
        yticklabels=[
            f"Test {row.test_label}\nSe {100 * row.sensitivity:.0f}%, Sp {100 * row.specificity:.0f}%"
            for row in equal_j.itertuples(index=False)
        ],
        xlabel="Expected information at 20% (bits)",
        xlim=(0, 0.225),
        ylim=(-0.55, 1.25),
    )
    axes[1].text(
        0.03,
        0.12,
        f"Both J = 0.60\nDifference = {delta:.6f} bits",
        transform=axes[1].transAxes,
        fontsize=8.5,
        color=base.MUTED,
    )
    base.panel_label(axes[1], "B", "Equal J, different information", "exact construction")
    base.grid_y(axes[1])
    fig.suptitle("Information is related to conventional accuracy without being identical", y=0.985)
    fig.subplots_adjust(left=0.085, right=0.985, bottom=0.18, top=0.77, wspace=0.34)
    return fig


def figure_3(data: dict[str, object], grayscale: bool) -> plt.Figure:
    fig, axes = base.new_figure("Figure 3", grayscale, nrows=1, ncols=2)
    palette = base.colors(grayscale)
    models = (
        ("v0.42.0 canonical REML", "Primary REML", 0.1280262761348617),
        ("v0.40.0/v0.41.0 regularized GH15", "Direct-binomial sensitivity", 0.38086130898547527),
    )
    threshold_styles = (
        ("CEA at 2.5µg/L", palette[0], "-", "o", "2.5 µg/L"),
        ("CEA at 5µg/L", palette[2], "--", "s", "5 µg/L"),
    )
    for axis, (model, title, crossing) in zip(axes, models):
        model_frame = data["cea_profiles"].loc[data["cea_profiles"].model_specification.eq(model)]
        for test, color, linestyle, marker, label in threshold_styles:
            frame = model_frame.loc[model_frame.test_name.eq(test)]
            axis.plot(
                100 * frame.starting_probability,
                frame.information_bits,
                color=color,
                ls=linestyle,
                lw=1.8,
                marker=marker,
                markevery=8,
                ms=3.3,
                label=label,
            )
            for probability in (0.05, 0.20):
                row = frame.loc[np.isclose(frame.starting_probability, probability)].iloc[0]
                axis.scatter(100 * probability, row.information_bits, color=color, marker=marker, s=28, zorder=4)
        axis.axvline(100 * crossing, color=palette[3], ls=":", lw=1.5)
        axis.text(
            100 * crossing + (0.8 if crossing < 0.2 else -1.2),
            0.018,
            f"Crossing {100 * crossing:.1f}%",
            rotation=90,
            va="bottom",
            ha="left" if crossing < 0.2 else "right",
            fontsize=8.0,
            color=palette[3],
        )
        axis.set(xlim=(0, 50), ylim=(0, 0.34), xlabel="Starting probability (%)", ylabel="Expected information (bits)")
        axis.legend(frameon=False, fontsize=8.2, loc="upper left")
        base.grid_both(axis)
        base.panel_label(axis, "A" if axis is axes[0] else "B", title, "empirical")
    fig.suptitle("Why starting probability matters: CEA thresholds for recurrent colorectal cancer", y=0.985)
    base.figure_footer(
        fig,
        "The 2.5 µg/L threshold is more sensitive and less specific. Crossings describe expected-information ordering; they are not action thresholds or clinical-superiority estimates.",
    )
    fig.subplots_adjust(left=0.08, right=0.985, bottom=0.22, top=0.75, wspace=0.27)
    return fig


def build_table_1(anchor_summary: pd.DataFrame) -> pd.DataFrame:
    rows: list[dict[str, str]] = []
    for row in anchor_summary.sort_values("starting_probability").itertuples(index=False):
        rows.append(
            {
                "Starting probability": f"{100 * row.starting_probability:.0f}%",
                "Starting entropy (bits)": f"{row.starting_entropy_bits:.3f}",
                "Median information, IQR (bits)": f"{row.information_bits_median:.3f} ({row.information_bits_q1:.3f}–{row.information_bits_q3:.3f})",
                "Median entropy removed, IQR": f"{100 * row.fraction_entropy_removed_median:.1f}% ({100 * row.fraction_entropy_removed_q1:.1f}–{100 * row.fraction_entropy_removed_q3:.1f}%)",
                "Spearman ρ with Youden's J": f"{row.spearman_information_vs_youden_j:.3f}",
            }
        )
    return pd.DataFrame.from_records(rows)


def build_captions(data: dict[str, object]) -> dict[str, str]:
    inherited = base.build_captions(data, base.build_table_2(data["profile"]))
    twenty = data["anchor_summary"].loc[np.isclose(data["anchor_summary"].starting_probability, 0.20)].iloc[0]
    canonical = data["cea_profiles"].loc[data["cea_profiles"].model_specification.eq("v0.42.0 canonical REML")]

    def cea_value(test: str, probability: float) -> float:
        return float(canonical.loc[canonical.test_name.eq(test) & np.isclose(canonical.starting_probability, probability), "information_bits"].iloc[0])

    inherited.update(
        {
            "Figure 1": (
                "Figure 1. The information in diagnostic tests at a 20% starting probability. "
                "(A) Expected-information contours in sensitivity-specificity space with all 273 primary source-defined pooled operating points from 210 reviews. "
                "(B) Complete distribution of the fraction of the 0.722 starting bits removed; the median was "
                f"{100 * float(twenty.fraction_entropy_removed_median):.1f}%. "
                "(C) Expected information versus post-negative probability for the same points. Four shape-coded clinical landmarks orient the scale and branch behaviour; they are not rankings of clinical utility. Primary estimator: continuity-corrected bivariate restricted maximum likelihood."
            ),
            "Figure 2": (
                "Figure 2. Expected information and conventional accuracy at a 20% starting probability. "
                f"(A) Across all 273 pooled operating points, mutual information and Youden's J had Spearman ρ={float(twenty.spearman_information_vs_youden_j):.3f}. "
                "(B) An exact construction gives both tests Youden's J=0.60 while expected information differs by 0.010414 bits: sensitivity/specificity 80%/80% gives 0.182 bits and 95%/65% gives 0.193 bits. "
                "The empirical association describes this sample; the construction shows that the measures are mathematically non-equivalent."
            ),
            "Figure 3": (
                "Figure 3. Starting probability changes the expected-information ordering of carcinoembryonic-antigen (CEA) thresholds for detecting recurrent colorectal cancer. "
                f"(A) Under the primary REML model, the 2.5 and 5 µg/L thresholds supplied {cea_value('CEA at 2.5µg/L', 0.05):.3f} and {cea_value('CEA at 5µg/L', 0.05):.3f} bits at a 5% starting probability and crossed at 12.8%; at 20%, they supplied {cea_value('CEA at 2.5µg/L', 0.20):.3f} and {cea_value('CEA at 5µg/L', 0.20):.3f} bits. "
                "(B) The regularised direct-binomial sensitivity analysis crossed at 38.1%. The 2.5 µg/L threshold was more sensitive and less specific. Youden's J remained 0.60 and 0.56 because it does not depend on starting probability. Neither crossing is an action threshold or a clinical-superiority estimate."
            ),
            "Figure 4": inherited["Figure 4"],
            "Table 1": (
                "Table 1. Diagnostic information across stated starting probabilities. Each row contains 273 source-defined pooled operating points from 210 reviews. Information and the fraction of starting entropy removed are medians with interquartile ranges (IQR). Primary estimator: continuity-corrected bivariate restricted maximum likelihood."
            ),
        }
    )
    return inherited


def write_table(manuscript_dir: Path, table: pd.DataFrame, caption: str) -> list[Path]:
    directory = manuscript_dir / "tables" / f"v{VERSION}"
    directory.mkdir(parents=True, exist_ok=True)
    stem = "table_1_diagnostic_information"
    footnote = "IQR = interquartile range. Starting probabilities are standardised analytical anchors, not prevalence estimates or action thresholds."
    csv_path = directory / f"{stem}.csv"
    md_path = directory / f"{stem}.md"
    docx_path = directory / f"{stem}.docx"
    base.write_csv(csv_path, table)
    md_path.write_text(f"{caption}\n\n{base.markdown_table(table)}\n\n*{footnote}*\n", encoding="utf-8", newline="\n")
    base.write_docx_table(docx_path, caption, table, footnote)
    captions_path = directory / "TABLE_CAPTIONS.md"
    captions_path.write_text(f"# Table captions\n\n{caption}\n", encoding="utf-8", newline="\n")
    return [csv_path, md_path, docx_path, captions_path]


def write_caption_files(manuscript_dir: Path, output_analysis: Path, captions: dict[str, str]) -> list[Path]:
    figures_dir = manuscript_dir / "figures" / f"v{VERSION}"
    figures_dir.mkdir(parents=True, exist_ok=True)
    lines = ["# Figure captions"]
    records: list[dict[str, str]] = []
    for display in DISPLAY_ORDER:
        if not display.startswith("Figure"):
            continue
        lines.extend(("", f"## {display}", "", captions[display], "", f"Sources: {'; '.join(DISPLAY_SOURCES[display])}", "", f"Claims: {DISPLAY_CLAIMS[display]}"))
        records.append({"display": display, "caption": captions[display], "source_files": "; ".join(DISPLAY_SOURCES[display]), "claim_ids": DISPLAY_CLAIMS[display]})
    md_path = figures_dir / "FIGURE_CAPTIONS.md"
    csv_path = output_analysis / "figure_captions.csv"
    md_path.write_text("\n".join(lines) + "\n", encoding="utf-8", newline="\n")
    base.write_csv(csv_path, pd.DataFrame.from_records(records))
    return [md_path, csv_path]


def write_display_manifest(root: Path, manuscript_dir: Path, output_analysis: Path, captions: dict[str, str]) -> Path:
    rows: list[dict[str, object]] = []
    for order, display in enumerate(DISPLAY_ORDER, start=1):
        if display.startswith("Figure"):
            spec = FIGURE_SPECS[display]
            outputs = "; ".join(
                (manuscript_dir / "figures" / f"v{VERSION}" / f"{spec['stem']}{gray}.{extension}").relative_to(root).as_posix()
                for gray in ("", "_grayscale")
                for extension in ("pdf", "svg", "png")
            )
            dimensions = f"180 × {float(spec['height_mm']):.0f} mm"
        else:
            outputs = "; ".join(
                (manuscript_dir / "tables" / f"v{VERSION}" / f"table_1_diagnostic_information.{extension}").relative_to(root).as_posix()
                for extension in ("csv", "md", "docx")
            )
            dimensions = "editable table; not rasterised"
        rows.append(
            {
                "order": order,
                "display": display,
                "source_files": "; ".join(DISPLAY_SOURCES[display]),
                "claim_ids": DISPLAY_CLAIMS[display],
                "outputs": outputs,
                "final_dimensions": dimensions,
                "caption": captions[display],
                "classification_encoding": "empirical, theoretical, or sensitivity status stated in panel labels and captions",
                "redundant_encoding": "colour plus markers, line styles, hatching, or direct labels",
            }
        )
    path = output_analysis / "display_manifest.csv"
    base.write_csv(path, pd.DataFrame.from_records(rows))
    return path


def build(root: Path, output_analysis: Path, manuscript_dir: Path, qa_dir: Path) -> None:
    configure_base()
    data = base.load_data(root)
    figures_dir = manuscript_dir / "figures" / f"v{VERSION}"
    figures_dir.mkdir(parents=True, exist_ok=True)
    output_analysis.mkdir(parents=True, exist_ok=True)
    qa_dir.mkdir(parents=True, exist_ok=True)
    builders = {
        "Figure 1": lambda gray: figure_1(data, gray),
        "Figure 2": lambda gray: figure_2(data, gray),
        "Figure 3": lambda gray: figure_3(data, gray),
        "Figure 4": lambda gray: base.figure_4(data["branches"], data["branch_summary"], gray),
        "Figure S1": lambda gray: base.figure_s1(data["run_manifest"], data["trace_summary"], data["exceptions"], gray),
        "Figure S2": lambda gray: base.figure_s2(data["fidelity"], data["audit_summary"], data["routes"], gray),
        "Figure S3": lambda gray: base.figure_s3(data["anchors"], data["anchor_summary"], gray),
        "Figure S4": lambda gray: base.figure_s4(data["recovery"], data["recovery_summary"], data["diagnostics"], data["diagnostic_summary"], gray),
        "Figure S5": lambda gray: base.figure_s5(data["sensitivity"], data["sensitivity_summary"], data["estimator_effect"], gray),
        "Figure S6": lambda gray: base.figure_s6(data["contrasts"], data["movement"], gray),
        "Figure S7": lambda gray: base.figure_s7(data["near_ties"], data["restricted"], gray),
        "Figure S8": lambda gray: base.figure_s8(data["crossings"], data["cea_profiles"], data["theory_summary"], gray),
    }
    figure_paths: list[Path] = []
    for display in FIGURE_SPECS:
        for grayscale in (False, True):
            figure_paths.extend(base.save_figure(builders[display](grayscale), figures_dir, display, grayscale))

    captions = build_captions(data)
    table_paths = write_table(manuscript_dir, build_table_1(data["anchor_summary"]), captions["Table 1"])
    caption_paths = write_caption_files(manuscript_dir, output_analysis, captions)
    display_source_paths = base.write_display_sources(output_analysis, data)
    cea_source = output_analysis / "display_sources/figure_3_cea_profiles.csv"
    base.write_csv(
        cea_source,
        data["cea_profiles"].loc[
            data["cea_profiles"].model_specification.isin(("v0.42.0 canonical REML", "v0.40.0/v0.41.0 regularized GH15"))
        ],
    )
    display_source_paths.append(cea_source)
    display_manifest = write_display_manifest(root, manuscript_dir, output_analysis, captions)
    bounds_paths = base.audit_pdf_text_bounds(figures_dir, qa_dir)
    previews = base.render_pdf_previews(figures_dir, qa_dir)
    contact_paths = base.write_contact_sheets(previews, qa_dir)
    visual_json = base.initial_visual_log(qa_dir, previews)
    visual_md = base.write_visual_markdown(qa_dir)
    visual_text = visual_md.read_text(encoding="utf-8").replace("analysis/bmj_rmr_v042", "analysis/bmj_rmr_v100")
    visual_md.write_text("\n".join(line for line in visual_text.splitlines() if "collision_contact_sheet" not in line) + "\n", encoding="utf-8", newline="\n")
    source_manifest = output_analysis / "source_hash_manifest.csv"
    base.write_csv(source_manifest, base.source_hash_rows(root))
    artifacts = [
        *figure_paths,
        *table_paths,
        *caption_paths,
        *display_source_paths,
        display_manifest,
        *bounds_paths,
        *previews,
        *contact_paths,
        visual_json,
        visual_md,
        source_manifest,
    ]
    base.write_csv(output_analysis / "display_artifact_manifest.csv", base.artifact_rows(root, artifacts))


def parse_args() -> argparse.Namespace:
    parser = argparse.ArgumentParser(description=__doc__)
    parser.add_argument("--root", type=Path, default=Path(__file__).resolve().parents[1])
    parser.add_argument("--output-analysis", type=Path)
    parser.add_argument("--manuscript-dir", type=Path)
    parser.add_argument("--qa-dir", type=Path)
    return parser.parse_args()


if __name__ == "__main__":
    arguments = parse_args()
    repository = arguments.root.resolve()
    build(
        repository,
        (arguments.output_analysis or repository / "analysis/bmj_rmr_v100").resolve(),
        (arguments.manuscript_dir or repository / "docs/manuscript/v20").resolve(),
        (arguments.qa_dir or repository / "analysis/bmj_rmr_v100/qa").resolve(),
    )
```
