## Supplementary material for "The information in diagnostic tests": Executable analysis code, derived inputs, source-acquisition records, tests, and release metadata: bmj_rmr_v190_visuals.py

```
"""Build the Information EBM 1.9.0 figures from the locked v0.42 analysis."""

from __future__ import annotations

import argparse
from pathlib import Path

import matplotlib as mpl
import matplotlib.pyplot as plt
import matplotlib.patheffects as path_effects
import networkx as nx
import numpy as np
import pandas as pd
from matplotlib.patches import FancyBboxPatch

from scripts import bmj_rmr_v042_visuals as base
from scripts import bmj_rmr_v100_visuals as previous


VERSION = "1.9.0"
RELEASE_DATE = "2026-08-29"

FIGURE_SPECS: dict[str, dict[str, object]] = {
    "Figure 1": {"stem": "figure_1_information_reference_scale", "height_mm": 142.0},
    "Figure 2": {"stem": "figure_2_information_and_accuracy", "height_mm": 124.0},
    "Figure 3": {"stem": "figure_3_cea_clinical_case", "height_mm": 112.0},
    "Figure 4": {"stem": "figure_4_observed_result_pathway", "height_mm": 132.0},
    "Figure S1": {"stem": "figure_s1_corpus_and_selection_flow", "height_mm": 150.0},
    **{key: value for key, value in previous.FIGURE_SPECS.items() if key.startswith("Figure S") and key != "Figure S1"},
    "Figure S9": {"stem": "figure_s9_result_branch_distributions", "height_mm": 120.0},
}

DISPLAY_ORDER = (
    "Figure 1", "Figure 2", "Figure 3", "Figure 4", "Table 1",
    "Figure S1", "Figure S2", "Figure S3", "Figure S4", "Figure S5",
    "Figure S6", "Figure S7", "Figure S8", "Figure S9",
)

DISPLAY_SOURCES: dict[str, tuple[str, ...]] = {
    "Figure 1": (
        "analysis/bmj_rmr_v042/standard_anchor_summary.csv",
        "analysis/bmj_rmr_v042/derived_operating_points_standard_anchors.csv",
        "analysis/bmj_rmr_v042/derived_operating_points_probability_profiles.csv",
        "analysis/bmj_rmr_v042/atlas_landmarks.csv",
    ),
    "Figure 2": (
        "analysis/bmj_rmr_v042/derived_operating_points_standard_anchors.csv",
        "analysis/bmj_rmr_v190/equal_youden_candidate_pairs.csv",
    ),
    "Figure 3": (
        "analysis/bmj_rmr_v042/empirical_cea_probability_profiles.csv",
        "analysis/bmj_rmr_v042/empirical_cea_crossing_sensitivity.csv",
    ),
    "Figure 4": (
        "analysis/bmj_rmr_v042/atlas_landmarks.csv",
        "analysis/bmj_rmr_v042/branch_entropy_all_primary.csv",
    ),
    "Table 1": ("analysis/bmj_rmr_v042/standard_anchor_summary.csv",),
    "Figure S1": (
        "analysis/full_atlas/v9/source_role_manifest.csv",
        "analysis/bmj_rmr_v042/source_trace_all_primary.csv",
        "analysis/bmj_rmr_v042/source_trace_summary.json",
        "analysis/bmj_rmr_v042/analysis_row_selection_exceptions.csv",
    ),
    **{key: value for key, value in previous.DISPLAY_SOURCES.items() if key.startswith("Figure S") and key != "Figure S1"},
    "Figure S9": (
        "analysis/bmj_rmr_v042/branch_entropy_all_primary.csv",
        "analysis/bmj_rmr_v042/branch_entropy_summary.csv",
    ),
}

DISPLAY_CLAIMS = {
    "Figure 1": "C001; C012; C013",
    "Figure 2": "C013; C014; C036",
    "Figure 3": "C017",
    "Figure 4": "C030",
    "Table 1": "C012; C013; C016",
    "Figure S1": "C001; C002; C003; C004",
    **{key: value for key, value in previous.DISPLAY_CLAIMS.items() if key.startswith("Figure S") and key != "Figure S1"},
    "Figure S9": "C016; C031",
}


def configure_base() -> None:
    base.VERSION = VERSION
    base.RELEASE_DATE = RELEASE_DATE
    base.FIGURE_SPECS = FIGURE_SPECS
    base.DISPLAY_ORDER = DISPLAY_ORDER
    base.DISPLAY_SOURCES = DISPLAY_SOURCES
    base.DISPLAY_CLAIMS = DISPLAY_CLAIMS
    previous.VERSION = VERSION
    previous.FIGURE_SPECS = FIGURE_SPECS
    previous.DISPLAY_ORDER = DISPLAY_ORDER
    previous.DISPLAY_SOURCES = DISPLAY_SOURCES
    previous.DISPLAY_CLAIMS = DISPLAY_CLAIMS


def entropy(probability: np.ndarray | float) -> np.ndarray:
    values = np.clip(np.asarray(probability, dtype=float), 1e-12, 1 - 1e-12)
    return -values * np.log2(values) - (1 - values) * np.log2(1 - values)


def information_percent(prior: float, sensitivity: np.ndarray, specificity: np.ndarray) -> np.ndarray:
    p_positive = prior * sensitivity + (1 - prior) * (1 - specificity)
    p_negative = 1 - p_positive
    post_positive = prior * sensitivity / np.clip(p_positive, 1e-12, None)
    post_negative = prior * (1 - sensitivity) / np.clip(p_negative, 1e-12, None)
    remaining = p_positive * entropy(post_positive) + p_negative * entropy(post_negative)
    return 100 * (float(entropy(prior)) - remaining) / float(entropy(prior))


def profile_percentiles(profiles: pd.DataFrame) -> pd.DataFrame:
    counts = profiles.groupby("starting_probability").operating_point_id.nunique()
    assert len(counts) == 50 and counts.eq(273).all()
    summary = (
        profiles.groupby("starting_probability").fraction_entropy_removed
        .quantile([0.10, 0.25, 0.50, 0.75, 0.90])
        .unstack()
        .rename(columns={0.10: "p10", 0.25: "p25", 0.50: "median", 0.75: "p75", 0.90: "p90"})
        .reset_index()
    )
    return summary


def hints_profiles(anchors: pd.DataFrame) -> pd.DataFrame:
    names = ("HINTS - subcomponent head impulse", "HINTS - subcomponent nystagmus")
    frame = anchors.loc[anchors.review_id.eq("CD015089") & anchors.test_name.isin(names)].copy()
    assert len(frame) == 6 and frame.group_id.nunique() == 2
    return frame


def rank_near_equal_youden_pairs(anchors: pd.DataFrame) -> pd.DataFrame:
    wide = anchors.pivot_table(
        index=["review_id", "review_title", "group_id", "test_name", "sensitivity", "specificity", "youden_j", "study_rows"],
        columns="starting_probability",
        values="fraction_entropy_removed",
    ).reset_index()
    rows: list[dict[str, object]] = []
    for (review_id, review_title), group in wide.groupby(["review_id", "review_title"]):
        records = list(group.to_dict("records"))
        for left_index, left in enumerate(records):
            for right in records[left_index + 1 :]:
                if (left["sensitivity"] - right["sensitivity"]) * (left["specificity"] - right["specificity"]) >= 0:
                    continue
                gaps = {prior: abs(float(left[prior]) - float(right[prior])) for prior in (0.05, 0.20, 0.50)}
                rows.append({
                    "review_id": review_id,
                    "review_title": review_title,
                    "test_a": left["test_name"],
                    "group_a": left["group_id"],
                    "sensitivity_a": left["sensitivity"],
                    "specificity_a": left["specificity"],
                    "youden_j_a": left["youden_j"],
                    "test_b": right["test_name"],
                    "group_b": right["group_id"],
                    "sensitivity_b": right["sensitivity"],
                    "specificity_b": right["specificity"],
                    "youden_j_b": right["youden_j"],
                    "absolute_j_difference": abs(float(left["youden_j"]) - float(right["youden_j"])),
                    "information_difference_5": gaps[0.05],
                    "information_difference_20": gaps[0.20],
                    "information_difference_50": gaps[0.50],
                    "maximum_information_difference": max(gaps.values()),
                })
    ranked = pd.DataFrame.from_records(rows).sort_values(
        ["absolute_j_difference", "maximum_information_difference"], ascending=[True, False]
    ).reset_index(drop=True)
    ranked.insert(0, "candidate_rank", np.arange(1, len(ranked) + 1))
    selected = (
        ranked.review_id.eq("CD015089")
        & ranked.test_a.str.contains("head impulse|nystagmus", regex=True)
        & ranked.test_b.str.contains("head impulse|nystagmus", regex=True)
    )
    assert selected.sum() == 1
    ranked.insert(1, "selected_for_main_figure", selected)
    ranked.insert(2, "selection_note", np.where(selected, "near-equal J; same review and target; large information gap; familiar emergency-medicine context", "candidate retained for audit"))
    return ranked


def figure_1(data: dict[str, object], grayscale: bool) -> plt.Figure:
    fig, ax = base.new_figure("Figure 1", grayscale)
    palette = base.colors(grayscale)
    profiles = data["probability_profiles"]
    percentiles = profile_percentiles(profiles)
    x = 100 * percentiles.starting_probability.to_numpy()
    neutral_light = "#E8E8E8" if grayscale else "#DCE8F1"
    neutral_mid = "#B8B8B8" if grayscale else "#9FC1D8"
    median_color = "#111111" if grayscale else "#005EA8"

    ax.fill_between(x, 100 * percentiles.p10, 100 * percentiles.p90, color=neutral_light, lw=0, label="10th–90th percentiles")
    ax.fill_between(x, 100 * percentiles.p25, 100 * percentiles.p75, color=neutral_mid, lw=0, label="25th–75th percentiles")
    ax.plot(x, 100 * percentiles["median"], color=median_color, lw=2.6, label="Median", zorder=3)

    roles = (
        ("Ottawa ankle rule", "Ottawa ankle rule", palette[0], "-", "o", 41, -3.2),
        ("Lung ultrasound pneumonia", "Lung ultrasound for pneumonia", palette[1], "--", "s", 43, 2.4),
        ("Non-contrast CT appendicitis", "Non-contrast CT for appendicitis", palette[2], "-.", "^", 31, 4.0),
        ("CT angiography intracranial lesion", "CT angiography:\nintracranial lesion", palette[3], ":", "D", 44, -5.0),
    )
    for label, display_label, color, linestyle, marker, label_x, offset in roles:
        landmark = data["landmarks"].loc[data["landmarks"].label.eq(label)].iloc[0]
        frame = profiles.loc[profiles.group_id.eq(landmark.group_id)].sort_values("starting_probability")
        ax.plot(100 * frame.starting_probability, 100 * frame.fraction_entropy_removed, color=color, ls=linestyle, lw=1.8, marker=marker, markevery=9, ms=3.5, zorder=4)
        point = frame.iloc[(100 * frame.starting_probability - label_x).abs().argmin()]
        ax.text(label_x, 100 * point.fraction_entropy_removed + offset, display_label, color=color, fontsize=7.3, va="center", ha="center", weight="bold", bbox={"facecolor": base.WHITE, "edgecolor": "none", "alpha": 0.82, "pad": 1.2}, zorder=6)

    reference_probabilities = (0.05, 0.10, 0.20, 0.30, 0.40, 0.50)
    label_offsets = (4.2, -5.0, 4.2, -5.0, 4.2, -5.0)
    for prior, offset in zip(reference_probabilities, label_offsets):
        row = percentiles.iloc[(percentiles.starting_probability - prior).abs().argmin()]
        value = 100 * row["median"]
        ax.scatter(100 * prior, value, s=31, facecolor=base.WHITE, edgecolor=median_color, linewidth=1.3, zorder=7)
        ax.text(100 * prior, value + offset, f"{value:.1f}%", color=median_color, fontsize=7.3, weight="bold", ha="center", va="center", zorder=7)

    ax.set(xlim=(1, 50.8), ylim=(0, 82), xlabel="Starting disease probability (%)", ylabel="Starting diagnostic uncertainty resolved (%)")
    ax.set_xticks([1, 5, 10, 20, 30, 40, 50])
    ax.set_yticks([0, 20, 40, 60, 80])
    ax.grid(axis="y", color=base.GRID, lw=0.55)
    ax.legend(frameon=False, loc="upper left", fontsize=7.6, ncol=3, handlelength=2.5, columnspacing=1.2)

    fig.suptitle("Empirical reference map of diagnostic information across starting probabilities", y=0.985)
    base.figure_footer(fig, "Bands summarise all 273 pooled estimates; direct labels show four source-traced clinical profiles.", y=0.025)
    fig.subplots_adjust(left=0.085, right=0.975, bottom=0.17, top=0.84)
    return fig


def figure_2(data: dict[str, object], grayscale: bool) -> plt.Figure:
    base.set_style(grayscale)
    fig = plt.figure(figsize=base.figure_size(float(FIGURE_SPECS["Figure 2"]["height_mm"])))
    grid_spec = fig.add_gridspec(2, 3, height_ratios=[1.0, 0.06], hspace=0.30, wspace=0.20)
    map_axes = [fig.add_subplot(grid_spec[0, index]) for index in range(3)]
    colorbar_axis = fig.add_subplot(grid_spec[1, :])
    palette = base.colors(grayscale)
    anchors = data["anchors"]
    hints = hints_profiles(anchors)
    priors = (0.05, 0.20, 0.50)
    grid = np.linspace(0.001, 0.999, 220)
    sensitivity, specificity = np.meshgrid(grid, grid)
    levels = np.arange(0, 101, 10)
    contour_set = None
    for index, (axis, prior) in enumerate(zip(map_axes, priors), start=1):
        removed = information_percent(prior, sensitivity, specificity)
        contour_set = axis.contourf(100 * sensitivity, 100 * specificity, removed, levels=levels, cmap="Greys" if grayscale else "cividis", vmin=0, vmax=100)
        frame = anchors.loc[np.isclose(anchors.starting_probability, prior)]
        axis.scatter(100 * frame.sensitivity, 100 * frame.specificity, s=8, facecolor=base.WHITE, edgecolor=base.INK, linewidth=0.35, alpha=0.78, zorder=4)
        for youden, linestyle in zip((0.20, 0.40, 0.60, 0.80), (":", "--", "-.", "-")):
            sens_line = np.linspace(youden, 1, 100)
            spec_line = 1 + youden - sens_line
            line = axis.plot(100 * sens_line, 100 * spec_line, color=base.WHITE, ls=linestyle, lw=1.1, zorder=5)[0]
            line.set_path_effects([path_effects.Stroke(linewidth=2.0, foreground="#222222"), path_effects.Normal()])
            label_sensitivity = 0.93
            label_specificity = 1 + youden - label_sensitivity
            if 0.03 < label_specificity < 0.97:
                text = axis.text(100 * label_sensitivity, 100 * label_specificity, f"J={youden:.1f}", color=base.WHITE, fontsize=6.4, ha="center", va="center", rotation=-34, zorder=6)
                text.set_path_effects([path_effects.Stroke(linewidth=1.7, foreground="#222222"), path_effects.Normal()])
        if np.isclose(prior, 0.05):
            pair = hints.loc[np.isclose(hints.starting_probability, prior)].sort_values("test_name")
            label_specs = (("Head impulse", "o", -25.0, -10.5), ("Nystagmus", "s", 2.5, -8.0))
            for row, (short, marker, dx, dy) in zip(pair.itertuples(index=False), label_specs):
                x_point, y_point = 100 * row.sensitivity, 100 * row.specificity
                axis.scatter(x_point, y_point, s=64, marker=marker, facecolor=palette[3], edgecolor=base.WHITE, linewidth=1.2, zorder=7)
                axis.plot([x_point, x_point + dx * 0.72], [y_point, y_point + dy * 0.72], color=palette[3], lw=0.9, zorder=7)
                axis.text(x_point + dx, y_point + dy, f"{short}\nJ={row.youden_j:.3f} | {100 * row.fraction_entropy_removed:.1f}%", fontsize=6.9, weight="bold", color=base.INK, ha="left", va="center", bbox={"facecolor": base.WHITE, "edgecolor": palette[3], "linewidth": 0.7, "alpha": 0.94, "pad": 1.8}, zorder=8)
        axis.set(xlim=(0, 100), ylim=(0, 100), xlabel="Sensitivity (%)")
        if index == 1:
            axis.set_ylabel("Specificity (%)")
        else:
            axis.tick_params(axis="y", labelleft=False)
        axis.set_aspect("equal")
        base.panel_label(axis, chr(64 + index), f"Starting probability {100 * prior:.0f}%", "all 273 pooled estimates")
    assert contour_set is not None
    colorbar = fig.colorbar(contour_set, cax=colorbar_axis, orientation="horizontal")
    colorbar.set_label("Starting diagnostic uncertainty resolved (%)", fontsize=7.4)
    colorbar.ax.tick_params(labelsize=6.8)

    fig.suptitle("What does diagnostic information add beyond sensitivity, specificity, and Youden's J?", y=0.987)
    fig.subplots_adjust(left=0.07, right=0.975, bottom=0.13, top=0.84)
    return fig


def figure_3(data: dict[str, object], grayscale: bool) -> plt.Figure:
    fig, axes = base.new_figure("Figure 3", grayscale, nrows=1, ncols=2)
    palette = base.colors(grayscale)
    models = (
        ("v0.42.0 canonical REML", "Primary analysis", 0.1280262761348617),
        ("v0.40.0/v0.41.0 regularized GH15", "Alternative synthesis model", 0.38086130898547527),
    )
    styles = (("CEA at 2.5µg/L", palette[0], "-", "o", "2.5 µg/L"), ("CEA at 5µg/L", palette[2], "--", "s", "5 µg/L"))
    for index, (axis, (model, title, crossing)) in enumerate(zip(axes, models)):
        model_frame = data["cea_profiles"].loc[data["cea_profiles"].model_specification.eq(model)]
        for test, color, linestyle, marker, label in styles:
            frame = model_frame.loc[model_frame.test_name.eq(test)]
            axis.plot(100 * frame.starting_probability, 100 * frame.fraction_entropy_removed, color=color, ls=linestyle, lw=1.8, marker=marker, markevery=8, ms=3.1, label=label)
            for probability in (0.05, 0.20):
                row = frame.loc[np.isclose(frame.starting_probability, probability)].iloc[0]
                axis.scatter(100 * probability, 100 * row.fraction_entropy_removed, color=color, marker=marker, s=26, zorder=4)
        axis.axvline(100 * crossing, color=palette[3], ls=":", lw=1.4)
        axis.text(100 * crossing + (-0.8 if index else 0.8), 3, f"Crossing {100 * crossing:.1f}%", rotation=90, va="bottom", ha="right" if index else "left", fontsize=7.8, color=palette[3])
        for anchor in (5, 20):
            axis.axvline(anchor, color=base.GRID, lw=0.7, zorder=0)
        axis.set(xlim=(1, 50), ylim=(0, 40), xlabel="Recurrence probability before testing (%)", ylabel="Starting uncertainty removed (%)")
        axis.legend(frameon=False, fontsize=8.0, loc="upper left")
        base.panel_label(axis, "A" if index == 0 else "B", title, "empirical")
        base.grid_both(axis)
    fig.suptitle("How can CEA thresholds change information ordering as recurrence probability changes?", y=0.985)
    base.figure_footer(fig, "The lower threshold favours sensitivity; the higher threshold favours specificity.", y=0.03)
    fig.subplots_adjust(left=0.08, right=0.985, bottom=0.22, top=0.75, wspace=0.27)
    return fig


def ottawa_row(data: dict[str, object]) -> pd.Series:
    landmark = data["landmarks"].loc[data["landmarks"].label.eq("Ottawa ankle rule")].iloc[0]
    return data["branches"].loc[data["branches"].group_id.eq(landmark.group_id) & np.isclose(data["branches"].starting_probability, 0.05)].iloc[0]


def build_table_1(anchors: pd.DataFrame) -> pd.DataFrame:
    rows: list[dict[str, str]] = []
    for prior in (0.05, 0.20, 0.50):
        values = anchors.loc[np.isclose(anchors.starting_probability, prior), "fraction_entropy_removed"]
        assert len(values) == 273
        quantiles = values.quantile([0.10, 0.25, 0.50, 0.75, 0.90])
        row = {
            "Starting disease probability": f"{100 * prior:.0f}%",
            "Median uncertainty resolved, % (IQR)": f"{100 * quantiles.loc[0.50]:.1f}% ({100 * quantiles.loc[0.25]:.1f}%–{100 * quantiles.loc[0.75]:.1f}%)",
            "10th–90th percentile range, %": f"{100 * quantiles.loc[0.10]:.1f}%–{100 * quantiles.loc[0.90]:.1f}%",
        }
        rows.append(row)
    table = pd.DataFrame.from_records(rows)
    assert table.iloc[:, 1].tolist() == ["23.3% (17.3%–37.6%)", "29.2% (22.1%–45.6%)", "31.4% (23.3%–48.3%)"]
    return table


def write_table(manuscript_dir: Path, table: pd.DataFrame, caption: str) -> list[Path]:
    directory = manuscript_dir / "tables" / f"v{VERSION}"
    directory.mkdir(parents=True, exist_ok=True)
    stem = "table_1_diagnostic_information"
    footnote = "IQR is the 25th–75th percentile. Every summary uses the same 273 pooled diagnostic estimates from 210 reviews. Starting probabilities are standardised analytical conditions, not estimates of disease prevalence or clinical action thresholds."
    csv_path = directory / f"{stem}.csv"
    md_path = directory / f"{stem}.md"
    docx_path = directory / f"{stem}.docx"
    base.write_csv(csv_path, table)
    md_path.write_text(f"{caption}\n\n{base.markdown_table(table)}\n\n*{footnote}*\n", encoding="utf-8", newline="\n")
    base.write_docx_table(docx_path, caption, table, footnote)
    captions_path = directory / "TABLE_CAPTIONS.md"
    captions_path.write_text(f"# Table captions\n\n{caption}\n", encoding="utf-8", newline="\n")
    return [csv_path, md_path, docx_path, captions_path]


def figure_4(data: dict[str, object], grayscale: bool) -> plt.Figure:
    base.set_style(grayscale)
    fig = plt.figure(figsize=base.figure_size(float(FIGURE_SPECS["Figure 4"]["height_mm"])))
    grid = fig.add_gridspec(2, 1, height_ratios=[3.2, 1.0], hspace=0.12)
    graph_axis = fig.add_subplot(grid[0, 0])
    summary_axis = fig.add_subplot(grid[1, 0])
    palette = base.colors(grayscale)
    row = ottawa_row(data)
    increase_color = "#666666" if grayscale else "#D97706"
    decrease_color = "#222222" if grayscale else "#0072B2"
    start_color = "#AAAAAA" if grayscale else "#2A9D8F"

    graph = nx.DiGraph()
    graph.add_node("before", layer=0)
    graph.add_node("positive", layer=1)
    graph.add_node("negative", layer=1)
    graph.add_edges_from((("before", "positive"), ("before", "negative")))
    positions = nx.multipartite_layout(graph, subset_key="layer", align="vertical", scale=1.0)
    positions["positive"][1] = 0.72
    positions["negative"][1] = -0.72
    graph_axis.axis("off")
    graph_axis.set(xlim=(-1.45, 1.45), ylim=(-1.22, 1.22))

    nx.draw_networkx_nodes(graph, positions, nodelist=["before"], node_size=10500, node_shape="s", node_color=[base.WHITE], edgecolors=[start_color], linewidths=2.2, ax=graph_axis)
    nx.draw_networkx_nodes(graph, positions, nodelist=["positive"], node_size=11200, node_shape="s", node_color=[base.WHITE], edgecolors=[increase_color], linewidths=2.2, ax=graph_axis)
    nx.draw_networkx_nodes(graph, positions, nodelist=["negative"], node_size=11200, node_shape="s", node_color=[base.WHITE], edgecolors=[decrease_color], linewidths=2.2, ax=graph_axis)
    nx.draw_networkx_edges(graph, positions, edgelist=[("before", "positive")], width=[1.8 + 5.5 * row.positive_result_probability], edge_color=[increase_color], style="dashed", arrows=True, arrowsize=18, connectionstyle="arc3,rad=0.04", ax=graph_axis)
    nx.draw_networkx_edges(graph, positions, edgelist=[("before", "negative")], width=[1.8 + 5.5 * row.negative_result_probability], edge_color=[decrease_color], style="solid", arrows=True, arrowsize=18, connectionstyle="arc3,rad=-0.04", ax=graph_axis)
    nx.draw_networkx_labels(
        graph,
        positions,
        labels={
            "before": f"BEFORE TESTING\n\nFracture  5.0%\n\nUncertainty  {row.starting_entropy_bits:.3f} bits",
            "positive": f"POSITIVE RESULT\n\nFracture  {100 * row.post_positive_probability:.1f}%\n\nUncertainty  +{row.positive_entropy_change_bits:.3f} bits\n(increases)",
            "negative": f"NEGATIVE RESULT\n\nFracture  {100 * row.post_negative_probability:.1f}%\n\nUncertainty  {row.negative_entropy_change_bits:.3f} bits\n(decreases)",
        },
        font_size=7.8,
        font_color=base.INK,
        font_weight="bold",
        ax=graph_axis,
    )
    nx.draw_networkx_edge_labels(
        graph,
        positions,
        edge_labels={
            ("before", "positive"): f"Positive result  {100 * row.positive_result_probability:.1f}%",
            ("before", "negative"): f"Negative result  {100 * row.negative_result_probability:.1f}%",
        },
        font_size=7.5,
        font_color=base.INK,
        label_pos=0.53,
        rotate=False,
        bbox={"facecolor": base.WHITE, "edgecolor": "none", "alpha": 0.9, "pad": 1.2},
        ax=graph_axis,
    )
    graph_axis.set(xlim=(-1.45, 1.45), ylim=(-1.22, 1.22))

    contributions = np.array([row.positive_information_contribution_bits, row.negative_information_contribution_bits])
    shares = 100 * contributions / contributions.sum()
    summary_axis.barh([0], [shares[0]], color=increase_color, height=0.42, label="Positive-result contribution")
    summary_axis.barh([0], [shares[1]], left=[shares[0]], color=decrease_color, height=0.42, label="Negative-result contribution")
    summary_axis.text(shares[0] / 2, 0, f"Positive\n{shares[0]:.1f}%", ha="center", va="center", color=base.WHITE, fontsize=7.3, weight="bold")
    summary_axis.text(shares[0] + shares[1] / 2, 0, f"Negative\n{shares[1]:.1f}%", ha="center", va="center", color=base.WHITE, fontsize=7.3, weight="bold")
    summary_axis.set(xlim=(0, 100), ylim=(-0.52, 0.72), yticks=[], xticks=[])
    summary_axis.spines[:].set_visible(False)
    summary_axis.set_title(f"Together, both possible results remove {100 * row.information_bits / row.starting_entropy_bits:.1f}% of starting uncertainty on average ({row.information_bits:.3f} bits)", fontsize=8.6, weight="bold", pad=3)
    summary_axis.set_xlabel("Share of expected diagnostic information", fontsize=7.3, labelpad=1)
    fig.suptitle("One observed result changes one patient's uncertainty\nDiagnostic information averages both possible results", y=0.985, linespacing=1.15)
    base.figure_footer(fig, "Ottawa ankle rule | source-traced pooled estimate from 16 studies", y=0.018)
    fig.subplots_adjust(left=0.035, right=0.965, bottom=0.10, top=0.82)
    return fig


def figure_s1(data: dict[str, object], grayscale: bool) -> plt.Figure:
    fig, ax = base.new_figure("Figure S1", grayscale)
    palette = base.colors(grayscale)
    ax.axis("off")
    ax.set(xlim=(0, 1), ylim=(0, 1))
    roles = data["source_role_manifest"]
    trace = data["source_trace"]
    parsed_reviews = int(roles.reviews_with_normalized_rows.sum())
    model_ready = int(roles.model_ready_groups.sum())
    primary_points = int(roles.primary_operating_points.sum())
    contributing_reviews = int(trace.groupby("source_route").review_id.nunique().sum())
    joined_rows = int(len(trace) + 23)
    final_rows = int(len(trace))
    assert (parsed_reviews, model_ready, primary_points, contributing_reviews, joined_rows, final_rows) == (382, 444, 273, 210, 4127, 4104)
    nodes = (
        (0.84, "Parsed review corpus", f"{parsed_reviews} review records represented after parsing"),
        (0.68, "Candidate evidence", "1,830 assessable source-defined analysis groups"),
        (0.52, "Structurally eligible", f"{model_ready} model-ready groups; all models converged"),
        (0.36, "Primary designation", f"{primary_points} pooled diagnostic estimates from {contributing_reviews} reviews"),
        (0.20, "Results joined", f"{joined_rows:,} normalised study-result appearances"),
        (0.04, "Final analysis", f"{final_rows:,} primary-analysis study-result appearances"),
    )
    for index, (y, title, body) in enumerate(nodes):
        color = palette[index % len(palette)]
        ax.add_patch(FancyBboxPatch((0.06, y), 0.62, 0.095, boxstyle="round,pad=0.012", fc=base.WHITE, ec=color, lw=1.4))
        ax.text(0.085, y + 0.064, title, fontsize=8.4, weight="bold", color=color)
        ax.text(0.085, y + 0.025, body, fontsize=7.5)
        if index < len(nodes) - 1:
            ax.annotate("", xy=(0.37, y - 0.055), xytext=(0.37, y), arrowprops={"arrowstyle": "-|>", "color": base.MUTED, "lw": 1.0})
    ax.text(0.73, 0.66, "1,386 did not meet structural eligibility\n1,219: fewer than five studies\n155: repeated study identifiers\n11: below chance in source direction\n1: fewer than three paired-state studies", fontsize=6.9, color=base.MUTED, va="top", linespacing=1.26)
    ax.text(0.71, 0.46, "171 model-ready alternatives\nretained in the audit record\nHistorical Cochrane: 225 to 83\nModern Cochrane: 13 to 9\nOSF: 13 to 8 | PMC: 182 to 169\nZenodo: 11 to 4 | Figshare: 0 to 0", fontsize=6.7, color=base.MUTED, va="top", linespacing=1.22)
    ax.text(0.73, 0.15, "23 noncanonical sheet rows excluded\nby one recorded Zenodo workbook rule", fontsize=6.9, color=base.MUTED, va="top", linespacing=1.26)
    fig.suptitle("Construction of the empirical reference sample", y=0.98)
    fig.subplots_adjust(left=0.04, right=0.96, bottom=0.03, top=0.91)
    return fig


def build_captions(data: dict[str, object]) -> dict[str, str]:
    captions = previous.build_captions(data)
    summary = data["anchor_summary"].set_index("starting_probability")
    twenty = summary.loc[0.20]
    hints = hints_profiles(data["anchors"])
    head = hints.loc[hints.test_name.eq("HINTS - subcomponent head impulse")].set_index("starting_probability")
    nystagmus = hints.loc[hints.test_name.eq("HINTS - subcomponent nystagmus")].set_index("starting_probability")
    cea = data["cea_profiles"].loc[data["cea_profiles"].model_specification.eq("v0.42.0 canonical REML")]
    row = ottawa_row(data)

    def cea_value(test: str, probability: float, field: str) -> float:
        return float(cea.loc[cea.test_name.eq(test) & np.isclose(cea.starting_probability, probability), field].iloc[0])

    captions.update({
        "Figure 1": "Figure 1. Empirical reference map of diagnostic information across starting probabilities. The median line and 25th–75th and 10th–90th percentile bands show the percentage of starting diagnostic uncertainty resolved by the same 273 pooled estimates from 210 reviews across starting disease probabilities from 1% to 50%. Median reference values at 5%, 10%, 20%, 30%, 40%, and 50% are labelled directly on the main map. The Ottawa ankle rule, lung ultrasound for pneumonia, non-contrast computed tomography for appendicitis, and computed tomography angiography for an intracranial lesion are source-traced examples used to orient the scale. The examples do not rank clinical value or represent how often tests are used.",
        "Figure 2": f"Figure 2. What diagnostic information adds beyond sensitivity, specificity, and Youden's J. (A–C) Matched surfaces show the percentage of starting diagnostic uncertainty resolved at 5%, 20%, and 50%; all 273 pooled diagnostic estimates are overlaid. Equal-Youden lines mark sensitivity-specificity combinations with J=0.20, 0.40, 0.60, or 0.80. Youden's J is unchanged by starting probability, while the information surface changes. In pooled component profiles from review CD015089, the directly labelled head-impulse and nystagmus profiles had nearly equal J values ({float(head.iloc[0].youden_j):.3f} and {float(nystagmus.iloc[0].youden_j):.3f}) but resolved {100 * float(head.loc[0.05].fraction_entropy_removed):.1f}% and {100 * float(nystagmus.loc[0.05].fraction_entropy_removed):.1f}% of starting uncertainty at 5%. The source-traced comparison describes expected uncertainty reduction and does not establish clinical superiority or net benefit.",
        "Figure 3": f"Figure 3. CEA information profiles for detecting recurrent colorectal cancer. (A) In the primary analysis, the 2.5 and 5 µg/L thresholds removed {100 * cea_value('CEA at 2.5µg/L', 0.05, 'fraction_entropy_removed'):.1f}% and {100 * cea_value('CEA at 5µg/L', 0.05, 'fraction_entropy_removed'):.1f}% of starting uncertainty at 5%, crossed at 12.8%, and removed {100 * cea_value('CEA at 2.5µg/L', 0.20, 'fraction_entropy_removed'):.1f}% and {100 * cea_value('CEA at 5µg/L', 0.20, 'fraction_entropy_removed'):.1f}% at 20%. (B) The alternative synthesis model crossed at 38.1%. The lower threshold favours sensitivity; the higher threshold favours specificity. The starting probability can therefore change which threshold is expected to resolve more uncertainty.",
        "Figure 4": f"Figure 4. Observed-result pathway for the Ottawa ankle rule at a 5% starting probability. Edge width represents how often each result occurs. A positive result occurred {100 * row.positive_result_probability:.1f}% of the time, raised fracture probability to {100 * row.post_positive_probability:.1f}%, and increased uncertainty by {row.positive_entropy_change_bits:.3f} bits. A negative result occurred {100 * row.negative_result_probability:.1f}% of the time, lowered fracture probability to {100 * row.post_negative_probability:.1f}%, and reduced uncertainty by {-row.negative_entropy_change_bits:.3f} bits. The lower bar shows each branch's contribution to expected information. Across both branches, expected information removed {100 * row.information_bits / row.starting_entropy_bits:.1f}% of starting uncertainty ({row.information_bits:.3f} bits). An individual observed result moves probability and may increase or decrease uncertainty; diagnostic information is the expectation across possible results before testing. Values use the source-traced primary pooled estimate from 16 studies.",
        "Figure S1": "Figure S1. Construction of the empirical reference sample. The parsed corpus represented 382 review records and 1,830 assessable source-defined analysis groups. Common structural rules identified 444 model-ready groups, and all models converged. Route-specific primary designation retained 273 pooled diagnostic estimates from 210 reviews. These groups joined to 4,127 normalised study-result appearances; one recorded canonical-sheet rule excluded 23 results, leaving 4,104 primary-analysis study-result appearances. Counts beside each transition give the exact structural and source-route accounting.",
        "Figure S9": previous.build_captions(data)["Figure 4"].replace("Figure 4.", "Figure S9."),
        "Table 1": "Table 1. Empirical reference scale for diagnostic uncertainty resolved. Each row reports the median with its interquartile range and the 10th–90th percentile range across the same 273 pooled diagnostic estimates from 210 reviews.",
    })
    return {
        display: caption.replace("operating points", "pooled diagnostic estimates")
        .replace("operating point", "pooled diagnostic estimate")
        .replace("operating-point", "pooled-estimate")
        .replace("study-row", "study-result")
        for display, caption in captions.items()
    }


def build(root: Path, output_analysis: Path, manuscript_dir: Path, qa_dir: Path) -> None:
    configure_base()
    data = base.load_data(root)
    data["probability_profiles"] = pd.read_csv(root / "analysis/bmj_rmr_v042/derived_operating_points_probability_profiles.csv")
    data["source_role_manifest"] = pd.read_csv(root / "analysis/full_atlas/v9/source_role_manifest.csv")
    data["source_trace"] = pd.read_csv(root / "analysis/bmj_rmr_v042/source_trace_all_primary.csv")
    candidate_pairs = rank_near_equal_youden_pairs(data["anchors"])
    base.write_csv(output_analysis / "equal_youden_candidate_pairs.csv", candidate_pairs)
    library_rows: list[dict[str, str]] = []
    for display in FIGURE_SPECS:
        library_rows.extend(
            (
                {"display": display, "library": "Matplotlib", "version": mpl.__version__, "repository_url": "https://github.com/matplotlib/matplotlib", "role": "vector-first plotting and export"},
                {"display": display, "library": "SciencePlots", "version": "2.2.2", "repository_url": "https://github.com/garrettj403/SciencePlots", "role": "publication style configuration"},
            )
        )
    library_rows.append({"display": "Figure 4", "library": "NetworkX", "version": nx.__version__, "repository_url": "https://github.com/networkx/networkx", "role": "directed probability graph, layout, nodes, edges, and labels"})
    library_manifest = output_analysis / "figure_library_manifest.csv"
    base.write_csv(library_manifest, pd.DataFrame.from_records(library_rows))
    figures_dir = manuscript_dir / "figures" / f"v{VERSION}"
    figures_dir.mkdir(parents=True, exist_ok=True)
    output_analysis.mkdir(parents=True, exist_ok=True)
    qa_dir.mkdir(parents=True, exist_ok=True)
    builders = {
        "Figure 1": lambda gray: figure_1(data, gray),
        "Figure 2": lambda gray: figure_2(data, gray),
        "Figure 3": lambda gray: figure_3(data, gray),
        "Figure 4": lambda gray: figure_4(data, gray),
        "Figure S1": lambda gray: figure_s1(data, gray),
        "Figure S2": lambda gray: base.figure_s2(data["fidelity"], data["audit_summary"], data["routes"], gray),
        "Figure S3": lambda gray: base.figure_s3(data["anchors"], data["anchor_summary"], gray),
        "Figure S4": lambda gray: base.figure_s4(data["recovery"], data["recovery_summary"], data["diagnostics"], data["diagnostic_summary"], gray),
        "Figure S5": lambda gray: base.figure_s5(data["sensitivity"], data["sensitivity_summary"], data["estimator_effect"], gray),
        "Figure S6": lambda gray: base.figure_s6(data["contrasts"], data["movement"], gray),
        "Figure S7": lambda gray: base.figure_s7(data["near_ties"], data["restricted"], gray),
        "Figure S8": lambda gray: base.figure_s8(data["crossings"], data["cea_profiles"], data["theory_summary"], gray),
        "Figure S9": lambda gray: base.figure_4(data["branches"], data["branch_summary"], gray),
    }
    figure_paths: list[Path] = []
    for display in FIGURE_SPECS:
        for grayscale in (False, True):
            figure_paths.extend(base.save_figure(builders[display](grayscale), figures_dir, display, grayscale))

    captions = build_captions(data)
    table_paths = write_table(manuscript_dir, build_table_1(data["anchors"]), captions["Table 1"])
    caption_paths = previous.write_caption_files(manuscript_dir, output_analysis, captions)
    display_source_paths = base.write_display_sources(output_analysis, data)
    for name, frame in {
        "figure_1_reference_percentiles.csv": profile_percentiles(data["probability_profiles"]),
        "figure_1_clinical_profiles.csv": data["probability_profiles"].loc[data["probability_profiles"].group_id.isin(data["landmarks"].group_id)],
        "figure_2_all_pooled_estimates.csv": data["anchors"],
        "figure_2_hints_component_profiles.csv": hints_profiles(data["anchors"]),
        "figure_3_cea_profiles.csv": data["cea_profiles"].loc[data["cea_profiles"].model_specification.isin(("v0.42.0 canonical REML", "v0.40.0/v0.41.0 regularized GH15"))],
        "figure_4_ottawa_pathway.csv": pd.DataFrame([ottawa_row(data)]),
    }.items():
        path = output_analysis / "display_sources" / name
        base.write_csv(path, frame)
        display_source_paths.append(path)
    display_manifest = previous.write_display_manifest(root, manuscript_dir, output_analysis, captions)
    bounds_paths = base.audit_pdf_text_bounds(figures_dir, qa_dir)
    previews = base.render_pdf_previews(figures_dir, qa_dir)
    contact_paths = base.write_contact_sheets(previews, qa_dir)
    visual_json = base.initial_visual_log(qa_dir, previews)
    visual_md = base.write_visual_markdown(qa_dir)
    source_manifest = output_analysis / "source_hash_manifest.csv"
    base.write_csv(source_manifest, base.source_hash_rows(root))
    artifacts = [*figure_paths, *table_paths, *caption_paths, *display_source_paths, display_manifest, library_manifest, *bounds_paths, *previews, *contact_paths, visual_json, visual_md, source_manifest]
    base.write_csv(output_analysis / "display_artifact_manifest.csv", base.artifact_rows(root, artifacts))


def parse_args() -> argparse.Namespace:
    parser = argparse.ArgumentParser(description=__doc__)
    parser.add_argument("--root", type=Path, default=Path(__file__).resolve().parents[1])
    parser.add_argument("--output-analysis", type=Path)
    parser.add_argument("--manuscript-dir", type=Path)
    parser.add_argument("--qa-dir", type=Path)
    return parser.parse_args()


if __name__ == "__main__":
    arguments = parse_args()
    repository = arguments.root.resolve()
    build(
        repository,
        (arguments.output_analysis or repository / "analysis/bmj_rmr_v190").resolve(),
        (arguments.manuscript_dir or repository / "docs/manuscript/v25").resolve(),
        (arguments.qa_dir or repository / "analysis/bmj_rmr_v190/qa").resolve(),
    )
```
