## Supplementary material for "The information in diagnostic tests": Executable analysis code, derived inputs, source-acquisition records, tests, and release metadata: bmj_rmr_v1100_bootstrap.py

```
"""Bootstrap confidence intervals for the Table 1 reference medians (manuscript 1.10.0).

Reads the locked v0.42.0 pooled estimates, resamples the 273 estimates with replacement,
and reports percentile intervals for the median percentage of starting uncertainty resolved
at 5%, 20%, and 50% starting probability. The intervals describe sampling of the reference
sample; uncertainty within each pooled estimate is not propagated.

Outputs (deterministic):
  analysis/bmj_rmr_v1100/reference_scale_bootstrap.csv
  analysis/bmj_rmr_v1100/reference_scale_bootstrap_summary.json
"""

from __future__ import annotations

import argparse
import json
from pathlib import Path

import numpy as np
import pandas as pd

REPLICATES = 2000
SEED = 20260830
ANCHORS = (0.05, 0.20, 0.50)
SOURCE = "analysis/bmj_rmr_v042/derived_operating_points_standard_anchors.csv"


def bootstrap_median(values: np.ndarray, replicates: int, seed: int) -> tuple[float, float, float]:
    rng = np.random.default_rng(seed)
    n = values.shape[0]
    draws = rng.integers(0, n, size=(replicates, n))
    medians = np.median(values[draws], axis=1)
    return float(np.median(values)), float(np.percentile(medians, 2.5)), float(np.percentile(medians, 97.5))


def run(root: Path) -> pd.DataFrame:
    anchors = pd.read_csv(root / SOURCE)
    rows = []
    for index, prior in enumerate(ANCHORS):
        frame = anchors.loc[(anchors.starting_probability.astype(float) - prior).abs() < 1e-12]
        values = frame.fraction_entropy_removed.to_numpy(dtype=float)
        assert values.shape[0] == 273, values.shape
        median, low, high = bootstrap_median(values, REPLICATES, SEED + index)
        rows.append(
            {
                "starting_probability": prior,
                "n_estimates": int(values.shape[0]),
                "median_fraction": median,
                "ci_low_fraction": low,
                "ci_high_fraction": high,
                "median_percent": round(100 * median, 1),
                "ci_low_percent": round(100 * low, 1),
                "ci_high_percent": round(100 * high, 1),
                "replicates": REPLICATES,
                "seed": SEED + index,
                "method": "bootstrap of pooled estimates; percentile interval (2.5th, 97.5th) of the resampled median",
                "source_file": SOURCE,
            }
        )
    table = pd.DataFrame.from_records(rows)
    out = root / "analysis/bmj_rmr_v1100"
    out.mkdir(parents=True, exist_ok=True)
    table.to_csv(out / "reference_scale_bootstrap.csv", index=False, lineterminator="\n")
    summary = {
        "analysis_version": "1.10.0",
        "replicates": REPLICATES,
        "seed_base": SEED,
        "source_file": SOURCE,
        "interval": "percentile 95% (2.5th to 97.5th percentile of resampled medians)",
        "scope": "sampling of the fixed 273-estimate reference sample; within-estimate uncertainty not propagated",
        "anchors": [
            {
                "starting_probability": row["starting_probability"],
                "median_percent": row["median_percent"],
                "ci_low_percent": row["ci_low_percent"],
                "ci_high_percent": row["ci_high_percent"],
            }
            for row in rows
        ],
    }
    (out / "reference_scale_bootstrap_summary.json").write_text(json.dumps(summary, indent=2) + "\n", encoding="utf-8")
    return table


def parse_args() -> argparse.Namespace:
    parser = argparse.ArgumentParser(description=__doc__)
    parser.add_argument("--root", type=Path, default=Path(__file__).resolve().parents[1])
    return parser.parse_args()


if __name__ == "__main__":
    result = run(parse_args().root.resolve())
    print(result[["starting_probability", "median_percent", "ci_low_percent", "ci_high_percent"]].to_string(index=False))
```
