## Supplementary material for "The information in diagnostic tests": Executable analysis code, derived inputs, source-acquisition records, tests, and release metadata: bmj_rmr_v1110_visuals.py

```
"""Redraw the four main figures; preserve the locked supplementary assets."""
from __future__ import annotations

import argparse
import hashlib
import json
import shutil
from pathlib import Path

import matplotlib as mpl
import matplotlib.pyplot as plt
import numpy as np
import pandas as pd

from scripts import bmj_rmr_v190_visuals as prior

VERSION = "1.11.0"
base = prior.base
FIGURE_SPECS = {
    **prior.FIGURE_SPECS,
    "Figure 1": {**prior.FIGURE_SPECS["Figure 1"], "height_mm": 120.0},
    "Figure 2": {**prior.FIGURE_SPECS["Figure 2"], "height_mm": 142.0},
    "Figure 3": {**prior.FIGURE_SPECS["Figure 3"], "height_mm": 110.0},
    "Figure 4": {**prior.FIGURE_SPECS["Figure 4"], "height_mm": 118.0},
}
BLUE, AMBER, INK = "#005EA8", "#B66A00", "#202B35"


def canvas(number: int, gray: bool):
    base.set_style(gray)
    mpl.rcParams.update({"font.size": 8.5, "axes.labelsize": 8.5,
                        "xtick.labelsize": 8.5, "ytick.labelsize": 8.5})
    return plt.figure(figsize=base.figure_size(FIGURE_SPECS[f"Figure {number}"]["height_mm"]))


def heading(fig, text):
    fig.text(0.075, 0.96, text, fontsize=9.5, weight="semibold", va="top")


def figure_1(data, gray=False):
    fig = canvas(1, gray)
    ax = fig.add_axes([0.10, 0.16, 0.62, 0.66])
    summary = prior.profile_percentiles(data["probability_profiles"])
    x = 100 * summary.starting_probability
    median = "#111111" if gray else BLUE
    ax.fill_between(x, 100*summary.p10, 100*summary.p90, color="#E8E8E8" if gray else "#E2ECF3", lw=0, label="10th-90th percentiles")
    ax.fill_between(x, 100*summary.p25, 100*summary.p75, color="#BDBDBD" if gray else "#AFCADC", lw=0, label="25th-75th percentiles")
    ax.plot(x, 100*summary["median"], color=median, lw=2.2, zorder=4)
    roles = (
        ("Ottawa ankle rule", "Ottawa ankle rule", "#58666E", "-", "o", 8.5),
        ("Lung ultrasound pneumonia", "Lung ultrasound\nfor pneumonia", "#007F68", "--", "s", 43),
        ("Non-contrast CT appendicitis", "Non-contrast CT\nfor appendicitis", "#8F5DA2", "-.", "^", 73),
        ("CT angiography intracranial lesion", "CT angiography\nintracranial lesion", AMBER, ":", "D", 61),
    )
    for label, name, color, style, marker, label_y in roles:
        color = "#444444" if gray else color
        landmark = data["landmarks"].loc[lambda f: f.label.eq(label)].iloc[0]
        frame = data["probability_profiles"].loc[lambda f: f.group_id.eq(landmark.group_id)].sort_values("starting_probability")
        ax.plot(100*frame.starting_probability, 100*frame.fraction_entropy_removed,
                color=color, ls=style, marker=marker, markevery=[9,29,49], ms=3.3, lw=1.4)
        ax.annotate(name, xy=(50, 100*frame.fraction_entropy_removed.iloc[-1]),
                    xytext=(53, label_y), fontsize=8.5, color=color, va="center",
                    annotation_clip=False, arrowprops={"arrowstyle":"-", "color":color, "lw":0.7})
    for p, offset in ((0.05, -16), (0.20, -18), (0.50, -16)):
        row = summary.loc[np.isclose(summary.starting_probability, p)].iloc[0]
        y = 100*row["median"]
        ax.plot(100*p, y, "o", ms=4.5, mfc="white", mec=median, mew=1.2, zorder=6)
        ax.annotate(f"{y:.1f}%", (100*p,y), xytext=(0,offset), textcoords="offset points",
                    ha="center", color=median, fontsize=9.5 if p==0.20 else 8.5,
                    weight="bold" if p==0.20 else "normal", va="center")
    ax.annotate("Median", (50,100*summary["median"].iloc[-1]), xytext=(53,31.5),
                color=median, fontsize=8.5, weight="semibold", va="center", annotation_clip=False,
                arrowprops={"arrowstyle":"-", "color":median, "lw":0.7})
    ax.set(xlim=(1,50), ylim=(0,80), xlabel="Starting disease probability (%)",
           ylabel="Starting diagnostic uncertainty resolved (%)")
    ax.set_xticks([1,10,20,30,40,50]); ax.set_yticks([0,20,40,60,80])
    ax.grid(axis="y", color="#E3E7EB", lw=0.5)
    ax.legend(loc="lower left", bbox_to_anchor=(0,1.015), ncol=2, fontsize=8.0,
              borderaxespad=0, handlelength=1.5, columnspacing=1.0)
    heading(fig, "Diagnostic information across starting probabilities")
    fig.text(0.10,0.035,"273 pooled estimates from 210 reviews; bands describe the reference sample.",fontsize=8.0)
    return fig


def figure_2(data, gray=False):
    fig = canvas(2, gray)
    anchors = data["anchors"]
    hints = prior.hints_profiles(anchors)
    grid = np.linspace(0.001,0.999,220)
    se, sp = np.meshgrid(grid,grid)
    for i,p in enumerate((0.05,0.20,0.50)):
        ax=fig.add_axes([0.095+i*0.305,0.545,0.265,0.336])
        contour=ax.contourf(100*se,100*sp,prior.information_percent(p,se,sp),
                            levels=np.arange(0,101,10),cmap="Greys_r" if gray else "cividis",vmin=0,vmax=100)
        rows=anchors.loc[np.isclose(anchors.starting_probability,p)]
        ax.scatter(100*rows.sensitivity,100*rows.specificity,s=7,fc="white",ec="#303030",lw=.3,alpha=.65)
        line=np.linspace(.5,1,100)
        ax.plot(100*line,100*(1.5-line),"--",color="white",lw=1.0)
        if i==0:
            for name,marker in (("head impulse","o"),("nystagmus","s")):
                r=hints.loc[hints.test_name.str.endswith(name)&np.isclose(hints.starting_probability,p)].iloc[0]
                ax.scatter(100*r.sensitivity,100*r.specificity,s=46,marker=marker,fc="none",ec="white",lw=1.6,zorder=6)
        ax.set(xlim=(0,100),ylim=(0,100),xlabel="Sensitivity (%)",xticks=[0,50,100],yticks=[0,50,100])
        if i==0: ax.set_ylabel("Specificity (%)")
        else: ax.tick_params(labelleft=False)
        ax.set_aspect("equal")
        ax.set_title(f"{chr(65+i)}  Starting probability {100*p:.0f}%",loc="left",fontsize=8.5,pad=8)
    cax=fig.add_axes([0.22,0.445,0.65,0.022])
    cb=fig.colorbar(contour,cax=cax,orientation="horizontal",ticks=[0,20,40,60,80,100])
    cb.set_label("Starting diagnostic uncertainty resolved (%)",fontsize=8.5)
    cb.ax.tick_params(labelsize=8.0)
    fig.text(.095,.35,"D  Nearly equal Youden's J, different information at 5%",fontsize=8.5,weight="semibold")
    ax=fig.add_axes([.37,.15,.47,.16])
    labels=[]
    for y,(name,marker,color) in enumerate((("head impulse","o",BLUE),("nystagmus","s",AMBER))):
        r=hints.loc[hints.test_name.str.endswith(name)&np.isclose(hints.starting_probability,.05)].iloc[0]
        color="#333333" if gray else color
        value=100*r.fraction_entropy_removed
        ax.hlines(y,0,value,color=color,lw=1.4)
        ax.plot(value,y,marker,ms=5,color=color)
        ax.text(value+1.5,y,f"{value:.1f}%",va="center",fontsize=8.5,color=color)
        labels.append(f"{name.capitalize()}  (J={r.youden_j:.3f})")
    ax.set(yticks=[0,1],yticklabels=labels,ylim=(1.6,-.6),xlim=(0,40),xticks=[0,10,20,30,40],
           xlabel="Starting diagnostic uncertainty resolved (%)")
    ax.spines["left"].set_visible(False); ax.tick_params(axis="y",length=0,pad=10)
    heading(fig,"Information depends on both accuracy and starting probability")
    fig.text(.095,.055,"A-C: all 273 estimates; dashed line: J=0.5. D: pooled HINTS components.",fontsize=8.0)
    return fig


def figure_3(data, gray=False):
    fig=canvas(3,gray)
    models=(("v0.42.0 canonical REML","Primary analysis","canonical_reml_probability"),
            ("v0.40.0/v0.41.0 regularized GH15","Alternative synthesis model","regularized_gh15_probability"))
    for i,(model,title,key) in enumerate(models):
        ax=fig.add_axes([.10+.47*i,.21,.35,.58])
        for name,label,color,style,dy in (("CEA at 2.5µg/L","2.5 µg/L",BLUE,"-",2.1),
                                          ("CEA at 5µg/L","5 µg/L",AMBER,"--",-2.3)):
            frame=data["cea_profiles"].loc[lambda f: f.model_specification.eq(model)&f.test_name.eq(name)].sort_values("starting_probability")
            color="#222222" if gray else color
            ax.plot(100*frame.starting_probability,100*frame.fraction_entropy_removed,color=color,ls=style,lw=1.8)
            y=100*frame.fraction_entropy_removed.iloc[-1]
            ax.annotate(label,xy=(50,y),xytext=(52,y+dy),fontsize=8.5,color=color,va="center",
                        annotation_clip=False,arrowprops={"arrowstyle":"-","color":color,"lw":.7})
        crossing=100*data["crossings_exact"][key]
        ax.axvline(crossing,color="#65717B",ls=":",lw=1.0)
        ax.text(crossing,36,f"Crossing\n{crossing:.1f}%",ha="center",va="top",fontsize=8.5,
                bbox={"fc":"white","ec":"none","pad":1.5})
        ax.set(xlim=(1,62),ylim=(0,40),xticks=[1,10,20,30,40,50],yticks=[0,10,20,30,40])
        ax.set_xlabel("Recurrence probability before testing (%)",fontsize=8.0)
        if i==0: ax.set_ylabel("Starting diagnostic uncertainty resolved (%)")
        ax.set_title(f"{chr(65+i)}  {title}",loc="left",fontsize=8.5,pad=9)
        ax.grid(axis="y",color="#E3E7EB",lw=.5)
        ax.spines["bottom"].set_bounds(1,50)
    heading(fig,"CEA thresholds change information ordering as starting probability changes")
    fig.text(.10,.065,"Both panels use the same scale; crossings describe expected information.",fontsize=8.0)
    return fig


def ottawa_summary(data):
    r=prior.ottawa_row(data)
    remaining=sum(float(r[f"{s}_result_probability"])*float(r[f"{s}_branch_entropy_bits"]) for s in ("positive","negative"))
    removed=float(r.starting_entropy_bits)-remaining
    if not np.isclose(removed,r.information_bits,rtol=0,atol=1e-12):
        raise ValueError("Ottawa expected entropy does not match the locked information value")
    return r, remaining, 100*removed/float(r.starting_entropy_bits)


def figure_4(data,gray=False):
    fig=canvas(4,gray)
    r,remaining,percent=ottawa_summary(data)
    heading(fig,"One result can increase uncertainty; the test reduces it on average")
    fig.text(.075,.875,f"Ottawa ankle rule  |  Before testing: {100*r.starting_probability:.1f}% fracture probability",fontsize=8.5)
    ax=fig.add_axes([.075,.47,.86,.33]); ax.set(xlim=(0,1),ylim=(0,1)); ax.axis("off")
    ax.text(0,1.05,"A  After one result",fontsize=9,weight="semibold")
    for x,label in ((.30,"How often"),(.56,"Fracture probability"),(.83,"Uncertainty change")):
        ax.text(x,.86,label,ha="center",fontsize=8.5)
    ax.axhline(.78,color="#D5DDE3",lw=.6); ax.axhline(.10,color="#D5DDE3",lw=.6)
    for state,y,color in (("positive",.58,AMBER),("negative",.26,BLUE)):
        color="#222222" if gray else color
        change=float(r[f"{state}_entropy_change_bits"])
        ax.text(0,y,state.capitalize(),va="center",fontsize=9,weight="semibold",color=color)
        ax.text(.30,y,f"{100*r[f'{state}_result_probability']:.1f}%",ha="center",va="center",fontsize=9)
        ax.text(.56,y,f"{100*r[f'post_{state}_probability']:.1f}%",ha="center",va="center",fontsize=9)
        ax.text(.83,y,f"{change:+.3f} bits\n{'increases' if change>0 else 'decreases'}",ha="center",va="center",fontsize=8.5,color=color)
    fig.text(.075,.425,"B  Before knowing the result",fontsize=9,weight="semibold")
    ax=fig.add_axes([.255,.18,.45,.17])
    ax.barh([1,0],[r.starting_entropy_bits,remaining],height=.36,
            color=["#C7CDD2","#444444" if gray else BLUE])
    for y,value in ((1,float(r.starting_entropy_bits)),(0,remaining)):
        ax.text(value+.007,y,f"{value:.3f}",va="center",fontsize=8.5)
    ax.set(yticks=[1,0],yticklabels=["Before testing","After testing\n(expected)"],
           xlim=(0,.35),ylim=(-.6,1.6),xticks=[0,.1,.2,.3],xlabel="Diagnostic uncertainty (bits)")
    ax.spines["left"].set_visible(False); ax.spines["bottom"].set_bounds(0,.3); ax.tick_params(axis="y",length=0,pad=8)
    fig.text(.775,.29,f"{percent:.1f}% removed",fontsize=9.5,weight="bold",color="#222222" if gray else BLUE,ha="center")
    fig.text(.775,.245,f"{r.information_bits:.3f} bits\non average",fontsize=8.5,ha="center",va="top")
    fig.text(.075,.055,"The expected value weights both outcomes by how often they occur.",fontsize=8.0)
    return fig


def load_data(root):
    data=base.load_data(root)
    data["probability_profiles"]=pd.read_csv(root/"analysis/bmj_rmr_v042/derived_operating_points_probability_profiles.csv")
    data["crossings_exact"]=json.loads((root/"analysis/bmj_rmr_v042/phase3_summary.json").read_text())["empirical_cea_crossing"]
    return data


def build(root):
    data=load_data(root)
    source=root/"docs/manuscript/v27"
    figures=source/f"figures/v{VERSION}"
    analysis=root/"analysis/bmj_rmr_v1110"
    qa=analysis/"qa"
    # Retain released supplemental masters byte for byte; redraw only Figures 1-4.
    shutil.copytree(root/"docs/manuscript/v26/figures/v1.10.0",figures,dirs_exist_ok=True)
    shutil.copytree(root/"docs/manuscript/v26/tables/v1.10.0",source/f"tables/v{VERSION}",dirs_exist_ok=True)
    base.VERSION=VERSION; base.RELEASE_DATE="2026-09-08"; base.FIGURE_SPECS=FIGURE_SPECS
    for i,builder in enumerate((figure_1,figure_2,figure_3,figure_4),1):
        for gray in (False,True): base.save_figure(builder(data,gray),figures,f"Figure {i}",gray)
    captions=pd.read_csv(root/"analysis/bmj_rmr_v1100/figure_captions.csv")
    replacements={
        "Figure 1":("Median reference values at 5%, 10%, 20%, 30%, 40%, and 50% are labelled directly on the main map.","Median reference values at 5%, 20%, and 50% are labelled; the 29.2% median at 20% is emphasized. The bands describe the distribution of pooled estimates, not confidence intervals."),
        "Figure 2":("Equal-Youden lines mark sensitivity-specificity combinations with J=0.20, 0.40, 0.60, or 0.80.","A dashed line in each map marks J=0.50. (D) A separate comparison shows the two highlighted HINTS component profiles at a 5% starting probability."),
    }
    for display,(old,new) in replacements.items():
        mask=captions.display.eq(display)
        assert old in captions.loc[mask,"caption"].iloc[0]
        captions.loc[mask,"caption"]=captions.loc[mask,"caption"].str.replace(old,new,regex=False)
    mask=captions.display.eq("Figure 3")
    captions.loc[mask,"caption"] += " Both panels use the same axes; the crossings do not imply a large information difference or clinical superiority."
    r,remaining,percent=ottawa_summary(data)
    captions.loc[captions.display.eq("Figure 4"),"caption"]=(
        f"Figure 4. Individual outcomes and expected uncertainty for the Ottawa ankle rule at a 5% starting probability. "
        f"(A) A positive result has probability {100*r.positive_result_probability:.1f}%, raises fracture probability to {100*r.post_positive_probability:.1f}%, and increases uncertainty by {r.positive_entropy_change_bits:.3f} bits. "
        f"A negative result has probability {100*r.negative_result_probability:.1f}%, lowers fracture probability to {100*r.post_negative_probability:.1f}%, and reduces uncertainty by {-r.negative_entropy_change_bits:.3f} bits. "
        f"(B) Weighting the two conditional entropies by their result probabilities gives {remaining:.3f} bits of expected remaining uncertainty, compared with {r.starting_entropy_bits:.3f} bits before testing. "
        f"The expected reduction is {r.information_bits:.3f} bits, or {percent:.1f}% of starting uncertainty. An individual result can increase uncertainty while the test reduces it on average. "
        "Values use the primary pooled estimate from 16 studies; they describe a standardized 5% starting probability. Differences and percentages are calculated before rounding.")
    base.write_csv(analysis/"figure_captions.csv",captions)
    (figures/"FIGURE_CAPTIONS.md").write_text("# Figure captions\n\n"+"\n\n".join(captions.caption)+"\n",encoding="utf-8")
    (analysis/"ottawa_expected_uncertainty.json").write_text(json.dumps({"starting_entropy_bits":float(r.starting_entropy_bits),"expected_remaining_entropy_bits":remaining,"information_bits":float(r.information_bits),"percent_removed":percent},indent=2)+"\n")
    base.audit_pdf_text_bounds(figures,qa)
    paths=list(figures.glob("*"))+list((source/f"tables/v{VERSION}").glob("*"))
    paths += [root/p for p in sorted({p for i in range(1,5) for p in prior.DISPLAY_SOURCES[f"Figure {i}"]})]
    paths += [Path(__file__).resolve(),root/"analysis/bmj_rmr_v042/phase3_summary.json"]
    base.write_csv(analysis/"figure_source_and_artifact_hashes.csv",pd.DataFrame([
        {"path":p.relative_to(root).as_posix(),"bytes":p.stat().st_size,"sha256":hashlib.sha256(p.read_bytes()).hexdigest()} for p in paths if p.is_file()]))
    print(f"Built four main figures in colour and grayscale: {figures}")


if __name__=="__main__":
    parser=argparse.ArgumentParser(description=__doc__)
    parser.add_argument("--root",type=Path,default=Path(__file__).resolve().parents[1])
    build(parser.parse_args().root.resolve())
```
