## Supplementary material for "The information in diagnostic tests": Executable analysis code, derived inputs, source-acquisition records, tests, and release metadata: bmj_rmr_v1120_visuals.py

```
"""Add the hypothetical entropy chord; retain the empirical figure sequence."""
import argparse
import hashlib
import json
import shutil
from pathlib import Path

import matplotlib.pyplot as plt
import numpy as np
import pandas as pd

from scripts import bmj_rmr_v1110_visuals as previous
from scripts.bmj_rmr_v042 import binary_entropy, expanded_diagnostic_metrics

VERSION = "1.12.0"
S10 = {"stem": "figure_s10_entropy_chord", "height_mm": 138.0}


def teaching_values():
    rows = []
    for p in (0.20, 0.05):
        metrics = expanded_diagnostic_metrics(0.90, 0.95, p)
        rows.append({"starting_probability": p, "sensitivity": .90, "specificity": .95,
                     "starting_entropy_bits": float(binary_entropy(p)),
                     **{key: float(value) for key, value in metrics.items()}})
    return pd.DataFrame(rows)


def entropy_chord(gray=False):
    previous.base.set_style(gray)
    plt.rcParams.update({"font.size": 8.5, "axes.labelsize": 8.5,
                        "xtick.labelsize": 8, "ytick.labelsize": 8})
    fig = plt.figure(figsize=previous.base.figure_size(S10["height_mm"]))
    blue, amber = ("#222222", "#666666") if gray else (previous.BLUE, previous.AMBER)
    fig.text(.085, .965, "Expected learning is the gap between the curve and the chord",
             fontsize=9.5, weight="semibold", va="top")
    fig.text(.085, .912, "Hypothetical test: sensitivity 90%, specificity 95%", fontsize=8.5)
    x = np.linspace(0, 1, 1001)
    for i, r in enumerate(teaching_values().itertuples(index=False)):
        ax = fig.add_axes([.09 + i * .48, .31, .37, .49])
        ax.plot(100*x, binary_entropy(x), color="#444444", lw=1.1)
        neg, pos, p = r.post_negative_probability, r.post_positive_probability, r.starting_probability
        hn, hp, h = float(binary_entropy(neg)), float(binary_entropy(pos)), r.starting_entropy_bits
        remaining = r.expected_posterior_entropy_bits
        ax.plot([100*neg, 100*pos], [hn, hp], "--", color="#777777", lw=1)
        ax.scatter([100*neg, 100*pos], [hn, hp], c="#333333", marker="s", s=22, zorder=4)
        ax.scatter([100*p], [h], c=blue, marker="o", s=28, zorder=5)
        ax.scatter([100*p], [remaining], c=amber, marker="D", s=25, zorder=5)
        ax.annotate("", (100*p, h), (100*p, remaining),
                    arrowprops={"arrowstyle": "<->", "color": amber, "lw": 1.3,
                                "shrinkA": 4, "shrinkB": 4})
        # Label positions are explicit for these two stipulated examples only.
        locations = ((37,.69), (39,.26), (20,.04), (62,.72)) if i == 0 else ((24,.38), (24,.19), (30,.03), (65,.72))
        points = ((p,h,f"Prior {100*p:.0f}%",blue),
                  (p,remaining,"Average after\ntesting",amber),
                  (neg,hn,f"Negative\n{100*neg:.2f}%","#333333"),
                  (pos,hp,f"Positive\n{100*pos:.2f}%","#333333"))
        for (px,py,label,color),location in zip(points,locations):
            ax.annotate(label, (100*px,py), location, fontsize=8, color=color,
                        ha="center", va="center",
                        arrowprops={"arrowstyle":"-", "lw":.6, "color":color})
        ax.set(xlim=(0,100), ylim=(-.03,1.07), xticks=[0,25,50,75,100],
               yticks=[0,.25,.5,.75,1], xlabel="Disease probability (%)")
        if i == 0:
            ax.set_ylabel("Binary entropy (bits)")
        ax.set_title(f"{'AB'[i]}  Starting probability {100*p:.0f}%", loc="left", fontsize=9, pad=9)
        left = .09 + i*.48
        fig.text(left, .19, f"Result weights: + {100*r.positive_result_probability:g}%\n"
                 f"                          − {100*r.negative_result_probability:g}%", fontsize=8.5)
        fig.text(left, .11, f"Expected reduction: {r.information_bits:.3f} bits\n"
                 f"{100*r.fraction_entropy_removed:.1f}% of starting entropy", fontsize=8.5,
                 color=blue, weight="semibold")
    fig.text(.085,.035,"The diamond is a weighted average, not an additional possible result.",fontsize=8)
    return fig


def build(root):
    source = root / "docs/manuscript/v28"
    figures = source / f"figures/v{VERSION}"
    analysis = root / "analysis/bmj_rmr_v1120"
    analysis.mkdir(parents=True, exist_ok=True)
    shutil.copytree(root / "docs/manuscript/v27/figures/v1.11.0", figures, dirs_exist_ok=True)
    base = previous.base
    base.VERSION, base.RELEASE_DATE = VERSION, "2026-09-13"
    base.FIGURE_SPECS = {**previous.FIGURE_SPECS, "Figure S10": S10}
    data = previous.load_data(root)
    for gray in (False, True):
        fig = previous.figure_4(data, gray)
        for text in fig.findobj(match=plt.Text):
            text.set_text(text.get_text().replace("How often", "Expected frequency")
                          .replace("The expected value weights both outcomes by how often they occur.",
                                   "Result frequencies are model-implied at a standardized 5% prior."))
        base.save_figure(fig, figures, "Figure 4", gray)
        base.save_figure(entropy_chord(gray), figures, "Figure S10", gray)
    values = teaching_values()
    values.to_csv(analysis / "teaching_values.csv", index=False, lineterminator="\n")
    captions = pd.read_csv(root / "analysis/bmj_rmr_v1110/figure_captions.csv")
    r, remaining, percent = previous.ottawa_summary(data)
    captions.loc[captions.display.eq("Figure 4"), "caption"] += (
        " Result frequencies are implied by the pooled sensitivity and specificity at this assumed prior,"
        " not observed frequencies in a 5% prevalence cohort. Uncertainty change means posterior entropy"
        " minus prior entropy; its weighted mean is minus mutual information."
        " The negative branch supplies 80.1% of probability-weighted posterior-to-prior KL gain;"
        " this share does not partition entropy reduction.")
    captions.loc[captions.display.eq("Figure S9"), "caption"] += (
        " Entropy change is posterior minus prior; negative values indicate uncertainty reduction."
        " Its result-weighted mean is minus mutual information, whereas the weighted mean of"
        " posterior-to-prior KL gains is mutual information.")
    caption = (
        "Figure S10. How expected diagnostic information appears on the binary entropy curve. "
        "An original hypothetical illustration uses stipulated sensitivity 90% and specificity 95%, "
        "with (A) a 20% prior and (B) a 5% prior. Squares mark posterior states after negative and "
        "positive results; the circle marks the prior. The diamond lies on the dashed chord joining "
        "the posterior points, at their result-probability-weighted mean entropy and mean probability "
        "(which equals the prior). It is an average, not a patient outcome. The vertical gap to the "
        "prior on the curve is mutual information: 0.437 bits (60.6%) at 20%, and 0.149 bits (52.1%) "
        "at 5%. In B, a positive result moves probability to 48.65%, increasing entropy despite "
        "positive KL gain and a positive expected entropy reduction. Axes are linear and identical; "
        "weights and values are calculated before rounding. These stipulated examples are distinct "
        "from the empirical Ottawa example in Figure 4.")
    captions = pd.concat([captions, pd.DataFrame([{"display":"Figure S10","caption":caption}])], ignore_index=True)
    captions.to_csv(analysis / "figure_captions.csv", index=False, lineterminator="\n")
    (figures / "FIGURE_CAPTIONS.md").write_text("# Figure captions\n\n" + "\n\n".join(captions.caption) + "\n", encoding="utf-8", newline="\n")
    (analysis / "ottawa_expected_uncertainty.json").write_text(json.dumps({
        "starting_entropy_bits":float(r.starting_entropy_bits), "expected_remaining_entropy_bits":remaining,
        "information_bits":float(r.information_bits), "percent_removed":percent,
        "negative_weighted_kl_share":float(r.negative_information_contribution_bits/r.information_bits)}, indent=2)+"\n",encoding="utf-8",newline="\n")
    base.audit_pdf_text_bounds(figures, analysis / "qa")
    paths = [*figures.iterdir(), *(source/f"tables/v{VERSION}").iterdir(),
             Path(__file__).resolve(), root/"styles/bmj.mplstyle",
             root/"scripts/bmj_rmr_v042.py", root/"scripts/bmj_rmr_v1110_visuals.py",
             analysis/"teaching_values.csv", analysis/"figure_captions.csv",
             root/"analysis/bmj_rmr_v042/branch_entropy_all_primary.csv"]
    pd.DataFrame([{"path":p.relative_to(root).as_posix(),"bytes":p.stat().st_size,
                   "sha256":hashlib.sha256(p.read_bytes()).hexdigest()} for p in paths if p.is_file()]).to_csv(
        analysis/"figure_source_and_artifact_hashes.csv",index=False,lineterminator="\n")
    print(values[["starting_probability","information_bits","fraction_entropy_removed"]].to_string(index=False))


if __name__ == "__main__":
    parser = argparse.ArgumentParser(description=__doc__)
    parser.add_argument("--root", type=Path, default=Path(__file__).resolve().parents[1])
    build(parser.parse_args().root.resolve())
```
