## Supplementary material for "The information in diagnostic tests": Executable analysis code, derived inputs, source-acquisition records, tests, and release metadata: bmj_rmr_v1130_robustness.py

```
"""Supplementary review-weighting and whole-review bootstrap checks (A4/A5)."""
import argparse
import hashlib
import json
from pathlib import Path

import numpy as np
import pandas as pd

from scripts.bmj_rmr_v1100_bootstrap import ANCHORS, REPLICATES, SOURCE

SEED = 20260913
PRIMARY = "analysis/bmj_rmr_v1100/reference_scale_bootstrap.csv"


def weighted_median(values, weights):
    values, weights = np.asarray(values, dtype=float), np.asarray(weights, dtype=float)
    if (values.ndim != 1 or weights.shape != values.shape or not values.size
            or not np.isfinite(values).all() or not np.isfinite(weights).all()
            or (weights <= 0).any()):
        raise ValueError("Expected finite one-dimensional values and positive matching weights")
    order = np.argsort(values, kind="stable")
    values, weights = values[order], weights[order]
    cumulative = np.cumsum(weights)
    half = weights.sum() / 2
    index = int(np.searchsorted(cumulative, half - 1e-12))
    if index + 1 < len(values) and abs(cumulative[index] - half) <= 1e-12:
        return float((values[index] + values[index + 1]) / 2)
    return float(values[index])


def review_resample_medians(values, groups, draws):
    return np.asarray([np.median(values[np.concatenate([groups[i] for i in draw])], axis=0)
                       for draw in draws])


def run(root, output):
    output.mkdir(parents=True, exist_ok=True)
    if (output / "review_robustness.csv").exists():
        raise FileExistsError("Use a new output directory; do not overwrite released analyses")
    frame = pd.read_csv(root / SOURCE)
    assert len(frame) == 819
    assert frame.review_id.notna().all() and frame.source.notna().all()
    assert not frame.duplicated(["operating_point_id", "starting_probability"]).any()
    assert frame.groupby("review_id").source.nunique().eq(1).all()
    assert frame.groupby("operating_point_id").review_id.nunique().eq(1).all()
    assert set(frame.starting_probability) == set(ANCHORS)
    assert frame.groupby("starting_probability").operating_point_id.nunique().eq(273).all()
    assert frame.groupby("starting_probability").review_id.nunique().eq(210).all()
    values = frame.pivot(index="operating_point_id", columns="starting_probability",
                         values="fraction_entropy_removed").loc[:, list(ANCHORS)]
    profiles = frame.drop_duplicates("operating_point_id").set_index("operating_point_id").loc[values.index]
    assert profiles.study_rows.sum() == 4104
    values = values.to_numpy(dtype=float)
    assert values.shape == (273, 3) and np.isfinite(values).all()
    assert ((values >= 0) & (values <= 1)).all()
    counts = profiles.groupby(["review_id", "source"]).size().rename("n_profiles").reset_index()
    counts = counts.sort_values("review_id").reset_index(drop=True)
    review_ids = counts.review_id.to_list()
    weights = 1 / profiles.review_id.map(counts.set_index("review_id").n_profiles).to_numpy()
    assert np.isclose(weights.sum(), 210, rtol=0, atol=1e-12)
    groups = [np.flatnonzero(profiles.review_id.to_numpy() == review) for review in review_ids]
    assert all(np.isclose(weights[group].sum(), 1) for group in groups)
    draws = np.random.default_rng(SEED).integers(0, len(groups), size=(REPLICATES, len(groups)))
    medians = review_resample_medians(values, groups, draws)
    intervals = np.percentile(medians, [2.5, 97.5], axis=0, method="linear")
    primary = pd.read_csv(root / PRIMARY).set_index("starting_probability")
    rows = []
    for index, prior in enumerate(ANCHORS):
        median = float(np.median(values[:, index]))
        alternate = weighted_median(values[:, index], weights)
        assert np.isclose(median, primary.loc[prior, "median_fraction"], rtol=0, atol=1e-14)
        rows.append(dict(starting_probability=prior, n_profiles=273, n_reviews=210,
                         primary_median_fraction=median, equal_review_median_fraction=alternate,
                         difference_percentage_points=100 * (alternate - median),
                         primary_ci_low_fraction=float(primary.loc[prior, "ci_low_fraction"]),
                         primary_ci_high_fraction=float(primary.loc[prior, "ci_high_fraction"]),
                         review_ci_low_fraction=float(intervals[0, index]),
                         review_ci_high_fraction=float(intervals[1, index])))
    table = pd.DataFrame(rows)
    table.to_csv(output / "review_robustness.csv", index=False, lineterminator="\n")
    counts.to_csv(output / "review_contributions.csv", index=False, lineterminator="\n")
    replicates = pd.DataFrame(medians, columns=[f"median_fraction_{int(p*100)}" for p in ANCHORS])
    replicates.insert(0, "replicate", np.arange(1, REPLICATES + 1))
    replicates["sampled_profile_count"] = np.asarray([len(g) for g in groups])[draws].sum(axis=1)
    replicates.to_csv(output / "review_bootstrap_replicates.csv", index=False, lineterminator="\n")
    summary = dict(analysis_version="1.13.0", seed=SEED, replicates=REPLICATES,
                   numpy_version=np.__version__, review_order="lexically sorted review_id",
                   draws_sha256=hashlib.sha256(draws.astype("<i8").tobytes()).hexdigest(),
                   n_profiles=273, n_reviews=210, single_profile_reviews=int((counts.n_profiles == 1).sum()),
                   multiple_profile_reviews=int((counts.n_profiles > 1).sum()),
                   maximum_profiles_per_review=int(counts.n_profiles.max()),
                   weighted_median="first cumulative weight reaching half; midpoint at exact half (absolute tolerance 1e-12)",
                   bootstrap="ordinary profile median; complete sampled reviews with multiplicities; shared draws across priors; linear percentiles",
                   interpretation="supplementary analyses planned after exploratory results were seen; original primary estimates and intervals retained",
                   limits="within-profile estimation error and primary-study overlap across reviews are not propagated",
                   inputs=[dict(path=p, sha256=hashlib.sha256((root / p).read_bytes()).hexdigest())
                           for p in (SOURCE, PRIMARY)])
    (output / "review_robustness_summary.json").write_bytes((json.dumps(summary, indent=2) + "\n").encode())
    return table


if __name__ == "__main__":
    parser = argparse.ArgumentParser(description=__doc__)
    parser.add_argument("--root", type=Path, default=Path(__file__).resolve().parents[1])
    parser.add_argument("--output", type=Path)
    args = parser.parse_args()
    root = args.root.resolve()
    print(run(root, args.output or root / "analysis/bmj_rmr_v1130").to_string(index=False))
```
