## Supplementary material for "The information in diagnostic tests": Executable analysis code, derived inputs, source-acquisition records, tests, and release metadata: bmj_rmr_v1131_visuals.py

```
"""Repair S1/S2/S9 layout while preserving the published numerical inputs."""
from pathlib import Path
import hashlib
import json

import matplotlib.pyplot as plt
from matplotlib.patches import Rectangle
import numpy as np
import pandas as pd

from scripts import bmj_rmr_v1110_visuals as prior

ROOT = Path(__file__).resolve().parents[1]
base = prior.base
VERSION = "1.13.1"
SPECS = {
    "Figure S1": {"stem": "figure_s1_corpus_and_selection_flow", "height_mm": 185},
    "Figure S2": {"stem": "figure_s2_validation_and_route_stability", "height_mm": 150},
    "Figure S9": {"stem": "figure_s9_result_branch_distributions", "height_mm": 145},
}


def canvas(display, gray):
    base.set_style(gray)
    plt.rcParams.update({"font.size": 8, "axes.labelsize": 8,
                         "xtick.labelsize": 8, "ytick.labelsize": 8})
    return plt.figure(figsize=base.figure_size(SPECS[display]["height_mm"]))


def selection(data, gray=False):
    fig = canvas("Figure S1", gray)
    ax = fig.add_axes([.035, .025, .93, .91])
    ax.set(xlim=(0, 1), ylim=(0, 1)); ax.axis("off")
    blue = "#333333" if gray else prior.BLUE
    roles, trace = data["roles"], data["trace"]
    counts = (int(roles.reviews_with_normalized_rows.sum()), int(roles.model_ready_groups.sum()),
              int(roles.primary_operating_points.sum()), trace.review_id.nunique(), len(trace))
    assert counts == (382, 444, 273, 210, 4104), counts
    nodes = [(0.85, "382 review records", "Parsed structured evidence"),
             (0.68, "1,830 candidate groups", "Source-defined tests or analyses"),
             (0.51, "444 model-ready groups", "All 444 model fits converged"),
             (0.34, "273 pooled profiles", "From 210 contributing reviews"),
             (0.17, "4,127 joined results", "Study-result appearances"),
             (0.00, "4,104 included results", "Final analysis sample")]
    for i, (y, title, subtitle) in enumerate(nodes):
        ax.add_patch(Rectangle((.015, y+.02), .48, .105, fc="#F5F7F9", ec=blue, lw=.9))
        ax.text(.04, y+.085, title, fontsize=9, weight="semibold", color=blue, va="center")
        ax.text(.04, y+.045, subtitle, fontsize=8, va="center")
        if i < 5:
            ax.annotate("", (.255, y-.045), (.255, y+.018),
                        arrowprops={"arrowstyle": "-|>", "lw": .8, "color": "#666666"})
    exclusions = [(.65, "1,386 structurally ineligible", "1,219: fewer than five studies\n155: repeated study identifiers\n11: below chance in source direction\n1: fewer than three studies with both states"),
                  (.39, "171 alternative analyses", "Historical Cochrane: 142\nModern Cochrane: 4\nOSF: 5; PubMed Central: 13; Zenodo: 7\nRetained in the audit record"),
                  (.105, "23 results excluded", "Noncanonical workbook sheet\nRecorded Zenodo selection rule")]
    for y, title, detail in exclusions:
        ax.plot([.51, .55], [y+.022, y+.022], color="#777777", lw=.8)
        ax.text(.565, y+.052, title, weight="semibold", fontsize=8, va="top")
        ax.text(.565, y+.015, detail, fontsize=7.5, va="top", linespacing=1.45)
    fig.text(.05, .965, "How the reference sample was selected", fontsize=9.5, weight="semibold")
    return fig


def fidelity(data, gray=False):
    fig = canvas("Figure S2", gray)
    palette = base.colors(gray)
    left = fig.add_axes([.10, .40, .32, .41])
    right = fig.add_axes([.70, .40, .265, .41])
    frame = data["fidelity"]
    for field, marker, color in (("sensitivity", "o", palette[0]), ("specificity", "s", palette[1])):
        left.scatter(frame["published_"+field], frame[field], s=18, marker=marker,
                     facecolors="none" if marker == "o" else color, edgecolors=color,
                     alpha=.7, linewidth=.8, label=field.capitalize())
    large = frame.large_discrepancy.astype(str).str.lower().eq("true")
    for field in ("sensitivity", "specificity"):
        left.scatter(frame.loc[large, "published_"+field], frame.loc[large, field],
                     s=40, marker="x", color=palette[3], linewidth=1)
    left.plot([0, 1], [0, 1], color="#777777", ls="--", lw=.8)
    left.set(xlim=(0, 1.01), ylim=(0, 1.01), xlabel="Source-reported pooled value", ylabel="Primary REML estimate")
    left.set_title(f"A  Source agreement (n={len(frame)})", loc="left", fontsize=8.5, pad=9)
    left.legend(loc="lower right", fontsize=7.5, frameon=False)
    fig.text(.10, .30, "Crosses: absolute difference of at least 0.10", fontsize=7.5)
    frame = data["routes"].loc[np.isclose(data["routes"].starting_probability, .2)].copy()
    order = ["historical Cochrane", "modern Cochrane", "OSF", "Zenodo", "PMC"]
    frame = frame.set_index("cohort").loc[order]
    y = np.arange(len(frame))
    right.hlines(y, frame.information_bits_q1, frame.information_bits_q3, lw=3, color=palette[1])
    right.scatter(frame.information_bits_median, y, s=28, marker="D", color=palette[1], zorder=3)
    for i, row in enumerate(frame.itertuples()):
        right.text(row.information_bits_q3+.009, i, f"n={int(row.operating_points)}", va="center", fontsize=7.5)
    right.set(yticks=y, yticklabels=["Historical Cochrane", "Modern Cochrane", "OSF", "Zenodo", "PubMed Central"],
              xlabel="Information (bits)", xlim=(0, max(frame.information_bits_q3)+.13), ylim=(-.6,4.6))
    right.invert_yaxis()
    right.set_title("C  Source routes at 20% prior", loc="left", fontsize=8.5, pad=9)
    fig.text(.62, .30, "Diamonds: medians; bars: interquartile ranges", fontsize=7.5)
    audit = data["audit_summary"]
    fig.text(.10, .215, "B  Recorded author audit", fontsize=8.5, weight="semibold")
    fig.text(.10, .15, f"Transcription: {audit['transcription_error_numerator']}/{audit['transcription_error_denominator_cells']} cells with errors", fontsize=8.5)
    fig.text(.55, .15, f"Acceptance: {audit['acceptance_error_numerator']}/{audit['acceptance_error_denominator_decisions']} decisions with errors", fontsize=8.5)
    fig.text(.10, .08, "OSF = Open Science Framework. Audits were conducted by the authors.", fontsize=8)
    fig.text(.05, .945, "Source agreement and route-specific reference values", fontsize=9.5, weight="semibold")
    return fig


def branches(data, gray=False):
    # Reuse the original violin, jitter and summary calculations; relocate only text/layout.
    fig = base.figure_4(data["branches"], data["branch_summary"], gray)
    fig.set_size_inches(*base.figure_size(SPECS["Figure S9"]["height_mm"]))
    for text in list(fig.texts):
        text.remove()
    for i, ax in enumerate(fig.axes):
        note = next(t for t in ax.texts if t.get_text().startswith("Entropy increase"))
        label = note.get_text().replace("Entropy increase", "Profiles with entropy increase")
        note.remove()
        ax.set_position([.10+i*.305, .35, .255, .46])
        ax.set_xlabel("")
        ax.set_title(f"{'ABC'[i]}  Starting probability {int([5,20,50][i])}%", loc="left", fontsize=8.5, pad=10)
        note = fig.text(.10+i*.305, .17, label, fontsize=7.5, va="top", linespacing=1.3)
        note.set_gid("branch_count_annotation")
    fig.text(.055, .94, "One result can increase uncertainty", fontsize=9.5, weight="semibold")
    fig.text(.055, .035, "Entropy change = posterior minus prior. Each panel includes all 273 profiles.", fontsize=8)
    fig.canvas.draw()
    for text in fig.texts:
        if text.get_gid() == "branch_count_annotation":
            assert not any(text.get_window_extent().overlaps(ax.get_window_extent()) for ax in fig.axes)
    return fig


def build(root=ROOT):
    data = prior.load_data(root)
    data["roles"] = pd.read_csv(root/"analysis/full_atlas/v9/source_role_manifest.csv")
    data["trace"] = pd.read_csv(root/"analysis/bmj_rmr_v042/source_trace_all_primary.csv")
    output = root/"docs/manuscript/v31/figures/v1.13.1"
    analysis = root/"analysis/bmj_rmr_v1131"
    analysis.mkdir(parents=True, exist_ok=True)
    base.VERSION, base.RELEASE_DATE = VERSION, "2026-09-13"
    base.FIGURE_SPECS = {**base.FIGURE_SPECS, **SPECS}
    for display, fn in (("Figure S1", selection), ("Figure S2", fidelity), ("Figure S9", branches)):
        for gray in (False, True):
            base.save_figure(fn(data, gray), output, display, gray)
    captions = pd.read_csv(root/"analysis/bmj_rmr_v1120/figure_captions.csv")
    captions.loc[captions.display.eq("Figure 2"), "caption"] += " HINTS denotes head impulse, nystagmus and test of skew."
    captions.loc[captions.display.eq("Figure 3"), "caption"] += " CEA denotes carcinoembryonic antigen."
    for display in ("Figure 4", "Figure S9", "Figure S10"):
        captions.loc[captions.display.eq(display), "caption"] += " KL denotes Kullback-Leibler divergence."
    captions.loc[captions.display.eq("Figure S1"), "caption"] += " The diagram describes selection within the accessible source collection, not a comprehensive search of diagnostic medicine."
    captions.to_csv(analysis/"figure_captions.csv",index=False,lineterminator="\n")
    (output/"FIGURE_CAPTIONS.md").write_bytes(("# Figure captions\n\n"+"\n\n".join(captions.caption)+"\n").encode())
    paths = [*output.iterdir(), Path(__file__).resolve(), root/"analysis/bmj_rmr_v042/branch_entropy_all_primary.csv",
             root/"analysis/bmj_rmr_v042/source_summary_fidelity.csv", root/"analysis/bmj_rmr_v042/source_route_stability.csv"]
    pd.DataFrame([{"path":p.relative_to(root).as_posix(),"bytes":p.stat().st_size,
                   "sha256":hashlib.sha256(p.read_bytes()).hexdigest()} for p in paths if p.is_file()]).to_csv(
        analysis/"figure_source_and_artifact_hashes.csv",index=False,lineterminator="\n")
    base.audit_pdf_text_bounds(output, analysis/"qa")
    print(json.dumps({"version":VERSION,"changed_figures":list(SPECS),"S9_annotations_outside_data":True}))


if __name__ == "__main__":
    build()
```
