## Supplementary material for "The information in diagnostic tests": Executable analysis code, derived inputs, source-acquisition records, tests, and release metadata: bmj_rmr_v1133_visuals.py

```
"""Replace production terminology in four supplementary figures; retain all data."""
from pathlib import Path

import matplotlib.pyplot as plt
import pandas as pd

from scripts import bmj_rmr_v1110_visuals as current
from scripts import bmj_rmr_v1131_visuals as previous

REPLACEMENTS = {
    "Parsed structured evidence": "Structured review data",
    "4,127 joined results": "4,127 linked results",
    "Historical Cochrane": "2018 Cochrane corpus",
    "Modern Cochrane": "Recent Cochrane reviews",
    "Retained in the audit record": "Documented alternative analyses",
    "Noncanonical workbook sheet": "Alternative extraction sheet",
    "limited smoke diagnostic": "limited numerical check",
    "Historical REML": "Alternative REML",
    "The CEA crossings are inherited model-specific examples.": "CEA crossings depend on the synthesis model.",
    "One dataset per 24 scenarios:": "24 scenarios, one dataset each:",
}


def apply_labels(figure):
    before = [(a.get_xlim(), a.get_ylim(), [line.get_xydata().tobytes() for line in a.lines]) for a in figure.axes]
    changed = 0
    for axes in figure.axes:
        for dimension in ("x", "y"):
            labels = [label.get_text() for label in getattr(axes, f"get_{dimension}ticklabels")()]
            updated = labels[:]
            for old, new in REPLACEMENTS.items():
                updated = [label.replace(old, new) for label in updated]
            if updated != labels:
                getattr(axes, f"set_{dimension}ticks")(getattr(axes, f"get_{dimension}ticks")(), labels=updated)
                changed += sum(a != b for a, b in zip(labels, updated))
    for label in figure.findobj(match=plt.Text):
        original = label.get_text()
        text = original
        for old, new in REPLACEMENTS.items():
            text = text.replace(old, new)
        if text != original:
            label.set_text(text)
            changed += 1
    after = [(a.get_xlim(), a.get_ylim(), [line.get_xydata().tobytes() for line in a.lines]) for a in figure.axes]
    assert before == after, "Text edits changed plotted data"
    return changed


def prepare(root):
    data = current.load_data(root)
    data["roles"] = pd.read_csv(root / "analysis/full_atlas/v9/source_role_manifest.csv")
    data["trace"] = pd.read_csv(root / "analysis/bmj_rmr_v042/source_trace_all_primary.csv")
    base = current.base
    base.VERSION = "1.13.3"
    base.FIGURE_SPECS = {**current.FIGURE_SPECS, **previous.SPECS}
    functions = {
        "Figure S1": lambda g: previous.selection(data, g),
        "Figure S2": lambda g: previous.fidelity(data, g),
        "Figure S4": lambda g: base.figure_s4(data["recovery"], data["recovery_summary"], data["diagnostics"], data["diagnostic_summary"], g),
        "Figure S8": lambda g: base.figure_s8(data["crossings"], data["cea_profiles"], data["theory_summary"], g),
    }
    for display, function in functions.items():
        for gray in (False, True):
            figure = function(gray)
            changed = apply_labels(figure)
            assert changed, display
            base.save_figure(figure, root / "docs/manuscript/v33/figures/v1.13.1", display, gray)
            print(display, "grayscale" if gray else "colour", changed, "labels; plotted values unchanged")


if __name__ == "__main__":
    prepare(Path(__file__).resolve().parents[1])
```
