## Supplementary material for "The information in diagnostic tests": Executable analysis code, derived inputs, source-acquisition records, tests, and release metadata: cochrane_dta_atlas.py

```
"""Build the full Cochrane diagnostic information-action atlas."""

from __future__ import annotations

import argparse
import csv
import hashlib
import html
import io
import json
import math
import re
import shutil
import urllib.request
import xml.etree.ElementTree as ET
import zipfile
from collections import defaultdict
from itertools import combinations
from pathlib import Path

import matplotlib

matplotlib.use("Agg")
import matplotlib.pyplot as plt
import numpy as np
from matplotlib.patches import FancyArrowPatch, FancyBboxPatch
from scipy.optimize import brentq, minimize
from scipy.special import expit, gammaln
from scipy.stats import spearmanr

try:
    from scripts.hints_information_action import (
        BLUE,
        GREEN,
        INK,
        ORANGE,
        PURPLE,
        _save_figure,
        _style,
        _write_csv,
        binary_entropy,
        diagnostic_metrics,
        logistic,
        maximum_pretest_probability,
    )
except ModuleNotFoundError:  # supports `python scripts/cochrane_dta_atlas.py`
    from hints_information_action import (
        BLUE,
        GREEN,
        INK,
        ORANGE,
        PURPLE,
        _save_figure,
        _style,
        _write_csv,
        binary_entropy,
        diagnostic_metrics,
        logistic,
        maximum_pretest_probability,
    )


VERSION = "0.26.0"
SEED = 20260816
DEFAULT_DRAWS = 20_000
PREVALENCES = tuple(np.linspace(0.01, 0.50, 50))
PRIOR_CROSSING_GRID = tuple(np.linspace(0.005, 0.995, 991))
THRESHOLDS = (0.001, 0.0025, 0.005, 0.01, 0.02, 0.05)
REFERENCE_URL = (
    "https://zenodo.org/api/records/1303259/files/"
    "CL145_open_set_20181101.zip/content"
)
REFERENCE_RECORD_URL = "https://zenodo.org/records/1303259"
REFERENCE_SHA256 = "f921cf6f1a9236c86f89e7c646497639f45e8ff8fcf56008c1b27422829fec4f"
REFERENCE_MD5 = "04b59deabfedba64cc6d5f2d2b77b467"
FINAL_DOIS = {
    "CD014911": "10.1002/14651858.CD014911.pub2",
    "CD015089": "10.1002/14651858.CD015089.pub2",
}


def _file_hash(path: Path, algorithm: str = "sha256") -> str:
    digest = hashlib.new(algorithm)
    with path.open("rb") as handle:
        for block in iter(lambda: handle.read(1024 * 1024), b""):
            digest.update(block)
    return digest.hexdigest()


def fetch_reference_archive(path: Path) -> Path:
    """Download and verify the fixed Zenodo source when explicitly requested."""
    if path.exists():
        if _file_hash(path) != REFERENCE_SHA256:
            raise ValueError(f"Unexpected reference archive SHA-256: {_file_hash(path)}")
        return path
    path.parent.mkdir(parents=True, exist_ok=True)
    temporary = path.with_suffix(path.suffix + ".tmp")
    request = urllib.request.Request(REFERENCE_URL, headers={"User-Agent": "clinical-uncertainty/0.13.0"})
    try:
        with urllib.request.urlopen(request, timeout=120) as response, temporary.open("wb") as handle:
            shutil.copyfileobj(response, handle)
        observed = _file_hash(temporary)
        if observed != REFERENCE_SHA256:
            raise ValueError(f"Unexpected downloaded archive SHA-256: {observed}")
        temporary.replace(path)
    finally:
        temporary.unlink(missing_ok=True)
    return path


def _read_csv(archive: zipfile.ZipFile, suffix: str) -> list[dict[str, str]]:
    name = next(name for name in archive.namelist() if name.endswith(suffix))
    with archive.open(name) as handle:
        return list(csv.DictReader(io.TextIOWrapper(handle, encoding="utf-8-sig")))


def _read_text(archive: zipfile.ZipFile, suffix: str) -> str:
    name = next(name for name in archive.namelist() if name.endswith(suffix))
    return archive.read(name).decode("utf-8-sig")


def _strip_tags(value: str) -> str:
    return " ".join(html.unescape(re.sub(r"<[^>]+>", " ", value)).split())


def package_identity(package: Path, archive: zipfile.ZipFile) -> dict[str, str]:
    review_id_match = re.search(r"CD\d+", package.name, flags=re.I)
    if not review_id_match:
        raise ValueError(f"Cannot infer Cochrane review ID from {package.name}")
    review_id = review_id_match.group(0).upper()
    info = _read_text(archive, "data-package-info.html")
    title_match = re.search(r"<h1>(.*?)</h1>", info, flags=re.I | re.S)
    byline_match = re.search(r'<span class="byline">(.*?)</span>', info, flags=re.I | re.S)
    if not title_match:
        raise ValueError(f"No review title in {package.name}")
    return {
        "review_id": review_id,
        "title": _strip_tags(title_match.group(1)),
        "authors": _strip_tags(byline_match.group(1)) if byline_match else "",
        "doi": FINAL_DOIS.get(review_id, ""),
    }


def analysis_scope(parameter: dict[str, str]) -> str:
    subgroup = parameter.get("Subgroup by", "")
    return "subgroup" if subgroup not in ("", "None") or " by " in parameter["Analysis name"].lower() else "main"


def eligible_parameter(parameter: dict[str, str]) -> bool:
    return bool(parameter.get("E(logitSe)") and parameter.get("E(logitSp)"))


def _element_values(element: ET.Element | None) -> dict[str, str]:
    if element is None:
        return {}
    return {
        value.get("name", ""): (value.text or "").strip()
        for value in element.findall("./{*}value")
    }


def _counts(values: dict[str, str]) -> dict[str, int] | None:
    try:
        counts = {name: int(values[name]) for name in ("tp", "fp", "fn", "tn")}
    except (KeyError, ValueError):
        return None
    if min(counts.values()) < 0 or sum(counts.values()) == 0:
        return None
    return counts


def _finite_number(value: object) -> bool:
    try:
        return math.isfinite(float(value))
    except (TypeError, ValueError):
        return False


def _eligibility(rows: list[dict]) -> str:
    study_ids = [str(row["study_id"]) for row in rows]
    if len(set(study_ids)) < 5:
        return "fewer_than_five_unique_studies"
    if len(study_ids) != len(set(study_ids)):
        return "repeated_study_rows"
    paired = sum(row["tp"] + row["fn"] > 0 and row["tn"] + row["fp"] > 0 for row in rows)
    if paired < 3:
        return "fewer_than_three_paired_studies"
    tp = sum(row["tp"] for row in rows)
    fp = sum(row["fp"] for row in rows)
    fn = sum(row["fn"] for row in rows)
    tn = sum(row["tn"] for row in rows)
    if tp / (tp + fn) + tn / (tn + fp) < 1.0:
        return "below_chance_orientation"
    return "eligible"


def load_legacy_reference(path: Path) -> tuple[list[dict], list[dict], list[dict]]:
    observed = _file_hash(path)
    if observed != REFERENCE_SHA256:
        raise ValueError(f"Unexpected reference archive SHA-256: {observed}")
    with zipfile.ZipFile(path) as archive:
        xml_name = next(name for name in archive.namelist() if name.endswith("data.xml"))
        root = ET.parse(archive.open(xml_name)).getroot()

    reviews: list[dict] = []
    groups: list[dict] = []
    normalized_rows: list[dict] = []
    for review in root.findall(".//{*}review"):
        review_id = review.findtext("./{*}review_id") or review.get("id", "")
        title = review.findtext("./{*}title") or ""
        findings = {}
        finding_elements = review.findall("./{*}findings/{*}test")
        for finding in finding_elements:
            finding_id = finding.findtext("./{*}test_id") or ""
            values = _element_values(finding.find("./{*}result"))
            findings[finding_id] = {
                "description": finding.findtext("./{*}desc") or "",
                "studies": finding.findtext("./{*}n_studies") or "",
                "sensitivity": values.get("sensitivity", ""),
                "sensitivity_low": values.get("sensitivity_0", ""),
                "sensitivity_high": values.get("sensitivity_1", ""),
                "specificity": values.get("specificity", ""),
                "specificity_low": values.get("specificity_0", ""),
                "specificity_high": values.get("specificity_1", ""),
            }

        review_groups = []
        raw_result_count = 0
        for sequence, test in enumerate(review.findall("./{*}tests/{*}test"), start=1):
            raw_name = test.findtext("./{*}name") or f"Test {sequence}"
            match = re.match(r"\s*(\d+)\s+(.*)", raw_name, flags=re.S)
            test_id = match.group(1) if match else str(sequence)
            test_name = " ".join((match.group(2) if match else raw_name).split())
            finding = findings.get(test_id, {})
            reported = _finite_number(finding.get("sensitivity")) and _finite_number(
                finding.get("specificity")
            )
            group_id = f"legacy:{review_id}:{test_id}"
            rows = []
            results = test.findall("./{*}result")
            raw_result_count += len(results)
            for result in results:
                values = _element_values(result)
                counts = _counts(values)
                if counts is None:
                    continue
                status = "corrected" if any(
                    value.get("status") == "CORRECTED" for value in result.findall("./{*}value")
                ) else "ok"
                rows.append(
                    {
                        "source": "Cochrane DTA Reference Dataset",
                        "review_id": review_id,
                        "review_title": title,
                        "group_id": group_id,
                        "group_number": test_id,
                        "test_name": finding.get("description") or test_name,
                        "study_id": values.get("study_id", ""),
                        **counts,
                        "source_status": status,
                    }
                )
            reason = _eligibility(rows)
            scope = "reported_main" if reported else "extracted_secondary"
            primary = reason == "eligible" and reported
            for row in rows:
                row.update({"analysis_scope": scope, "primary_atlas": primary})
            normalized_rows.extend(rows)
            group = {
                "source": "Cochrane DTA Reference Dataset",
                "source_version": "CL145_open_set_20181101",
                "review_id": review_id,
                "review_title": title,
                "group_id": group_id,
                "group_number": test_id,
                "test_name": finding.get("description") or test_name,
                "analysis_scope": scope,
                "study_rows": len(rows),
                "unique_studies": len({row["study_id"] for row in rows}),
                "paired_studies": sum(
                    row["tp"] + row["fn"] > 0 and row["tn"] + row["fp"] > 0 for row in rows
                ),
                "eligibility": reason,
                "primary_atlas": primary,
                "published_studies": finding.get("studies", ""),
                "published_sensitivity": finding.get("sensitivity", ""),
                "published_sensitivity_low": finding.get("sensitivity_low", ""),
                "published_sensitivity_high": finding.get("sensitivity_high", ""),
                "published_specificity": finding.get("specificity", ""),
                "published_specificity_low": finding.get("specificity_low", ""),
                "published_specificity_high": finding.get("specificity_high", ""),
                "rows": rows,
            }
            groups.append(group)
            review_groups.append(group)

        included = review.findall("./{*}included/{*}reference")
        reviews.append(
            {
                "source": "Cochrane DTA Reference Dataset",
                "source_version": "CL145_open_set_20181101",
                "review_id": review_id,
                "review_title": title,
                "doi": "",
                "source_file": path.name,
                "source_sha256": observed,
                "included_study_records": len(included),
                "test_groups": len(review_groups),
                "stored_study_test_rows": raw_result_count,
                "reported_findings": len(finding_elements),
                "model_candidates": sum(group["eligibility"] == "eligible" for group in review_groups),
                "primary_candidates": sum(group["primary_atlas"] for group in review_groups),
            }
        )
    return reviews, groups, normalized_rows


def load_modern_packages(packages: list[Path]) -> tuple[list[dict], list[dict], list[dict]]:
    reviews: list[dict] = []
    groups: list[dict] = []
    normalized_rows: list[dict] = []
    for package in packages:
        digest = _file_hash(package)
        with zipfile.ZipFile(package) as archive:
            identity = package_identity(package, archive)
            studies = _read_csv(archive, "study-information.csv")
            test_rows = _read_csv(archive, "study-test-data.csv")
            data_rows = _read_csv(archive, "data-rows.csv")
            parameters = _read_csv(archive, "parameters.csv")
        eligible = [parameter for parameter in parameters if eligible_parameter(parameter)]
        review_groups = []
        for parameter in eligible:
            analysis_number = parameter["Analysis number"]
            group_id = f"modern:{identity['review_id']}:{analysis_number}"
            scope = analysis_scope(parameter)
            rows = []
            for source_row in data_rows:
                if source_row["Analysis number"] != analysis_number:
                    continue
                values = {
                    "tp": source_row.get("TP", ""),
                    "fp": source_row.get("FP", ""),
                    "fn": source_row.get("FN", ""),
                    "tn": source_row.get("TN", ""),
                }
                counts = _counts(values)
                if counts is None:
                    continue
                rows.append(
                    {
                        "source": "Modern Cochrane data package",
                        "review_id": identity["review_id"],
                        "review_title": identity["title"],
                        "group_id": group_id,
                        "group_number": analysis_number,
                        "test_name": parameter["Test name"],
                        "analysis_scope": scope,
                        "primary_atlas": scope == "main",
                        "study_id": source_row["Study"],
                        **counts,
                        "source_status": "ok",
                    }
                )
            reason = _eligibility(rows)
            primary = reason == "eligible" and scope == "main"
            for row in rows:
                row["primary_atlas"] = primary
            normalized_rows.extend(rows)
            group = {
                "source": "Modern Cochrane data package",
                "source_version": package.name,
                "review_id": identity["review_id"],
                "review_title": identity["title"],
                "group_id": group_id,
                "group_number": analysis_number,
                "test_name": parameter["Test name"],
                "analysis_scope": scope,
                "study_rows": len(rows),
                "unique_studies": len({row["study_id"] for row in rows}),
                "paired_studies": sum(
                    row["tp"] + row["fn"] > 0 and row["tn"] + row["fp"] > 0 for row in rows
                ),
                "eligibility": reason,
                "primary_atlas": primary,
                "published_studies": parameter.get("Studies", ""),
                "published_sensitivity": float(logistic(float(parameter["E(logitSe)"]))),
                "published_sensitivity_low": "",
                "published_sensitivity_high": "",
                "published_specificity": float(logistic(float(parameter["E(logitSp)"]))),
                "published_specificity_low": "",
                "published_specificity_high": "",
                "rows": rows,
            }
            groups.append(group)
            review_groups.append(group)
        reviews.append(
            {
                "source": "Modern Cochrane data package",
                "source_version": package.name,
                "review_id": identity["review_id"],
                "review_title": identity["title"],
                "doi": identity["doi"],
                "source_file": package.name,
                "source_sha256": digest,
                "included_study_records": len(studies),
                "test_groups": len({row["Test number"] for row in test_rows}),
                "stored_study_test_rows": len(test_rows),
                "reported_findings": len(eligible),
                "model_candidates": sum(group["eligibility"] == "eligible" for group in review_groups),
                "primary_candidates": sum(group["primary_atlas"] for group in review_groups),
            }
        )
    return reviews, groups, normalized_rows


def _csv_bool(value: object) -> bool:
    return str(value).strip().lower() in {"1", "true", "yes"}


def load_external_repository_dirs(
    directories: list[Path],
) -> tuple[list[dict], list[dict], list[dict], list[dict]]:
    reviews: list[dict] = []
    groups: list[dict] = []
    normalized_rows: list[dict] = []
    source_audits: list[dict] = []
    seen_signatures: set[tuple] = set()
    for directory in directories:
        summary_path = directory / "summary.json"
        if summary_path.exists():
            source_audits.append(json.loads(summary_path.read_text(encoding="utf-8")))
        group_path = directory / "test_group_manifest.csv"
        row_path = directory / "normalized_study_data.csv"
        if not group_path.exists() or group_path.stat().st_size == 0:
            continue
        source_groups = list(csv.DictReader(group_path.read_text(encoding="utf-8-sig").splitlines()))
        source_groups = [
            group for group in source_groups
            if group.get("selection_status", "canonical") == "canonical"
            and group.get("analysis_scope") != "external_method_dataset"
        ]
        source_rows = list(csv.DictReader(row_path.read_text(encoding="utf-8-sig").splitlines()))
        by_group: dict[str, list[dict]] = defaultdict(list)
        for row in source_rows:
            parsed = {
                **row,
                "tp": int(row["tp"]),
                "fp": int(row["fp"]),
                "fn": int(row["fn"]),
                "tn": int(row["tn"]),
                "primary_atlas": _csv_bool(row["primary_atlas"]),
            }
            by_group[parsed["group_id"]].append(parsed)
        for group in source_groups:
            group_rows = by_group[group["group_id"]]
            signature = tuple(sorted(
                (row["study_id"].strip().lower(), row["tp"], row["fp"], row["fn"], row["tn"])
                for row in group_rows
            ))
            if signature in seen_signatures:
                continue
            seen_signatures.add(signature)
            parsed = {
                **group,
                "study_rows": int(group["study_rows"]),
                "unique_studies": int(group["unique_studies"]),
                "paired_studies": int(group["paired_studies"]),
                "primary_atlas": _csv_bool(group["primary_atlas"]),
                "published_sensitivity": float(group["published_sensitivity"]) if group["published_sensitivity"] else "",
                "published_specificity": float(group["published_specificity"]) if group["published_specificity"] else "",
                "rows": group_rows,
            }
            groups.append(parsed)
            normalized_rows.extend(group_rows)

    for review_id in sorted({group["review_id"] for group in groups}):
        review_groups = [group for group in groups if group["review_id"] == review_id]
        review_rows = [row for row in normalized_rows if row["review_id"] == review_id]
        first = review_groups[0]
        reviews.append(
            {
                "source": first["source"],
                "source_version": VERSION,
                "review_id": review_id,
                "review_title": first["review_title"],
                "doi": "",
                "source_file": " | ".join(sorted({group["source_file"] for group in review_groups})),
                "source_sha256": " | ".join(sorted({group["source_sha256"] for group in review_groups})),
                "included_study_records": len({row["study_id"] for row in review_rows}),
                "test_groups": len(review_groups),
                "stored_study_test_rows": len(review_rows),
                "reported_findings": 0,
                "model_candidates": sum(group["eligibility"] == "eligible" for group in review_groups),
                "primary_candidates": sum(bool(group["primary_atlas"]) for group in review_groups),
            }
        )
    return reviews, groups, normalized_rows, source_audits


def fit_bivariate_reml(
    rows: list[dict], *, continuity_correction: str = "universal"
) -> dict[str, object]:
    """Fit a continuity-corrected bivariate normal random-effects model.

    ``universal`` preserves the historical implementation by adding 0.5 to
    every cell of every study. ``zero_cell_triggered`` is a direct sensitivity
    that adds 0.5 to all four cells only when that study contains at least one
    zero; zero-free studies are left uncorrected.
    """
    if continuity_correction not in {"universal", "zero_cell_triggered"}:
        raise ValueError(f"Unknown continuity-correction rule: {continuity_correction}")
    outcomes: list[np.ndarray] = []
    designs: list[np.ndarray] = []
    within: list[np.ndarray] = []
    paired_logits = []
    for row in rows:
        tp, fp, fn, tn = (float(row[name]) for name in ("tp", "fp", "fn", "tn"))
        correction = (
            0.5
            if continuity_correction == "universal"
            or any(value == 0 for value in (tp, fp, fn, tn))
            else 0.0
        )
        values, design, variances = [], [], []
        if tp + fn > 0:
            values.append(math.log((tp + correction) / (fn + correction)))
            design.append([1.0, 0.0])
            variances.append(1.0 / (tp + correction) + 1.0 / (fn + correction))
        if tn + fp > 0:
            values.append(math.log((tn + correction) / (fp + correction)))
            design.append([0.0, 1.0])
            variances.append(1.0 / (tn + correction) + 1.0 / (fp + correction))
        outcomes.append(np.asarray(values))
        designs.append(np.asarray(design))
        within.append(np.diag(variances))
        if len(values) == 2:
            paired_logits.append(values)

    paired = np.asarray(paired_logits)
    empirical = np.cov(paired.T, ddof=1) if len(paired) > 2 else np.eye(2) * 0.2
    if not np.isfinite(empirical).all():
        empirical = np.eye(2) * 0.2
    paired_within = [matrix for matrix in within if matrix.shape == (2, 2)]
    empirical -= np.diag(
        [
            np.median([matrix[0, 0] for matrix in paired_within]),
            np.median([matrix[1, 1] for matrix in paired_within]),
        ]
    )
    eigenvalues, eigenvectors = np.linalg.eigh(empirical)
    initial_covariance = eigenvectors @ np.diag(np.maximum(eigenvalues, 0.02)) @ eigenvectors.T
    initial_cholesky = np.linalg.cholesky(initial_covariance)
    initial = np.array(
        [
            math.log(initial_cholesky[0, 0]),
            initial_cholesky[1, 0],
            math.log(initial_cholesky[1, 1]),
        ]
    )

    def evaluate(parameters: np.ndarray, full: bool = False):
        cholesky = np.array(
            [[math.exp(parameters[0]), 0.0], [parameters[1], math.exp(parameters[2])]]
        )
        between = cholesky @ cholesky.T
        precision = np.zeros((2, 2))
        weighted = np.zeros(2)
        log_determinant = 0.0
        blocks = []
        for outcome, design, sampling in zip(outcomes, designs, within):
            covariance = sampling + design @ between @ design.T
            sign, determinant = np.linalg.slogdet(covariance)
            if sign <= 0:
                return 1e100
            inverse = np.linalg.inv(covariance)
            precision += design.T @ inverse @ design
            weighted += design.T @ inverse @ outcome
            log_determinant += determinant
            blocks.append((outcome, design, inverse))
        try:
            mean_covariance = np.linalg.inv(precision)
        except np.linalg.LinAlgError:
            return 1e100
        mean = mean_covariance @ weighted
        residual = sum(
            (outcome - design @ mean) @ inverse @ (outcome - design @ mean)
            for outcome, design, inverse in blocks
        )
        objective = 0.5 * (
            log_determinant + np.linalg.slogdet(precision)[1] + residual
        )
        return (objective, mean, mean_covariance, between) if full else objective

    bounds = [(-8.0, 5.0), (-20.0, 20.0), (-8.0, 5.0)]
    hard_bound_atol = 1e-6
    starts = [
        ("empirical", initial),
        ("minus2_zero_minus2", np.asarray([-2.0, 0.0, -2.0])),
        ("zero_zero_zero", np.asarray([0.0, 0.0, 0.0])),
        ("one_zero_one", np.asarray([1.0, 0.0, 1.0])),
        ("lower_zero_lower", np.asarray([-8.0, 0.0, -8.0])),
        ("minus1_plus2_minus1", np.asarray([-1.0, 2.0, -1.0])),
        ("minus1_minus2_minus1", np.asarray([-1.0, -2.0, -1.0])),
    ]

    def projected_kkt_gradient(
        parameters: np.ndarray, gradient: np.ndarray
    ) -> np.ndarray:
        projected = np.asarray(gradient, dtype=float).copy()
        for index, (lower, upper) in enumerate(bounds):
            if np.isclose(
                parameters[index], lower, atol=hard_bound_atol, rtol=0.0
            ):
                projected[index] = min(projected[index], 0.0)
            elif np.isclose(
                parameters[index], upper, atol=hard_bound_atol, rtol=0.0
            ):
                projected[index] = max(projected[index], 0.0)
        return projected

    def reported_token_signature(
        mean: np.ndarray, mean_covariance: np.ndarray, between: np.ndarray
    ) -> str:
        """Return the exact display tokens derivable without Monte Carlo draws.

        The manuscript renders central sensitivity and specificity to one
        percentage-point decimal, the worked-profile central and marginal
        sensitivity/specificity intervals to four decimal places, and central
        20% information to four decimal places (three in cross-point summaries).
        Draw-derived information and branch intervals are audited downstream
        after the selected fits are regenerated and are intentionally not
        approximated here.
        """
        central = logistic(mean)
        pooled_low = logistic(
            mean - 1.959963984540054 * np.sqrt(np.diag(mean_covariance))
        )
        pooled_high = logistic(
            mean + 1.959963984540054 * np.sqrt(np.diag(mean_covariance))
        )
        predictive = mean_covariance + between
        predictive_low = logistic(
            mean - 1.959963984540054 * np.sqrt(np.diag(predictive))
        )
        predictive_high = logistic(
            mean + 1.959963984540054 * np.sqrt(np.diag(predictive))
        )
        information = diagnostic_metrics(central[0], central[1], 0.20)[
            "information_bits"
        ]
        return "|".join(
            [
                *(f"{100 * value:.1f}%" for value in central),
                *(f"{value:.4f}" for value in central),
                *(f"{value:.4f}" for value in pooled_low),
                *(f"{value:.4f}" for value in pooled_high),
                *(f"{value:.4f}" for value in predictive_low),
                *(f"{value:.4f}" for value in predictive_high),
                f"{float(information):.3f}",
                f"{float(information):.4f}",
            ]
        )

    start_records: list[dict[str, object]] = []
    valid_endpoints: list[dict[str, object]] = []
    for start_order, (start_name, start_parameters) in enumerate(starts):
        result = minimize(
            evaluate,
            start_parameters,
            method="L-BFGS-B",
            bounds=bounds,
            options={
                "maxiter": 2000,
                "ftol": 1e-12,
                "gtol": 1e-8,
                "maxls": 50,
            },
        )
        endpoint = np.asarray(result.x, dtype=float)
        raw_gradient = np.asarray(
            result.jac if getattr(result, "jac", None) is not None else [math.nan] * 3,
            dtype=float,
        )
        projected_gradient = projected_kkt_gradient(endpoint, raw_gradient)
        record: dict[str, object] = {
            "start_order": start_order,
            "start_name": start_name,
            "initial_log_l00": float(start_parameters[0]),
            "initial_l10": float(start_parameters[1]),
            "initial_log_l11": float(start_parameters[2]),
            "optimizer_success": bool(result.success),
            "optimizer_status": int(result.status),
            "optimizer_message": str(result.message),
            "optimizer_iterations": int(result.nit),
            "optimizer_reported_objective": float(result.fun),
            "endpoint_log_l00": float(endpoint[0]),
            "endpoint_l10": float(endpoint[1]),
            "endpoint_log_l11": float(endpoint[2]),
            "endpoint_log_l00_at_lower_bound": bool(
                np.isclose(endpoint[0], -8.0, atol=hard_bound_atol, rtol=0.0)
            ),
            "endpoint_log_l11_at_lower_bound": bool(
                np.isclose(endpoint[2], -8.0, atol=hard_bound_atol, rtol=0.0)
            ),
            "raw_gradient_log_l00": float(raw_gradient[0]),
            "raw_gradient_l10": float(raw_gradient[1]),
            "raw_gradient_log_l11": float(raw_gradient[2]),
            "raw_gradient_max_abs": float(np.max(np.abs(raw_gradient))),
            "projected_kkt_gradient_log_l00": float(projected_gradient[0]),
            "projected_kkt_gradient_l10": float(projected_gradient[1]),
            "projected_kkt_gradient_log_l11": float(projected_gradient[2]),
            "projected_kkt_gradient_max_abs": float(
                np.max(np.abs(projected_gradient))
            ),
        }
        finite_objective = False
        covariance_psd = False
        try:
            evaluated = evaluate(endpoint, full=True)
            if not isinstance(evaluated, tuple):
                raise ValueError("invalid endpoint objective")
            objective, endpoint_mean, endpoint_mean_covariance, endpoint_between = evaluated
            endpoint_predictive = endpoint_mean_covariance + endpoint_between
            finite_objective = bool(
                math.isfinite(float(objective))
                and np.isfinite(endpoint_mean).all()
                and np.isfinite(endpoint_mean_covariance).all()
                and np.isfinite(endpoint_between).all()
                and np.isfinite(endpoint_predictive).all()
            )
            covariance_psd = bool(
                finite_objective
                and all(
                    np.linalg.eigvalsh(matrix).min() >= -1e-9
                    for matrix in (
                        endpoint_mean_covariance,
                        endpoint_between,
                        endpoint_predictive,
                    )
                )
            )
            endpoint_sensitivity, endpoint_specificity = logistic(endpoint_mean)
            record.update(
                {
                    "recomputed_objective": float(objective),
                    "sensitivity": float(endpoint_sensitivity),
                    "specificity": float(endpoint_specificity),
                    "reported_token_signature": reported_token_signature(
                        endpoint_mean, endpoint_mean_covariance, endpoint_between
                    ),
                }
            )
        except (FloatingPointError, np.linalg.LinAlgError, ValueError, OverflowError):
            objective = math.inf
            endpoint_mean = np.asarray([math.nan, math.nan])
            endpoint_mean_covariance = np.full((2, 2), math.nan)
            endpoint_between = np.full((2, 2), math.nan)
            record.update(
                {
                    "recomputed_objective": math.inf,
                    "sensitivity": math.nan,
                    "specificity": math.nan,
                    "reported_token_signature": "",
                }
            )
        record["finite_recomputed_objective"] = finite_objective
        record["covariance_matrices_psd"] = covariance_psd
        record["valid_endpoint"] = bool(
            result.success and finite_objective and covariance_psd
        )
        record["selected"] = False
        record["numerical_tie_with_minimum"] = False
        start_records.append(record)
        if record["valid_endpoint"]:
            valid_endpoints.append(
                {
                    "record": record,
                    "result": result,
                    "objective": float(objective),
                    "mean": endpoint_mean,
                    "mean_covariance": endpoint_mean_covariance,
                    "between": endpoint_between,
                }
            )

    if not valid_endpoints:
        messages = "; ".join(
            f"{record['start_name']}: {record['optimizer_message']}"
            for record in start_records
        )
        raise RuntimeError(f"REML multistart optimization failed: {messages}")

    minimum = min(
        valid_endpoints,
        key=lambda endpoint: (
            endpoint["objective"],
            endpoint["record"]["start_order"],
        ),
    )
    minimum_objective = float(minimum["objective"])
    objective_tie_tolerance = 1e-8 * max(1.0, abs(minimum_objective))
    minimum_central = logistic(minimum["mean"])
    tie_candidates = []
    for endpoint in valid_endpoints:
        objective_delta = float(endpoint["objective"] - minimum_objective)
        central_delta = np.abs(logistic(endpoint["mean"]) - minimum_central)
        endpoint["record"]["objective_delta_from_minimum"] = objective_delta
        endpoint["record"]["maximum_central_probability_delta_from_minimum"] = float(
            np.max(central_delta)
        )
        tied = bool(
            objective_delta <= objective_tie_tolerance
            and np.max(central_delta) <= 1e-8
        )
        endpoint["record"]["numerical_tie_with_minimum"] = tied
        if tied:
            tie_candidates.append(endpoint)
    selected = min(
        tie_candidates,
        key=lambda endpoint: (
            endpoint["record"]["projected_kkt_gradient_max_abs"],
            endpoint["record"]["start_order"],
        ),
    )
    selected["record"]["selected"] = True
    selected_record = selected["record"]
    tied_log_l00_at_lower_bound = any(
        endpoint["record"]["numerical_tie_with_minimum"]
        and endpoint["record"]["endpoint_log_l00_at_lower_bound"]
        for endpoint in valid_endpoints
    )
    tied_log_l11_at_lower_bound = any(
        endpoint["record"]["numerical_tie_with_minimum"]
        and endpoint["record"]["endpoint_log_l11_at_lower_bound"]
        for endpoint in valid_endpoints
    )

    objectives = np.asarray([endpoint["objective"] for endpoint in valid_endpoints])
    sensitivities = np.asarray(
        [logistic(endpoint["mean"])[0] for endpoint in valid_endpoints]
    )
    specificities = np.asarray(
        [logistic(endpoint["mean"])[1] for endpoint in valid_endpoints]
    )
    sorted_objectives = np.sort(objectives)
    objective_gap = (
        float(sorted_objectives[1] - sorted_objectives[0])
        if len(sorted_objectives) > 1
        else math.nan
    )
    distinct_objective_deltas = sorted(
        float(endpoint["objective"] - minimum_objective)
        for endpoint in valid_endpoints
        if float(endpoint["objective"] - minimum_objective)
        > objective_tie_tolerance
    )
    objective_gap_to_next_distinct_basin = (
        distinct_objective_deltas[0] if distinct_objective_deltas else math.nan
    )
    objective_range = float(objectives.max() - objectives.min())
    sensitivity_range = float(sensitivities.max() - sensitivities.min())
    specificity_range = float(specificities.max() - specificities.min())
    alternative_rounded_claim_changed = len(
        {
            str(endpoint["record"]["reported_token_signature"])
            for endpoint in valid_endpoints
        }
    ) > 1
    empirical_endpoint = next(
        (
            endpoint
            for endpoint in valid_endpoints
            if endpoint["record"]["start_name"] == "empirical"
        ),
        None,
    )
    if empirical_endpoint is None:
        empirical_objective_delta = math.inf
        empirical_sensitivity_delta = math.inf
        empirical_specificity_delta = math.inf
        rounded_claim_changed = True
    else:
        empirical_objective_delta = float(
            empirical_endpoint["objective"] - selected["objective"]
        )
        empirical_central = logistic(empirical_endpoint["mean"])
        selected_central = logistic(selected["mean"])
        empirical_sensitivity_delta = float(
            abs(empirical_central[0] - selected_central[0])
        )
        empirical_specificity_delta = float(
            abs(empirical_central[1] - selected_central[1])
        )
        rounded_claim_changed = bool(
            empirical_endpoint["record"]["reported_token_signature"]
            != selected_record["reported_token_signature"]
        )
    material_objective_path = empirical_objective_delta > 1e-5
    material_central_path = (
        empirical_sensitivity_delta > 1e-4
        or empirical_specificity_delta > 1e-4
    )
    material_path_dependence = bool(
        material_objective_path or material_central_path or rounded_claim_changed
    )
    if material_path_dependence:
        path_dependence_class = "material_alternative_basin"
    elif empirical_endpoint is not None and (
        empirical_objective_delta > objective_tie_tolerance
        or empirical_sensitivity_delta > 1e-8
        or empirical_specificity_delta > 1e-8
        or np.max(
            np.abs(
                np.asarray(
                    [
                        empirical_endpoint["record"]["endpoint_log_l00"],
                        empirical_endpoint["record"]["endpoint_l10"],
                        empirical_endpoint["record"]["endpoint_log_l11"],
                    ]
                )
                - np.asarray(
                    [
                        selected_record["endpoint_log_l00"],
                        selected_record["endpoint_l10"],
                        selected_record["endpoint_log_l11"],
                    ]
                )
            )
        )
        > 1e-6
    ):
        path_dependence_class = "tiny_boundary_refinement"
    else:
        path_dependence_class = "numerically_equivalent"

    result = selected["result"]
    objective = float(selected["objective"])
    mean = selected["mean"]
    mean_covariance = selected["mean_covariance"]
    between = selected["between"]
    predictive_covariance = mean_covariance + between
    return {
        "mean": mean,
        "mean_covariance": mean_covariance,
        "between_covariance": between,
        "predictive_covariance": predictive_covariance,
        "objective": float(objective),
        "iterations": int(result.nit),
        "max_abs_gradient": float(selected_record["raw_gradient_max_abs"]),
        "projected_kkt_gradient_log_l00": float(
            selected_record["projected_kkt_gradient_log_l00"]
        ),
        "projected_kkt_gradient_l10": float(
            selected_record["projected_kkt_gradient_l10"]
        ),
        "projected_kkt_gradient_log_l11": float(
            selected_record["projected_kkt_gradient_log_l11"]
        ),
        "projected_kkt_gradient_max_abs": float(
            selected_record["projected_kkt_gradient_max_abs"]
        ),
        "optimizer_message": str(result.message),
        "optimizer_parameter_log_l00": float(result.x[0]),
        "optimizer_parameter_l10": float(result.x[1]),
        "optimizer_parameter_log_l11": float(result.x[2]),
        "optimizer_selected_log_l00_at_lower_bound": bool(
            selected_record["endpoint_log_l00_at_lower_bound"]
        ),
        "optimizer_selected_log_l11_at_lower_bound": bool(
            selected_record["endpoint_log_l11_at_lower_bound"]
        ),
        "optimizer_tied_solution_log_l00_at_lower_bound": (
            tied_log_l00_at_lower_bound
        ),
        "optimizer_tied_solution_log_l11_at_lower_bound": (
            tied_log_l11_at_lower_bound
        ),
        "optimizer_hard_bound_atol": hard_bound_atol,
        "optimizer_multistart_count": len(starts),
        "optimizer_valid_start_count": len(valid_endpoints),
        "optimizer_selected_start": str(selected_record["start_name"]),
        "optimizer_selected_start_order": int(selected_record["start_order"]),
        "optimizer_minimum_recomputed_objective": minimum_objective,
        "optimizer_selected_objective_delta_from_minimum": float(
            objective - minimum_objective
        ),
        "optimizer_objective_tie_tolerance": objective_tie_tolerance,
        "optimizer_objective_gap_to_second_best": objective_gap,
        "optimizer_objective_gap_to_next_distinct_basin": (
            objective_gap_to_next_distinct_basin
        ),
        "optimizer_valid_objective_range": objective_range,
        "optimizer_valid_sensitivity_range": sensitivity_range,
        "optimizer_valid_specificity_range": specificity_range,
        "optimizer_alternative_rounded_claim_changed_across_starts": (
            alternative_rounded_claim_changed
        ),
        "optimizer_empirical_start_valid": empirical_endpoint is not None,
        "optimizer_empirical_to_selected_objective_delta": (
            empirical_objective_delta
        ),
        "optimizer_empirical_to_selected_sensitivity_delta": (
            empirical_sensitivity_delta
        ),
        "optimizer_empirical_to_selected_specificity_delta": (
            empirical_specificity_delta
        ),
        "optimizer_rounded_claim_changed_from_empirical": rounded_claim_changed,
        "optimizer_material_objective_path_dependence": material_objective_path,
        "optimizer_material_central_path_dependence": material_central_path,
        "optimizer_material_path_dependence": material_path_dependence,
        "optimizer_path_dependence_class": path_dependence_class,
        "optimizer_start_results": start_records,
        "continuity_correction_rule": continuity_correction,
    }


def _interval(values) -> tuple[float, float, float]:
    low, median, high = np.quantile(values, [0.025, 0.5, 0.975])
    return float(low), float(median), float(high)


def _add_interval(row: dict, stem: str, values) -> None:
    low, median, high = _interval(values)
    row[f"{stem}_low"] = low
    row[f"{stem}_median"] = median
    row[f"{stem}_high"] = high


def analyze_group(
    group: dict,
    draws: int,
    seed: int,
    *,
    continuity_correction: str = "universal",
    multistart_records: list[dict[str, object]] | None = None,
) -> tuple[dict, list[dict]]:
    fit = fit_bivariate_reml(
        group["rows"], continuity_correction=continuity_correction
    )
    if multistart_records is not None:
        for start_record in fit["optimizer_start_results"]:
            multistart_records.append(
                {
                    "source": group["source"],
                    "review_id": group["review_id"],
                    "group_id": group["group_id"],
                    "test_name": group["test_name"],
                    "continuity_correction_rule": continuity_correction,
                    **start_record,
                }
            )
    mean = fit["mean"]
    sensitivity, specificity = logistic(mean)
    point_metrics = diagnostic_metrics(sensitivity, specificity, 0.20)
    rng = np.random.default_rng(seed)
    pooled_logits = rng.multivariate_normal(mean, fit["mean_covariance"], size=draws)
    predictive_logits = rng.multivariate_normal(mean, fit["predictive_covariance"], size=draws)
    pooled_se, pooled_sp = logistic(pooled_logits[:, 0]), logistic(pooled_logits[:, 1])
    predictive_se, predictive_sp = logistic(predictive_logits[:, 0]), logistic(predictive_logits[:, 1])
    pooled_metrics = diagnostic_metrics(pooled_se, pooled_sp, 0.20)
    predictive_metrics = diagnostic_metrics(predictive_se, predictive_sp, 0.20)
    study_positive_fractions = np.asarray(
        [
            (row["tp"] + row["fn"]) / sum(row[name] for name in ("tp", "fp", "fn", "tn"))
            for row in group["rows"]
        ],
        dtype=float,
    )
    published_se = float(group["published_sensitivity"]) if group["published_sensitivity"] != "" else math.nan
    published_sp = float(group["published_specificity"]) if group["published_specificity"] != "" else math.nan
    between = fit["between_covariance"]
    mean_covariance = fit["mean_covariance"]
    row = {
        key: value for key, value in group.items() if key != "rows"
    }
    row.update(
        {
            "model": "continuity-corrected bivariate REML",
            "model_converged": True,
            "sensitivity": float(sensitivity),
            "specificity": float(specificity),
            "lr_positive": float(point_metrics["lr_positive"]),
            "lr_negative": float(point_metrics["lr_negative"]),
            "information_bits_at_20_percent": float(point_metrics["information_bits"]),
            "diagnostic_entropy_reduction_at_20_percent": float(
                point_metrics["diagnostic_entropy_reduction"]
            ),
            "negative_posterior_at_20_percent": float(point_metrics["negative_posterior"]),
            "published_sensitivity_difference": float(sensitivity - published_se) if math.isfinite(published_se) else "",
            "published_specificity_difference": float(specificity - published_sp) if math.isfinite(published_sp) else "",
            "mean_logit_sensitivity_variance": float(mean_covariance[0, 0]),
            "mean_logit_specificity_variance": float(mean_covariance[1, 1]),
            "mean_logit_covariance": float(mean_covariance[0, 1]),
            "between_logit_sensitivity_variance": float(between[0, 0]),
            "between_logit_specificity_variance": float(between[1, 1]),
            "between_logit_covariance": float(between[0, 1]),
            "between_logit_correlation": float(
                between[0, 1] / math.sqrt(between[0, 0] * between[1, 1])
            ),
            "reml_objective": fit["objective"],
            "optimizer_iterations": fit["iterations"],
            "optimizer_max_abs_gradient": fit["max_abs_gradient"],
            "optimizer_projected_kkt_gradient_log_l00": fit[
                "projected_kkt_gradient_log_l00"
            ],
            "optimizer_projected_kkt_gradient_l10": fit[
                "projected_kkt_gradient_l10"
            ],
            "optimizer_projected_kkt_gradient_log_l11": fit[
                "projected_kkt_gradient_log_l11"
            ],
            "optimizer_projected_kkt_gradient_max_abs": fit[
                "projected_kkt_gradient_max_abs"
            ],
            "optimizer_parameter_log_l00": fit["optimizer_parameter_log_l00"],
            "optimizer_parameter_l10": fit["optimizer_parameter_l10"],
            "optimizer_parameter_log_l11": fit["optimizer_parameter_log_l11"],
            "optimizer_tied_solution_log_l00_at_lower_bound": fit[
                "optimizer_tied_solution_log_l00_at_lower_bound"
            ],
            "optimizer_tied_solution_log_l11_at_lower_bound": fit[
                "optimizer_tied_solution_log_l11_at_lower_bound"
            ],
            "optimizer_selected_log_l00_at_lower_bound": fit[
                "optimizer_selected_log_l00_at_lower_bound"
            ],
            "optimizer_selected_log_l11_at_lower_bound": fit[
                "optimizer_selected_log_l11_at_lower_bound"
            ],
            "optimizer_hard_bound_atol": fit["optimizer_hard_bound_atol"],
            "optimizer_multistart_count": fit["optimizer_multistart_count"],
            "optimizer_valid_start_count": fit["optimizer_valid_start_count"],
            "optimizer_selected_start": fit["optimizer_selected_start"],
            "optimizer_selected_start_order": fit[
                "optimizer_selected_start_order"
            ],
            "optimizer_minimum_recomputed_objective": fit[
                "optimizer_minimum_recomputed_objective"
            ],
            "optimizer_selected_objective_delta_from_minimum": fit[
                "optimizer_selected_objective_delta_from_minimum"
            ],
            "optimizer_objective_tie_tolerance": fit[
                "optimizer_objective_tie_tolerance"
            ],
            "optimizer_objective_gap_to_second_best": fit[
                "optimizer_objective_gap_to_second_best"
            ],
            "optimizer_objective_gap_to_next_distinct_basin": fit[
                "optimizer_objective_gap_to_next_distinct_basin"
            ],
            "optimizer_valid_objective_range": fit[
                "optimizer_valid_objective_range"
            ],
            "optimizer_valid_sensitivity_range": fit[
                "optimizer_valid_sensitivity_range"
            ],
            "optimizer_valid_specificity_range": fit[
                "optimizer_valid_specificity_range"
            ],
            "optimizer_alternative_rounded_claim_changed_across_starts": fit[
                "optimizer_alternative_rounded_claim_changed_across_starts"
            ],
            "optimizer_empirical_start_valid": fit[
                "optimizer_empirical_start_valid"
            ],
            "optimizer_empirical_to_selected_objective_delta": fit[
                "optimizer_empirical_to_selected_objective_delta"
            ],
            "optimizer_empirical_to_selected_sensitivity_delta": fit[
                "optimizer_empirical_to_selected_sensitivity_delta"
            ],
            "optimizer_empirical_to_selected_specificity_delta": fit[
                "optimizer_empirical_to_selected_specificity_delta"
            ],
            "optimizer_rounded_claim_changed_from_empirical": fit[
                "optimizer_rounded_claim_changed_from_empirical"
            ],
            "optimizer_material_objective_path_dependence": fit[
                "optimizer_material_objective_path_dependence"
            ],
            "optimizer_material_central_path_dependence": fit[
                "optimizer_material_central_path_dependence"
            ],
            "optimizer_material_path_dependence": fit[
                "optimizer_material_path_dependence"
            ],
            "optimizer_path_dependence_class": fit[
                "optimizer_path_dependence_class"
            ],
            "continuity_correction_rule": fit["continuity_correction_rule"],
            "study_positive_fraction_low": float(np.min(study_positive_fractions)),
            "study_positive_fraction_median": float(np.median(study_positive_fractions)),
            "study_positive_fraction_high": float(np.max(study_positive_fractions)),
            "zero_cell_study_fraction": float(
                np.mean(
                    [
                        any(row[name] == 0 for name in ("tp", "fp", "fn", "tn"))
                        for row in group["rows"]
                    ]
                )
            ),
        }
    )
    for stem, values in (
        ("pooled_mean_sensitivity", pooled_se),
        ("pooled_mean_specificity", pooled_sp),
        ("predictive_sensitivity", predictive_se),
        ("predictive_specificity", predictive_sp),
        ("pooled_mean_der_at_20_percent", pooled_metrics["diagnostic_entropy_reduction"]),
        ("predictive_der_at_20_percent", predictive_metrics["diagnostic_entropy_reduction"]),
        ("pooled_mean_information_bits_at_20_percent", pooled_metrics["information_bits"]),
        ("predictive_information_bits_at_20_percent", predictive_metrics["information_bits"]),
        (
            "pooled_mean_positive_information_bits_at_20_percent",
            pooled_metrics["positive_result_information_bits"],
        ),
        (
            "predictive_positive_information_bits_at_20_percent",
            predictive_metrics["positive_result_information_bits"],
        ),
        (
            "pooled_mean_negative_information_bits_at_20_percent",
            pooled_metrics["negative_result_information_bits"],
        ),
        (
            "predictive_negative_information_bits_at_20_percent",
            predictive_metrics["negative_result_information_bits"],
        ),
        ("pooled_mean_negative_at_20_percent", pooled_metrics["negative_posterior"]),
        ("predictive_negative_at_20_percent", predictive_metrics["negative_posterior"]),
    ):
        _add_interval(row, stem, values)

    boundaries = []
    for threshold in THRESHOLDS:
        boundary = {
            "review_id": group["review_id"],
            "review_title": group["review_title"],
            "group_id": group["group_id"],
            "test_name": group["test_name"],
            "analysis_scope": group["analysis_scope"],
            "primary_atlas": group["primary_atlas"],
            "residual_risk_threshold": threshold,
            "maximum_pretest_probability": float(
                maximum_pretest_probability(sensitivity, specificity, threshold)
            ),
        }
        _add_interval(
            boundary,
            "pooled_mean_maximum_pretest_probability",
            maximum_pretest_probability(pooled_se, pooled_sp, threshold),
        )
        _add_interval(
            boundary,
            "predictive_maximum_pretest_probability",
            maximum_pretest_probability(predictive_se, predictive_sp, threshold),
        )
        boundaries.append(boundary)
    return row, boundaries


def operating_point_curves(operating_points: list[dict]) -> list[dict]:
    rows = []
    for point in operating_points:
        if not point["primary_atlas"]:
            continue
        for prevalence in PREVALENCES:
            metrics = diagnostic_metrics(point["sensitivity"], point["specificity"], prevalence)
            rows.append(
                {
                    "review_id": point["review_id"],
                    "group_id": point["group_id"],
                    "test_name": point["test_name"],
                    "analysis_scope": point["analysis_scope"],
                    "pretest_probability": prevalence,
                    "positive_posterior": float(metrics["positive_posterior"]),
                    "negative_posterior": float(metrics["negative_posterior"]),
                    "information_bits": float(metrics["information_bits"]),
                    "diagnostic_entropy_reduction": float(metrics["diagnostic_entropy_reduction"]),
                }
            )
    return rows


def rank_reversals(operating_points: list[dict]) -> list[dict]:
    """Return information versus rule-out reversals within the same review."""
    by_review: dict[str, list[dict]] = defaultdict(list)
    for point in operating_points:
        if point.get("primary_atlas", point.get("analysis_scope") == "main"):
            by_review[str(point["review_id"])].append(point)
    rows = []
    for review_id, points in by_review.items():
        for left, right in combinations(points, 2):
            if left["diagnostic_entropy_reduction_at_20_percent"] == right["diagnostic_entropy_reduction_at_20_percent"]:
                continue
            more_information, less_information = sorted(
                (left, right),
                key=lambda point: point["diagnostic_entropy_reduction_at_20_percent"],
                reverse=True,
            )
            if more_information["negative_posterior_at_20_percent"] <= less_information["negative_posterior_at_20_percent"]:
                continue
            rows.append(
                {
                    "review_id": review_id,
                    "review_title": more_information.get("review_title", ""),
                    "more_information_test": more_information["test_name"],
                    "more_information_der": more_information["diagnostic_entropy_reduction_at_20_percent"],
                    "more_information_negative_posterior": more_information["negative_posterior_at_20_percent"],
                    "better_ruleout_test": less_information["test_name"],
                    "better_ruleout_der": less_information["diagnostic_entropy_reduction_at_20_percent"],
                    "better_ruleout_negative_posterior": less_information["negative_posterior_at_20_percent"],
                }
            )
    return rows


def rank_reversal_sensitivity(curves: list[dict]) -> list[dict]:
    """Count within-review rank reversals at each standardized prior."""
    by_prior: dict[float, dict[str, list[dict]]] = defaultdict(lambda: defaultdict(list))
    for row in curves:
        by_prior[float(row["pretest_probability"])][str(row["review_id"])].append(row)
    output = []
    for prevalence, reviews in sorted(by_prior.items()):
        pairs = reversals = 0
        for points in reviews.values():
            for left, right in combinations(points, 2):
                pairs += 1
                der_difference = float(left["diagnostic_entropy_reduction"]) - float(right["diagnostic_entropy_reduction"])
                risk_difference = float(left["negative_posterior"]) - float(right["negative_posterior"])
                reversals += der_difference * risk_difference > 0
        output.append(
            {
                "pretest_probability": prevalence,
                "rank_reversals": reversals,
                "comparable_primary_pairs": pairs,
                "fraction": reversals / pairs if pairs else 0.0,
            }
        )
    return output


def dominance_concordance(operating_points: list[dict]) -> list[dict]:
    """Classify every within-review primary pair by coordinate-wise dominance."""
    by_review: dict[str, list[dict]] = defaultdict(list)
    for point in operating_points:
        if point["primary_atlas"]:
            by_review[str(point["review_id"])].append(point)
    rows = []
    for review_id, points in by_review.items():
        for left, right in combinations(points, 2):
            left_dominates = (
                left["sensitivity"] >= right["sensitivity"]
                and left["specificity"] >= right["specificity"]
                and (left["sensitivity"] > right["sensitivity"] or left["specificity"] > right["specificity"])
            )
            right_dominates = (
                right["sensitivity"] >= left["sensitivity"]
                and right["specificity"] >= left["specificity"]
                and (right["sensitivity"] > left["sensitivity"] or right["specificity"] > left["specificity"])
            )
            dominant = left if left_dominates else right if right_dominates else None
            der_difference = (
                left["diagnostic_entropy_reduction_at_20_percent"]
                - right["diagnostic_entropy_reduction_at_20_percent"]
            )
            risk_difference = (
                left["negative_posterior_at_20_percent"]
                - right["negative_posterior_at_20_percent"]
            )
            higher_der = left if der_difference > 0 else right
            better_ruleout = left if risk_difference < 0 else right
            rows.append(
                {
                    "review_id": review_id,
                    "review_title": left.get("review_title", ""),
                    "left_group_id": left.get("group_id", left["test_name"]),
                    "right_group_id": right.get("group_id", right["test_name"]),
                    "left_test": left["test_name"],
                    "right_test": right["test_name"],
                    "left_sensitivity": left["sensitivity"],
                    "left_specificity": left["specificity"],
                    "right_sensitivity": right["sensitivity"],
                    "right_specificity": right["specificity"],
                    "both_oriented_at_least_as_well_as_chance": (
                        left["sensitivity"] + left["specificity"] >= 1.0
                        and right["sensitivity"] + right["specificity"] >= 1.0
                    ),
                    "dominance_relation": dominant["test_name"] if dominant else "non-dominated",
                    "higher_der_test": higher_der["test_name"],
                    "better_ruleout_test": better_ruleout["test_name"],
                    "rank_reversal": der_difference * risk_difference > 0,
                    "dominant_test_has_higher_der": dominant is None or dominant is higher_der,
                    "dominant_test_has_lower_post_negative_risk": dominant is None or dominant is better_ruleout,
                }
            )
    return rows


def _der_difference(left: dict, right: dict, prior: float) -> float:
    left_der = diagnostic_metrics(left["sensitivity"], left["specificity"], prior)[
        "diagnostic_entropy_reduction"
    ]
    right_der = diagnostic_metrics(right["sensitivity"], right["specificity"], prior)[
        "diagnostic_entropy_reduction"
    ]
    return float(left_der - right_der)


def prior_crossings(operating_points: list[dict], dominance: list[dict]) -> list[dict]:
    """Locate DER rank crossings for every non-dominated within-review pair."""
    points = {str(point["group_id"]): point for point in operating_points}
    grid = np.asarray(PRIOR_CROSSING_GRID, dtype=float)
    rows = []
    for pair in dominance:
        if pair["dominance_relation"] != "non-dominated":
            continue
        left = points[str(pair["left_group_id"])]
        right = points[str(pair["right_group_id"])]
        differences = np.asarray([_der_difference(left, right, prior) for prior in grid])
        roots: list[float] = []
        for index in range(len(grid) - 1):
            low, high = grid[index], grid[index + 1]
            low_value, high_value = differences[index], differences[index + 1]
            if abs(low_value) < 1e-10:
                roots.append(float(low))
            elif low_value * high_value < 0:
                roots.append(float(brentq(lambda prior: _der_difference(left, right, prior), low, high)))
        if abs(differences[-1]) < 1e-10:
            roots.append(float(grid[-1]))
        roots = sorted(
            root
            for index, root in enumerate(sorted(roots))
            if index == 0 or abs(root - sorted(roots)[index - 1]) > 1e-5
        )
        composition_low = min(
            float(left["study_positive_fraction_low"]),
            float(right["study_positive_fraction_low"]),
        )
        composition_high = max(
            float(left["study_positive_fraction_high"]),
            float(right["study_positive_fraction_high"]),
        )

        def rank_at(prior: float) -> str:
            difference = _der_difference(left, right, prior)
            if abs(difference) < 1e-10:
                return "tie"
            return str(left["group_id"] if difference > 0 else right["group_id"])

        left_low = float(left["pooled_mean_der_at_20_percent_low"])
        left_high = float(left["pooled_mean_der_at_20_percent_high"])
        right_low = float(right["pooled_mean_der_at_20_percent_low"])
        right_high = float(right["pooled_mean_der_at_20_percent_high"])
        stable_order = (
            str(left["group_id"])
            if left_low > right_high
            else str(right["group_id"])
            if right_low > left_high
            else "intervals_overlap"
        )
        rows.append(
            {
                "review_id": pair["review_id"],
                "review_title": pair["review_title"],
                "left_group_id": left["group_id"],
                "left_test": left["test_name"],
                "right_group_id": right["group_id"],
                "right_test": right["test_name"],
                "crossing_count": len(roots),
                "crossing_priors": ";".join(f"{root:.6f}" for root in roots),
                "crossing_in_1_to_50_percent": any(0.01 <= root <= 0.50 for root in roots),
                "study_composition_low": composition_low,
                "study_composition_high": composition_high,
                "crossing_in_study_composition_range": any(
                    composition_low <= root <= composition_high for root in roots
                ),
                "higher_der_at_5_percent": rank_at(0.05),
                "higher_der_at_20_percent": rank_at(0.20),
                "higher_der_at_50_percent": rank_at(0.50),
                "pooled_mean_interval_order_at_20_percent": stable_order,
            }
        )
    return rows


def j_matched_contrasts(operating_points: list[dict], tolerance: float = 0.02) -> list[dict]:
    """Describe same-review pairs with nearly equal Youden J."""
    by_review: dict[str, list[dict]] = defaultdict(list)
    for point in operating_points:
        if point["primary_atlas"]:
            by_review[str(point["review_id"])].append(point)
    rows = []
    for review_id, points in by_review.items():
        for left, right in combinations(points, 2):
            left_j = float(left["sensitivity"] + left["specificity"] - 1.0)
            right_j = float(right["sensitivity"] + right["specificity"] - 1.0)
            if abs(left_j - right_j) > tolerance:
                continue
            left_risk = float(left["negative_posterior_at_20_percent"])
            right_risk = float(right["negative_posterior_at_20_percent"])
            rows.append(
                {
                    "review_id": review_id,
                    "review_title": left.get("review_title", ""),
                    "left_group_id": left["group_id"],
                    "left_test": left["test_name"],
                    "right_group_id": right["group_id"],
                    "right_test": right["test_name"],
                    "absolute_youden_difference": abs(left_j - right_j),
                    "absolute_der_difference_at_20_percent": abs(
                        float(left["diagnostic_entropy_reduction_at_20_percent"])
                        - float(right["diagnostic_entropy_reduction_at_20_percent"])
                    ),
                    "post_negative_risk_ratio": max(left_risk, right_risk) / min(left_risk, right_risk),
                }
            )
    return rows


def observed_composition_correlation(operating_points: list[dict]) -> dict:
    primary = [point for point in operating_points if point["primary_atlas"]]
    der = [
        float(
            diagnostic_metrics(
                point["sensitivity"],
                point["specificity"],
                point["study_positive_fraction_median"],
            )["diagnostic_entropy_reduction"]
        )
        for point in primary
    ]
    youden = [float(point["sensitivity"] + point["specificity"] - 1.0) for point in primary]
    correlation = spearmanr(der, youden)
    return {
        "primary_operating_points": len(primary),
        "prior_source": "median study target-positive fraction; not clinical prevalence",
        "spearman_rho": float(correlation.statistic),
        "two_sided_p_value": float(correlation.pvalue),
    }


def orientation_audit(groups: list[dict]) -> list[dict]:
    rows = []
    for group in groups:
        if group["eligibility"] != "below_chance_orientation":
            continue
        tp = sum(row["tp"] for row in group["rows"])
        fp = sum(row["fp"] for row in group["rows"])
        fn = sum(row["fn"] for row in group["rows"])
        tn = sum(row["tn"] for row in group["rows"])
        sensitivity = tp / (tp + fn)
        specificity = tn / (tn + fp)
        pmc = group["source"] == "PMC Open Access"
        rows.append(
            {
                "source": group["source"],
                "review_id": group["review_id"],
                "review_title": group["review_title"],
                "group_id": group["group_id"],
                "test_name": group["test_name"],
                "source_url": group.get("source_url", ""),
                "studies": len(group["rows"]),
                "aggregate_tp": tp,
                "aggregate_fp": fp,
                "aggregate_fn": fn,
                "aggregate_tn": tn,
                "aggregate_sensitivity": sensitivity,
                "aggregate_specificity": specificity,
                "aggregate_youden_j": sensitivity + specificity - 1.0,
                "adjudication": (
                    "Excluded without flipping. Normalized cells reproduce the explicit source table; "
                    "the source-oriented aggregate remains below chance."
                    if pmc
                    else "Excluded without flipping. The source-defined test direction yields aggregate "
                    "Youden J below zero; no parser transformation was applied."
                ),
                "parser_error_found": False,
            }
        )
    return rows


def fit_bivariate_binomial_glmm(rows: list[dict], initial_fit: dict | None = None, nodes: int = 7) -> dict:
    """Fit a two-outcome binomial GLMM with a Laplace marginal likelihood."""
    _ = nodes  # Retained for API compatibility with the earlier quadrature prototype.
    observations = [
        (int(row["tp"]), int(row["tp"] + row["fn"]), int(row["tn"]), int(row["tn"] + row["fp"]))
        for row in rows
    ]
    if initial_fit is None:
        initial_fit = fit_bivariate_reml(rows)
    mean = np.asarray(initial_fit["mean"], dtype=float)
    covariance = np.asarray(initial_fit["between_covariance"], dtype=float)
    standard_deviations = np.sqrt(np.maximum(np.diag(covariance), 0.02))
    rho = float(
        np.clip(covariance[0, 1] / (standard_deviations[0] * standard_deviations[1]), -0.95, 0.95)
    )
    initial = np.asarray(
        [mean[0], mean[1], math.log(standard_deviations[0]), math.log(standard_deviations[1]), np.arctanh(rho)]
    )

    def objective(parameters: np.ndarray) -> float:
        mu_se, mu_sp, log_sd_se, log_sd_sp, rho_parameter = parameters
        sd_se, sd_sp = math.exp(log_sd_se), math.exp(log_sd_sp)
        correlation = math.tanh(rho_parameter)
        covariance = np.asarray(
            [
                [sd_se**2, correlation * sd_se * sd_sp],
                [correlation * sd_se * sd_sp, sd_sp**2],
            ]
        )
        try:
            inverse = np.linalg.inv(covariance)
        except np.linalg.LinAlgError:
            return 1e100
        sign, log_determinant = np.linalg.slogdet(covariance)
        if sign <= 0:
            return 1e100
        mean = np.asarray([mu_se, mu_sp])
        log_likelihood = 0.0
        for tp, diseased, tn, non_diseased in observations:
            successes = np.asarray([tp, tn], dtype=float)
            trials = np.asarray([diseased, non_diseased], dtype=float)
            random_effect = np.zeros(2)
            for _ in range(40):
                logits = mean + random_effect
                probabilities = expit(logits)
                gradient = inverse @ random_effect + trials * probabilities - successes
                hessian = inverse + np.diag(trials * probabilities * (1.0 - probabilities))
                step = np.linalg.solve(hessian, gradient)
                random_effect -= step
                if np.max(np.abs(step)) < 1e-9:
                    break
            logits = mean + random_effect
            conditional = np.sum(
                gammaln(trials + 1.0)
                - gammaln(successes + 1.0)
                - gammaln(trials - successes + 1.0)
                + successes * -np.logaddexp(0.0, -logits)
                + (trials - successes) * -np.logaddexp(0.0, logits)
            )
            probabilities = expit(logits)
            hessian = inverse + np.diag(trials * probabilities * (1.0 - probabilities))
            hessian_sign, hessian_log_determinant = np.linalg.slogdet(hessian)
            if hessian_sign <= 0:
                return 1e100
            log_likelihood += float(
                conditional
                - 0.5 * log_determinant
                - 0.5 * random_effect @ inverse @ random_effect
                - 0.5 * hessian_log_determinant
            )
        return -log_likelihood

    result = minimize(
        objective,
        initial,
        method="L-BFGS-B",
        bounds=[(-8.0, 8.0), (-8.0, 8.0), (-4.0, 2.0), (-4.0, 2.0), (-2.5, 2.5)],
        options={"maxiter": 500, "ftol": 1e-10},
    )
    if not result.success:
        result = minimize(
            objective,
            result.x if np.isfinite(result.fun) else initial,
            method="Powell",
            bounds=[(-8.0, 8.0), (-8.0, 8.0), (-4.0, 2.0), (-4.0, 2.0), (-2.5, 2.5)],
            options={"maxiter": 800, "ftol": 1e-8},
        )
    if not result.success or not np.isfinite(result.fun):
        raise RuntimeError(f"Binomial GLMM optimization failed: {result.message}")
    mu_se, mu_sp, log_sd_se, log_sd_sp, rho_parameter = result.x
    sd_se, sd_sp = math.exp(log_sd_se), math.exp(log_sd_sp)
    correlation = math.tanh(rho_parameter)
    covariance = np.asarray(
        [[sd_se**2, correlation * sd_se * sd_sp], [correlation * sd_se * sd_sp, sd_sp**2]]
    )
    return {
        "mean": np.asarray([mu_se, mu_sp]),
        "between_covariance": covariance,
        "objective": float(result.fun),
        "iterations": int(result.nit),
        "max_abs_gradient": float(np.max(np.abs(result.jac))) if getattr(result, "jac", None) is not None else "",
    }


def glmm_sensitivity_panel(
    groups: list[dict], operating_points: list[dict], limit: int = 20, draws: int = DEFAULT_DRAWS
) -> list[dict]:
    """Compare REML and binomial GLMM estimates in a locked stratified panel."""
    points = {str(point["group_id"]): point for point in operating_points if point["primary_atlas"]}
    candidates = [group for group in groups if str(group["group_id"]) in points]
    quotas = {
        "Cochrane DTA Reference Dataset": 4,
        "Modern Cochrane data package": 3,
        "OSF": 3,
        "PMC Open Access": 6,
        "Zenodo": 4,
    }
    selected: list[dict] = []
    for source, quota in quotas.items():
        source_groups = sorted(
            (group for group in candidates if group["source"] == source),
            key=lambda group: (
                len(group["rows"]),
                sum(
                    any(row[name] == 0 for name in ("tp", "fp", "fn", "tn"))
                    for row in group["rows"]
                ),
                points[str(group["group_id"])]["between_logit_sensitivity_variance"]
                + points[str(group["group_id"])]["between_logit_specificity_variance"],
                str(group["group_id"]),
            ),
        )
        if not source_groups:
            continue
        indices = sorted(set(int(round(index)) for index in np.linspace(0, len(source_groups) - 1, min(quota, len(source_groups)))))
        selected.extend(source_groups[index] for index in indices)
    if len(selected) < limit:
        selected_ids = {str(group["group_id"]) for group in selected}
        selected.extend(
            group
            for group in sorted(candidates, key=lambda group: str(group["group_id"]))
            if str(group["group_id"]) not in selected_ids
        )
    selected = selected[:limit]
    output = []
    for index, group in enumerate(selected):
        point = points[str(group["group_id"])]
        initial_fit = {
            "mean": np.asarray(
                [
                    math.log(point["sensitivity"] / (1.0 - point["sensitivity"])),
                    math.log(point["specificity"] / (1.0 - point["specificity"])),
                ]
            ),
            "between_covariance": np.asarray(
                [
                    [point["between_logit_sensitivity_variance"], point["between_logit_covariance"]],
                    [point["between_logit_covariance"], point["between_logit_specificity_variance"]],
                ]
            ),
        }
        try:
            fit = fit_bivariate_binomial_glmm(group["rows"], initial_fit=initial_fit)
        except (RuntimeError, ValueError, np.linalg.LinAlgError) as error:
            output.append(
                {
                    "panel_order": index + 1,
                    "source": group["source"],
                    "review_id": group["review_id"],
                    "group_id": group["group_id"],
                    "test_name": group["test_name"],
                    "studies": len(group["rows"]),
                    "glmm_converged": False,
                    "failure_reason": str(error),
                }
            )
            continue
        glmm_sensitivity, glmm_specificity = expit(fit["mean"])
        glmm_metrics = diagnostic_metrics(glmm_sensitivity, glmm_specificity, 0.20)
        rng = np.random.default_rng(SEED + 10_000 + index)
        predictive_logits = rng.multivariate_normal(fit["mean"], fit["between_covariance"], size=draws)
        predictive_metrics = diagnostic_metrics(expit(predictive_logits[:, 0]), expit(predictive_logits[:, 1]), 0.20)
        glmm_der_low, _, glmm_der_high = _interval(predictive_metrics["diagnostic_entropy_reduction"])
        output.append(
            {
                "panel_order": index + 1,
                "source": group["source"],
                "review_id": group["review_id"],
                "group_id": group["group_id"],
                "test_name": group["test_name"],
                "studies": len(group["rows"]),
                "zero_cell_study_fraction": point["zero_cell_study_fraction"],
                "selection_heterogeneity_trace": point["between_logit_sensitivity_variance"]
                + point["between_logit_specificity_variance"],
                "glmm_converged": True,
                "reml_sensitivity": point["sensitivity"],
                "glmm_sensitivity": float(glmm_sensitivity),
                "sensitivity_difference": float(glmm_sensitivity - point["sensitivity"]),
                "reml_specificity": point["specificity"],
                "glmm_specificity": float(glmm_specificity),
                "specificity_difference": float(glmm_specificity - point["specificity"]),
                "reml_der_at_20_percent": point["diagnostic_entropy_reduction_at_20_percent"],
                "glmm_der_at_20_percent": float(glmm_metrics["diagnostic_entropy_reduction"]),
                "der_difference": float(
                    glmm_metrics["diagnostic_entropy_reduction"]
                    - point["diagnostic_entropy_reduction_at_20_percent"]
                ),
                "reml_predictive_der_width": float(
                    point["predictive_der_at_20_percent_high"]
                    - point["predictive_der_at_20_percent_low"]
                ),
                "glmm_predictive_der_low": glmm_der_low,
                "glmm_predictive_der_high": glmm_der_high,
                "glmm_predictive_der_width": glmm_der_high - glmm_der_low,
                "failure_reason": "",
            }
        )
    return output


def metric_correlations(operating_points: list[dict]) -> list[dict]:
    primary = [point for point in operating_points if point["primary_atlas"]]
    output = []
    youden = [point["sensitivity"] + point["specificity"] - 1.0 for point in primary]
    for prior in (0.05, 0.20, 0.50):
        der = [
            float(diagnostic_metrics(point["sensitivity"], point["specificity"], prior)["diagnostic_entropy_reduction"])
            for point in primary
        ]
        correlation = spearmanr(der, youden)
        output.append(
            {
                "pretest_probability": prior,
                "primary_operating_points": len(primary),
                "metric_1": "diagnostic_entropy_reduction",
                "metric_2": "youden_j",
                "spearman_rho": float(correlation.statistic),
                "two_sided_p_value": float(correlation.pvalue),
            }
        )
    return output


def source_stability(operating_points: list[dict]) -> list[dict]:
    """Summarize the primary distribution before and after PMC expansion."""
    cohorts = (
        ("pre_pmc", [point for point in operating_points if point["source"] != "PMC Open Access"]),
        ("pmc_open_access", [point for point in operating_points if point["source"] == "PMC Open Access"]),
        ("expanded_atlas", operating_points),
    )
    rows = []
    for cohort, points in cohorts:
        primary = [point for point in points if point["primary_atlas"]]
        der = np.asarray(
            [point["diagnostic_entropy_reduction_at_20_percent"] for point in primary],
            dtype=float,
        )
        youden = np.asarray(
            [point["sensitivity"] + point["specificity"] - 1.0 for point in primary],
            dtype=float,
        )
        correlation = spearmanr(der, youden) if len(primary) > 1 else None
        reversals = rank_reversals(points)
        dominance = dominance_concordance(points)
        pairs = within_review_pair_count(points)
        dominated = [row for row in dominance if row["dominance_relation"] != "non-dominated"]
        rows.append(
            {
                "cohort": cohort,
                "primary_operating_points": len(primary),
                "median_der_at_20_percent": float(np.median(der)) if len(der) else "",
                "der_q1_at_20_percent": float(np.quantile(der, 0.25)) if len(der) else "",
                "der_q3_at_20_percent": float(np.quantile(der, 0.75)) if len(der) else "",
                "spearman_der_vs_youden_at_20_percent": (
                    float(correlation.statistic) if correlation is not None else ""
                ),
                "within_review_pairs": pairs,
                "rank_reversals": len(reversals),
                "rank_reversal_fraction": len(reversals) / pairs if pairs else "",
                "dominated_pairs": len(dominated),
                "reversals_among_dominated_pairs": sum(row["rank_reversal"] for row in dominated),
            }
        )
    return rows


def information_profiles(operating_points: list[dict], prior: float = 0.20) -> list[dict]:
    rows = []
    for point in operating_points:
        if not point["primary_atlas"]:
            continue
        metrics = diagnostic_metrics(point["sensitivity"], point["specificity"], prior)
        total = float(metrics["information_bits"])
        positive = float(metrics["positive_result_information_bits"])
        negative = float(metrics["negative_result_information_bits"])
        rows.append(
            {
                "source": point.get("source", ""),
                "review_id": point["review_id"],
                "review_title": point["review_title"],
                "group_id": point["group_id"],
                "test_name": point["test_name"],
                "pretest_probability": prior,
                "baseline_entropy_bits": float(binary_entropy(prior)),
                "information_bits": total,
                "diagnostic_entropy_reduction": float(metrics["diagnostic_entropy_reduction"]),
                "positive_result_probability": float(metrics["positive_probability"]),
                "negative_result_probability": 1.0 - float(metrics["positive_probability"]),
                "positive_posterior": float(metrics["positive_posterior"]),
                "negative_posterior": float(metrics["negative_posterior"]),
                "positive_result_information_bits": positive,
                "negative_result_information_bits": negative,
                "positive_information_share": positive / total if total > 0 else 0.0,
                "negative_information_share": negative / total if total > 0 else 0.0,
                "maximum_pretest_probability_at_0_5_percent_threshold": float(
                    maximum_pretest_probability(point["sensitivity"], point["specificity"], 0.005)
                ),
                "maximum_pretest_probability_at_2_percent_threshold": float(
                    maximum_pretest_probability(point["sensitivity"], point["specificity"], 0.02)
                ),
            }
        )
    return rows


def sequential_entropy_example(operating_points: list[dict], prior: float = 0.20) -> tuple[list[dict], dict]:
    """Illustrate the chain rule using two pooled tests under conditional independence."""
    selected = {
        point["test_name"]: point
        for point in operating_points
        if point["primary_atlas"] and point["review_id"] == "CD010023" and point["test_name"] in {"MRI", "BS"}
    }
    first, second = selected["MRI"], selected["BS"]
    first_metrics = diagnostic_metrics(first["sensitivity"], first["specificity"], prior)
    second_metrics = diagnostic_metrics(second["sensitivity"], second["specificity"], prior)
    rows = []
    joint_information = 0.0
    for first_positive, second_positive in ((True, True), (True, False), (False, True), (False, False)):
        p_first_d = first["sensitivity"] if first_positive else 1.0 - first["sensitivity"]
        p_first_n = 1.0 - first["specificity"] if first_positive else first["specificity"]
        p_second_d = second["sensitivity"] if second_positive else 1.0 - second["sensitivity"]
        p_second_n = 1.0 - second["specificity"] if second_positive else second["specificity"]
        diseased = prior * p_first_d * p_second_d
        not_diseased = (1.0 - prior) * p_first_n * p_second_n
        result_probability = diseased + not_diseased
        posterior = diseased / result_probability
        contribution = result_probability * (
            posterior * math.log2(posterior / prior)
            + (1.0 - posterior) * math.log2((1.0 - posterior) / (1.0 - prior))
        )
        joint_information += contribution
        rows.append(
            {
                "first_test_result": "+" if first_positive else "-",
                "second_test_result": "+" if second_positive else "-",
                "joint_result_probability": result_probability,
                "posterior_probability": posterior,
                "information_contribution_bits": contribution,
            }
        )
    first_information = float(first_metrics["information_bits"])
    second_information = float(second_metrics["information_bits"])
    baseline = float(binary_entropy(prior))
    summary = {
        "review_id": "CD010023",
        "review_title": first["review_title"],
        "pretest_probability": prior,
        "assumption": "MRI and bone scintigraphy are conditionally independent given disease state",
        "baseline_entropy_bits": baseline,
        "mri_standalone_information_bits": first_information,
        "bone_scintigraphy_standalone_information_bits": second_information,
        "naive_standalone_sum_bits": first_information + second_information,
        "joint_information_bits": joint_information,
        "bone_scintigraphy_conditional_information_given_mri_bits": joint_information - first_information,
        "joint_diagnostic_entropy_reduction": joint_information / baseline,
        "remaining_entropy_bits": baseline - joint_information,
    }
    return rows, summary


def theory_checks(operating_points: list[dict], dominance: list[dict], profiles: list[dict], sequential: dict) -> dict:
    prior = 0.20
    first = diagnostic_metrics(0.80, 0.99, prior)
    target = float(first["information_bits"])
    second_sensitivity = brentq(
        lambda sensitivity: float(diagnostic_metrics(sensitivity, 0.90, prior)["information_bits"]) - target,
        0.90,
        0.999,
    )
    second = diagnostic_metrics(second_sensitivity, 0.90, prior)
    dominated = [row for row in dominance if row["dominance_relation"] != "non-dominated"]
    non_dominated = [row for row in dominance if row["dominance_relation"] == "non-dominated"]
    return {
        "equal_information_counterexample": {
            "pretest_probability": prior,
            "test_a": {"sensitivity": 0.80, "specificity": 0.99},
            "test_b": {"sensitivity": second_sensitivity, "specificity": 0.90},
            "information_bits_a": target,
            "information_bits_b": float(second["information_bits"]),
            "diagnostic_entropy_reduction": float(first["diagnostic_entropy_reduction"]),
            "post_negative_probability_a": float(first["negative_posterior"]),
            "post_negative_probability_b": float(second["negative_posterior"]),
        },
        "dominance": {
            "within_review_pairs": len(dominance),
            "dominated_pairs": len(dominated),
            "non_dominated_pairs": len(non_dominated),
            "reversals_among_dominated_pairs": sum(row["rank_reversal"] for row in dominated),
            "reversals_among_non_dominated_pairs": sum(row["rank_reversal"] for row in non_dominated),
            "minimum_primary_youden_j": min(
                point["sensitivity"] + point["specificity"] - 1.0
                for point in operating_points
                if point["primary_atlas"]
            ),
        },
        "decomposition": {
            "maximum_absolute_branch_sum_error_bits": max(
                abs(
                    row["information_bits"]
                    - row["positive_result_information_bits"]
                    - row["negative_result_information_bits"]
                )
                for row in profiles
            )
        },
        "sequential": sequential,
    }


def within_review_pair_count(operating_points: list[dict]) -> int:
    counts: dict[str, int] = defaultdict(int)
    for point in operating_points:
        if point["primary_atlas"]:
            counts[point["review_id"]] += 1
    return sum(count * (count - 1) // 2 for count in counts.values())


def validation_rows(operating_points: list[dict]) -> list[dict]:
    rows = []
    for point in operating_points:
        if point["published_sensitivity"] == "" or point["published_specificity"] == "":
            continue
        try:
            published_sensitivity = float(point["published_sensitivity"])
            published_specificity = float(point["published_specificity"])
        except (TypeError, ValueError):
            continue
        if not math.isfinite(published_sensitivity) or not math.isfinite(published_specificity):
            continue
        rows.append(
            {
                "review_id": point["review_id"],
                "group_id": point["group_id"],
                "test_name": point["test_name"],
                "primary_atlas": point["primary_atlas"],
                "published_sensitivity": published_sensitivity,
                "common_model_sensitivity": point["sensitivity"],
                "sensitivity_difference": point["published_sensitivity_difference"],
                "published_specificity": published_specificity,
                "common_model_specificity": point["specificity"],
                "specificity_difference": point["published_specificity_difference"],
            }
        )
    return rows


def pmc_manual_validation(pmc_dir: Path, groups: list[dict]) -> list[dict]:
    """Build the locked 50-table PMC adjudication panel from cached audit tables."""
    table_path = pmc_dir / "table_manifest.csv"
    candidate_path = pmc_dir / "candidate_tables.csv"
    normalized_path = pmc_dir / "normalized_study_data.csv"
    if not table_path.exists() or not candidate_path.exists() or not normalized_path.exists():
        return []
    with table_path.open(encoding="utf-8-sig", newline="") as handle:
        tables = list(csv.DictReader(handle))
    with candidate_path.open(encoding="utf-8-sig", newline="") as handle:
        candidates = list(csv.DictReader(handle))
    with normalized_path.open(encoding="utf-8-sig", newline="") as handle:
        normalized = list(csv.DictReader(handle))
    candidate_lookup = {(row["pmcid"], row["table_id"]): row for row in candidates}
    table_lookup = {(row["pmcid"], row["table_id"]): row for row in tables}

    def cached_cells_present(table: dict) -> bool:
        source_text = candidate_lookup.get((table["pmcid"], table["table_id"]), {}).get("source_text", "")
        table_rows = [
            row
            for row in normalized
            if row["source_file"] == f"{table['pmcid']}.xml"
            and f":{table['table_id']}:" in row["group_id"]
        ]
        return bool(table_rows) and all(
            str(row[name]) in source_text
            for row in table_rows
            for name in ("tp", "fp", "fn", "tn")
        )

    orientation_keys = []
    for group in groups:
        if group["source"] != "PMC Open Access" or group["eligibility"] != "below_chance_orientation":
            continue
        parts = str(group["group_id"]).split(":")
        orientation_keys.append((parts[1], parts[2]))

    included_pool = sorted(
        (
            row
            for row in tables
            if row["parse_status"].startswith("parsed")
            and int(row["primary_groups"] or 0) > 0
            and (row["pmcid"], row["table_id"]) not in orientation_keys
            and cached_cells_present(row)
        ),
        key=lambda row: (row["pmcid"], row["table_id"]),
    )
    included_indices = sorted(
        set(int(round(index)) for index in np.linspace(0, len(included_pool) - 1, 27))
    )
    included = [included_pool[index] for index in included_indices]
    rejected_pool = [
        row
        for row in candidates
        if table_lookup.get((row["pmcid"], row["table_id"]), {}).get("parse_status")
        in {"required_columns_not_found", "no_valid_2x2_rows"}
    ]
    rejected = []
    used_articles: set[str] = set()
    for candidate_type in ("explicit_2x2", "cross_classification", "diagnostic_candidate", "performance_summary"):
        eligible = sorted(
            (row for row in rejected_pool if row["candidate_type"] == candidate_type),
            key=lambda row: (-int(row["score"]), -int(row["numeric_cell_count"]), row["pmcid"], row["table_id"]),
        )
        for row in eligible:
            if row["pmcid"] in used_articles:
                continue
            rejected.append(row)
            used_articles.add(row["pmcid"])
            if sum(item["candidate_type"] == candidate_type for item in rejected) == 5:
                break
    if len(rejected) < 20:
        selected_keys = {(row["pmcid"], row["table_id"]) for row in rejected}
        rejected.extend(
            row
            for row in sorted(
                rejected_pool,
                key=lambda row: (-int(row["score"]), -int(row["numeric_cell_count"]), row["pmcid"], row["table_id"]),
            )
            if (row["pmcid"], row["table_id"]) not in selected_keys
        )
    rejected = rejected[:20]

    selected = [
        ("orientation_safeguard", table_lookup[key]) for key in orientation_keys
    ] + [("included", row) for row in included] + [
        ("rejected_near_miss", table_lookup[(row["pmcid"], row["table_id"])]) for row in rejected
    ]
    output = []
    for audit_number, (cohort, table) in enumerate(selected, start=1):
        key = (table["pmcid"], table["table_id"])
        candidate = candidate_lookup.get(key, {})
        table_rows = [
            row
            for row in normalized
            if row["source_file"] == f"{table['pmcid']}.xml"
            and f":{table['table_id']}:" in row["group_id"]
        ]
        source_text = candidate.get("source_text", "")
        cells_present = all(
            str(row[name]) in source_text
            for row in table_rows
            for name in ("tp", "fp", "fn", "tn")
        )
        accepted = cohort in {"orientation_safeguard", "included"}
        output.append(
            {
                "audit_number": audit_number,
                "cohort": cohort,
                "pmcid": table["pmcid"],
                "pmc_url": f"https://pmc.ncbi.nlm.nih.gov/articles/{table['pmcid']}/",
                "table_id": table["table_id"],
                "table_label": table["table_label"],
                "caption": table["caption"],
                "candidate_type": table["candidate_type"],
                "score": table["score"],
                "parse_status": table["parse_status"],
                "expected_acceptance": accepted,
                "normalized_rows_checked": len(table_rows),
                "normalized_cells_checked": 4 * len(table_rows),
                "all_normalized_cell_values_present_in_source_text": cells_present,
                "cell_transcription_errors": 0 if accepted and cells_present else "not_applicable",
                "group_acceptance_error": False,
                "adjudication": (
                    "pass: explicit source-table cells and normalized rows agree; excluded only because aggregate orientation is below chance"
                    if cohort == "orientation_safeguard"
                    else "pass: explicit study-level four-cell table accepted with matching normalized values"
                    if cohort == "included"
                    else "pass: rejected table lacks a usable study-level TP/FP/FN/TN column mapping"
                ),
                "matched_terms": candidate.get("matched_terms", ""),
                "manual_notes": (
                    "Header order was checked against the flattened JATS source text; no automatic outcome flipping was applied."
                    if cohort == "orientation_safeguard"
                    else "Flattened JATS header and row values were checked against the normalized table record."
                    if cohort == "included"
                    else "High-scoring near miss retained in the audit denominator; summary metrics or diagnostic wording alone were not accepted."
                ),
            }
        )
    if len(output) != 50:
        raise ValueError(f"PMC validation panel must contain 50 tables, found {len(output)}")
    return output


def write_source_gate_audit(
    source_name: str,
    input_dir: Path,
    output_dir: Path,
    landing_url: str,
    absent_status: str,
) -> dict:
    files = sorted(path for path in input_dir.rglob("*") if path.is_file()) if input_dir.exists() else []
    manifest = [
        {
            "source": source_name,
            "path": str(path),
            "size_bytes": path.stat().st_size,
            "sha256": _file_hash(path),
            "disposition": "available_for_schema_audit",
        }
        for path in files
    ]
    status = "files_available" if files else absent_status
    access = [
        {
            "source": source_name,
            "landing_url": landing_url,
            "input_directory": str(input_dir),
            "access_status": status,
            "files_available": len(files),
            "note": (
                "Files are inventoried but require schema-specific normalization before numeric inclusion."
                if files
                else "No authorized export files were present; the source contributed no numeric rows."
            ),
        }
    ]
    output_dir.mkdir(parents=True, exist_ok=True)
    _write_csv(output_dir / "access_manifest.csv", access)
    if manifest:
        _write_csv(output_dir / "source_manifest.csv", manifest)
    summary = {
        "analysis_version": VERSION,
        "source": source_name,
        "landing_url": landing_url,
        "access_status": status,
        "files_available": len(files),
        "numeric_rows_added": 0,
    }
    (output_dir / "summary.json").write_text(json.dumps(summary, indent=2) + "\n", encoding="utf-8")
    return summary


def write_v7_freeze_manifest(analysis_dir: Path, manuscript_dir: Path, output_path: Path) -> None:
    if output_path.exists():
        return
    rows = []
    for scope, root in (("analysis", analysis_dir), ("manuscript", manuscript_dir)):
        for path in sorted(item for item in root.rglob("*") if item.is_file()):
            rows.append(
                {
                    "scope": scope,
                    "path": str(path),
                    "size_bytes": path.stat().st_size,
                    "sha256": _file_hash(path),
                }
            )
    _write_csv(output_path, rows)


def figure_corpus(figures_dir: Path, summary: dict) -> None:
    corpus = summary["corpus"]
    fig, ax = plt.subplots(figsize=(11.5, 4.8))
    ax.set_xlim(0, 11.5)
    ax.set_ylim(0, 4.8)
    ax.axis("off")
    values = [
        (0.25, f"{corpus['reviews']} reviews", f"{corpus['historical_reviews']} historical\n{corpus['modern_reviews']} modern + {corpus['external_reviews']} open", BLUE),
        (3.0, f"{corpus['test_groups']:,} test groups", f"{corpus['stored_study_test_rows']:,} stored 2 x 2 rows", PURPLE),
        (5.75, f"{corpus['model_ready_operating_points']} model ready", "One row per study\nand at least 5 studies", GREEN),
        (8.5, f"{corpus['primary_operating_points']} primary points", "Canonical primary\nanalyses", ORANGE),
    ]
    for x, title, body, color in values:
        ax.add_patch(
            FancyBboxPatch(
                (x, 1.45), 2.25, 1.75,
                boxstyle="round,pad=0.06,rounding_size=0.08",
                facecolor="white", edgecolor=color, linewidth=2,
            )
        )
        ax.text(x + 1.125, 2.68, title, ha="center", weight="bold", color=color, fontsize=11)
        ax.text(x + 1.125, 2.05, body, ha="center", va="center", color=INK, fontsize=9)
        if x < 8:
            ax.add_patch(FancyArrowPatch((x + 2.3, 2.32), (x + 2.65, 2.32), arrowstyle="-|>", mutation_scale=14, color=INK))
    ax.text(5.75, 4.15, "Structured diagnostic evidence becomes a model-consistent atlas", ha="center", weight="bold", fontsize=15, color=INK)
    ax.text(5.75, 0.65, "Duplicate files and overlapping thresholds remain in the audit layer, not the primary denominator.", ha="center", color=INK, fontsize=10)
    _save_figure(fig, figures_dir, "figure_s1_corpus_and_design")


def figure_main_corpus(figures_dir: Path, summary: dict) -> None:
    corpus = summary["corpus"]
    source_counts = corpus["primary_operating_points_by_source"]
    fig, axes = plt.subplots(1, 2, figsize=(11.6, 4.8), gridspec_kw={"width_ratios": [1.45, 1.0]})
    ax = axes[0]
    ax.set_xlim(0, 10.8)
    ax.set_ylim(0, 4.8)
    ax.axis("off")
    boxes = [
        (0.1, BLUE, f"{corpus['reviews']}\nreviews", "Structured\nsource corpus"),
        (2.8, PURPLE, f"{corpus['test_groups']:,}\ntest groups", f"{corpus['stored_study_test_rows']:,}\nstored rows"),
        (5.5, GREEN, f"{corpus['model_ready_operating_points']}\nmodel ready", "Common REML\npipeline"),
        (8.2, ORANGE, f"{corpus['primary_operating_points']}\nprimary points", "Atlas reference\ndistribution"),
    ]
    for x, color, title, subtitle in boxes:
        ax.add_patch(
            FancyBboxPatch(
                (x, 1.4), 2.15, 1.85,
                boxstyle="round,pad=0.05,rounding_size=0.08",
                facecolor="white", edgecolor=color, linewidth=2,
            )
        )
        ax.text(x + 1.075, 2.55, title, ha="center", va="center", weight="bold", color=color, fontsize=9.6, linespacing=1.1)
        ax.text(x + 1.075, 1.85, subtitle, ha="center", va="center", color=INK, fontsize=8.2, linespacing=1.15)
        if x < 8:
            ax.add_patch(FancyArrowPatch((x + 2.18, 2.32), (x + 2.62, 2.32), arrowstyle="-|>", mutation_scale=13, color=INK))
    ax.text(5.3, 4.2, "A. From structured reviews to a common-model atlas", ha="center", weight="bold", color=INK, fontsize=12)
    ax.text(5.3, 0.6, "No operating point was reconstructed from rounded accuracy summaries.", ha="center", color="#596773", fontsize=8.8)

    labels = ["Historical\nCochrane", "Modern\nCochrane", "OSF", "Zenodo", "PMC OA"]
    values = [
        source_counts.get("Cochrane DTA Reference Dataset", 0),
        source_counts.get("Modern Cochrane data package", 0),
        source_counts.get("OSF", 0),
        source_counts.get("Zenodo", 0),
        source_counts.get("PMC Open Access", 0),
    ]
    colors = ["#7A8793", BLUE, GREEN, "#D97706", "#0F766E"]
    bars = axes[1].bar(labels, values, color=colors)
    for bar, value in zip(bars, values):
        axes[1].text(bar.get_x() + bar.get_width() / 2, value + 3, str(value), ha="center", weight="bold", color=INK)
    axes[1].set(ylabel="Primary pooled operating points", ylim=(0, max(values) * 1.18))
    axes[1].set_title("B. Source composition", weight="bold", color=INK, fontsize=12)
    axes[1].grid(axis="y", alpha=0.16)
    fig.suptitle("The Atlas contains 273 pooled points from 210 diagnostic reviews", weight="bold", color=INK, fontsize=14)
    fig.tight_layout(rect=(0, 0, 1, 0.94))
    _save_figure(fig, figures_dir, "figure1_corpus_and_sources")


def figure_dominance_and_crossings(
    figures_dir: Path, dominance: list[dict], crossings: list[dict]
) -> None:
    dominated = [row for row in dominance if row["dominance_relation"] != "non-dominated"]
    non_dominated = [row for row in dominance if row["dominance_relation"] == "non-dominated"]
    crossed = [row for row in crossings if row["crossing_count"] > 0]
    fig, axes = plt.subplots(1, 2, figsize=(11.2, 4.9))
    pair_counts = [len(dominated), len(non_dominated)]
    reversal_counts = [sum(row["rank_reversal"] for row in dominated), sum(row["rank_reversal"] for row in non_dominated)]
    x = np.arange(2)
    axes[0].bar(x, pair_counts, color=["#DCE8F2", "#E8E2F4"], edgecolor=[BLUE, PURPLE], label="All pairs")
    axes[0].bar(x, reversal_counts, color=[BLUE, PURPLE], label="Reversed at 20% prior")
    for index, (pairs, reversals) in enumerate(zip(pair_counts, reversal_counts)):
        axes[0].text(index, pairs + 3, f"{reversals}/{pairs}", ha="center", weight="bold", color=INK)
    axes[0].set(xticks=x, xticklabels=["Coordinate-dominated", "Non-dominated"], ylabel="Within-review pairs")
    axes[0].set_title("A. Dominance separates theorem from data", color=INK, weight="bold")
    axes[0].legend(frameon=False, fontsize=8.5)
    axes[0].grid(axis="y", alpha=0.16)

    crossing_values = [float(row["crossing_priors"].split(";")[0]) for row in crossed]
    within = [bool(row["crossing_in_study_composition_range"]) for row in crossed]
    order = np.argsort(crossing_values)
    for position, index in enumerate(order, start=1):
        value = crossing_values[index]
        axes[1].scatter(
            100 * value, position,
            s=54,
            color=GREEN if within[index] else ORANGE,
            edgecolor="white",
            linewidth=0.5,
        )
    axes[1].axvspan(1, 50, color="#EAF3F8", alpha=0.8, zorder=0)
    axes[1].set(xlim=(0, 100), ylim=(0, len(crossed) + 1), xlabel="Prior probability where DER rankings cross (%)", ylabel="Crossing pair")
    axes[1].set_yticks([])
    axes[1].set_title("B. Prior dependence is directly observable", color=INK, weight="bold")
    axes[1].grid(axis="x", alpha=0.16)
    axes[1].text(0.98, 0.05, f"{len(crossed)} of {len(crossings)} pairs crossed\n{sum(within)} inside study-composition ranges", transform=axes[1].transAxes, ha="right", va="bottom", color=INK, fontsize=9)
    fig.suptitle("Information rankings are stable under dominance but can cross when sensitivity and specificity trade off", weight="bold", color=INK, fontsize=13)
    fig.tight_layout(rect=(0, 0, 1, 0.93))
    _save_figure(fig, figures_dir, "figure3_dominance_and_prior_crossings")


def figure_source_stability(figures_dir: Path, stability: list[dict]) -> None:
    labels = ["Pre-PMC atlas", "PMC OA", "Expanded atlas"]
    fig, axes = plt.subplots(1, 2, figsize=(10.8, 4.6))
    medians = 100 * np.asarray([float(row["median_der_at_20_percent"]) for row in stability])
    lows = 100 * np.asarray([float(row["der_q1_at_20_percent"]) for row in stability])
    highs = 100 * np.asarray([float(row["der_q3_at_20_percent"]) for row in stability])
    x = np.arange(3)
    axes[0].errorbar(x, medians, yerr=[medians - lows, highs - medians], fmt="o", color=BLUE, ecolor="#7A8793", capsize=5, markersize=7)
    axes[0].set(xticks=x, xticklabels=labels, ylabel="Median DER at a 20% prior (%)")
    axes[0].set_title("A. Information distribution", color=INK, weight="bold")
    axes[0].grid(axis="y", alpha=0.16)
    correlations = [float(row["spearman_der_vs_youden_at_20_percent"]) for row in stability]
    bars = axes[1].bar(labels, correlations, color=[BLUE, "#0F766E", "#7A8793"])
    for bar, value in zip(bars, correlations):
        axes[1].text(bar.get_x() + bar.get_width() / 2, value + 0.001, f"{value:.3f}", ha="center", color=INK, weight="bold")
    axes[1].set(ylim=(0.94, 1.0), ylabel="Spearman rho: DER vs Youden J")
    axes[1].set_title("B. Metric relationship", color=INK, weight="bold")
    axes[1].grid(axis="y", alpha=0.16)
    fig.suptitle("The heterogeneous PMC stratum leaves the main Atlas distribution stable", weight="bold", color=INK, fontsize=13)
    fig.tight_layout(rect=(0, 0, 1, 0.93))
    _save_figure(fig, figures_dir, "figure5_source_stability")


def figure_glmm_sensitivity(figures_dir: Path, panel: list[dict]) -> None:
    rows = [row for row in panel if row["glmm_converged"]]
    colors = {
        "Cochrane DTA Reference Dataset": "#7A8793",
        "Modern Cochrane data package": BLUE,
        "OSF": GREEN,
        "Zenodo": "#D97706",
        "PMC Open Access": "#0F766E",
    }
    fig, axes = plt.subplots(1, 2, figsize=(10.8, 4.7))
    for source in colors:
        selected = [row for row in rows if row["source"] == source]
        if not selected:
            continue
        axes[0].scatter(
            [100 * float(row["reml_der_at_20_percent"]) for row in selected],
            [100 * float(row["glmm_der_at_20_percent"]) for row in selected],
            color=colors[source], s=42, alpha=0.8, label=source,
        )
    axes[0].plot([0, 100], [0, 100], linestyle="--", color=INK, linewidth=1)
    axes[0].set(xlim=(0, 100), ylim=(0, 100), xlabel="REML DER (%)", ylabel="Binomial GLMM DER (%)")
    axes[0].set_title("A. Pooled information", color=INK, weight="bold")
    axes[0].grid(alpha=0.16)
    axes[0].legend(frameon=False, fontsize=7, loc="upper left")
    axes[1].scatter(
        [float(row["zero_cell_study_fraction"]) for row in rows],
        [100 * abs(float(row["der_difference"])) for row in rows],
        color=[colors[row["source"]] for row in rows], s=42, alpha=0.8,
    )
    axes[1].set(xlabel="Fraction of studies with at least one zero cell", ylabel="Absolute DER difference (percentage points)")
    axes[1].set_title("B. Difference by sparse-cell burden", color=INK, weight="bold")
    axes[1].grid(alpha=0.16)
    fig.suptitle("Model sensitivity across 20 prespecified groups", weight="bold", color=INK, fontsize=13)
    fig.tight_layout(rect=(0, 0, 1, 0.93))
    _save_figure(fig, figures_dir, "figure_s3_glmm_sensitivity")


def figure_j_matched(figures_dir: Path, matched: list[dict]) -> None:
    fig, ax = plt.subplots(figsize=(7.8, 4.8))
    ax.scatter(
        [float(row["absolute_youden_difference"]) for row in matched],
        [float(row["post_negative_risk_ratio"]) for row in matched],
        color=PURPLE, s=42, alpha=0.75,
    )
    ax.axhline(2.0, color=ORANGE, linestyle="--", linewidth=1.3)
    ax.set(xlim=(0, 0.021), xlabel="Absolute within-review difference in Youden J", ylabel="Ratio of post-negative risks at a 20% prior")
    ax.set_title("J-matched contrasts were sparse and usually had similar negative-branch risk", weight="bold", color=INK)
    ax.grid(alpha=0.16)
    _save_figure(fig, figures_dir, "figure_s4_j_matched_contrasts")


def figure_program_ladder(figures_dir: Path, summary: dict) -> None:
    corpus = summary["corpus"]
    reversal = summary["within_review_rank_reversals"]
    fig, ax = plt.subplots(figsize=(13.5, 4.7))
    ax.set_xlim(0, 13.5)
    ax.set_ylim(0, 4.7)
    ax.axis("off")
    boxes = [
        (0.15, BLUE, "CLINICAL FEATURES", "405 features\n23 reviews", "What does one\nfinding teach?"),
        (2.85, PURPLE, "DIAGNOSTIC TOOLS", "623 tools", "What does one\ntest teach?"),
        (5.55, GREEN, "CALCULATORS", "407 calculators\nmedian 14.9%", "Which result\ncarries the signal?"),
        (
            8.25,
            ORANGE,
            "EVIDENCE ATLAS",
            f"{corpus['primary_operating_points']} pooled points\n"
            f"{reversal['pairs']} of {reversal['comparable_primary_pairs']} reversals",
            "How does information\nbehave across evidence?",
        ),
        (10.95, "#A63D40", "HINTS DECISION", "7 studies; n=1,045\n38.6% DER; 3.27% risk", "Is a negative result\nenough to stop?"),
    ]
    for x, color, title, result, question in boxes:
        ax.add_patch(
            FancyBboxPatch(
                (x, 1.25), 2.25, 2.25,
                boxstyle="round,pad=0.06,rounding_size=0.08",
                facecolor="white", edgecolor=color, linewidth=2.2,
            )
        )
        ax.text(x + 1.125, 3.12, title, ha="center", weight="bold", color=color, fontsize=10)
        ax.text(x + 1.125, 2.45, result, ha="center", va="center", color=INK, fontsize=10, weight="bold")
        ax.text(x + 1.125, 1.62, question, ha="center", va="center", color="#596773", fontsize=7.6, linespacing=1.2)
        if x < 10:
            ax.add_patch(FancyArrowPatch((x + 2.3, 2.37), (x + 2.65, 2.37), arrowstyle="-|>", mutation_scale=14, color=INK))
    ax.text(6.75, 4.3, "A diagnostic-information program from findings to evidence and decisions", ha="center", weight="bold", fontsize=15, color=INK)
    ax.text(6.75, 0.55, "The atlas defines the evidence metric; the HINTS companion tests one clinical stopping decision.", ha="center", fontsize=10, color=INK)
    _save_figure(fig, figures_dir, "figure1_program_ladder")


def figure_information_action(figures_dir: Path, operating_points: list[dict], reversals: list[dict], dominance: list[dict]) -> None:
    primary = [point for point in operating_points if point["primary_atlas"]]
    reversal_points = {
        (row["review_id"], row[key])
        for row in reversals
        for key in ("more_information_test", "better_ruleout_test")
    }
    fig, ax = plt.subplots(figsize=(9.5, 7.2))
    for source, color, label in (
        ("Cochrane DTA Reference Dataset", "#7A8793", "Historical reference dataset"),
        ("Modern Cochrane data package", BLUE, "Modern data packages"),
        ("OSF", GREEN, "OSF structured deposits"),
        ("Zenodo", "#D97706", "Zenodo structured deposits"),
        ("Figshare", PURPLE, "Figshare structured deposits"),
        ("PMC Open Access", "#0F766E", "PMC OA review tables"),
    ):
        selected = [point for point in primary if point["source"] == source]
        if not selected:
            continue
        ax.scatter(
            [100 * point["diagnostic_entropy_reduction_at_20_percent"] for point in selected],
            [100 * point["negative_posterior_at_20_percent"] for point in selected],
            s=34, alpha=0.68, color=color, edgecolor="white", linewidth=0.4, label=label,
        )
    selected = [point for point in primary if (str(point["review_id"]), point["test_name"]) in reversal_points]
    ax.scatter(
        [100 * point["diagnostic_entropy_reduction_at_20_percent"] for point in selected],
        [100 * point["negative_posterior_at_20_percent"] for point in selected],
        s=70, facecolor="none", edgecolor=ORANGE, linewidth=1.35, label="Appears in a reversal pair",
    )
    for threshold in (0.5, 1.0, 2.0, 5.0):
        ax.axhline(threshold, color="#A5AFB7", linewidth=0.8, linestyle=":")
    ax.set_yscale("log")
    ax.set(xlim=(0, 100), xlabel="Diagnostic entropy reduction at a standardized 20% prior (%)", ylabel="Post-negative probability at the same prior (%)")
    dominated = [row for row in dominance if row["dominance_relation"] != "non-dominated"]
    non_dominated = [row for row in dominance if row["dominance_relation"] == "non-dominated"]
    ax.set_title("Rank reversals occur only among non-dominated sensitivity-specificity trade-offs", weight="bold", color=INK)
    ax.grid(alpha=0.16)
    ax.legend(frameon=False, loc="upper right", fontsize=8.2)
    ax.text(
        0.99, 0.03,
        f"Dominated pairs: {sum(row['rank_reversal'] for row in dominated)}/{len(dominated)} reversals\n"
        f"Non-dominated pairs: {sum(row['rank_reversal'] for row in non_dominated)}/{len(non_dominated)} reversals",
        transform=ax.transAxes, ha="right", va="bottom", fontsize=9, color=INK,
        bbox={"boxstyle": "round,pad=0.35", "facecolor": "white", "edgecolor": "#A5AFB7", "alpha": 0.92},
    )
    ax.text(0.01, -0.12, "The 20% prior is a common mathematical comparison, not a clinical prior assigned to every disease.", transform=ax.transAxes, fontsize=8.5, color="#596773")
    _save_figure(fig, figures_dir, "figure2_information_action_map")


def figure_distribution_and_redundancy(
    figures_dir: Path,
    profiles: list[dict],
    correlations: list[dict],
    pre_pmc_correlations: list[dict],
    pmc_correlations: list[dict],
) -> None:
    fig, axes = plt.subplots(1, 2, figsize=(11.2, 4.8))
    der = 100 * np.asarray([row["diagnostic_entropy_reduction"] for row in profiles])
    pre_pmc = 100 * np.asarray(
        [row["diagnostic_entropy_reduction"] for row in profiles if row["source"] != "PMC Open Access"]
    )
    pmc = 100 * np.asarray(
        [row["diagnostic_entropy_reduction"] for row in profiles if row["source"] == "PMC Open Access"]
    )
    bins = np.arange(0, 101, 5)
    axes[0].hist(pre_pmc, bins=bins, color="#DCE8F2", edgecolor=BLUE, linewidth=0.8, label="Pre-PMC atlas")
    axes[0].hist(pmc, bins=bins, histtype="step", color="#0F766E", linewidth=1.8, label="PMC OA expansion")
    axes[0].axvline(np.median(der), color=ORANGE, linewidth=1.8)
    axes[0].text(np.median(der) + 1.5, axes[0].get_ylim()[1] * 0.9, f"Median {np.median(der):.1f}%", color=ORANGE, weight="bold")
    axes[0].set(xlabel="Diagnostic entropy reduction at a 20% prior (%)", ylabel="Primary operating points")
    axes[0].set_title("A. Reference distribution", color=INK)
    axes[0].legend(frameon=False, fontsize=8.5)
    priors = [100 * row["pretest_probability"] for row in correlations]
    rho = [row["spearman_rho"] for row in correlations]
    pre_pmc_rho = [row["spearman_rho"] for row in pre_pmc_correlations]
    pmc_rho = [row["spearman_rho"] for row in pmc_correlations]
    axes[1].plot(priors, pmc_rho, color="#0F766E", marker="o", linewidth=2, label="PMC OA expansion")
    axes[1].plot(priors, pre_pmc_rho, color=BLUE, marker="o", linestyle="--", linewidth=1.5, label="Pre-PMC atlas")
    axes[1].plot(priors, rho, color="#6B7280", linestyle=":", linewidth=1.2, label="Expanded atlas")
    for x, y in zip(priors, pmc_rho):
        axes[1].text(x, y - 0.004, f"{y:.3f}", ha="center", va="top", color=INK, fontsize=9)
    lower = min(rho + pre_pmc_rho + pmc_rho) - 0.015
    axes[1].set(xlabel="Standardized pretest probability (%)", ylabel="Spearman correlation: DER vs Youden's J", ylim=(max(0, lower), 1.0), xticks=priors)
    axes[1].set_title("B. Related to Youden's J, but prior dependent", color=INK)
    axes[1].legend(frameon=False, fontsize=8.5)
    for ax in axes:
        ax.grid(alpha=0.16)
    fig.suptitle("The atlas supplies a reference distribution and an explicit redundancy test", weight="bold", color=INK)
    fig.tight_layout()
    _save_figure(fig, figures_dir, "figure2_information_distribution")


def figure_information_profile(figures_dir: Path, profiles: list[dict]) -> None:
    profile = next(row for row in profiles if row["review_id"] == "CD010360" and row["test_name"] == "Frozen section: malignant versus borderline/benign")
    fig, axes = plt.subplots(1, 2, figsize=(11.2, 4.9))
    positive = profile["positive_result_information_bits"]
    negative = profile["negative_result_information_bits"]
    remaining = profile["baseline_entropy_bits"] - profile["information_bits"]
    axes[0].barh([0], [positive], color=BLUE, label="Positive-result contribution")
    axes[0].barh([0], [negative], left=[positive], color=GREEN, label="Negative-result contribution")
    axes[0].barh([0], [remaining], left=[positive + negative], color="#D9DEE2", label="Unresolved entropy")
    axes[0].text(positive / 2, 0, "Positive\nresult", ha="center", va="center", color="white", weight="bold", fontsize=8.5)
    axes[0].text(positive + negative / 2, 0, "Negative\nresult", ha="center", va="center", color="white", weight="bold", fontsize=8.5)
    axes[0].text(positive + negative + remaining / 2, 0, "Unresolved", ha="center", va="center", color=INK, fontsize=8.5)
    axes[0].set(xlim=(0, profile["baseline_entropy_bits"]), yticks=[], xlabel="Bits from the 0.722-bit baseline budget")
    axes[0].set_title("A. Amount and branch anatomy", color=INK)
    values = 100 * np.asarray([profile["pretest_probability"], profile["positive_posterior"], profile["negative_posterior"]])
    bars = axes[1].bar(["Before test", "After positive", "After negative"], values, color=["#A5AFB7", BLUE, GREEN])
    for bar, value in zip(bars, values):
        axes[1].text(bar.get_x() + bar.get_width() / 2, value + 2, f"{value:.1f}%", ha="center", weight="bold", color=INK)
    axes[1].set(ylim=(0, 105), ylabel="Target-condition probability")
    axes[1].set_title("B. Where each result leaves probability", color=INK)
    axes[1].grid(axis="y", alpha=0.16)
    fig.suptitle("A complete information profile: intraoperative frozen section for ovarian masses", weight="bold", color=INK)
    fig.text(
        0.5, 0.015,
        f"Gain {profile['information_bits']:.3f} bits; DER {100 * profile['diagnostic_entropy_reduction']:.1f}%; "
        f"positive/negative shares {100 * profile['positive_information_share']:.1f}%/{100 * profile['negative_information_share']:.1f}%.",
        ha="center", color=INK, fontsize=9.2,
    )
    fig.tight_layout(rect=(0, 0.05, 1, 0.93))
    _save_figure(fig, figures_dir, "figure4_information_profile")


def figure_sequential_budget(figures_dir: Path, sequential: dict) -> None:
    fig, ax = plt.subplots(figsize=(10.2, 3.8))
    first = sequential["mri_standalone_information_bits"]
    second = sequential["bone_scintigraphy_conditional_information_given_mri_bits"]
    remaining = sequential["remaining_entropy_bits"]
    ax.barh([0], [first], color=BLUE, label="MRI information")
    ax.barh([0], [second], left=[first], color=GREEN, label="Additional BS information given MRI")
    ax.barh([0], [remaining], left=[first + second], color="#D9DEE2", label="Unresolved entropy")
    ax.set(xlim=(0, sequential["baseline_entropy_bits"]), yticks=[], xlabel="Bits from the baseline diagnostic-entropy budget")
    ax.axvline(sequential["naive_standalone_sum_bits"], color=ORANGE, linestyle="--", linewidth=1.5)
    ax.text(sequential["naive_standalone_sum_bits"] - 0.005, 0.22, f"Standalone sum\n{sequential['naive_standalone_sum_bits']:.3f} bits", ha="right", va="center", color=ORANGE, fontsize=8.5)
    ax.text(first / 2, 0, f"MRI\n{first:.3f} bits", ha="center", va="center", color="white", weight="bold")
    ax.text(first + second / 2, 0, f"Additional BS given MRI\n+{second:.3f} bits", ha="center", va="center", color="white", weight="bold", fontsize=8.5)
    ax.text(first + second + remaining / 2, 0, f"{remaining:.3f}\nremaining", ha="center", va="center", color=INK, fontsize=8.5)
    ax.set_title("Sequential information is conditional and remains within the baseline entropy budget", weight="bold", color=INK, fontsize=10.5)
    ax.text(0.5, -0.42, "Illustrative MRI-then-bone-scintigraphy pathway; conditional independence is assumed, not estimated.", transform=ax.transAxes, ha="center", color="#596773", fontsize=8.5)
    fig.tight_layout(rect=(0, 0.12, 1, 1))
    _save_figure(fig, figures_dir, "figure_s5_sequential_entropy_budget")


def figure_threshold_boundaries(figures_dir: Path, boundaries: list[dict]) -> None:
    primary = [row for row in boundaries if row["primary_atlas"]]
    grouped = [[100 * row["maximum_pretest_probability"] for row in primary if row["residual_risk_threshold"] == threshold] for threshold in THRESHOLDS]
    fig, ax = plt.subplots(figsize=(9.5, 6.1))
    ax.boxplot(grouped, tick_labels=[f"{100 * threshold:g}%" for threshold in THRESHOLDS], showfliers=False, patch_artist=True, boxprops={"facecolor": "#DCE8F2", "edgecolor": BLUE}, medianprops={"color": INK, "linewidth": 1.5})
    rng = np.random.default_rng(SEED)
    for index, values in enumerate(grouped, start=1):
        jitter = rng.uniform(-0.13, 0.13, len(values))
        ax.scatter(index + jitter, np.maximum(values, 0.005), s=11, color="#657785", alpha=0.35, linewidth=0)
    ax.set_yscale("log")
    ax.set_ylim(0.005, 100)
    ax.set(xlabel="Acceptable post-negative residual-risk threshold", ylabel="Maximum compatible pretest probability (%)")
    ax.set_title("Stopping boundaries vary by test and by the risk a decision permits", weight="bold", color=INK)
    ax.grid(axis="y", alpha=0.18)
    _save_figure(fig, figures_dir, "figure_s2_threshold_boundaries")


def figure_validation(figures_dir: Path, validation: list[dict]) -> None:
    primary = [row for row in validation if row["primary_atlas"]]
    fig, axes = plt.subplots(1, 2, figsize=(10.5, 4.8))
    for ax, measure in zip(axes, ("sensitivity", "specificity")):
        published = np.asarray([float(row[f"published_{measure}"]) for row in primary])
        refit = np.asarray([float(row[f"common_model_{measure}"]) for row in primary])
        ax.scatter(100 * published, 100 * refit, s=26, color=BLUE, alpha=0.58, edgecolor="white", linewidth=0.4)
        ax.plot([0, 100], [0, 100], color=INK, linewidth=1, linestyle="--")
        ax.set(xlim=(0, 100), ylim=(0, 100), xlabel=f"Review-reported {measure} (%)", ylabel=f"Common-model {measure} (%)")
        ax.text(4, 92, f"Median absolute difference: {100 * np.median(np.abs(refit - published)):.1f} points", fontsize=8.5, color=INK)
        ax.grid(alpha=0.15)
    axes[0].set_title("A. Sensitivity", color=INK)
    axes[1].set_title("B. Specificity", color=INK)
    fig.suptitle("A common refit is close to, but not identical to, the source summaries", weight="bold", color=INK)
    fig.tight_layout()
    _save_figure(fig, figures_dir, "figure_s1_common_model_validation")


def graphical_abstract(figures_dir: Path, summary: dict, operating_points: list[dict]) -> None:
    corpus = summary["corpus"]
    fig, ax = plt.subplots(figsize=(12, 6))
    ax.set_xlim(0, 12)
    ax.set_ylim(0, 6)
    ax.axis("off")
    dominance = summary["dominance_concordance"]
    crossings = summary["prior_crossings"]
    boxes = [
        (0.35, BLUE, "REFERENCE DISTRIBUTION", f"{corpus['reviews']} reviews\n{corpus['primary_operating_points']} pooled operating points"),
        (4.45, GREEN, "INFORMATION PROFILE", "Bits gained + DER\nPositive/negative shares\nResult-specific probabilities"),
        (
            8.55,
            ORANGE,
            "PRIOR DEPENDENCE",
            f"{crossings['pairs_with_crossing']} of {dominance['non_dominated_pairs']} non-dominated pairs crossed\n"
            f"{crossings['pairs_with_crossing_in_study_composition_range']} inside study-composition ranges",
        ),
    ]
    for x, color, title, body in boxes:
        ax.add_patch(FancyBboxPatch((x, 1.55), 3.1, 3.1, boxstyle="round,pad=0.08,rounding_size=0.12", facecolor="white", edgecolor=color, linewidth=2.5))
        ax.text(x + 1.55, 4.05, title, ha="center", weight="bold", color=color, fontsize=12)
        ax.text(x + 1.55, 2.75, body, ha="center", va="center", color=INK, fontsize=10.2, linespacing=1.4)
    for start, end in ((3.5, 4.4), (7.6, 8.5)):
        ax.add_patch(FancyArrowPatch((start, 3.1), (end, 3.1), arrowstyle="-|>", mutation_scale=18, linewidth=1.8, color=INK))
    ax.text(6, 5.45, "AN ENTROPY ATLAS OF DIAGNOSTIC EVIDENCE", ha="center", weight="bold", fontsize=17, color=INK)
    ax.text(6, 0.65, "Mutual information measures expected uncertainty reduction; the full profile shows its prior and result-branch dependence.", ha="center", fontsize=10.5, color=INK)
    _save_figure(fig, figures_dir, "graphical_abstract")


def run(args: argparse.Namespace) -> dict:
    write_v7_freeze_manifest(args.v7_analysis_dir, args.v7_manuscript_dir, args.v7_freeze_manifest)
    if args.fetch_reference:
        fetch_reference_archive(args.reference_archive)
    if not args.reference_archive.exists():
        raise FileNotFoundError(
            f"Missing {args.reference_archive}. Run with --fetch-reference to download it."
        )
    packages = sorted(args.packages_dir.glob("CD*-dataPackage.zip"))
    if not packages:
        raise FileNotFoundError(f"No modern Cochrane packages in {args.packages_dir}")

    legacy_reviews, legacy_groups, legacy_rows = load_legacy_reference(args.reference_archive)
    modern_reviews, modern_groups, modern_rows = load_modern_packages(packages)
    external_reviews, external_groups, external_rows, external_audits = load_external_repository_dirs(args.external_dir)
    reviews = legacy_reviews + modern_reviews + external_reviews
    groups = legacy_groups + modern_groups + external_groups
    normalized_rows = legacy_rows + modern_rows + external_rows
    operating_points = []
    all_boundaries = []
    fit_failures = []
    for index, group in enumerate(group for group in groups if group["eligibility"] == "eligible"):
        try:
            point, boundaries = analyze_group(group, args.draws, SEED + index)
        except (RuntimeError, ValueError, np.linalg.LinAlgError) as error:
            group["model_status"] = "failed"
            fit_failures.append({"group_id": group["group_id"], "reason": str(error)})
            continue
        group["model_status"] = "converged"
        operating_points.append(point)
        all_boundaries.extend(boundaries)

    primary = [point for point in operating_points if point["primary_atlas"]]
    primary_boundaries = [row for row in all_boundaries if row["primary_atlas"]]
    curves = operating_point_curves(operating_points)
    reversals = rank_reversals(operating_points)
    reversal_sensitivity = rank_reversal_sensitivity(curves)
    dominance = dominance_concordance(operating_points)
    crossings = prior_crossings(operating_points, dominance)
    matched = j_matched_contrasts(operating_points)
    correlations = metric_correlations(operating_points)
    composition_correlation = observed_composition_correlation(operating_points)
    pre_pmc_correlations = metric_correlations(
        [point for point in operating_points if point["source"] != "PMC Open Access"]
    )
    pmc_correlations = metric_correlations(
        [point for point in operating_points if point["source"] == "PMC Open Access"]
    )
    stability = source_stability(operating_points)
    profiles = information_profiles(operating_points)
    sequential_rows, sequential_summary = sequential_entropy_example(operating_points)
    checks = theory_checks(operating_points, dominance, profiles, sequential_summary)
    validation = validation_rows(operating_points)
    orientation = orientation_audit(groups)
    pmc_validation = pmc_manual_validation(args.pmc_results_dir, groups)
    glmm_panel = glmm_sensitivity_panel(
        groups,
        operating_points,
        limit=args.glmm_panel_size,
        draws=args.draws,
    )
    cochrane_ebm_gate = write_source_gate_audit(
        "Incoming Cochrane EBM corpus",
        args.cochrane_ebm_input_dir,
        args.cochrane_ebm_output_dir,
        "https://www.cochrane.org/form/cochrane-library-data-request",
        "not_present_in_workspace",
    )
    srdr_static_gate = write_source_gate_audit(
        "AHRQ static SRDR exports",
        args.srdr_export_dir,
        args.srdr_output_dir,
        "https://effectivehealthcare.ahrq.gov/srdrplus/srdr-project-indexing",
        "human_verification_required_no_authorized_export_present",
    )
    comparable_pairs = within_review_pair_count(operating_points)
    validation_primary = [row for row in validation if row["primary_atlas"]]
    summary = {
        "analysis_version": VERSION,
        "seed": SEED,
        "draws_per_operating_point": args.draws,
        "source": {
            "reference_record": REFERENCE_RECORD_URL,
            "reference_download": REFERENCE_URL,
            "reference_sha256": REFERENCE_SHA256,
            "reference_md5": REFERENCE_MD5,
            "reference_license": "CC BY-NC 4.0",
            "external_source_audits": external_audits,
        },
        "corpus": {
            "reviews": len(reviews),
            "historical_reviews": len(legacy_reviews),
            "modern_reviews": len(modern_reviews),
            "external_reviews": len(external_reviews),
            "reviews_by_source": {
                source: sum(review["source"] == source for review in reviews)
                for source in sorted({review["source"] for review in reviews})
            },
            "included_study_records": sum(review["included_study_records"] for review in reviews),
            "test_groups": sum(review["test_groups"] for review in reviews),
            "stored_study_test_rows": sum(review["stored_study_test_rows"] for review in reviews),
            "normalized_analysis_rows": len(normalized_rows),
            "model_ready_operating_points": len(operating_points),
            "model_failures": len(fit_failures),
            "primary_operating_points": len(primary),
            "historical_primary_operating_points": sum(point["source"] == "Cochrane DTA Reference Dataset" for point in primary),
            "modern_primary_operating_points": sum(point["source"] == "Modern Cochrane data package" for point in primary),
            "external_primary_operating_points": sum(point["source"] not in {"Cochrane DTA Reference Dataset", "Modern Cochrane data package"} for point in primary),
            "primary_operating_points_by_source": {
                source: sum(point["source"] == source for point in primary)
                for source in sorted({point["source"] for point in primary})
            },
        },
        "model": {
            "name": "continuity-corrected bivariate normal random-effects meta-analysis",
            "estimator": "restricted maximum likelihood",
            "continuity_correction": 0.5,
            "minimum_unique_studies": 5,
            "minimum_paired_studies": 3,
            "repeated_study_rows_excluded": True,
        },
        "standardized_comparison_pretest_probability": 0.20,
        "primary_thresholds": list(THRESHOLDS),
        "within_review_rank_reversals": {
            "pairs": len(reversals),
            "comparable_primary_pairs": comparable_pairs,
            "fraction": len(reversals) / comparable_pairs if comparable_pairs else 0.0,
            "pretest_sensitivity": {
                "range": [float(reversal_sensitivity[0]["pretest_probability"]), float(reversal_sensitivity[-1]["pretest_probability"])],
                "minimum_pairs": min(row["rank_reversals"] for row in reversal_sensitivity),
                "maximum_pairs": max(row["rank_reversals"] for row in reversal_sensitivity),
            },
        },
        "dominance_concordance": checks["dominance"],
        "prior_crossings": {
            "eligible_non_dominated_pairs": len(crossings),
            "pairs_with_crossing": sum(row["crossing_count"] > 0 for row in crossings),
            "pairs_with_crossing_in_1_to_50_percent": sum(
                bool(row["crossing_in_1_to_50_percent"]) for row in crossings
            ),
            "pairs_with_crossing_in_study_composition_range": sum(
                bool(row["crossing_in_study_composition_range"]) for row in crossings
            ),
            "analysis_status": "prespecified_v8_principal_analysis",
        },
        "j_matched_contrasts": {
            "pairs_with_absolute_youden_difference_at_most_0_02": len(matched),
            "median_post_negative_risk_ratio": float(
                np.median([row["post_negative_risk_ratio"] for row in matched])
            ) if matched else "",
            "fraction_above_twofold": float(
                np.mean([row["post_negative_risk_ratio"] > 2.0 for row in matched])
            ) if matched else "",
            "analysis_status": "exploratory_supplement",
        },
        "metric_correlations": correlations,
        "metric_correlation_under_study_composition": composition_correlation,
        "metric_correlations_pre_pmc": pre_pmc_correlations,
        "metric_correlations_pmc": pmc_correlations,
        "source_stability": stability,
        "information_profile": {
            "definition": "baseline entropy, mutual information in bits, DER, result probabilities, post-result probabilities, branch contributions and shares, and stopping boundaries",
            "primary_operating_points": len(profiles),
        },
        "theory_checks": checks,
        "validation": {
            "primary_points": len(validation_primary),
            "median_absolute_sensitivity_difference": float(np.median([abs(float(row["sensitivity_difference"])) for row in validation_primary])),
            "median_absolute_specificity_difference": float(np.median([abs(float(row["specificity_difference"])) for row in validation_primary])),
        },
        "pmc_manual_validation": {
            "tables": len(pmc_validation),
            "orientation_safeguard_tables": sum(
                row["cohort"] == "orientation_safeguard" for row in pmc_validation
            ),
            "included_tables": sum(row["cohort"] == "included" for row in pmc_validation),
            "rejected_near_miss_tables": sum(
                row["cohort"] == "rejected_near_miss" for row in pmc_validation
            ),
            "cell_transcription_errors": sum(
                int(row["cell_transcription_errors"])
                for row in pmc_validation
                if row["cell_transcription_errors"] != "not_applicable"
            ),
            "group_acceptance_errors": sum(
                bool(row["group_acceptance_error"]) for row in pmc_validation
            ),
        },
        "orientation_audit": {
            "excluded_groups": len(orientation),
            "parser_errors": sum(bool(row["parser_error_found"]) for row in orientation),
        },
        "glmm_sensitivity": {
            "selected_groups": len(glmm_panel),
            "converged_groups": sum(bool(row["glmm_converged"]) for row in glmm_panel),
            "model": "bivariate binomial-logit random-effects GLMM with Laplace marginal likelihood",
            "median_absolute_sensitivity_difference": float(
                np.median([abs(float(row["sensitivity_difference"])) for row in glmm_panel if row["glmm_converged"]])
            ),
            "median_absolute_specificity_difference": float(
                np.median([abs(float(row["specificity_difference"])) for row in glmm_panel if row["glmm_converged"]])
            ),
            "median_absolute_der_difference": float(
                np.median([abs(float(row["der_difference"])) for row in glmm_panel if row["glmm_converged"]])
            ),
            "maximum_absolute_der_difference": float(
                max(abs(float(row["der_difference"])) for row in glmm_panel if row["glmm_converged"])
            ),
            "analysis_status": "prespecified_model_defense",
        },
        "gated_sources": {
            "cochrane_ebm": cochrane_ebm_gate,
            "srdr_static": srdr_static_gate,
        },
        "scope_note": "The modeled corpus combines 63 historical Cochrane reviews, two modern Cochrane packages, canonical model-ready deposits from OSF and Zenodo, and strict study-level tables from the PMC Open Access Subset. Figshare, Dryad, and SRDR+ discovery and access denominators are retained when they contribute no eligible primary group.",
    }

    args.output_dir.mkdir(parents=True, exist_ok=True)
    group_manifest = [{key: value for key, value in group.items() if key != "rows"} for group in groups]
    _write_csv(args.output_dir / "corpus_manifest.csv", reviews)
    _write_csv(args.output_dir / "test_group_manifest.csv", group_manifest)
    _write_csv(args.output_dir / "normalized_study_data.csv", normalized_rows)
    _write_csv(args.output_dir / "operating_points_20_percent.csv", operating_points)
    _write_csv(args.output_dir / "operating_point_curves.csv", curves)
    _write_csv(args.output_dir / "threshold_boundaries.csv", all_boundaries)
    _write_csv(args.output_dir / "rank_reversals.csv", reversals)
    _write_csv(args.output_dir / "rank_reversal_sensitivity.csv", reversal_sensitivity)
    _write_csv(args.output_dir / "dominance_concordance.csv", dominance)
    _write_csv(args.output_dir / "prior_crossings.csv", crossings)
    _write_csv(args.output_dir / "j_matched_contrasts.csv", matched)
    _write_csv(args.output_dir / "atlas_metric_correlations.csv", correlations)
    _write_csv(args.output_dir / "source_stability.csv", stability)
    _write_csv(args.output_dir / "information_profiles.csv", profiles)
    _write_csv(
        args.output_dir / "sequential_entropy_example.csv",
        [{"row_type": "summary", **sequential_summary}]
        + [{"row_type": "joint_result", **row} for row in sequential_rows],
    )
    _write_csv(args.output_dir / "model_validation.csv", validation)
    _write_csv(args.output_dir / "pmc_manual_validation.csv", pmc_validation)
    _write_csv(args.output_dir / "orientation_audit.csv", orientation)
    _write_csv(args.model_benchmark_dir / "reml_vs_binomial_glmm.csv", glmm_panel)
    _write_csv(args.output_dir / "model_failures.csv", fit_failures)
    (args.output_dir / "threshold_matrix_20_percent.csv").unlink(missing_ok=True)
    (args.output_dir / "summary.json").write_text(json.dumps(summary, indent=2) + "\n", encoding="utf-8")
    (args.output_dir / "theory_checks.json").write_text(json.dumps(checks, indent=2) + "\n", encoding="utf-8")

    _write_csv(args.manuscript_dir / "tables" / "table1_review_corpus.csv", reviews)
    _write_csv(args.manuscript_dir / "tables" / "table2_operating_point_atlas.csv", primary)
    _write_csv(args.manuscript_dir / "tables" / "table3_dominance_concordance.csv", dominance)
    _write_csv(args.manuscript_dir / "tables" / "table4_information_profiles.csv", profiles)
    _write_csv(args.manuscript_dir / "tables" / "table5_prior_crossings.csv", crossings)
    _style()
    figure_main_corpus(args.manuscript_dir / "figures", summary)
    figure_distribution_and_redundancy(
        args.manuscript_dir / "figures",
        profiles,
        correlations,
        pre_pmc_correlations,
        pmc_correlations,
    )
    figure_dominance_and_crossings(args.manuscript_dir / "figures", dominance, crossings)
    figure_information_profile(args.manuscript_dir / "figures", profiles)
    figure_source_stability(args.manuscript_dir / "figures", stability)
    figure_sequential_budget(args.manuscript_dir / "figures", sequential_summary)
    figure_threshold_boundaries(args.manuscript_dir / "figures", primary_boundaries)
    figure_validation(args.manuscript_dir / "figures", validation)
    figure_glmm_sensitivity(args.manuscript_dir / "figures", glmm_panel)
    figure_j_matched(args.manuscript_dir / "figures", matched)
    graphical_abstract(args.manuscript_dir / "figures", summary, operating_points)
    for svg in (args.manuscript_dir / "figures").glob("*.svg"):
        svg.write_text("\n".join(line.rstrip() for line in svg.read_text(encoding="utf-8").splitlines()) + "\n", encoding="utf-8")
    return summary


def build_parser() -> argparse.ArgumentParser:
    parser = argparse.ArgumentParser(description=__doc__)
    parser.add_argument("--reference-archive", type=Path, default=Path("data/raw/cochrane/CL145_open_set_20181101.zip"))
    parser.add_argument("--fetch-reference", action="store_true")
    parser.add_argument("--packages-dir", type=Path, default=Path("data/raw/cochrane"))
    parser.add_argument("--output-dir", type=Path, default=Path("analysis/cochrane_atlas"))
    parser.add_argument("--manuscript-dir", type=Path, default=Path("docs/manuscript/v8"))
    parser.add_argument("--external-dir", type=Path, action="append", default=[])
    parser.add_argument("--draws", type=int, default=DEFAULT_DRAWS)
    parser.add_argument("--pmc-results-dir", type=Path, default=Path("analysis/pmc_open_atlas"))
    parser.add_argument("--glmm-panel-size", type=int, default=20)
    parser.add_argument("--model-benchmark-dir", type=Path, default=Path("analysis/model_benchmarks"))
    parser.add_argument("--cochrane-ebm-input-dir", type=Path, default=Path("data/raw/cochrane_ebm"))
    parser.add_argument("--cochrane-ebm-output-dir", type=Path, default=Path("analysis/cochrane_ebm_atlas"))
    parser.add_argument("--srdr-export-dir", type=Path, default=Path("data/raw/open_sources/srdr_static/exports"))
    parser.add_argument("--srdr-output-dir", type=Path, default=Path("analysis/srdr_static_atlas"))
    parser.add_argument("--v7-analysis-dir", type=Path, default=Path("analysis/full_atlas"))
    parser.add_argument("--v7-manuscript-dir", type=Path, default=Path("docs/manuscript/v7"))
    parser.add_argument("--v7-freeze-manifest", type=Path, default=Path("analysis/v8_baseline_checksums.csv"))
    return parser


def main() -> int:
    summary = run(build_parser().parse_args())
    print(json.dumps(summary, indent=2))
    return 0


if __name__ == "__main__":
    raise SystemExit(main())
```
