## Supplementary material for "The information in diagnostic tests": Executable analysis code, derived inputs, source-acquisition records, tests, and release metadata: hints_information_action.py

```
"""Reproduce the HINTS information-action analysis and publication figures."""

from __future__ import annotations

import argparse
import csv
import hashlib
import html
import io
import json
import math
import re
import zipfile
from collections import Counter, defaultdict
from pathlib import Path

import matplotlib

matplotlib.use("Agg")
import matplotlib.pyplot as plt
import numpy as np
from matplotlib.patches import FancyArrowPatch, FancyBboxPatch, Rectangle


VERSION = "0.11.0"
SEED = 20260815
DEFAULT_DRAWS = 1_000_000
EXPECTED_SHA256 = "238d207faa3de9e6b67a6eac311dd8ce1781db6e8b60fd9b42ceb6e9eafeb940"
PRIMARY_ANALYSIS = "1"
SUMMARY_ANALYSES = {"1", "3", "5", "7", "8"}
SENSITIVITY_ANALYSES = {"9", "11", "12", "13"}
BLUE = "#0072B2"
ORANGE = "#D55E00"
GREEN = "#009E73"
PURPLE = "#CC79A7"
GOLD = "#E69F00"
INK = "#243447"
LIGHT = "#F3F6F8"


def logistic(value):
    return 1.0 / (1.0 + np.exp(-np.asarray(value)))


def binary_entropy(probability):
    p = np.asarray(probability, dtype=float)
    clipped = np.clip(p, np.finfo(float).tiny, 1.0 - np.finfo(float).eps)
    entropy = -(clipped * np.log2(clipped) + (1.0 - clipped) * np.log2(1.0 - clipped))
    return np.where((p <= 0.0) | (p >= 1.0), 0.0, entropy)


def diagnostic_metrics(sensitivity, specificity, prevalence):
    se = np.asarray(sensitivity, dtype=float)
    sp = np.asarray(specificity, dtype=float)
    p = np.asarray(prevalence, dtype=float)
    positive_probability = p * se + (1.0 - p) * (1.0 - sp)
    negative_probability = p * (1.0 - se) + (1.0 - p) * sp
    positive_posterior = p * se / positive_probability
    negative_posterior = p * (1.0 - se) / negative_probability
    information = (
        binary_entropy(p)
        - positive_probability * binary_entropy(positive_posterior)
        - negative_probability * binary_entropy(negative_posterior)
    )
    baseline = binary_entropy(p)
    diagnostic_entropy_reduction = np.divide(
        information,
        baseline,
        out=np.zeros_like(np.asarray(information, dtype=float)),
        where=baseline > 0,
    )
    clipped_prior = np.clip(p, np.finfo(float).tiny, 1.0 - np.finfo(float).eps)

    def kl_bernoulli(posterior):
        value = np.clip(posterior, np.finfo(float).tiny, 1.0 - np.finfo(float).eps)
        return value * np.log2(value / clipped_prior) + (1.0 - value) * np.log2(
            (1.0 - value) / (1.0 - clipped_prior)
        )

    positive_information = positive_probability * kl_bernoulli(positive_posterior)
    negative_information = negative_probability * kl_bernoulli(negative_posterior)
    with np.errstate(divide="ignore", invalid="ignore"):
        lr_positive = se / (1.0 - sp)
        lr_negative = (1.0 - se) / sp
    return {
        "sensitivity": se,
        "specificity": sp,
        "lr_positive": lr_positive,
        "lr_negative": lr_negative,
        "youden": se + sp - 1.0,
        "positive_probability": positive_probability,
        "positive_posterior": positive_posterior,
        "negative_posterior": negative_posterior,
        "information_bits": information,
        "diagnostic_entropy_reduction": diagnostic_entropy_reduction,
        "positive_result_information_bits": positive_information,
        "negative_result_information_bits": negative_information,
    }


def maximum_pretest_probability(sensitivity: float, specificity: float, threshold: float) -> float:
    lr_negative = (1.0 - sensitivity) / specificity
    return threshold / (threshold + lr_negative * (1.0 - threshold))


def threshold_status(low: float, high: float, threshold: float) -> str:
    """Classify whether an interval lies below, above, or across a threshold."""
    if high < threshold:
        return "below"
    if low > threshold:
        return "above"
    return "crosses"


def _read_csv(archive: zipfile.ZipFile, suffix: str) -> list[dict[str, str]]:
    name = next(name for name in archive.namelist() if name.endswith(suffix))
    with archive.open(name) as handle:
        return list(csv.DictReader(io.TextIOWrapper(handle, encoding="utf-8-sig")))


def _field(fragment: str, label: str) -> str:
    match = re.search(rf"{re.escape(label)}:</b>\s*([^<]+)", fragment, flags=re.I)
    if not match:
        return "Not reported"
    return " ".join(html.unescape(match.group(1)).replace("&nbsp;", " ").split())


def canonical_exam(test_number: str) -> str:
    number = int(test_number)
    if number == 1 or 9 <= number <= 21:
        return "Clinical HINTS"
    if number == 2 or 22 <= number <= 30:
        return "Video-assisted HINTS"
    if number == 3 or 31 <= number <= 40:
        return "Clinical HINTS Plus"
    if number == 4 or 41 <= number <= 48:
        return "Video-assisted HINTS Plus"
    return {
        5: "Head impulse",
        6: "Video head impulse",
        7: "Nystagmus",
        8: "Test of skew",
    }[number]


def canonicalize(rows: list[dict[str, str]]) -> list[dict[str, object]]:
    grouped: dict[tuple[object, ...], dict[str, object]] = {}
    for row in rows:
        key = (
            row["Study"],
            canonical_exam(row["Test number"]),
            int(row["TP"]),
            int(row["FP"]),
            int(row["FN"]),
            int(row["TN"]),
        )
        record = grouped.setdefault(
            key,
            {
                "study": key[0],
                "exam": key[1],
                "tp": key[2],
                "fp": key[3],
                "fn": key[4],
                "tn": key[5],
                "source_test_numbers": [],
                "source_test_names": [],
            },
        )
        record["source_test_numbers"].append(int(row["Test number"]))
        record["source_test_names"].append(row["Test"])
    output = []
    for record in grouped.values():
        record["source_test_numbers"] = ";".join(
            str(value) for value in sorted(set(record["source_test_numbers"]))
        )
        record["source_test_names"] = "; ".join(sorted(set(record["source_test_names"])))
        record["n"] = record["tp"] + record["fp"] + record["fn"] + record["tn"]
        output.append(record)
    return sorted(output, key=lambda row: (row["study"], row["exam"], row["n"]))


def covariance_matrices(parameter: dict[str, str]) -> tuple[np.ndarray, np.ndarray]:
    mean = np.array(
        [
            [float(parameter["SE(E(logitSe))"]) ** 2, float(parameter["Cov(Es)"])],
            [float(parameter["Cov(Es)"]), float(parameter["SE(E(logitSp))"]) ** 2],
        ]
    )
    var_se = float(parameter["Var(logitSe)"])
    var_sp = float(parameter["Var(logitSp)"])
    between_covariance = float(parameter["Corr(logits)"]) * math.sqrt(var_se * var_sp)
    between = np.array([[var_se, between_covariance], [between_covariance, var_sp]])
    for label, matrix in (("mean", mean), ("between-study", between), ("predictive", mean + between)):
        if np.linalg.eigvalsh(matrix).min() < -1e-9:
            raise ValueError(f"{label} covariance matrix is not positive semidefinite")
    return mean, between


def parameter_draws(
    parameter: dict[str, str], draws: int, seed: int
) -> tuple[np.ndarray, np.ndarray, np.ndarray, np.ndarray, np.ndarray, np.ndarray]:
    mean_covariance, between_covariance = covariance_matrices(parameter)
    mean = np.array([float(parameter["E(logitSe)"]), float(parameter["E(logitSp)"])])
    rng = np.random.default_rng(seed)
    pooled = rng.multivariate_normal(mean, mean_covariance, size=draws)
    predictive = rng.multivariate_normal(mean, mean_covariance + between_covariance, size=draws)
    return (
        logistic(pooled[:, 0]),
        logistic(pooled[:, 1]),
        logistic(predictive[:, 0]),
        logistic(predictive[:, 1]),
        mean_covariance,
        between_covariance,
    )


def _interval(values) -> tuple[float, float, float]:
    low, median, high = np.quantile(values, [0.025, 0.5, 0.975])
    return float(low), float(median), float(high)


def _write_csv(path: Path, rows: list[dict[str, object]]) -> None:
    path.parent.mkdir(parents=True, exist_ok=True)
    if not rows:
        return
    with path.open("w", encoding="utf-8", newline="") as handle:
        writer = csv.DictWriter(handle, fieldnames=list(dict.fromkeys(key for row in rows for key in row)))
        writer.writeheader()
        writer.writerows(rows)


def _save_figure(fig: plt.Figure, figures_dir: Path, stem: str) -> None:
    figures_dir.mkdir(parents=True, exist_ok=True)
    for extension in ("png", "pdf", "svg"):
        kwargs = {"dpi": 300} if extension == "png" else {}
        fig.savefig(figures_dir / f"{stem}.{extension}", bbox_inches="tight", **kwargs)
    plt.close(fig)


def _style() -> None:
    plt.rcParams.update(
        {
            "font.family": "DejaVu Sans",
            "font.size": 9,
            "axes.titlesize": 11,
            "axes.labelsize": 9,
            "axes.spines.top": False,
            "axes.spines.right": False,
            "figure.facecolor": "white",
            "axes.facecolor": "white",
            "savefig.facecolor": "white",
        }
    )


def figure_framework(figures_dir: Path) -> None:
    fig, ax = plt.subplots(figsize=(11.5, 4.8))
    ax.set_xlim(0, 11.5)
    ax.set_ylim(0, 4.8)
    ax.axis("off")
    boxes = [
        (0.25, "Accuracy", "Sensitivity\nand specificity", BLUE),
        (2.45, "Information", "Bits and diagnostic\nentropy reduction", GREEN),
        (4.65, "Residual risk", "Probability after\nthe observed result", ORANGE),
        (6.85, "Transportability", "Uncertainty in the mean\nand a new setting", PURPLE),
        (9.05, "Action threshold", "Stop, observe, consult,\nimage, or repeat", GOLD),
    ]
    for x, title, body, color in boxes:
        patch = FancyBboxPatch(
            (x, 1.55), 1.85, 1.65, boxstyle="round,pad=0.05,rounding_size=0.08",
            facecolor="white", edgecolor=color, linewidth=2
        )
        ax.add_patch(patch)
        ax.text(x + 0.925, 2.75, title, ha="center", va="center", weight="bold", color=color)
        ax.text(x + 0.925, 2.12, body, ha="center", va="center", color=INK, linespacing=1.35)
        if x < 9:
            ax.add_patch(FancyArrowPatch((x + 1.88, 2.38), (x + 2.17, 2.38), arrowstyle="-|>", mutation_scale=13, color="#6B7785"))
    ax.text(5.75, 4.15, "Diagnostic information is not diagnostic permission", ha="center", weight="bold", fontsize=16, color=INK)
    ax.text(5.75, 0.68, "Information describes what the test tells us. Consequences determine how much residual risk is acceptable.", ha="center", fontsize=10.5, color=INK)
    ax.plot([2.8, 9.65], [0.38, 0.38], color="#C8D0D8", lw=1.2)
    _save_figure(fig, figures_dir, "figure1_information_action_framework")


def graphical_abstract(figures_dir: Path, primary: dict[str, float], interval: dict[str, float]) -> None:
    fig, ax = plt.subplots(figsize=(12, 5.2))
    ax.set_xlim(0, 12)
    ax.set_ylim(0, 5.2)
    ax.axis("off")
    ax.text(6, 4.72, "HINTS can be highly informative while stopping remains threshold-dependent", ha="center", fontsize=17, weight="bold", color=INK)
    items = [
        (0.45, BLUE, "20%", "Worked scenario\n(not a universal prior)"),
        (3.25, GREEN, f"{100*primary['diagnostic_entropy_reduction']:.1f}%", "Uncertainty removed\nby clinical HINTS"),
        (6.05, ORANGE, f"{100*primary['negative_posterior']:.2f}%", "Residual risk after a\nperipheral HINTS pattern"),
        (8.85, PURPLE, f"{100*interval['predictive_low']:.2f}% to\n{100*interval['predictive_high']:.1f}%", "95% predictive interval\nfor a new setting"),
    ]
    for x, color, number, label in items:
        patch = FancyBboxPatch((x, 1.25), 2.25, 2.55, boxstyle="round,pad=0.08,rounding_size=0.12", facecolor="white", edgecolor=color, linewidth=2.5)
        ax.add_patch(patch)
        ax.text(x + 1.125, 2.72, number, ha="center", va="center", fontsize=21, weight="bold", color=color)
        ax.text(x + 1.125, 1.77, label, ha="center", va="center", fontsize=10.5, color=INK, linespacing=1.3)
        if x < 8:
            ax.add_patch(FancyArrowPatch((x + 2.3, 2.53), (x + 2.72, 2.53), arrowstyle="-|>", mutation_scale=15, color="#6B7785"))
    ax.text(6, 0.57, "The full probability-threshold surface is the result. At the pooled estimate, the 0.5% survey anchor is crossed only below 6.8% pretest risk.", ha="center", fontsize=10.3, color=INK)
    _save_figure(fig, figures_dir, "graphical_abstract")


def figure_curves(
    figures_dir: Path,
    sensitivity: float,
    specificity: float,
    pooled_se: np.ndarray,
    pooled_sp: np.ndarray,
) -> list[dict[str, float]]:
    prevalence = np.linspace(0.01, 0.50, 99)
    point = diagnostic_metrics(sensitivity, specificity, prevalence)
    band_se = pooled_se[:100_000]
    band_sp = pooled_sp[:100_000]
    rows = []
    information_low, information_high, residual_low, residual_high = [], [], [], []
    for p, info, residual in zip(prevalence, point["diagnostic_entropy_reduction"], point["negative_posterior"]):
        simulated = diagnostic_metrics(band_se, band_sp, p)
        info_interval = np.quantile(simulated["diagnostic_entropy_reduction"], [0.025, 0.975])
        residual_interval = np.quantile(simulated["negative_posterior"], [0.025, 0.975])
        information_low.append(info_interval[0])
        information_high.append(info_interval[1])
        residual_low.append(residual_interval[0])
        residual_high.append(residual_interval[1])
        rows.append(
            {
                "pretest_probability": p,
                "diagnostic_entropy_reduction": float(info),
                "diagnostic_entropy_reduction_low": float(info_interval[0]),
                "diagnostic_entropy_reduction_high": float(info_interval[1]),
                "negative_posterior": float(residual),
                "negative_posterior_low": float(residual_interval[0]),
                "negative_posterior_high": float(residual_interval[1]),
            }
        )
    fig, axes = plt.subplots(1, 2, figsize=(11.5, 4.4))
    axes[0].plot(100 * prevalence, 100 * point["diagnostic_entropy_reduction"], color=GREEN, lw=2.3)
    axes[0].fill_between(100 * prevalence, 100 * np.array(information_low), 100 * np.array(information_high), color=GREEN, alpha=0.18, linewidth=0)
    axes[0].set(title="A. Fraction of uncertainty removed", xlabel="Pretest probability (%)", ylabel="Diagnostic entropy reduction (%)", xlim=(1, 50), ylim=(0, 100))
    axes[1].plot(100 * prevalence, 100 * point["negative_posterior"], color=ORANGE, lw=2.3)
    axes[1].fill_between(100 * prevalence, 100 * np.array(residual_low), 100 * np.array(residual_high), color=ORANGE, alpha=0.18, linewidth=0)
    for threshold, line_style in ((0.5, "-"), (1.0, "--"), (2.0, ":")):
        axes[1].axhline(threshold, color="#697783", lw=1, ls=line_style)
        axes[1].text(49.4, threshold + 0.12, f"{threshold:g}%", ha="right", va="bottom", color="#596773", fontsize=8)
    axes[1].set(title="B. Residual risk after a negative result", xlabel="Pretest probability (%)", ylabel="Post-negative probability (%)", xlim=(1, 50), ylim=(0, 6))
    for ax in axes:
        ax.axvline(20, color="#9AA5AF", ls="--", lw=1)
        ax.grid(axis="y", color="#E5E9EC", lw=0.7)
    fig.suptitle("Clinical HINTS remains informative while residual risk rises with pretest probability", fontsize=13, weight="bold", color=INK)
    fig.tight_layout()
    _save_figure(fig, figures_dir, "figure2_information_and_residual_risk")
    return rows


def figure_decision_surface(
    figures_dir: Path, sensitivity: float, specificity: float, pooled_se: np.ndarray, pooled_sp: np.ndarray
) -> list[dict[str, object]]:
    prevalence = np.linspace(0.01, 0.50, 99)
    thresholds = np.linspace(0.001, 0.05, 99)
    lr_negative = (1.0 - pooled_se) / pooled_sp
    lr_low, lr_high = np.quantile(lr_negative, [0.025, 0.975])
    surface = np.empty((len(thresholds), len(prevalence)))
    rows = []
    point_lr_negative = (1.0 - sensitivity) / specificity
    point_residual = prevalence * point_lr_negative / (1.0 - prevalence + prevalence * point_lr_negative)
    residual_low = prevalence * lr_low / (1.0 - prevalence + prevalence * lr_low)
    residual_high = prevalence * lr_high / (1.0 - prevalence + prevalence * lr_high)
    for row_index, threshold in enumerate(thresholds):
        status_codes = np.where(residual_high < threshold, 2, np.where(residual_low > threshold, 0, 1))
        surface[row_index] = status_codes
        rows.extend(
            {
                "pretest_probability": float(p),
                "residual_risk_threshold": float(threshold),
                "point_estimate_below_threshold": bool(point < threshold),
                "pooled_mean_95_interval_low": float(low),
                "pooled_mean_95_interval_high": float(high),
                "confidence_status": ("below" if code == 2 else "above" if code == 0 else "crosses"),
            }
            for p, point, low, high, code in zip(prevalence, point_residual, residual_low, residual_high, status_codes)
        )
    fig, ax = plt.subplots(figsize=(8.2, 5.4))
    cmap = matplotlib.colors.ListedColormap([ORANGE, "#C8D0D8", GREEN])
    image = ax.imshow(
        surface,
        origin="lower",
        aspect="auto",
        extent=(1, 50, 0.1, 5.0),
        vmin=-0.5,
        vmax=2.5,
        cmap=cmap,
    )
    point_boundary = [100 * maximum_pretest_probability(sensitivity, specificity, t) for t in thresholds]
    ax.plot(point_boundary, 100 * thresholds, color="white", lw=2.1, label="Pooled point-estimate boundary")
    ax.scatter([20], [0.5], s=45, facecolor=ORANGE, edgecolor="white", linewidth=0.8, zorder=3, label="20% worked scenario, 0.5% threshold")
    ax.set(xlabel="Pretest probability (%)", ylabel="Residual-risk threshold (%)", title="Threshold status using the pooled-mean 95% confidence interval")
    colorbar = fig.colorbar(image, ax=ax, pad=0.02)
    colorbar.set_ticks([0, 1, 2], labels=["Above", "Crosses", "Below"])
    colorbar.set_label("Position of 95% interval relative to threshold")
    ax.legend(loc="lower right", frameon=True, facecolor="white", fontsize=8)
    fig.tight_layout()
    _save_figure(fig, figures_dir, "figure3_decision_sufficiency_surface")
    return rows


def figure_uncertainty(
    figures_dir: Path, pooled_residual: np.ndarray, predictive_residual: np.ndarray
) -> None:
    fig, ax = plt.subplots(figsize=(8.4, 5.1))
    intervals = [np.quantile(values, [0.025, 0.5, 0.975]) * 100 for values in (pooled_residual, predictive_residual)]
    for y, (low, median, high), color in zip((1, 0), intervals, (BLUE, PURPLE)):
        ax.errorbar(median, y, xerr=[[median - low], [high - median]], fmt="o", color=color, capsize=6, lw=2.4, markersize=7)
    for threshold, line_style in ((0.5, "-"), (1.0, "--"), (2.0, ":")):
        ax.axvline(threshold, color=ORANGE if threshold == 0.5 else "#697783", lw=1.5, ls=line_style)
        ax.text(threshold, 1.32, f"{threshold:g}%", ha="center", va="bottom", fontsize=8, color="#596773")
    ax.set_xscale("log")
    ax.set_yticks([0, 1], ["New-setting predictive interval", "Pooled-mean confidence interval"])
    ax.set(xlabel="Post-negative probability in the 20% worked scenario (%)", ylim=(-0.55, 1.55))
    ax.grid(axis="x", color="#E5E9EC", lw=0.7, which="both")
    fig.suptitle("Between-study heterogeneity widens the range of residual risk", fontsize=13, weight="bold", color=INK)
    fig.tight_layout(rect=(0, 0, 1, 0.96))
    _save_figure(fig, figures_dir, "figure4_average_vs_new_setting")


def _setting_label(location: str) -> str:
    lower = location.lower()
    if "emergency department and" in lower:
        return "ED + outpatient"
    if "emergency department" in lower:
        return "ED"
    if "outpatient and inpatient" in lower:
        return "Outpatient + inpatient"
    return "Hospital specialty unit"


def _specialty_label(value: str) -> str:
    lower = value.lower()
    if "emergency" in lower:
        return "Emergency medicine"
    if "neuro-ot" in lower or "neuro-oph" in lower:
        return "Neurologic subspecialty"
    if lower == "neurology":
        return "Neurology"
    if "otolaryngology" in lower:
        return "Otolaryngology"
    return "Not reported"


def _training_label(value: str) -> str:
    lower = value.lower()
    if "not reported" in lower:
        return "Not reported"
    if "resident" in lower and "attending" in lower:
        return "Resident + attending"
    if "resident" in lower:
        return "Resident"
    if "attending" in lower:
        return "Attending"
    return value


def study_table(
    study_information: list[dict[str, str]],
    study_test_rows: list[dict[str, str]],
    risk_rows: list[dict[str, str]],
) -> list[dict[str, object]]:
    risk = {row["Study"]: row for row in risk_rows}
    by_study: dict[str, list[dict[str, str]]] = defaultdict(list)
    for row in study_test_rows:
        by_study[row["Study"]].append(row)
    risk_fields = {
        "selection_risk": "Domain (judgement): Could the selection of patients have introduced bias?",
        "index_test_risk": "Domain (judgement): Could the conduct or interpretation of the index test have introduced bias?: All tests",
        "reference_standard_risk": "Domain (judgement): Could the reference standard, its conduct, or its interpretation have introduced bias?",
        "flow_risk": "Domain (judgement): Could the patient flow have introduced bias?",
    }
    output = []
    for row in study_information:
        study = row["Study"]
        location = _field(row["Char: Patient characteristics and setting"], "Location")
        specialty = _field(row["Char: Index test"], "Clinician specialty")
        training = _field(row["Char: Index test"], "Clinician experience level")
        tests = sorted({canonical_exam(test["Test number"]) for test in by_study[study] if int(test["Test number"]) <= 8})
        n_max = max(sum(int(test[key]) for key in ("TP", "FP", "FN", "TN")) for test in by_study[study])
        result = {
            "study": study,
            "year": int(row["Year"]),
            "participants": n_max,
            "setting": _setting_label(location),
            "location_verbatim": location,
            "examiner_specialty": _specialty_label(specialty),
            "specialty_verbatim": specialty,
            "training": _training_label(training),
            "tests": "; ".join(tests),
            "target_condition": _field(row["Char: Target condition and reference standard(s)"], "Target condition"),
            "reference_standard": _field(row["Char: Target condition and reference standard(s)"], "Reference standard"),
        }
        result.update({name: risk[study][field] for name, field in risk_fields.items()})
        output.append(result)
    return sorted(output, key=lambda item: (item["year"], item["study"]))


def figure_transportability(figures_dir: Path, studies: list[dict[str, object]]) -> None:
    columns = ["setting", "examiner_specialty", "training", "selection_risk"]
    titles = ["Setting", "Examiner", "Training", "Selection bias"]
    palette = {
        "ED": BLUE,
        "ED + outpatient": "#56B4E9",
        "Outpatient + inpatient": "#8FB9A8",
        "Hospital specialty unit": "#A7B0B8",
        "Emergency medicine": ORANGE,
        "Neurology": PURPLE,
        "Neurologic subspecialty": "#8C6BB1",
        "Otolaryngology": GOLD,
        "Not reported": "#C7CDD2",
        "Resident": "#4DAF4A",
        "Attending": "#1B9E77",
        "Resident + attending": "#80B1D3",
        "Low risk": "#2CA25F",
        "Unclear risk": GOLD,
        "High risk": "#D7301F",
    }
    abbreviations = {
        "ED": "ED", "ED + outpatient": "ED+OP", "Outpatient + inpatient": "OP+IP", "Hospital specialty unit": "Unit",
        "Emergency medicine": "EM", "Neurology": "Neuro", "Neurologic subspecialty": "Subspec", "Otolaryngology": "ENT", "Not reported": "NR",
        "Resident": "Res", "Attending": "Att", "Resident + attending": "Mixed", "Low risk": "Low", "Unclear risk": "Unclear", "High risk": "High",
    }
    fig, ax = plt.subplots(figsize=(10.5, 8.5))
    ax.set_xlim(0, len(columns))
    ax.set_ylim(0, len(studies))
    for row_index, study in enumerate(reversed(studies)):
        for column_index, column in enumerate(columns):
            value = str(study[column])
            ax.add_patch(Rectangle((column_index, row_index), 1, 1, facecolor=palette[value], edgecolor="white", linewidth=1.5))
            ax.text(column_index + 0.5, row_index + 0.5, abbreviations[value], ha="center", va="center", fontsize=8, color="white" if value not in {"Not reported", "Unclear risk"} else INK, weight="bold")
    ax.set_xticks(np.arange(len(columns)) + 0.5, titles)
    ax.xaxis.tick_top()
    ax.set_yticks(np.arange(len(studies)) + 0.5, [str(study["study"]) for study in reversed(studies)])
    ax.tick_params(length=0)
    for spine in ax.spines.values():
        spine.set_visible(False)
    ax.set_title("The evidence was collected in emergency settings, but rarely by emergency physicians", pad=35, fontsize=13, weight="bold", color=INK)
    ax.text(2, -1.0, "ED: emergency department. OP: outpatient. IP: inpatient. NR: not reported.", ha="center", fontsize=8.5, color="#596773")
    fig.tight_layout()
    _save_figure(fig, figures_dir, "figure5_evidence_transportability")


def parameter_result(
    parameter: dict[str, str], prevalence: float, draws: int, seed: int
) -> tuple[dict[str, object], dict[str, np.ndarray]]:
    sensitivity = float(logistic(float(parameter["E(logitSe)"])))
    specificity = float(logistic(float(parameter["E(logitSp)"])))
    point = diagnostic_metrics(sensitivity, specificity, prevalence)
    pooled_se, pooled_sp, predictive_se, predictive_sp, mean_covariance, between_covariance = parameter_draws(parameter, draws, seed)
    pooled = diagnostic_metrics(pooled_se, pooled_sp, prevalence)
    predictive = diagnostic_metrics(predictive_se, predictive_sp, prevalence)
    pooled_residual = _interval(pooled["negative_posterior"])
    predictive_residual = _interval(predictive["negative_posterior"])
    pooled_information = _interval(pooled["diagnostic_entropy_reduction"])
    predictive_information = _interval(predictive["diagnostic_entropy_reduction"])
    pooled_sensitivity = _interval(pooled_se)
    pooled_specificity = _interval(pooled_sp)
    predictive_sensitivity = _interval(predictive_se)
    predictive_specificity = _interval(predictive_sp)
    result = {
        "analysis_number": int(parameter["Analysis number"]),
        "analysis_name": parameter["Analysis name"],
        "test_name": parameter["Test name"],
        "parameter_studies_field": int(parameter["Studies"]),
        "pretest_probability": prevalence,
        "sensitivity": sensitivity,
        "specificity": specificity,
        "lr_positive": float(point["lr_positive"]),
        "lr_negative": float(point["lr_negative"]),
        "youden": float(point["youden"]),
        "positive_posterior": float(point["positive_posterior"]),
        "negative_posterior": float(point["negative_posterior"]),
        "information_bits": float(point["information_bits"]),
        "diagnostic_entropy_reduction": float(point["diagnostic_entropy_reduction"]),
        "positive_result_information_bits": float(point["positive_result_information_bits"]),
        "negative_result_information_bits": float(point["negative_result_information_bits"]),
        "pooled_mean_sensitivity_low": pooled_sensitivity[0],
        "pooled_mean_sensitivity_median": pooled_sensitivity[1],
        "pooled_mean_sensitivity_high": pooled_sensitivity[2],
        "pooled_mean_specificity_low": pooled_specificity[0],
        "pooled_mean_specificity_median": pooled_specificity[1],
        "pooled_mean_specificity_high": pooled_specificity[2],
        "predictive_sensitivity_low": predictive_sensitivity[0],
        "predictive_sensitivity_median": predictive_sensitivity[1],
        "predictive_sensitivity_high": predictive_sensitivity[2],
        "predictive_specificity_low": predictive_specificity[0],
        "predictive_specificity_median": predictive_specificity[1],
        "predictive_specificity_high": predictive_specificity[2],
        "pooled_mean_negative_low": pooled_residual[0],
        "pooled_mean_negative_median": pooled_residual[1],
        "pooled_mean_negative_high": pooled_residual[2],
        "predictive_negative_low": predictive_residual[0],
        "predictive_negative_median": predictive_residual[1],
        "predictive_negative_high": predictive_residual[2],
        "pooled_mean_der_low": pooled_information[0],
        "pooled_mean_der_median": pooled_information[1],
        "pooled_mean_der_high": pooled_information[2],
        "predictive_der_low": predictive_information[0],
        "predictive_der_median": predictive_information[1],
        "predictive_der_high": predictive_information[2],
        "mean_covariance_min_eigenvalue": float(np.linalg.eigvalsh(mean_covariance).min()),
        "between_covariance_min_eigenvalue": float(np.linalg.eigvalsh(between_covariance).min()),
    }
    arrays = {
        "pooled_se": pooled_se,
        "pooled_sp": pooled_sp,
        "predictive_se": predictive_se,
        "predictive_sp": predictive_sp,
        "pooled_residual": pooled["negative_posterior"],
        "predictive_residual": predictive["negative_posterior"],
    }
    return result, arrays


def risk_summary(risk_rows: list[dict[str, str]]) -> list[dict[str, object]]:
    domains = {
        "Patient selection": "Domain (judgement): Could the selection of patients have introduced bias?",
        "Index test": "Domain (judgement): Could the conduct or interpretation of the index test have introduced bias?: All tests",
        "Reference standard": "Domain (judgement): Could the reference standard, its conduct, or its interpretation have introduced bias?",
        "Flow and timing": "Domain (judgement): Could the patient flow have introduced bias?",
    }
    output = []
    for domain, field in domains.items():
        counts = Counter(row[field] for row in risk_rows)
        output.append(
            {
                "domain": domain,
                "low_risk": counts["Low risk"],
                "unclear_risk": counts["Unclear risk"],
                "high_risk": counts["High risk"],
            }
        )
    return output


def run(args: argparse.Namespace) -> dict[str, object]:
    package = Path(args.package)
    digest = hashlib.sha256(package.read_bytes()).hexdigest()
    if digest != EXPECTED_SHA256:
        raise ValueError(f"Unexpected package SHA-256: {digest}")
    with zipfile.ZipFile(package) as archive:
        study_information = _read_csv(archive, "study-information.csv")
        study_test_rows = _read_csv(archive, "study-test-data.csv")
        risk_rows = _read_csv(archive, "risk-of-bias.csv")
        parameter_rows = _read_csv(archive, "parameters.csv")
        analysis_rows = _read_csv(archive, "data-rows.csv")

    output_dir = Path(args.output_dir)
    figures_dir = Path(args.figures_dir)
    tables_dir = Path(args.tables_dir)
    output_dir.mkdir(parents=True, exist_ok=True)
    tables_dir.mkdir(parents=True, exist_ok=True)
    _style()

    canonical = canonicalize(study_test_rows)
    studies = study_table(study_information, study_test_rows, risk_rows)
    parameters = {row["Analysis number"]: row for row in parameter_rows if row["E(logitSe)"]}
    primary_rows = [row for row in analysis_rows if row["Analysis number"] == PRIMARY_ANALYSIS]
    primary_counts = {key: sum(int(row[key]) for row in primary_rows) for key in ("TP", "FP", "FN", "TN")}
    primary_n = sum(primary_counts.values())
    observed_prevalence = (primary_counts["TP"] + primary_counts["FN"]) / primary_n

    primary, arrays = parameter_result(parameters[PRIMARY_ANALYSIS], 0.20, args.draws, SEED)
    summary_results = [primary]
    for index, analysis_number in enumerate(sorted(SUMMARY_ANALYSES - {PRIMARY_ANALYSIS}, key=int)):
        result, _ = parameter_result(parameters[analysis_number], 0.20, args.draws, SEED + 100 + index)
        summary_results.append(result)
    sensitivity_results = []
    for index, analysis_number in enumerate(sorted(SENSITIVITY_ANALYSES, key=int)):
        result, _ = parameter_result(parameters[analysis_number], 0.20, args.draws, SEED + 200 + index)
        sensitivity_results.append(result)

    thresholds = []
    for threshold in (0.001, 0.0025, 0.005, 0.01, 0.02, 0.05):
        thresholds.append(
            {
                "residual_risk_threshold": threshold,
                "maximum_pretest_probability": maximum_pretest_probability(primary["sensitivity"], primary["specificity"], threshold),
            }
        )

    anchor_results = []
    for prevalence in (0.05, 0.10, 0.20, observed_prevalence, 0.40):
        point = diagnostic_metrics(primary["sensitivity"], primary["specificity"], prevalence)
        pooled = diagnostic_metrics(arrays["pooled_se"], arrays["pooled_sp"], prevalence)
        predictive = diagnostic_metrics(arrays["predictive_se"], arrays["predictive_sp"], prevalence)
        pooled_interval = _interval(pooled["negative_posterior"])
        predictive_interval = _interval(predictive["negative_posterior"])
        anchor_results.append(
            {
                "pretest_probability": prevalence,
                "negative_posterior": float(point["negative_posterior"]),
                "information_bits": float(point["information_bits"]),
                "diagnostic_entropy_reduction": float(point["diagnostic_entropy_reduction"]),
                "pooled_mean_negative_low": pooled_interval[0],
                "pooled_mean_negative_median": pooled_interval[1],
                "pooled_mean_negative_high": pooled_interval[2],
                "predictive_negative_low": predictive_interval[0],
                "predictive_negative_median": predictive_interval[1],
                "predictive_negative_high": predictive_interval[2],
            }
        )

    threshold_stability = []
    for threshold in (0.001, 0.0025, 0.005, 0.01, 0.02, 0.05):
        threshold_stability.append(
            {
                "pretest_probability": 0.20,
                "residual_risk_threshold": threshold,
                "point_estimate_below_threshold": primary["negative_posterior"] < threshold,
                "pooled_mean_95_interval_status": threshold_status(primary["pooled_mean_negative_low"], primary["pooled_mean_negative_high"], threshold),
                "new_setting_95_predictive_interval_status": threshold_status(primary["predictive_negative_low"], primary["predictive_negative_high"], threshold),
            }
        )

    integrity = []
    for number, parameter in sorted(parameters.items(), key=lambda item: int(item[0])):
        rows = [row for row in analysis_rows if row["Analysis number"] == number]
        integrity.append(
            {
                "analysis_number": int(number),
                "analysis_name": parameter["Analysis name"],
                "parameter_studies_field": int(parameter["Studies"]),
                "exported_analysis_rows": len(rows),
                "unique_studies_in_exported_rows": len({row["Study"] for row in rows}),
            }
        )

    _write_csv(output_dir / "canonical_evidence.csv", canonical)
    _write_csv(output_dir / "parameter_integrity.csv", integrity)
    _write_csv(output_dir / "risk_of_bias_summary.csv", risk_summary(risk_rows))
    _write_csv(output_dir / "threshold_crossings.csv", thresholds)
    _write_csv(output_dir / "anchor_prevalences.csv", anchor_results)
    _write_csv(output_dir / "threshold_stability_20_percent.csv", threshold_stability)
    _write_csv(output_dir / "sensitivity_analyses.csv", sensitivity_results)
    _write_csv(tables_dir / "table1_study_characteristics.csv", studies)
    _write_csv(tables_dir / "table2_information_action_bundle.csv", summary_results)

    figure_framework(figures_dir)
    curve_rows = figure_curves(figures_dir, primary["sensitivity"], primary["specificity"], arrays["pooled_se"], arrays["pooled_sp"])
    surface_rows = figure_decision_surface(figures_dir, primary["sensitivity"], primary["specificity"], arrays["pooled_se"], arrays["pooled_sp"])
    figure_uncertainty(figures_dir, arrays["pooled_residual"], arrays["predictive_residual"])
    figure_transportability(figures_dir, studies)
    graphical_abstract(
        figures_dir,
        primary,
        {
            "predictive_low": primary["predictive_negative_low"],
            "predictive_high": primary["predictive_negative_high"],
        },
    )
    _write_csv(output_dir / "prevalence_curves.csv", curve_rows)
    _write_csv(output_dir / "decision_surface.csv", surface_rows)

    summary = {
        "analysis_version": VERSION,
        "source_package": package.name,
        "source_sha256": digest,
        "seed": SEED,
        "monte_carlo_draws": args.draws,
        "source_counts": {
            "studies": len(study_information),
            "participants": sum(int(study["participants"]) for study in studies),
            "stored_test_definitions": len({row["Test number"] for row in study_test_rows}),
            "stored_study_test_rows": len(study_test_rows),
            "canonical_rows": len(canonical),
            "repeated_analysis_views_removed": len(study_test_rows) - len(canonical),
            "primary_hints_exported_studies": len({row["Study"] for row in primary_rows}),
            "primary_hints_participants": primary_n,
            "primary_hints_observed_prevalence": observed_prevalence,
            "exclusive_ed_records": sum(study["setting"] == "ED" for study in studies),
            "mixed_ed_outpatient_records": sum(study["setting"] == "ED + outpatient" for study in studies),
            "emergency_medicine_examiner_records": sum(study["examiner_specialty"] == "Emergency medicine" for study in studies),
        },
        "primary_20_percent": primary,
        "threshold_crossings": thresholds,
        "anchor_prevalences": anchor_results,
        "threshold_stability_20_percent": threshold_stability,
        "risk_of_bias": risk_summary(risk_rows),
        "integrity_note": "The Cochrane parameter metadata reports 18 in the Studies field for primary clinical HINTS, while both exported primary tables contain 12 unique studies and 1,890 participants, matching the published review.",
    }
    (output_dir / "summary.json").write_text(json.dumps(summary, indent=2) + "\n", encoding="utf-8")
    return summary


def build_parser() -> argparse.ArgumentParser:
    parser = argparse.ArgumentParser(description=__doc__)
    parser.add_argument("--package", default="data/raw/cochrane/CD015089-dataPackage.zip")
    parser.add_argument("--output-dir", default="analysis/hints")
    parser.add_argument("--figures-dir", default="docs/manuscript/v1/figures")
    parser.add_argument("--tables-dir", default="docs/manuscript/v1/tables")
    parser.add_argument("--draws", type=int, default=DEFAULT_DRAWS)
    return parser


def main() -> int:
    summary = run(build_parser().parse_args())
    print(json.dumps(summary, indent=2))
    return 0


if __name__ == "__main__":
    raise SystemExit(main())
```
