## Supplementary material for "The information in diagnostic tests": Executable analysis code, derived inputs, source-acquisition records, tests, and release metadata: open_repository_dta.py

```
"""Acquire and normalize public repository diagnostic 2 x 2 data."""

from __future__ import annotations

import argparse
import csv
import hashlib
import io
import json
import math
import os
import re
import time
import urllib.error
import urllib.parse
import urllib.request
import xml.etree.ElementTree as ET
import zipfile
from collections import defaultdict
from datetime import datetime, timezone
from pathlib import Path

import duckdb
from openpyxl import load_workbook

try:
    from scripts.cochrane_dta_atlas import _eligibility, _file_hash
except ModuleNotFoundError:  # supports `python scripts/open_repository_dta.py`
    from cochrane_dta_atlas import _eligibility, _file_hash


VERSION = "0.24.0"
USER_AGENT = "diagnostic-information/0.24.0 (+https://github.com/ShuhanCS/diagnostic-information)"
OSF_DB_SHA256 = "9cc33c599dfb9052258f76e6200370c2f5d1df80c075c89e552c5a5287ab36dc"
OSF_AZURE_URL = (
    "https://stsharescoreprodca6829.blob.core.windows.net/score-releases/"
    "share2/0.4.0/inputs/snapshots/osf.db?"
    "versionid=2026-07-18T19%3A11%3A43.2775331Z"
)
OSF_QUERIES = (
    '"diagnostic test accuracy"',
    '"diagnostic accuracy"',
    '"sensitivity and specificity"',
    "sensitivity specificity meta-analysis",
    '"true positive" "false positive"',
    '"2x2" diagnostic',
)
FILE_LIKELY_TERMS = (
    "2x2",
    "2 x 2",
    "data and materials",
    "data repository",
    "dataset",
    "extraction workbook",
    "raw data",
    "source data",
    "supplementary material",
)
SRDR_PUBLISHERS = {
    "systematic review data repository",
    "systematic review data repository srdr",
}
SRDR_DIAGNOSTIC_TERMS = (
    "accuracy",
    "blood gas",
    "diagnos",
    "screening",
    "sensitivity",
    "serology",
    "specificity",
)
OPEN_REPOSITORY_QUERIES = (
    '"diagnostic test accuracy"',
    '"diagnostic accuracy" AND "systematic review"',
    '"diagnostic accuracy" AND "meta-analysis"',
    '"test accuracy" AND "meta-analysis"',
    '"sensitivity and specificity" AND "meta-analysis"',
    '"true positive" AND "false positive" AND "meta-analysis"',
    '"2x2" AND diagnostic AND "meta-analysis"',
)
DIAGNOSTIC_REVIEW_TERMS = (
    "diagnostic accuracy",
    "diagnostic test accuracy",
    "sensitivity and specificity",
    "test accuracy",
    "true positive",
    "true-positive",
    "2x2",
    "2 x 2",
)
REVIEW_TERMS = ("systematic review", "meta-analysis", "meta analysis", "metadta")
METHOD_RECORD_TERMS = (
    "metadata of",
    "r shiny",
    "simulation",
    "tutorial",
    "model for",
    "models for",
    "transformation",
)
SECONDARY_TABLE_TERMS = ("audit", "secondary", "sensitivity", "subgroup", "supplement")
SUPPORTED_EXTENSIONS = {".csv", ".tsv", ".xlsx", ".json", ".zip", ".docx"}
COUNT_ALIASES = {
    "tp": {"tp", "true positive", "true positives", "truepositive", "truepositives"},
    "fp": {"fp", "false positive", "false positives", "falsepositive", "falsepositives"},
    "fn": {"fn", "false negative", "false negatives", "falsenegative", "falsenegatives"},
    "tn": {"tn", "true negative", "true negatives", "truenegative", "truenegatives"},
}
STUDY_ALIASES = {
    "author",
    "citation",
    "first author",
    "reference",
    "study",
    "study id",
    "study identifier",
    "study name",
}
STUDY_IDENTITY_PRIORITY = (
    "first author",
    "author",
    "citation",
    "reference",
    "study name",
    "study id",
    "study identifier",
    "study",
)
AGGREGATE_STUDY_ROWS = {"all studies", "overall", "pooled", "summary", "total"}
GROUP_ALIASES = {
    "assay",
    "cutoff",
    "index test",
    "modality",
    "test",
    "test name",
    "threshold",
}


def utc_now() -> str:
    return datetime.now(timezone.utc).replace(microsecond=0).isoformat().replace("+00:00", "Z")


def normalize_label(value: object) -> str:
    text = re.sub(r"[_\-/]+", " ", str(value or "").strip().lower())
    return " ".join(re.sub(r"[^a-z0-9 ]+", " ", text).split())


def _slug(value: object) -> str:
    return re.sub(r"[^a-z0-9]+", "-", str(value or "").lower()).strip("-")[:80] or "overall"


def _write_csv(path: Path, rows: list[dict]) -> None:
    path.parent.mkdir(parents=True, exist_ok=True)
    if not rows:
        path.write_text("", encoding="utf-8")
        return
    fields: list[str] = []
    for row in rows:
        fields.extend(key for key in row if key not in fields)
    with path.open("w", newline="", encoding="utf-8-sig") as handle:
        writer = csv.DictWriter(handle, fieldnames=fields, extrasaction="ignore")
        writer.writeheader()
        writer.writerows(rows)


class HttpCache:
    def __init__(self, root: Path, token: str = "", delay: float = 0.0):
        self.root = root
        self.token = token
        self.delay = delay
        self.live_requests = 0
        root.mkdir(parents=True, exist_ok=True)

    def _path(self, url: str, suffix: str = ".json") -> Path:
        return self.root / f"{hashlib.sha256(url.encode()).hexdigest()}{suffix}"

    def _json_request(
        self, url: str, method: str = "GET", request_payload: dict | None = None
    ) -> tuple[int, object, bool]:
        serialized = json.dumps(request_payload, sort_keys=True) if request_payload is not None else ""
        cache_key = url if method == "GET" and request_payload is None else f"{method}\n{url}\n{serialized}"
        path = self._path(cache_key)
        if path.exists():
            cached = json.loads(path.read_text(encoding="utf-8"))
            cached_status = int(cached["status"])
            if cached_status not in {0, 429, 500, 502, 503, 504}:
                return cached_status, cached.get("payload"), True
        headers = {"User-Agent": USER_AGENT, "Accept": "application/json"}
        data = serialized.encode("utf-8") if request_payload is not None else None
        if data is not None:
            headers["Content-Type"] = "application/json"
        if self.token:
            headers["Authorization"] = f"Bearer {self.token}"
            headers["api-key"] = self.token
        status, payload = 0, None
        for attempt in range(4):
            try:
                with urllib.request.urlopen(
                    urllib.request.Request(url, headers=headers, data=data, method=method), timeout=90
                ) as response:
                    status = response.status
                    body = response.read()
                try:
                    payload = json.loads(body)
                except json.JSONDecodeError:
                    payload = {"body_sha256": hashlib.sha256(body).hexdigest(), "body_prefix": body[:500].decode("utf-8", "replace")}
                break
            except urllib.error.HTTPError as error:
                status = error.code
                body = error.read()
                try:
                    payload = json.loads(body)
                except json.JSONDecodeError:
                    payload = {"body_sha256": hashlib.sha256(body).hexdigest(), "body_prefix": body[:500].decode("utf-8", "replace")}
                if status not in {429, 500, 502, 503, 504}:
                    break
            except (TimeoutError, urllib.error.URLError) as error:
                payload = {"transport_error": str(error)}
            if attempt < 3:
                time.sleep(2**attempt)
        receipt = {
            "method": method,
            "url": url,
            "request_payload": request_payload,
            "status": status,
            "retrieved_at": utc_now(),
            "payload": payload,
        }
        path.write_text(json.dumps(receipt, ensure_ascii=False) + "\n", encoding="utf-8")
        self.live_requests += 1
        if self.delay:
            time.sleep(self.delay)
        return status, payload, False

    def json(self, url: str) -> tuple[int, object, bool]:
        return self._json_request(url)

    def json_post(self, url: str, payload: dict) -> tuple[int, object, bool]:
        return self._json_request(url, "POST", payload)

    def download(self, url: str, target: Path, max_bytes: int) -> tuple[int, str]:
        if target.exists():
            return 200, _file_hash(target)
        headers = {"User-Agent": USER_AGENT}
        if self.token:
            headers["Authorization"] = f"Bearer {self.token}"
        temporary = target.with_suffix(target.suffix + ".tmp")
        target.parent.mkdir(parents=True, exist_ok=True)
        try:
            with urllib.request.urlopen(
                urllib.request.Request(url, headers=headers), timeout=120
            ) as response, temporary.open("wb") as handle:
                status = response.status
                total = 0
                while block := response.read(1024 * 1024):
                    total += len(block)
                    if total > max_bytes:
                        return 413, ""
                    handle.write(block)
            temporary.replace(target)
            self.live_requests += 1
            return status, _file_hash(target)
        except urllib.error.HTTPError as error:
            return error.code, ""
        finally:
            temporary.unlink(missing_ok=True)


def _datacite_pages(cache: HttpCache, url: str) -> list[dict]:
    records: list[dict] = []
    while url:
        status, payload, _ = cache.json(url)
        if status != 200 or not isinstance(payload, dict):
            raise RuntimeError(f"DataCite request failed ({status}): {url}")
        records.extend(payload.get("data", []))
        url = payload.get("links", {}).get("next")
    return records


def _datacite_row(item: dict) -> dict:
    attributes = item.get("attributes", {})
    title = " | ".join(value.get("title", "") for value in attributes.get("titles") or [])
    description = " ".join(value.get("description", "") for value in attributes.get("descriptions") or [])
    subjects = " | ".join(value.get("subject", "") for value in attributes.get("subjects") or [])
    rights = attributes.get("rightsList") or []
    first_right = rights[0] if rights else {}
    types = attributes.get("types") or {}
    return {
        "doi": item.get("id", "").lower(),
        "title": title,
        "description": re.sub(r"<[^>]+>", " ", description),
        "subjects": subjects,
        "publisher": attributes.get("publisher", ""),
        "publication_year": attributes.get("publicationYear", ""),
        "resource_type": types.get("resourceType", ""),
        "resource_type_general": types.get("resourceTypeGeneral", ""),
        "license": first_right.get("rights", ""),
        "license_url": first_right.get("rightsUri", ""),
        "source_url": attributes.get("url", "") or f"https://doi.org/{item.get('id', '')}",
    }


def discover_osf(cache: HttpCache) -> list[dict]:
    union: dict[str, dict] = {}
    matched: dict[str, set[str]] = defaultdict(set)
    for query in OSF_QUERIES:
        url = (
            "https://api.datacite.org/dois?client-id=cos.osf&query="
            f"{urllib.parse.quote(query)}&page%5Bsize%5D=1000"
        )
        for item in _datacite_pages(cache, url):
            union[item["id"].lower()] = item
            matched[item["id"].lower()].add(query)
    rows = []
    for doi, item in sorted(union.items()):
        row = _datacite_row(item)
        match = re.search(r"osf\.io/([a-z0-9]+)$", doi, flags=re.I)
        row["source"] = "OSF"
        row["source_id"] = match.group(1).lower() if match else ""
        row["matched_queries"] = " | ".join(sorted(matched[doi]))
        text = " ".join(str(row[key]) for key in ("title", "description", "subjects")).lower()
        row["file_probe_selected"] = bool(
            row["resource_type"] in {"Project", "ProjectComponent"}
            or row["resource_type_general"] in {"Dataset", "Project"}
            or any(term in text for term in FILE_LIKELY_TERMS)
        )
        rows.append(row)
    return rows


def _plain_text(value: object) -> str:
    return " ".join(re.sub(r"<[^>]+>", " ", str(value or "")).split())


def _is_diagnostic_review(row: dict) -> bool:
    text = " ".join(
        str(row.get(key, "")) for key in ("title", "description", "subjects")
    ).lower()
    return any(term in text for term in DIAGNOSTIC_REVIEW_TERMS) and any(
        term in text for term in REVIEW_TERMS
    )


def _is_primary_review_record(row: dict) -> bool:
    title = str(row.get("title", "")).lower().strip()
    return (
        _is_diagnostic_review(row)
        and not title.startswith(("metadta", "metabayesdta"))
        and not any(term in title for term in METHOD_RECORD_TERMS)
    )


def _absolute_url(base: str, href: object) -> str:
    return urllib.parse.urljoin(base, str(href or ""))


def discover_zenodo(cache: HttpCache) -> list[dict]:
    union: dict[str, dict] = {}
    matched: dict[str, set[str]] = defaultdict(set)
    for query in OPEN_REPOSITORY_QUERIES:
        url = f"https://zenodo.org/api/records?q={urllib.parse.quote(query)}&size=25"
        while url:
            status, payload, _ = cache.json(url)
            if status != 200 or not isinstance(payload, dict):
                raise RuntimeError(f"Zenodo request failed ({status}): {url}")
            for item in payload.get("hits", {}).get("hits", []):
                source_id = str(item.get("id", ""))
                if not source_id:
                    continue
                union[source_id] = item
                matched[source_id].add(query)
            url = str(payload.get("links", {}).get("next", ""))

    rows = []
    for source_id, item in sorted(union.items(), key=lambda pair: int(pair[0])):
        metadata = item.get("metadata", {})
        links = item.get("links", {})
        license_value = metadata.get("license") or {}
        license_name = license_value.get("id", "") if isinstance(license_value, dict) else str(license_value)
        row = {
            "source": "Zenodo",
            "source_id": source_id,
            "doi": metadata.get("doi", "") or item.get("doi", ""),
            "title": _plain_text(metadata.get("title", "")),
            "description": _plain_text(metadata.get("description", "")),
            "subjects": " | ".join(metadata.get("keywords") or []),
            "publisher": "Zenodo",
            "publication_year": str(metadata.get("publication_date", ""))[:4],
            "resource_type": (metadata.get("resource_type") or {}).get("title", ""),
            "resource_type_general": (metadata.get("resource_type") or {}).get("type", ""),
            "license": license_name,
            "license_url": links.get("license", ""),
            "source_url": links.get("self_html", "") or f"https://zenodo.org/records/{source_id}",
            "api_url": links.get("self", "") or f"https://zenodo.org/api/records/{source_id}",
            "matched_queries": " | ".join(sorted(matched[source_id])),
            "access_status": "not_probed",
        }
        row["file_probe_selected"] = _is_diagnostic_review(row)
        row["primary_candidate"] = _is_primary_review_record(row)
        rows.append(row)
    return rows


def discover_dryad(cache: HttpCache) -> list[dict]:
    base = "https://datadryad.org"
    union: dict[str, dict] = {}
    matched: dict[str, set[str]] = defaultdict(set)
    for query in OPEN_REPOSITORY_QUERIES:
        url = f"{base}/api/v2/search?q={urllib.parse.quote(query)}&per_page=100"
        while url:
            status, payload, _ = cache.json(url)
            if status != 200 or not isinstance(payload, dict):
                raise RuntimeError(f"Dryad request failed ({status}): {url}")
            for item in payload.get("_embedded", {}).get("stash:datasets", []):
                source_id = str(item.get("identifier", "")).removeprefix("doi:")
                if not source_id:
                    continue
                union[source_id] = item
                matched[source_id].add(query)
            next_link = payload.get("_links", {}).get("next", {}).get("href", "")
            url = _absolute_url(base, next_link) if next_link else ""

    rows = []
    for source_id, item in sorted(union.items()):
        links = item.get("_links", {})
        license_url = item.get("license", "")
        row = {
            "source": "Dryad",
            "source_id": source_id,
            "doi": source_id,
            "title": _plain_text(item.get("title", "")),
            "description": _plain_text(item.get("abstract", "")),
            "subjects": " | ".join(item.get("keywords") or []),
            "publisher": "Dryad",
            "publication_year": str(item.get("publicationDate", ""))[:4],
            "resource_type": "Dataset",
            "resource_type_general": "Dataset",
            "license": Path(urllib.parse.urlparse(license_url).path).stem,
            "license_url": license_url,
            "source_url": item.get("sharingLink", "") or f"https://doi.org/{source_id}",
            "version_url": _absolute_url(base, links.get("stash:version", {}).get("href", "")),
            "matched_queries": " | ".join(sorted(matched[source_id])),
            "access_status": "not_probed",
        }
        row["file_probe_selected"] = _is_diagnostic_review(row)
        row["primary_candidate"] = _is_primary_review_record(row)
        rows.append(row)
    return rows


def discover_figshare(cache: HttpCache) -> list[dict]:
    endpoint = "https://api.figshare.com/v2/articles/search"
    union: dict[str, dict] = {}
    matched: dict[str, set[str]] = defaultdict(set)
    for query in OPEN_REPOSITORY_QUERIES:
        page = 1
        while True:
            request_payload = {
                "search_for": query,
                "item_type": 3,
                "page": page,
                "page_size": 100,
            }
            status, payload, _ = cache.json_post(endpoint, request_payload)
            if status != 200 or not isinstance(payload, list):
                raise RuntimeError(f"Figshare request failed ({status}) on page {page}")
            for item in payload:
                source_id = str(item.get("id", ""))
                if not source_id:
                    continue
                union[source_id] = item
                matched[source_id].add(query)
            if len(payload) < request_payload["page_size"]:
                break
            page += 1

    rows = []
    for source_id, item in sorted(union.items(), key=lambda pair: int(pair[0])):
        title = item.get("resource_title", "") or item.get("title", "")
        row = {
            "source": "Figshare",
            "source_id": source_id,
            "doi": item.get("doi", ""),
            "related_article_doi": item.get("resource_doi", ""),
            "title": _plain_text(title),
            "description": _plain_text(item.get("description", "")),
            "subjects": "",
            "publisher": "Figshare",
            "publication_year": str(item.get("published_date", ""))[:4],
            "resource_type": item.get("defined_type_name", "Dataset"),
            "resource_type_general": "Dataset",
            "license": "",
            "license_url": "",
            "source_url": item.get("url_public_html", ""),
            "api_url": item.get("url_public_api", "") or item.get("url", ""),
            "matched_queries": " | ".join(sorted(matched[source_id])),
            "access_status": "not_probed",
        }
        row["file_probe_selected"] = _is_diagnostic_review(row)
        row["primary_candidate"] = _is_primary_review_record(row)
        rows.append(row)
    return rows


def load_osf_inventory(path: Path) -> set[str]:
    if not path.exists():
        raise FileNotFoundError(path)
    observed = _file_hash(path)
    if observed != OSF_DB_SHA256:
        raise ValueError(f"Unexpected OSF inventory SHA-256: {observed}")
    connection = duckdb.connect(str(path), read_only=True)
    try:
        return {row[0] for row in connection.execute("SELECT native_id FROM share_evidence").fetchall()}
    finally:
        connection.close()


def _relation_href(payload: dict, relationship: str) -> str:
    links = payload.get("data", {}).get("relationships", {}).get(relationship, {}).get("links", {})
    related = links.get("related", {})
    return related.get("href", "") if isinstance(related, dict) else str(related or "")


def _license(cache: HttpCache, payload: dict, fallback: tuple[str, str]) -> tuple[str, str]:
    url = _relation_href(payload, "license")
    if not url:
        return fallback
    status, result, _ = cache.json(url)
    if status != 200 or not isinstance(result, dict):
        return fallback
    attributes = result.get("data", {}).get("attributes", {})
    return attributes.get("name", fallback[0]), attributes.get("url", fallback[1])


def _walk_osf_files(cache: HttpCache, providers_url: str) -> tuple[list[dict], int]:
    status, payload, _ = cache.json(providers_url)
    if status != 200 or not isinstance(payload, dict):
        return [], status
    queue = []
    for provider in payload.get("data", []):
        href = _relation_href({"data": provider}, "files")
        if href:
            queue.append(href)
    files: list[dict] = []
    seen = set()
    while queue:
        url = queue.pop(0)
        if url in seen:
            continue
        seen.add(url)
        while url:
            page_status, page, _ = cache.json(url)
            if page_status != 200 or not isinstance(page, dict):
                return files, page_status
            for item in page.get("data", []):
                attributes = item.get("attributes", {})
                if attributes.get("kind") == "folder":
                    href = _relation_href({"data": item}, "files")
                    if href:
                        queue.append(href)
                    continue
                files.append(
                    {
                        "file_id": item.get("id", ""),
                        "file_name": attributes.get("name", ""),
                        "file_size": attributes.get("size", ""),
                        "download_url": item.get("links", {}).get("download", ""),
                    }
                )
            url = page.get("links", {}).get("next")
    return files, 200


def _column_map(header: list[object]) -> tuple[dict[str, int], int | None, list[int]]:
    normalized = [normalize_label(value) for value in header]
    counts: dict[str, int] = {}
    for name, aliases in COUNT_ALIASES.items():
        counts[name] = next((index for index, value in enumerate(normalized) if value in aliases), -1)
    study = next(
        (
            index
            for preferred in STUDY_IDENTITY_PRIORITY
            for index, value in enumerate(normalized)
            if value == preferred
        ),
        None,
    )
    groups = [index for index, value in enumerate(normalized) if value in GROUP_ALIASES]
    return counts, study, groups


def _count(value: object) -> int | None:
    if value is None or str(value).strip() == "":
        return None
    try:
        number = float(value)
    except (TypeError, ValueError):
        return None
    return int(number) if math.isfinite(number) and number >= 0 and number.is_integer() else None


def parse_table(matrix: list[list[object]], identity: dict, sheet: str) -> tuple[list[dict], list[dict], str]:
    header_index = None
    mapping: dict[str, int] = {}
    study_index = None
    group_indexes: list[int] = []
    for index, row in enumerate(matrix[:25]):
        counts, study, groups = _column_map(row)
        if study is not None and all(value >= 0 for value in counts.values()):
            header_index, mapping, study_index, group_indexes = index, counts, study, groups
            break
    if header_index is None or study_index is None:
        return [], [], "required_columns_not_found"

    parsed = []
    header = matrix[header_index]
    for row_number, values in enumerate(matrix[header_index + 1 :], start=header_index + 2):
        study_id = str(values[study_index] if study_index < len(values) else "").strip()
        counts = {
            name: _count(values[column] if column < len(values) else None)
            for name, column in mapping.items()
        }
        if (
            not study_id
            or normalize_label(study_id) in AGGREGATE_STUDY_ROWS
            or any(value is None for value in counts.values())
        ):
            continue
        parsed.append(
            {
                "study_id": study_id,
                **counts,
                "row_number": row_number,
                "group_values": tuple(
                    str(values[column] if column < len(values) else "").strip()
                    for column in group_indexes
                ),
            }
        )
    if not parsed:
        return [], [], "no_valid_2x2_rows"

    duplicates = len({row["study_id"] for row in parsed}) != len(parsed)
    grouped: dict[tuple[str, ...], list[dict]]
    primary = True
    if duplicates and group_indexes:
        grouped = defaultdict(list)
        for row in parsed:
            grouped[row["group_values"]].append(row)
        if any(len({row["study_id"] for row in rows}) != len(rows) for rows in grouped.values()):
            grouped = {("overall",): parsed}
        else:
            primary = False
    else:
        grouped = {("overall",): parsed}

    normalized_rows, groups = [], []
    table_text = f"{identity.get('file_name', '')} {sheet}".lower()
    secondary_table = any(term in table_text for term in SECONDARY_TABLE_TERMS)
    allow_primary = bool(identity.get("allow_primary", True))
    for sequence, (group_values, rows) in enumerate(grouped.items(), start=1):
        label = " | ".join(value for value in group_values if value) or "overall"
        group_id = (
            f"{identity['source'].lower()}:{identity['source_id']}:"
            f"{identity['file_id']}:{_slug(sheet)}:{sequence}:{_slug(label)}"
        )
        reason = _eligibility(rows)
        is_primary = primary and allow_primary and not secondary_table and reason == "eligible"
        if not primary:
            analysis_scope = "external_explicit_group"
        elif not allow_primary:
            analysis_scope = "external_method_dataset"
        elif secondary_table:
            analysis_scope = "external_secondary"
        else:
            analysis_scope = "external_main"
        for row in rows:
            normalized_rows.append(
                {
                    "source": identity["source"],
                    "review_id": identity["review_id"],
                    "review_title": identity["review_title"],
                    "group_id": group_id,
                    "group_number": sequence,
                    "test_name": "Overall structured 2 x 2 data" if label == "overall" else label,
                    "analysis_scope": analysis_scope,
                    "primary_atlas": is_primary,
                    "study_id": row["study_id"],
                    "tp": row["tp"],
                    "fp": row["fp"],
                    "fn": row["fn"],
                    "tn": row["tn"],
                    "source_status": "ok",
                    "source_file": identity["file_name"],
                    "source_sheet": sheet,
                    "source_row": row["row_number"],
                    "source_url": identity["source_url"],
                    "source_sha256": identity["sha256"],
                    "source_license": identity["license"],
                    "source_license_url": identity["license_url"],
                }
            )
        groups.append(
            {
                "source": identity["source"],
                "source_version": identity["sha256"],
                "review_id": identity["review_id"],
                "review_title": identity["review_title"],
                "group_id": group_id,
                "group_number": sequence,
                "test_name": "Overall structured 2 x 2 data" if label == "overall" else label,
                "analysis_scope": analysis_scope,
                "study_rows": len(rows),
                "unique_studies": len({row["study_id"] for row in rows}),
                "paired_studies": sum(row["tp"] + row["fn"] > 0 and row["tn"] + row["fp"] > 0 for row in rows),
                "eligibility": reason,
                "primary_atlas": is_primary,
                "published_studies": "",
                "published_sensitivity": "",
                "published_sensitivity_low": "",
                "published_sensitivity_high": "",
                "published_specificity": "",
                "published_specificity_low": "",
                "published_specificity_high": "",
                "source_file": identity["file_name"],
                "source_sheet": sheet,
                "source_url": identity["source_url"],
                "source_sha256": identity["sha256"],
                "source_license": identity["license"],
                "source_license_url": identity["license_url"],
            }
        )
    return normalized_rows, groups, "parsed"


def _docx_matrices(content: bytes, prefix: str = "") -> list[tuple[str, list[list[object]]]]:
    namespace = "{http://schemas.openxmlformats.org/wordprocessingml/2006/main}"
    with zipfile.ZipFile(io.BytesIO(content)) as archive:
        root = ET.fromstring(archive.read("word/document.xml"))
    matrices = []
    for table_number, table in enumerate(root.iter(f"{namespace}tbl"), start=1):
        matrix = []
        for row in table.findall(f"{namespace}tr"):
            values = []
            for cell in row.findall(f"{namespace}tc"):
                values.append(
                    " ".join(
                        node.text.strip()
                        for node in cell.iter(f"{namespace}t")
                        if node.text and node.text.strip()
                    )
                )
            matrix.append(values)
        if matrix:
            matrices.append((f"{prefix}Table {table_number}", matrix))
    return matrices


def _matrices(path: Path) -> list[tuple[str, list[list[object]]]]:
    suffix = path.suffix.lower()
    if suffix in {".csv", ".tsv"}:
        text = path.read_text(encoding="utf-8-sig", errors="replace")
        delimiter = "\t" if suffix == ".tsv" else csv.Sniffer().sniff(text[:4096], delimiters=",;\t").delimiter
        return [(path.name, list(csv.reader(io.StringIO(text), delimiter=delimiter)))]
    if suffix == ".xlsx":
        workbook = load_workbook(path, read_only=True, data_only=True)
        try:
            return [(sheet.title, [list(row) for row in sheet.iter_rows(values_only=True)]) for sheet in workbook.worksheets]
        finally:
            workbook.close()
    if suffix == ".json":
        payload = json.loads(path.read_text(encoding="utf-8-sig"))
        records = payload if isinstance(payload, list) else payload.get("data", []) if isinstance(payload, dict) else []
        if not records or not all(isinstance(row, dict) for row in records):
            return []
        header = list(dict.fromkeys(key for row in records for key in row))
        return [(path.name, [header] + [[row.get(key) for key in header] for row in records])]
    if suffix == ".docx":
        return _docx_matrices(path.read_bytes())
    if suffix == ".zip":
        matrices = []
        with zipfile.ZipFile(path) as archive:
            for info in archive.infolist():
                member_suffix = Path(info.filename).suffix.lower()
                if info.is_dir() or member_suffix not in {".csv", ".tsv", ".xlsx", ".json", ".docx"} or info.file_size > 25_000_000:
                    continue
                content = archive.read(info)
                if member_suffix in {".csv", ".tsv"}:
                    text = content.decode("utf-8-sig", "replace")
                    delimiter = "\t" if member_suffix == ".tsv" else csv.Sniffer().sniff(text[:4096], delimiters=",;\t").delimiter
                    matrices.append((info.filename, list(csv.reader(io.StringIO(text), delimiter=delimiter))))
                elif member_suffix == ".json":
                    payload = json.loads(content)
                    records = payload if isinstance(payload, list) else payload.get("data", []) if isinstance(payload, dict) else []
                    if records and all(isinstance(row, dict) for row in records):
                        header = list(dict.fromkeys(key for row in records for key in row))
                        matrices.append((info.filename, [header] + [[row.get(key) for key in header] for row in records]))
                elif member_suffix == ".docx":
                    matrices.extend(_docx_matrices(content, f"{info.filename}:"))
                else:
                    workbook = load_workbook(io.BytesIO(content), read_only=True, data_only=True)
                    try:
                        matrices.extend((f"{info.filename}:{sheet.title}", [list(row) for row in sheet.iter_rows(values_only=True)]) for sheet in workbook.worksheets)
                    finally:
                        workbook.close()
        return matrices
    return []


def parse_file(path: Path, identity: dict) -> tuple[list[dict], list[dict], str]:
    rows, groups = [], []
    try:
        matrices = _matrices(path)
    except (
        csv.Error,
        ET.ParseError,
        json.JSONDecodeError,
        KeyError,
        OSError,
        ValueError,
        zipfile.BadZipFile,
    ) as error:
        return [], [], f"parse_error:{type(error).__name__}"
    outcomes = []
    parsed_tables = []
    for sheet, matrix in matrices:
        table_rows, table_groups, outcome = parse_table(matrix, identity, sheet)
        parsed_tables.append((sheet, table_rows, table_groups))
        outcomes.append(outcome)
    if any("per_study" in sheet.lower() and table_rows for sheet, table_rows, _ in parsed_tables):
        parsed_tables = [
            item for item in parsed_tables
            if not ("pilot" in item[0].lower() and item[1])
        ]
    for _, table_rows, table_groups in parsed_tables:
        rows.extend(table_rows)
        groups.extend(table_groups)
    return rows, groups, "parsed" if rows else (outcomes[0] if len(outcomes) == 1 else "no_table_with_required_columns")


def _representation_priority(group: dict) -> tuple[int, str]:
    name = group["source_file"].lower()
    score = 0
    score += 8 if "locked" in name else 0
    score += 6 if "2x2" in name or "2 x 2" in name else 0
    score += 3 if name.endswith((".csv", ".tsv")) else 0
    score += 2 if "diagnostic" in name else 0
    score -= 5 if "study_characteristic" in name or "study characteristic" in name else 0
    return score, group["group_id"]


def canonicalize_representations(
    groups: list[dict], normalized_rows: list[dict], file_manifest: list[dict]
) -> tuple[list[dict], list[dict]]:
    """Keep one representation when a review deposits identical 2 x 2 rows twice."""
    rows_by_group: dict[str, list[dict]] = defaultdict(list)
    for row in normalized_rows:
        rows_by_group[row["group_id"]].append(row)
    signatures: dict[tuple, list[dict]] = defaultdict(list)
    for group in groups:
        signature = tuple(
            sorted(
                (
                    normalize_label(row["study_id"]),
                    int(row["tp"]),
                    int(row["fp"]),
                    int(row["fn"]),
                    int(row["tn"]),
                )
                for row in rows_by_group[group["group_id"]]
            )
        )
        signatures[signature].append(group)
        group["selection_status"] = "canonical"
        group["duplicate_of_group_id"] = ""
    for matches in signatures.values():
        if len(matches) < 2:
            continue
        keep = max(matches, key=_representation_priority)
        for group in matches:
            if group is keep:
                continue
            group["selection_status"] = "duplicate_structured_representation"
            group["duplicate_of_group_id"] = keep["group_id"]
            group["primary_atlas"] = False
    canonical_ids = {
        group["group_id"] for group in groups if group["selection_status"] == "canonical"
    }
    canonical_rows = [row for row in normalized_rows if row["group_id"] in canonical_ids]
    status_by_hash: dict[str, list[str]] = defaultdict(list)
    for group in groups:
        status_by_hash[group["source_sha256"]].append(group["selection_status"])
    for file in file_manifest:
        statuses = status_by_hash.get(file.get("sha256", ""), [])
        if statuses and all(status != "canonical" for status in statuses):
            file["parse_status"] = "parsed_duplicate_only"
            file["normalized_rows"] = 0
            file["test_groups"] = 0
        elif statuses and any(status != "canonical" for status in statuses):
            file["parse_status"] = "parsed_with_duplicate_tables"
    return groups, canonical_rows


def _primary_group_priority(group: dict) -> tuple[int, int, str]:
    text = f"{group.get('source_file', '')} {group.get('source_sheet', '')}".lower()
    score = 0
    score += 8 if "primary" in text else 0
    score += 6 if "master" in text else 0
    score += 4 if "overall" in text or "all_stud" in text or "data_studies" in text else 0
    score -= 8 if any(term in text for term in SECONDARY_TABLE_TERMS) else 0
    score -= 4 if "poolable" in text else 0
    return score, int(group["study_rows"]), group["group_id"]


def select_review_primaries(
    groups: list[dict], normalized_rows: list[dict]
) -> tuple[list[dict], list[dict]]:
    """Retain at most one canonical primary operating point per repository record."""
    by_review: dict[str, list[dict]] = defaultdict(list)
    for group in groups:
        if group.get("selection_status") == "canonical" and group.get("primary_atlas"):
            by_review[group["review_id"]].append(group)
    for candidates in by_review.values():
        keep = max(candidates, key=_primary_group_priority)
        for group in candidates:
            group["primary_atlas"] = group is keep
    primary_by_group = {group["group_id"]: bool(group.get("primary_atlas")) for group in groups}
    for row in normalized_rows:
        row["primary_atlas"] = primary_by_group.get(row["group_id"], False)
    return groups, normalized_rows


def run_osf(args: argparse.Namespace) -> dict:
    raw = args.raw_dir / "osf"
    datacite = HttpCache(raw / "datacite")
    candidates = discover_osf(datacite)
    inventory = load_osf_inventory(args.osf_inventory)
    for row in candidates:
        row["in_fixed_osf_inventory"] = row["source_id"] in inventory
        row["access_status"] = "not_probed"

    token = os.environ.get("OSF_TOKEN", "").strip()
    if not token and args.osf_token_file.exists():
        token = args.osf_token_file.read_text(encoding="utf-8").strip()
    api = HttpCache(raw / "api", token=token, delay=0.05)
    file_manifest, normalized_rows, groups = [], [], []
    for index, candidate in enumerate(row for row in candidates if row["file_probe_selected"]):
        if not token:
            candidate["access_status"] = "osf_token_unavailable"
            continue
        guid = candidate["source_id"]
        status, resolver, _ = api.json(f"https://api.osf.io/v2/guids/{guid}/?resolve=false")
        if status != 200 or not isinstance(resolver, dict):
            candidate["access_status"] = f"guid_http_{status}"
            continue
        referent = _relation_href(resolver, "referent")
        if not referent:
            candidate["access_status"] = "referent_missing"
            continue
        status, detail, _ = api.json(referent)
        if status != 200 or not isinstance(detail, dict):
            candidate["access_status"] = f"referent_http_{status}"
            continue
        public = detail.get("data", {}).get("attributes", {}).get("public")
        if public is False:
            candidate["access_status"] = "not_public"
            continue
        providers = _relation_href(detail, "files")
        if not providers:
            candidate["access_status"] = "files_relationship_missing"
            continue
        license_name, license_url = _license(
            api, detail, (candidate["license"], candidate["license_url"])
        )
        files, file_status = _walk_osf_files(api, providers)
        candidate["public_file_count"] = len(files)
        candidate["access_status"] = "files_enumerated" if file_status == 200 else f"files_http_{file_status}"
        for item in files:
            suffix = Path(item["file_name"]).suffix.lower()
            record = {
                "source": "OSF",
                "source_id": guid,
                "review_id": f"OSF:{guid}",
                "review_title": candidate["title"],
                "doi": candidate["doi"],
                **item,
                "format": suffix.lstrip("."),
                "license": license_name,
                "license_url": license_url,
                "parse_status": "unsupported_format" if suffix not in SUPPORTED_EXTENSIONS else "not_downloaded",
            }
            file_manifest.append(record)
            try:
                size = int(item["file_size"])
            except (TypeError, ValueError):
                size = 0
            if suffix not in SUPPORTED_EXTENSIONS:
                continue
            if size > args.max_file_bytes:
                record["parse_status"] = "file_too_large"
                continue
            safe_name = re.sub(r"[^A-Za-z0-9._-]+", "_", item["file_name"])
            target = raw / "files" / f"{item['file_id']}_{safe_name}"
            download_status, digest = api.download(item["download_url"], target, args.max_file_bytes)
            record["download_status"] = download_status
            record["sha256"] = digest
            if download_status != 200:
                record["parse_status"] = f"download_http_{download_status}"
                continue
            identity = {
                "source": "OSF",
                "source_id": guid,
                "review_id": f"OSF:{guid}",
                "review_title": candidate["title"],
                "file_id": item["file_id"],
                "file_name": item["file_name"],
                "source_url": item["download_url"],
                "sha256": digest,
                "license": license_name,
                "license_url": license_url,
            }
            parsed_rows, parsed_groups, outcome = parse_file(target, identity)
            record["parse_status"] = outcome
            record["normalized_rows"] = len(parsed_rows)
            record["test_groups"] = len(parsed_groups)
            normalized_rows.extend(parsed_rows)
            groups.extend(parsed_groups)
        if (index + 1) % 50 == 0:
            print(f"OSF: probed {index + 1} candidates", flush=True)

    raw_normalized_rows = len(normalized_rows)
    raw_groups = len(groups)
    groups, normalized_rows = canonicalize_representations(groups, normalized_rows, file_manifest)
    groups, normalized_rows = select_review_primaries(groups, normalized_rows)
    canonical_groups = [group for group in groups if group["selection_status"] == "canonical"]
    source_manifest = candidates
    summary = {
        "analysis_version": VERSION,
        "source": "OSF",
        "run_at": utc_now(),
        "fixed_inventory": {
            "path": str(args.osf_inventory),
            "azure_url": OSF_AZURE_URL,
            "sha256": OSF_DB_SHA256,
            "records": len(inventory),
        },
        "discovered_records": len(candidates),
        "file_probe_selected": sum(bool(row["file_probe_selected"]) for row in candidates),
        "fixed_inventory_matches": sum(bool(row["in_fixed_osf_inventory"]) for row in candidates),
        "files_enumerated_projects": sum(row["access_status"] == "files_enumerated" for row in candidates),
        "public_files": len(file_manifest),
        "structured_files": sum(row["format"] in {value.lstrip('.') for value in SUPPORTED_EXTENSIONS} for row in file_manifest),
        "raw_parsed_files": sum(row["parse_status"].startswith("parsed") for row in file_manifest),
        "canonical_parsed_files": sum(row["parse_status"] in {"parsed", "parsed_with_duplicate_tables"} for row in file_manifest),
        "raw_normalized_rows": raw_normalized_rows,
        "raw_test_groups": raw_groups,
        "normalized_rows": len(normalized_rows),
        "test_groups": len(canonical_groups),
        "duplicate_groups": sum(group["selection_status"] != "canonical" for group in groups),
        "eligible_groups": sum(row["eligibility"] == "eligible" for row in canonical_groups),
        "primary_groups": sum(bool(row["primary_atlas"]) for row in canonical_groups),
        "api_live_requests": api.live_requests,
        "token_available": bool(token),
    }
    probed_candidates = [row for row in candidates if row["file_probe_selected"]]
    write_outputs(args.output_dir, source_manifest, probed_candidates, file_manifest, groups, normalized_rows, summary)
    return summary


def _zenodo_files(cache: HttpCache, candidate: dict) -> tuple[list[dict], int]:
    status, detail, _ = cache.json(candidate["api_url"])
    if status != 200 or not isinstance(detail, dict):
        return [], status
    metadata = detail.get("metadata", {})
    license_value = metadata.get("license") or {}
    if isinstance(license_value, dict):
        candidate["license"] = license_value.get("id", candidate.get("license", ""))
    candidate["license_url"] = detail.get("links", {}).get(
        "license", candidate.get("license_url", "")
    )
    files = []
    for item in detail.get("files", []):
        links = item.get("links", {})
        files.append(
            {
                "file_id": item.get("id", "") or item.get("key", ""),
                "file_name": item.get("key", ""),
                "file_size": item.get("size", ""),
                "download_url": links.get("self", "") or links.get("download", ""),
                "upstream_checksum": item.get("checksum", ""),
            }
        )
    return files, 200


def _dryad_files(cache: HttpCache, candidate: dict) -> tuple[list[dict], int]:
    base = "https://datadryad.org"
    if not candidate.get("version_url"):
        return [], 404
    status, version, _ = cache.json(candidate["version_url"])
    if status != 200 or not isinstance(version, dict):
        return [], status
    files_link = version.get("_links", {}).get("stash:files", {}).get("href", "")
    url = _absolute_url(base, files_link)
    files = []
    while url:
        page_status, payload, _ = cache.json(url)
        if page_status != 200 or not isinstance(payload, dict):
            return files, page_status
        for item in payload.get("_embedded", {}).get("stash:files", []):
            links = item.get("_links", {})
            files.append(
                {
                    "file_id": str(item.get("id", "")) or Path(item.get("path", "")).name,
                    "file_name": item.get("path", ""),
                    "file_size": item.get("size", ""),
                    "download_url": _absolute_url(
                        base, links.get("stash:download", {}).get("href", "")
                    ),
                    "upstream_checksum": (
                        f"{item.get('digestType', '')}:{item.get('digest', '')}".strip(":")
                    ),
                }
            )
        next_link = payload.get("_links", {}).get("next", {}).get("href", "")
        url = _absolute_url(base, next_link) if next_link else ""
    return files, 200


def _figshare_files(cache: HttpCache, candidate: dict) -> tuple[list[dict], int]:
    status, detail, _ = cache.json(candidate["api_url"])
    if status != 200 or not isinstance(detail, dict):
        return [], status
    candidate["description"] = _plain_text(detail.get("description", candidate.get("description", "")))
    candidate["subjects"] = " | ".join(detail.get("keywords") or [])
    license_value = detail.get("license") or {}
    if isinstance(license_value, dict):
        candidate["license"] = license_value.get("name", "")
        candidate["license_url"] = license_value.get("url", "")
    files = []
    for item in detail.get("files", []):
        files.append(
            {
                "file_id": str(item.get("id", "")),
                "file_name": item.get("name", ""),
                "file_size": item.get("size", ""),
                "download_url": item.get("download_url", ""),
                "upstream_checksum": f"md5:{item.get('computed_md5', '')}".strip(":"),
            }
        )
    return files, 200


def run_public_repository(args: argparse.Namespace) -> dict:
    adapters = {
        "zenodo": ("Zenodo", discover_zenodo, _zenodo_files),
        "dryad": ("Dryad", discover_dryad, _dryad_files),
        "figshare": ("Figshare", discover_figshare, _figshare_files),
    }
    source, discover, enumerate_files = adapters[args.source]
    raw = args.raw_dir / args.source
    api = HttpCache(raw / "api", delay=0.05)
    projects = discover(api)
    for project in projects:
        if not project["file_probe_selected"]:
            project["access_status"] = "not_diagnostic_review"
    candidates = [row for row in projects if row["file_probe_selected"]]

    file_manifest, normalized_rows, groups = [], [], []
    for index, candidate in enumerate(candidates):
        files, status = enumerate_files(api, candidate)
        candidate["public_file_count"] = len(files)
        candidate["access_status"] = "files_enumerated" if status == 200 else f"files_http_{status}"
        review_id = f"{args.source.upper()}:{candidate['source_id']}"
        for item in files:
            suffix = Path(item["file_name"]).suffix.lower()
            record = {
                "source": source,
                "source_id": candidate["source_id"],
                "review_id": review_id,
                "review_title": candidate["title"],
                "doi": candidate["doi"],
                **item,
                "format": suffix.lstrip("."),
                "license": candidate.get("license", ""),
                "license_url": candidate.get("license_url", ""),
                "parse_status": (
                    "unsupported_format" if suffix not in SUPPORTED_EXTENSIONS else "not_downloaded"
                ),
            }
            file_manifest.append(record)
            try:
                size = int(item["file_size"])
            except (TypeError, ValueError):
                size = 0
            if suffix not in SUPPORTED_EXTENSIONS:
                continue
            if not item["download_url"]:
                record["parse_status"] = "download_url_missing"
                continue
            if size > args.max_file_bytes:
                record["parse_status"] = "file_too_large"
                continue
            safe_name = re.sub(r"[^A-Za-z0-9._-]+", "_", Path(item["file_name"]).name)
            safe_id = re.sub(r"[^A-Za-z0-9._-]+", "_", str(item["file_id"]))
            target = raw / "files" / f"{candidate['source_id'].replace('/', '_')}_{safe_id}_{safe_name}"
            download_status, digest = api.download(
                item["download_url"], target, args.max_file_bytes
            )
            record["download_status"] = download_status
            record["sha256"] = digest
            if download_status != 200:
                record["parse_status"] = f"download_http_{download_status}"
                continue
            identity = {
                "source": source,
                "source_id": candidate["source_id"],
                "review_id": review_id,
                "review_title": candidate["title"],
                "file_id": item["file_id"],
                "file_name": item["file_name"],
                "source_url": item["download_url"],
                "sha256": digest,
                "license": candidate.get("license", ""),
                "license_url": candidate.get("license_url", ""),
                "allow_primary": candidate.get("primary_candidate", False),
            }
            parsed_rows, parsed_groups, outcome = parse_file(target, identity)
            record["parse_status"] = outcome
            record["normalized_rows"] = len(parsed_rows)
            record["test_groups"] = len(parsed_groups)
            normalized_rows.extend(parsed_rows)
            groups.extend(parsed_groups)
        if (index + 1) % 50 == 0:
            print(f"{source}: probed {index + 1} candidates", flush=True)

    raw_normalized_rows = len(normalized_rows)
    raw_groups = len(groups)
    groups, normalized_rows = canonicalize_representations(groups, normalized_rows, file_manifest)
    groups, normalized_rows = select_review_primaries(groups, normalized_rows)
    canonical_groups = [group for group in groups if group["selection_status"] == "canonical"]
    access_note = "Public anonymous API search, file enumeration, and bounded structured-file downloads were executed; narrative PDFs were not transcribed."
    if args.source == "dryad" and not any(row.get("download_status") == 200 for row in file_manifest):
        access_note = "Anonymous search and file enumeration succeeded, but structured-file downloads returned HTTP 401 or a browser proof-of-work page; no numeric rows were available without bypassing that access control."
    summary = {
        "analysis_version": VERSION,
        "source": source,
        "run_at": utc_now(),
        "queries": list(OPEN_REPOSITORY_QUERIES),
        "discovered_records": len(projects),
        "diagnostic_review_candidates": len(candidates),
        "files_enumerated_projects": sum(
            row["access_status"] == "files_enumerated" for row in candidates
        ),
        "public_files": len(file_manifest),
        "structured_files": sum(
            row["format"] in {value.lstrip(".") for value in SUPPORTED_EXTENSIONS}
            for row in file_manifest
        ),
        "downloaded_files": sum(row.get("download_status") == 200 for row in file_manifest),
        "raw_parsed_files": sum(row.get("parse_status", "").startswith("parsed") for row in file_manifest),
        "canonical_parsed_files": sum(
            row.get("parse_status") in {"parsed", "parsed_with_duplicate_tables"}
            for row in file_manifest
        ),
        "raw_normalized_rows": raw_normalized_rows,
        "raw_test_groups": raw_groups,
        "normalized_rows": len(normalized_rows),
        "test_groups": len(canonical_groups),
        "duplicate_groups": sum(group["selection_status"] != "canonical" for group in groups),
        "eligible_groups": sum(row["eligibility"] == "eligible" for row in canonical_groups),
        "primary_groups": sum(bool(row["primary_atlas"]) for row in canonical_groups),
        "api_live_requests": api.live_requests,
        "access_note": access_note,
    }
    write_outputs(
        args.output_dir,
        projects,
        candidates,
        file_manifest,
        groups,
        normalized_rows,
        summary,
    )
    return summary


def discover_srdr(cache: HttpCache) -> list[dict]:
    query = urllib.parse.quote('publisher:"Systematic Review Data Repository"')
    url = f"https://api.datacite.org/dois?prefix=10.26300&query={query}&page%5Bsize%5D=1000"
    rows = []
    for item in _datacite_pages(cache, url):
        row = _datacite_row(item)
        if normalize_label(row["publisher"]) not in SRDR_PUBLISHERS:
            continue
        match = re.search(r"(?:[?&]id=|/projects/)(\d+)", row["source_url"])
        row["source"] = "SRDR+"
        row["source_id"] = match.group(1) if match else ""
        text = " ".join(str(row[key]) for key in ("title", "description", "subjects")).lower()
        row["diagnostic_candidate"] = any(term in text for term in SRDR_DIAGNOSTIC_TERMS)
        row["access_status"] = "not_probed"
        rows.append(row)
    return sorted(rows, key=lambda row: (row["source_id"], row["doi"]))


def run_srdr(args: argparse.Namespace) -> dict:
    raw = args.raw_dir / "srdr"
    datacite = HttpCache(raw / "datacite")
    projects = discover_srdr(datacite)
    candidates = [row for row in projects if row["diagnostic_candidate"]]
    token = os.environ.get("SRDR_API_KEY", "").strip()
    if not token and args.srdr_api_key_file and args.srdr_api_key_file.exists():
        token = args.srdr_api_key_file.read_text(encoding="utf-8").strip()
    api = HttpCache(raw / "api", token=token, delay=0.1)
    public_url = "https://srdrplus.ahrq.gov/api/v2/public_projects.json"
    status, payload, _ = api.json(public_url)
    for candidate in candidates:
        if not token:
            candidate["access_status"] = f"platform_closed_public_endpoint_http_{status}"
        elif status != 200:
            candidate["access_status"] = f"public_endpoint_http_{status}"
        elif not candidate["source_id"]:
            candidate["access_status"] = "project_id_missing"
        else:
            project_url = f"https://srdrplus.ahrq.gov/api/v3/projects/{candidate['source_id']}.json"
            project_status, _, _ = api.json(project_url)
            candidate["access_status"] = "api_project_cached" if project_status == 200 else f"project_http_{project_status}"

    file_manifest, normalized_rows, groups = [], [], []
    if args.srdr_export_dir.exists():
        by_project = {row["source_id"]: row for row in candidates if row["source_id"]}
        for path in sorted(item for item in args.srdr_export_dir.rglob("*") if item.is_file()):
            match = re.search(r"(?:project[_ -]?|^)(\d{2,})", path.name, flags=re.I)
            project_id = match.group(1) if match else ""
            candidate = by_project.get(project_id)
            digest = _file_hash(path)
            record = {
                "source": "SRDR+",
                "source_id": project_id,
                "review_id": f"SRDR:{project_id}" if project_id else "SRDR:unmapped",
                "review_title": candidate["title"] if candidate else "Unmapped SRDR export",
                "doi": candidate["doi"] if candidate else "",
                "file_id": path.name,
                "file_name": path.name,
                "file_size": path.stat().st_size,
                "download_url": candidate["source_url"] if candidate else "",
                "format": path.suffix.lower().lstrip("."),
                "license": candidate["license"] if candidate else "",
                "license_url": candidate["license_url"] if candidate else "",
                "sha256": digest,
            }
            file_manifest.append(record)
            identity = {
                "source": "SRDR+",
                "source_id": project_id or "unmapped",
                "review_id": record["review_id"],
                "review_title": record["review_title"],
                "file_id": path.name,
                "file_name": path.name,
                "source_url": record["download_url"],
                "sha256": digest,
                "license": record["license"],
                "license_url": record["license_url"],
            }
            parsed_rows, parsed_groups, outcome = parse_file(path, identity)
            record["parse_status"] = outcome
            record["normalized_rows"] = len(parsed_rows)
            record["test_groups"] = len(parsed_groups)
            normalized_rows.extend(parsed_rows)
            groups.extend(parsed_groups)

    summary = {
        "analysis_version": VERSION,
        "source": "SRDR+",
        "run_at": utc_now(),
        "datacite_published_projects": len(projects),
        "diagnostic_candidates": len(candidates),
        "public_api_url": public_url,
        "public_api_status": status,
        "platform_status": "ceased_operations_after_2025-11",
        "archive_announcement_url": "https://effectivehealthcare.ahrq.gov/about/srdr-reflections",
        "closure_url": "https://effectivehealthcare.ahrq.gov/news/srdr-closure-farewell",
        "official_archive_status": "announced_but_not_published_as_of_2026-08-17",
        "api_key_available": bool(token),
        "cached_api_projects": sum(row["access_status"] == "api_project_cached" for row in candidates),
        "provided_export_files": len(file_manifest),
        "parsed_files": sum(row.get("parse_status") == "parsed" for row in file_manifest),
        "normalized_rows": len(normalized_rows),
        "test_groups": len(groups),
        "eligible_groups": sum(row["eligibility"] == "eligible" for row in groups),
        "primary_groups": sum(bool(row["primary_atlas"]) for row in groups),
        "access_note": "SRDR+ ceased operations after November 2025. AHRQ says the data files will be available soon but publishes no archive URL, format, or release date; the former public endpoint returned HTTP 405 and no official export was available.",
    }
    write_outputs(args.output_dir, projects, candidates, file_manifest, groups, normalized_rows, summary)
    return summary


def write_outputs(
    output_dir: Path,
    source_manifest: list[dict],
    candidates: list[dict],
    file_manifest: list[dict],
    groups: list[dict],
    normalized_rows: list[dict],
    summary: dict,
) -> None:
    output_dir.mkdir(parents=True, exist_ok=True)
    _write_csv(output_dir / "source_manifest.csv", source_manifest)
    _write_csv(output_dir / "candidate_manifest.csv", candidates)
    _write_csv(output_dir / "file_manifest.csv", file_manifest)
    _write_csv(output_dir / "test_group_manifest.csv", groups)
    _write_csv(output_dir / "normalized_study_data.csv", normalized_rows)
    (output_dir / "access_receipts.json").write_text(
        json.dumps({"analysis_version": VERSION, "source": summary["source"], "run_at": summary["run_at"], "summary": summary}, indent=2) + "\n",
        encoding="utf-8",
    )
    (output_dir / "summary.json").write_text(json.dumps(summary, indent=2) + "\n", encoding="utf-8")


def build_parser() -> argparse.ArgumentParser:
    parser = argparse.ArgumentParser(description=__doc__)
    parser.add_argument(
        "--source",
        choices=("osf", "srdr", "zenodo", "figshare", "dryad"),
        required=True,
    )
    parser.add_argument("--raw-dir", type=Path, default=Path("data/raw/open_sources"))
    parser.add_argument("--output-dir", type=Path)
    parser.add_argument(
        "--osf-inventory",
        type=Path,
        default=Path(r"C:\Users\Shuha\shuputerdesktop\misc\share2-tier1\snapshots\osf.db"),
    )
    parser.add_argument(
        "--osf-token-file",
        type=Path,
        default=Path(r"C:\Users\Shuha\projects\misc\share2-tier1\work\osf.token"),
    )
    parser.add_argument("--max-file-bytes", type=int, default=25_000_000)
    parser.add_argument("--srdr-api-key-file", type=Path)
    parser.add_argument("--srdr-export-dir", type=Path, default=Path("data/raw/open_sources/srdr/exports"))
    return parser


def main() -> int:
    args = build_parser().parse_args()
    if args.output_dir is None:
        args.output_dir = Path("analysis/open_repository_atlas") / args.source
    if args.source == "osf":
        summary = run_osf(args)
    elif args.source == "srdr":
        summary = run_srdr(args)
    else:
        summary = run_public_repository(args)
    print(json.dumps(summary, indent=2))
    return 0


if __name__ == "__main__":
    raise SystemExit(main())
```
