## Supplementary material for "The information in diagnostic tests": Executable analysis code, derived inputs, source-acquisition records, tests, and release metadata: reproduce_bmj_rmr.py

```
"""Reproduce manuscript values and figures from the accompanying derived inputs.

Run in the extracted code supplement: python -m scripts.reproduce_bmj_rmr
Optional: --refit all --figures (all 273 primary fits and all 14 figures).
"""
import argparse
import json
from pathlib import Path

import numpy as np
import pandas as pd
from scipy.special import expit

from scripts import bmj_rmr_v042 as scientific
from scripts import bmj_rmr_v1100_bootstrap as bootstrap
from scripts import bmj_rmr_v1130_robustness as robustness


def reproduce(root, output, refit="3", figures=False):
    output.mkdir(parents=True, exist_ok=True)
    core = root/"analysis/bmj_rmr_v042"
    points = pd.read_csv(core/"operating_points_primary.csv")
    trace = pd.read_csv(core/"source_trace_all_primary.csv")
    anchors = pd.read_csv(core/"derived_operating_points_standard_anchors.csv")
    assert len(points) == 273 and points.review_id.nunique() == 210 and len(trace) == 4104
    computed = []
    for p in (.05, .2, .5):
        for row in points.itertuples():
            values = scientific.expanded_diagnostic_metrics(row.sensitivity, row.specificity, p)
            expected = anchors.loc[(anchors.source==row.source)&(anchors.review_id==row.review_id)&
                                   (anchors.group_id==row.group_id)&(anchors.starting_probability==p)].iloc[0]
            for key in ("information_bits", "fraction_entropy_removed", "post_positive_probability", "post_negative_probability"):
                assert np.isclose(values[key], expected[key], rtol=0, atol=1e-12), (row.group_id,p,key)
            computed.append(dict(source=row.source,review_id=row.review_id,group_id=row.group_id,
                                 starting_probability=p,information_bits=float(values["information_bits"]),
                                 fraction_entropy_removed=float(values["fraction_entropy_removed"])))
    computed = pd.DataFrame(computed)
    computed.to_csv(output/"recomputed_anchors.csv",index=False,lineterminator="\n")
    primary = pd.read_csv(root/"analysis/bmj_rmr_v1100/reference_scale_bootstrap.csv")
    boot = []
    for i,p in enumerate(bootstrap.ANCHORS):
        values=anchors.loc[anchors.starting_probability.eq(p),"fraction_entropy_removed"].to_numpy()
        median,lo,hi=bootstrap.bootstrap_median(values,bootstrap.REPLICATES,bootstrap.SEED+i)
        expected=primary.loc[primary.starting_probability.eq(p)].iloc[0]
        assert np.allclose([median,lo,hi],expected[["median_fraction","ci_low_fraction","ci_high_fraction"]].astype(float),rtol=0,atol=1e-14)
        boot.append(dict(prior=p,median=median,low=lo,high=hi))
    (output/"profile-bootstrap.json").write_text(json.dumps(boot,indent=2)+"\n",encoding="utf-8")
    actual=robustness.run(root,output/"review-robustness")
    expected=pd.read_csv(root/"analysis/bmj_rmr_v1130/review_robustness.csv")
    pd.testing.assert_frame_equal(actual,expected,check_exact=False,atol=1e-14,rtol=0)
    selected=points.sort_values(["source","review_id","group_id"])
    if refit != "all":
        selected=selected.head(int(refit))
    fits=[]
    for row in selected.itertuples():
        rows=trace.loc[(trace.source==row.source)&(trace.review_id==row.review_id)&(trace.group_id==row.group_id)]
        fit=scientific.fit_bivariate_reml(rows[["tp","fp","fn","tn"]].to_dict("records"))
        se,sp=expit(fit["mean"])
        assert np.allclose([se,sp],[row.sensitivity,row.specificity],rtol=0,atol=1e-7),(row.group_id,se,sp)
        fits.append(dict(group_id=row.group_id,sensitivity=float(se),specificity=float(sp)))
    pd.DataFrame(fits).to_csv(output/"refit-checks.csv",index=False,lineterminator="\n")
    if figures:
        import matplotlib.pyplot as plt
        from scripts import bmj_rmr_v1110_visuals as current
        from scripts import bmj_rmr_v1120_visuals as teaching
        from scripts import bmj_rmr_v1131_visuals as repaired
        from scripts.bmj_rmr_v1133_visuals import apply_labels
        data=current.load_data(root)
        data["roles"]=pd.read_csv(root/"analysis/full_atlas/v9/source_role_manifest.csv")
        data["trace"]=trace
        base=current.base
        base.VERSION="1.13.3";base.RELEASE_DATE="2026-09-13"
        base.FIGURE_SPECS={**current.FIGURE_SPECS,**repaired.SPECS,"Figure S10":teaching.S10}
        functions={f"Figure {i}":(lambda gray,i=i:getattr(current,f"figure_{i}")(data,gray)) for i in range(1,5)}
        functions.update({"Figure S1":lambda g:repaired.selection(data,g),"Figure S2":lambda g:repaired.fidelity(data,g),
            "Figure S3":lambda g:base.figure_s3(data["anchors"],data["anchor_summary"],g),
            "Figure S4":lambda g:base.figure_s4(data["recovery"],data["recovery_summary"],data["diagnostics"],data["diagnostic_summary"],g),
            "Figure S5":lambda g:base.figure_s5(data["sensitivity"],data["sensitivity_summary"],data["estimator_effect"],g),
            "Figure S6":lambda g:base.figure_s6(data["contrasts"],data["movement"],g),
            "Figure S7":lambda g:base.figure_s7(data["near_ties"],data["restricted"],g),
            "Figure S8":lambda g:base.figure_s8(data["crossings"],data["cea_profiles"],data["theory_summary"],g),
            "Figure S9":lambda g:repaired.branches(data,g),"Figure S10":teaching.entropy_chord})
        for display,fn in functions.items():
            for gray in (False,True):
                fig=fn(gray)
                apply_labels(fig)
                if display=="Figure 4":
                    for text in fig.findobj(match=plt.Text):
                        text.set_text(text.get_text().replace("How often","Expected frequency").replace(
                            "The expected value weights both outcomes by how often they occur.",
                            "Result frequencies are model-implied at a standardized 5% prior."))
                base.save_figure(fig,output/"figures",display,gray)
    report={"manuscript_version":"1.13.3","profiles":273,"reviews":210,"study_result_appearances":4104,
            "anchor_rows_checked":len(computed),"profile_bootstrap":"PASS","review_weighting_and_bootstrap":"PASS",
            "primary_refits_checked":len(fits),"figures_regenerated":14 if figures else 0}
    (output/"verification.json").write_text(json.dumps(report,indent=2)+"\n",encoding="utf-8")
    print(json.dumps(report,indent=2))


if __name__=="__main__":
    parser=argparse.ArgumentParser(description=__doc__)
    parser.add_argument("--root",type=Path,default=Path(__file__).resolve().parents[1])
    parser.add_argument("--output",type=Path,default=Path("reproduced"))
    parser.add_argument("--refit",default="3",help="number of primary fits to check, or all")
    parser.add_argument("--figures",action="store_true")
    args=parser.parse_args()
    if args.refit!="all" and (not args.refit.isdigit() or not 0<=int(args.refit)<=273):parser.error("--refit must be 0..273 or all")
    reproduce(args.root.resolve(),args.output.resolve(),args.refit,args.figures)
```
